# Factors affecting the impact of COVID-19 vaccination on post COVID-19 conditions among adults: A Systematic Literature Review

**DOI:** 10.1101/2024.10.02.24314603

**Authors:** Abby E. Rudolph, Nadine Al Akoury, Natalija Bogdanenko, Kristen Markus, Isabelle Whittle, Olivia Wright, Hammam Haridy, Julia R. Spinardi, John M. McLaughlin, Moe H. Kyaw

**Affiliations:** Pfizer Inc., New York, USA; Pfizer Emerging Markets, Lebanon; Adelphi Values PROVE, Bollington, SK10 5JB, UK; Pfizer Emerging Markets, UAE; Pfizer Emerging Markets, Brazil

**Author notes:** CORRESPONDING AUTHOR: Abby Rudolph, PhD.

**Keywords:** Post-COVID Conditions, COVID-19 vaccine, SARS-CoV-2, Long COVID, vaccine effectiveness

## Abstract

**Importance:** Existing systematic literature reviews (SLRs) on COVID-19 vaccine effectiveness (VE) against post-COVID-19 conditions (PCC) document high heterogeneity across studies, but have not compared VE across design features known to impact PCC burden or VE against other COVID-19 endpoints.

**Objective:** This SLR summarizes the evidence across studies among predominately adults that report an adjusted measure of association for the relationship between vaccination and PCC, by timing of vaccination relative to infection or PCC-onset and across different study characteristics.

**Evidence review:** A comprehensive search strategy was developed within the OVID platform across EMBASE, MEDLINE® and Evidence-Based Medicine reviews, and supplemented with WHO COVID library and Google Scholar® searches, to collate evidence on vaccination and PCC published or posted as pre-prints between January 1^st^, 2020 and July 18^th^, 2023. JBI Critical Appraisal Checklists were used to assess each study’s risk of bias.

**Findings:** This review included 97 studies and synthesized results from 56 studies with low risk of bias that reported adjusted measures for the association between vaccination and PCC. Overall, 77% of pre-infection adjusted VE (aVE) estimates (vs. unvaccinated) were statistically significant (range: 7%–95%), 80% of estimates reflecting a mix of those vaccinated before and after infection were statistically significant (range: 62%–73%), one of five estimates reflecting vaccination after PCC onset was statistically significant (aVE=41%), 43% of post-infection vaccination estimates were statistically significant (two were protective [range: 28%–40%] and one was not [aVE=-47%]), and 46% of estimates not specifying vaccination timing were statistically significant (23 were protective [range: 29%–75%] and one was not [aVE=-132%]). Statistically significant pre-infection aVE estimates were slightly higher for mRNA (range: 14%–84%) than non-mRNA vaccines (range: 16%–38%) and aVE ranges during (4 studies; range: 10%–70%) and before Omicron predominance (10 studies; range: 7%–50%) overlapped. Pre-infection vaccination was protective regardless of vaccine type, number of doses received, PCC definition, predominant variant, and severity of acute infections included.

**Conclusions and Relevance:** Collectively our findings suggest that COVID-19 vaccination received prior to SARS-CoV-2 infection reduces the subsequent risk of developing PCC regardless of the predominant variant circulating.

**Key points:** *Question:* Do measures of COVID-19 vaccine effectiveness against post-COVID-19 conditions (PCC) vary by timing of vaccine relative to SARS-CoV-2 infection or PCC onset, vaccine type and number of doses received, PCC definition, predominant SARS-CoV-2 variant, and disease severity?

*Findings:* COVID-19 vaccination before SARS-CoV-2 infection appeared to reduce the risk of PCC (vs. unvaccinated). Compared with other COVID-19 vaccine types, mRNA vaccines seemed to offer greater protection, and a dose response was observed for mRNA vaccines.

*Meaning:* Despite heterogeneity across included studies, pre-infection vaccination reduced the risk of ≥1 PCC, regardless of SARS-CoV-2 variant, proportion of sample hospitalized, and PCC definition.

## Introduction

By July 21, 2024, there were ∼776 million confirmed COVID-19 cases globally.^1^ Due to under-reporting of cases post-pandemic and under-detection of cases globally, this is likely an underestimate. An estimated 10%–30% of cases experience new-onset or persisting signs, symptoms, or conditions following the acute phase of a SARS-CoV-2 infection^2^ and those with more severe acute infections are at greater risk.^3^ Although the definition varies by country and across the literature, this is commonly referred to as long COVID, post-COVID-19 conditions (PCC), or post-acute sequelae of COVID-19 (PASC).^4,5^ Al-Aly and colleagues estimated that by the end of 2023, ∼400 million people had experienced PCC at 3 months post-infection, corresponding with an economic impact of $1 trillion each year.^6^ The cumulative PCC burden, economic impact, and disability-adjusted-life-years lost due to PCC^7–9^ will continue to rise as new infections occur each day.

Because COVID-19 vaccines prevent both symptomatic infection and the development of severe COVID-191Zrelated outcomes,^10–14^ pre-infection vaccination may reduce the risk of PCC, as many systematic literature reviews (SLRs) have suggested.^15–23^ Vaccination after PCC-onset could resolve or ameliorate PCC stemming from persisting viral load,^24,25^ but prior SLRs report inconclusive findings.^18,23,26,27^ Existing SLRs are limited, as they have not synthesized findings across key study characteristics known to impact PCC burden or vaccine effectiveness (VE) (i.e., PCC data source and follow-up period; vaccine type and manufacturer, number of doses received, and time since vaccination; population; inclusion of reinfections; predominant variant).

This SLR summarizes the evidence across studies on predominately adults that report adjusted measures of association for the relationship between vaccination and PCC, by timing of vaccination and across different study design features.

## Materials and Methods

This SLR was conducted in accordance with the Cochrane group^28^ and Preferred Reporting Items for Systematic Reviews and Meta-Analyses (PRISMA) guidelines for SLRs.^29^ Two reviewers (IW and OW) assessed each study’s risk of bias using JBI Critical Appraisal Checklists.^30^ The study protocol was registered in PROSPERO (CRD42023446417).

A comprehensive search strategy was developed within the OVID platform across EMBASE, MEDLINE® and Evidence-Based Medicine reviews, and supplemented with WHO COVID library and Google Scholar® searches to collate evidence relating to vaccination and PCC published or posted on pre-print servers between January 1^st^, 2020 and July 18^th^ 2023 (English language only) (**eTable 1**).

Abstracts of retrieved citations were screened according to study PICOS criteria (**eTable 2**) by two independent reviewers (IW and OW); a third reviewer (KM) resolved discrepancies. Data from studies meeting the PICOS criteria were extracted by one reviewer and verified by a second. For each eligible study, methodology, participant characteristics, acute infection severity, definitions for vaccination and PCC, and other study design attributes were extracted (**eTable 3**).

The primary outcome was presence of ≥1 PCC. Adjusted vaccine effectiveness (aVE) against ≥1 PCC was calculated from adjusted estimates of relative risk (aRR) (e.g., risk ratios, odds ratios, or hazard ratios) using the equation aVE=(1-aRR)x100.

## Results

### Summary of included articles

This review included 97 studies (**Figure 1**), which were categorized according to timing of vaccination relative to infection or PCC-onset, vaccine type and manufacturer, number of doses received, time since vaccination, PCC data source and follow-up period, proportion of sample hospitalized during the acute infection, predominant variant circulating when diagnosed with COVID-19 or testing positive for SARS-CoV-2, and whether or not reinfections were included. The predominant variant during infection was reported in 30 of the 97 studies; when not reported (n=67 studies), it was determined using CoVariants, an online tracking database for predominating variant sequences by country.^31^ Variant of infection could not be determined for only one (1.8%) study. Of the 97 included studies, 80 were rated as having low risk of bias (**eTables 4-6**). Of these 80 studies, 24 studies presented only unadjusted estimates and were not included in analyses. Thus, findings from the 56 studies reporting adjusted estimates for the relationship between vaccination and PCC are summarized below (**Figure 1**).

**Figure 1.**
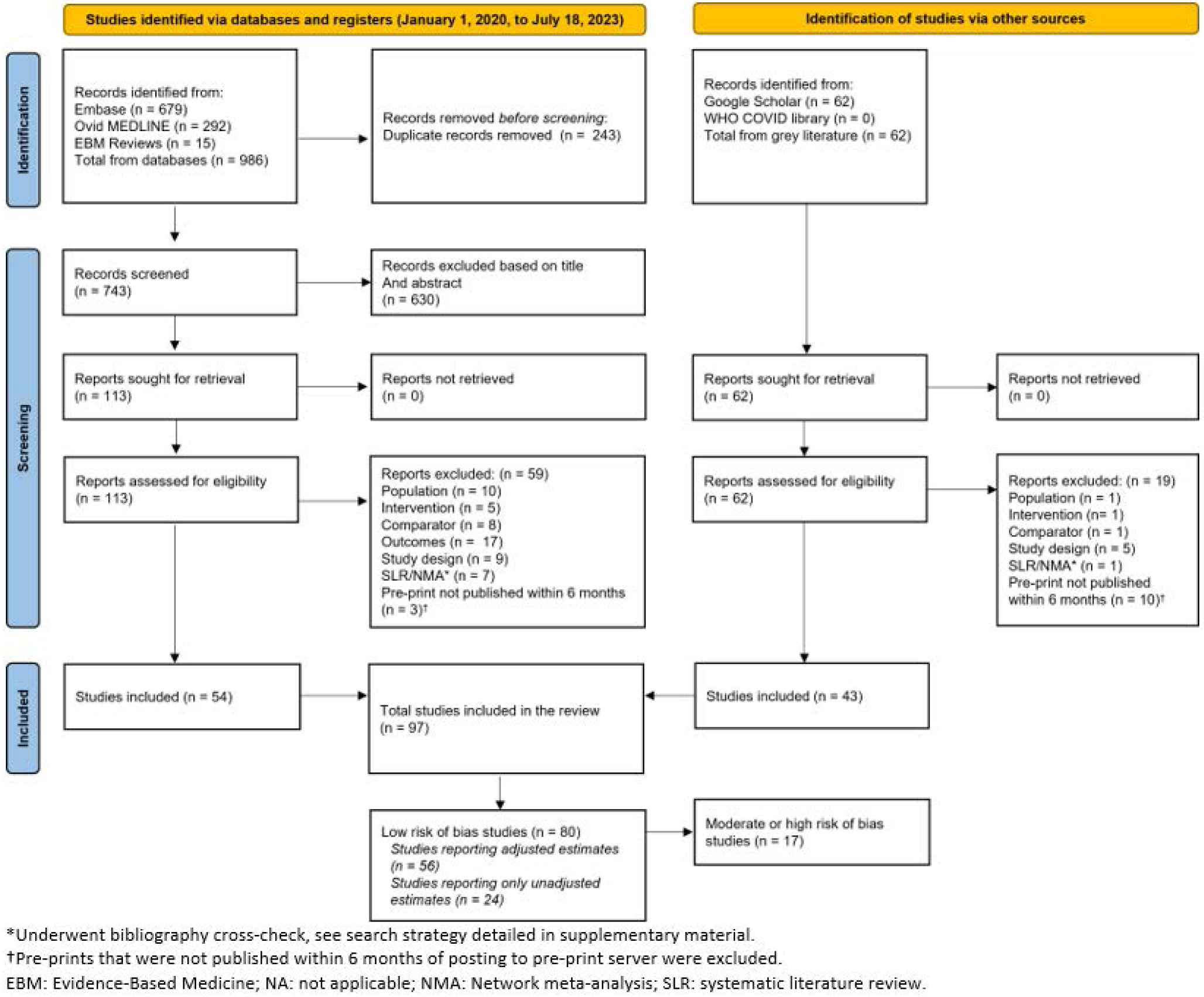
PRISMA flow diagram of included studies.

Across these 56 studies, estimates for the relationship between vaccination and PCC were calculated when vaccination occurred before infection (n=39 studies), after infection (n=4 studies), after PCC onset (n=3 studies), and either before or after infection (i.e., ‘mixed timing’, n=7 studies). Three studies reported estimates for more than one vaccination timepoint and 6 studies did not report the timing of vaccination relative to infection or PCC onset. The most common study design was a retrospective cohort (35.7%, n=20), followed by cross-sectional (32.1%, n=18), prospective cohort (25.0%, n=14), case-control (5.4%, n=3), and ambidirectional cohort (1.8%, n=1). Most studies were conducted in Europe (39.3%), followed by the Americas (37.5%), South-East Asia (7.1%), the Western Pacific and global (each 5.4%), the Eastern Mediterranean (3.6%), and Africa (1.8%). Twenty-six studies (46.4%) were conducted before Omicron predominance, 8 (14.3%) during Omicron predominance, and 21 (39.3%) both before and during Omicron predominance. Most studies (n=37; 66.1%) measured patient-reported PCC and 18 (32.1%) relied on diagnostic and procedural codes from electronic health records or health insurance claims data (**eTable 7**).

In the 56 studies with low risk of bias, adjusted estimates for the relationship between vaccination and PCC were used to compute aVE against ≥1 PCC (vs. unvaccinated; n=39 studies), comparative aVEs against ≥1 PCC (n=11 studies; **eFigure 1; eTable 8**), and aVE against ≥1 specific PCC symptom/condition across 4 organ systems (n=9 studies; **eFigure 2; eTable 9**). Ten studies estimated associations that could not be converted to aVE (**eTable 10**). Thirteen studies reported adjusted estimates for ≥2 of the above relationship types (**eTable 11**).

**Figure 2.**
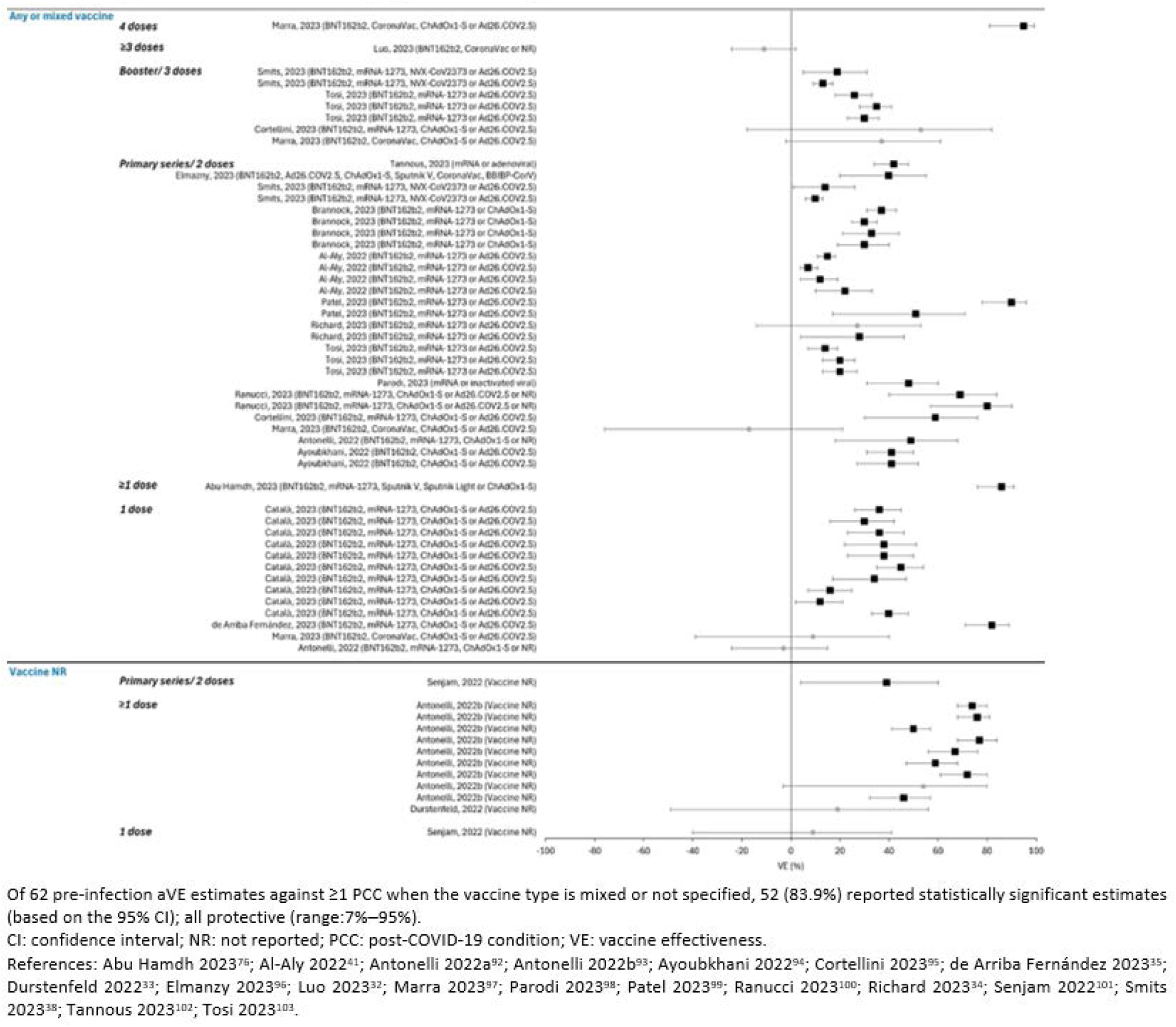
Adjusted Pre-infection VE against ≥1 PCC (vs. Unvaccinated) for Any or Unspecified Vaccine Type or Manufacturer, by Vaccine Type and Number of Doses.

### Adjusted VE against ≥1 PCC, by timing of vaccination relative to infection or PCC onset

Thirty-nine studies reported 134 adjusted estimates where aVE against ≥1 PCC (vs. unvaccinated) could be computed. Overall, 93 estimates (n=31 publications) evaluated pre-infection vaccination, 7 estimates (n=4 publications) evaluated post-infection vaccination, 5 estimates (n=1 publication) evaluated vaccination after PCC-onset, 5 estimates (n=2 publications) reflect a mix of those vaccinated before and after infection, and 24 estimates (n=4 publications) did not report vaccination timing (**Figures 2 and 3; eFigure 3; eTables 12, 13 and 14**). Three studies reported estimates for two different vaccination timepoints (two reported both pre- and post-infection estimates;^32,33^ one reported pre-infection and post-PCC-onset estimates).^34^

**Figure 3.**
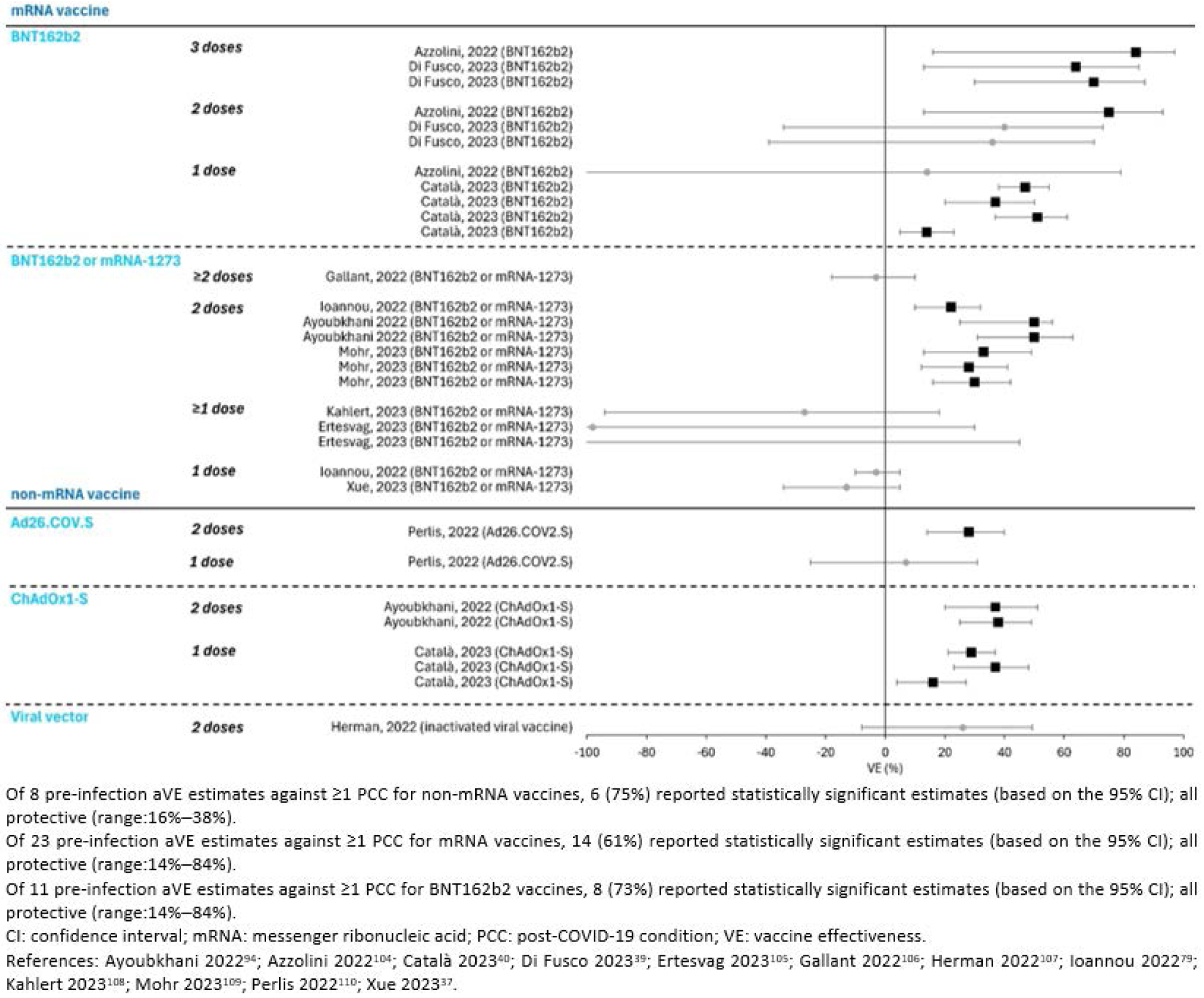
Adjusted Pre-infection VE against ≥1 PCC (vs. Unvaccinated) for mRNA and non-mRNA vaccines, by Vaccine Type, Manufacturer and Dose.

Overall, 77% of pre-infection aVE estimates (vs. unvaccinated) were statistically significant (aVE range: 7%– 95%; **Figures 2 and 3**). Similarly, over half of the aVE estimates that were not statistically significant (n=9/21) suggested a protective effect of pre-infection vaccination. At other timepoints, 80% of estimates reflecting a mix of those vaccinated before and after infection were statistically significant (aVE range: 62%–73%), 43% of estimates reflecting post-infection vaccination were statistically significant (two were protective [aVE range: 28%–40%] and one was not [aVE=-47%]), and 20% of estimates assessing vaccination after PCC onset were statistically significant (aVE=41%). Of estimates not specifying vaccination timing, 46% were statistically significant (23 estimates were protective [aVE range: 29%–75%] and one was not [aVE=-132%]) (**eFigure 3**).

Due to the number of studies focusing on vaccination before infection relative to other timeframes, aVE comparisons across subgroups were only possible for studies where vaccination occurred before infection.

### Adjusted VE against ≥1 PCC when vaccinated prior to an infection By number of doses

Statistically significant aVE estimates (vs. unvaccinated) varied by the number of vaccine doses received before infection. For example, 18 of 25 estimates for one dose were statistically significant (aVE range: 12%– 82%), 9 of 14 estimates for ≥1 dose were statistically significant (aVE range: 46%–86%), 36 of 41 estimates for 2 doses or primary series completion were statistically significant (aVE range: 7%–90%), 8 of 10 estimates for receipt of 3 doses or a booster were statistically significant (aVE range: 13%–84%), and the only estimate for 4 doses was statistically significant (aVE=95%). Additionally, all three studies that compared complete vs. incomplete vaccination against ≥1 PCC reported greater protection with complete vaccination^35–37^ and all three studies comparing the protection offered by a booster dose (relative to the primary series or <3 doses) reported no statistically significant difference (**eFigure 1; eTable 8**).^35,38,39^

### By vaccine manufacturer/platform and number of doses

Statistically significant estimates were slightly higher for mRNA vaccines (range: 14%–84%; n=14/23 estimates from 10 different studies were statistically significant) than non-mRNA vaccines (range: 16%–38%; n=6/8 estimates from 4 unique studies were statistically significant). For mRNA-specific estimates, aVE appeared to increase with additional doses. For example, 4 of 7 mRNA-specific estimates for 1 dose were statistically significant (aVE range: 14%–51%; 4 of 5 BNT162b2-specific estimates from 3 unique studies were statistically significant), 7 of 9 mRNA-specific estimates for 2 doses were statistically significant (aVE range: 22%-75%) and 1 of 3 BNT162b2-specific estimates for 2 doses was statistically significant (aVE=75%), and all 3 estimates for 3 doses were statistically significant (aVE range: 64%–84%; all BNT162b2) (**Figure 2; eTable 14 and 15**). Although data are more limited, estimates that directly compare the effectiveness of one or two vaccine doses (vs. unvaccinated) against ≥1 PCC for mRNA vs. non-mRNA vaccines suggest significantly greater protection from mRNA vaccines than other vaccine types (**eFigure 1; eTable 8**).^40,41^

### By variant

The range of statistically significant aVE overlapped for infections pre-Omicron (aVE range: 7%–50%; n=18/25 estimates from 10 studies were statistically significant) and during Omicron (aVE range: 10%–70%; n=12/15 estimates from 4 studies were statistically significant). For estimates reflecting infections during both Omicron and pre-Omicron periods, 41 of 52 estimates from 16 studies were statistically significant (aVE range: 12–95%) (**Figure 4; eTable 16**).

**Figure 4.**
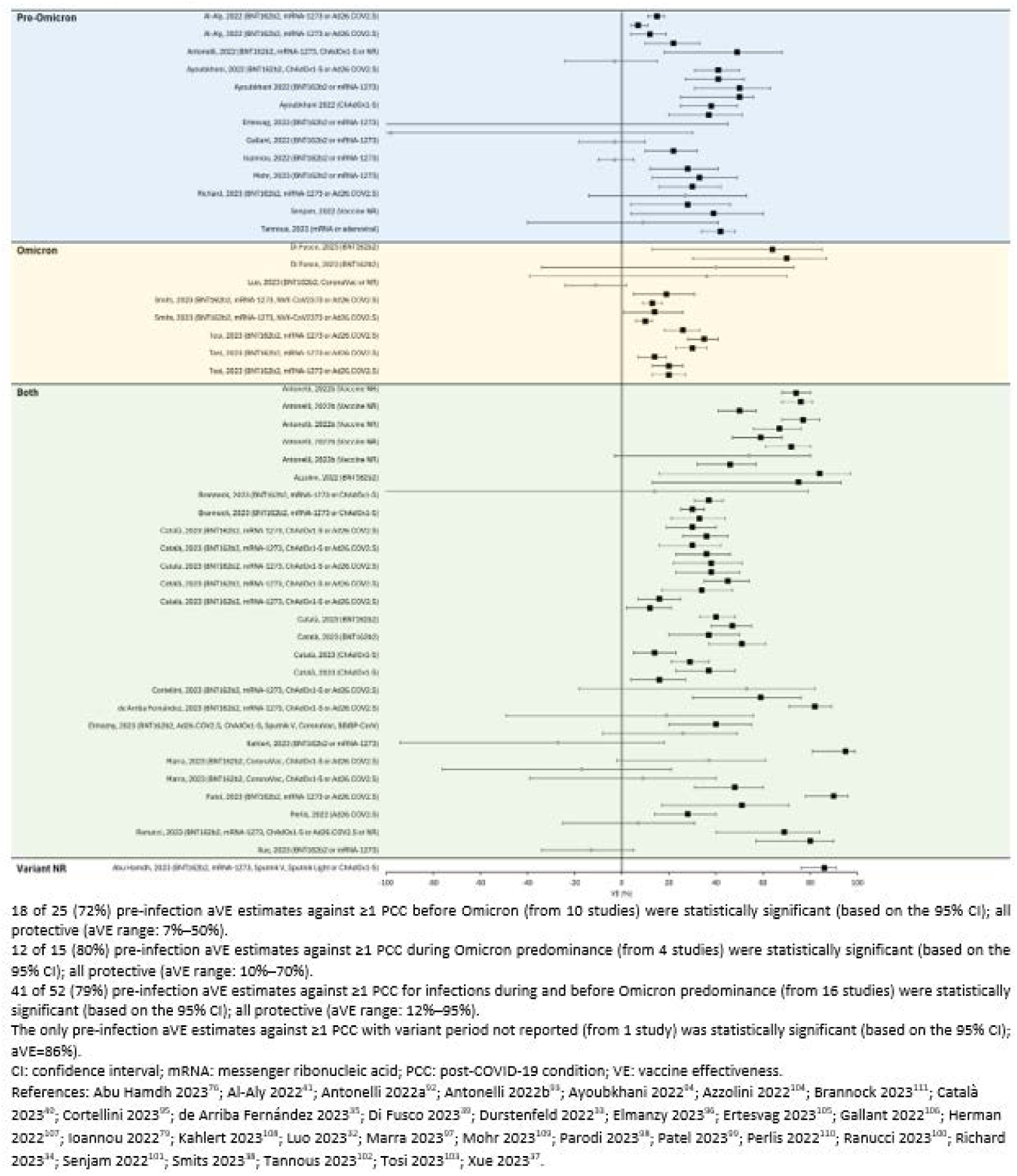
Adjusted Pre-infection VE against ≥1 PCC (vs.Unvaccinated), by Predominant Variant Circulating During Scute Infection.

### By PCC definition

Overall, more studies used patient-reported PCC (n=19) than medical codes/claims (n=11); one study used a combination of PCC based on patient-report and claims/codes. Estimates based on claims/codes were more likely to be statistically significant (89% vs. 67%), but the range of statistically significant aVE estimates overlapped for PCC based on medical/claims codes (range: 7%–95%; n=41/46 estimates statistically significant) and patient-report (range: 28%–90%; n=31/46 estimates statistically significant). Estimates based on patient-reported PCC (and particularly those reported at one-month post-infection) tended to be higher but were often less precise due to smaller sample sizes (**Figure 5; eTable 17 and 18**). Slightly lower aVEs were observed for time-to-event analyses (longer follow-up periods and codes/claims-based PCC), which typically have larger sample sizes and more precise estimates (**Figure 5**).

**Figure 5.**
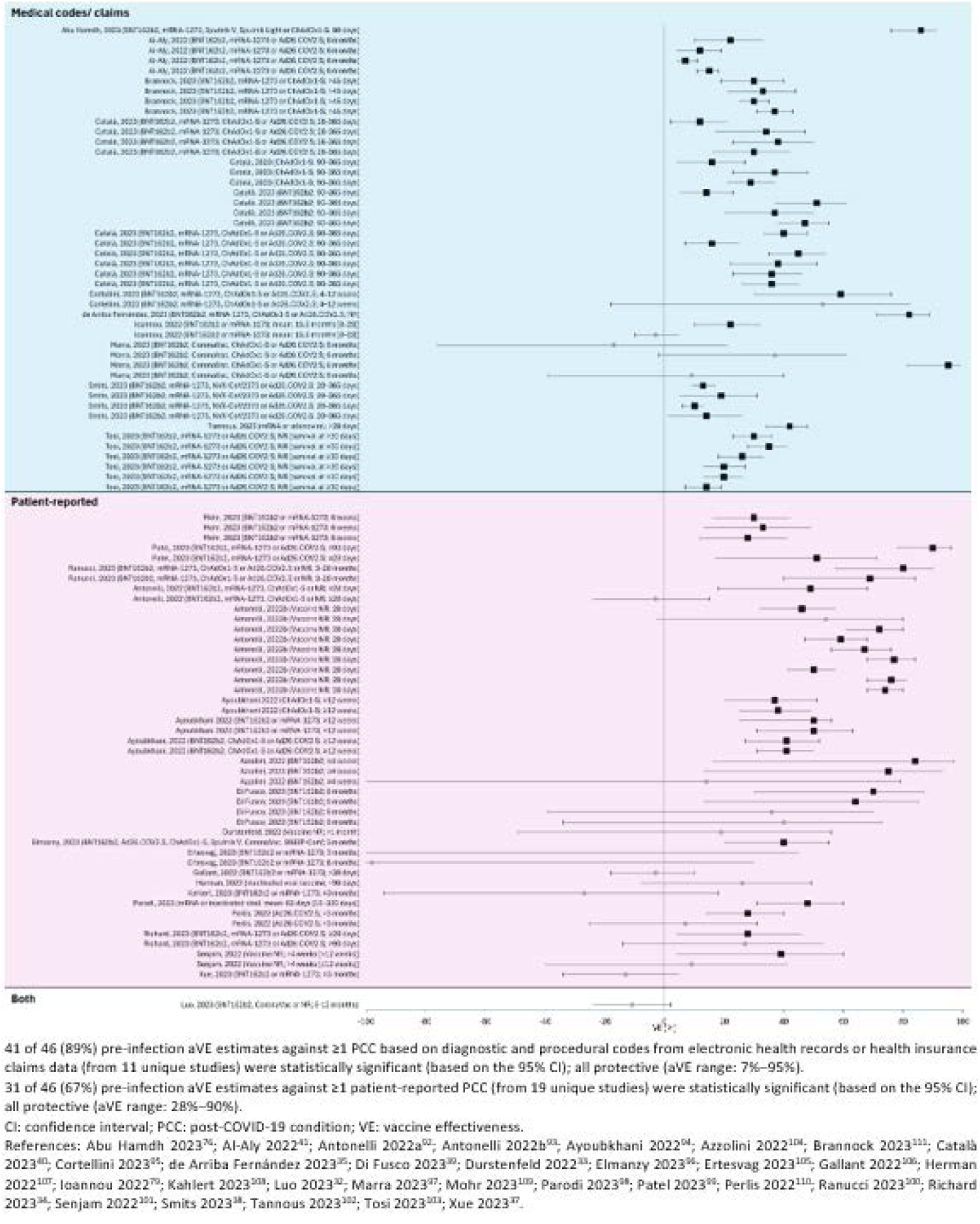
Adjusted Pre-infection VE against ≥1 PCC (vs. Unvaccinated), by PCC data source.

### By proportion of sample hospitalized during acute infection

Pre-infection vaccination was protective against PCC in studies including a mix of patients who were and were not hospitalized during the acute phase (aVE range: 15%–90%; n=17/24 estimates statistically significant), among those hospitalized during the acute phase (aVE range: 12%–80%; n=5/5 estimates statistically significant), and among those not hospitalized during the acute phase (aVE range: 7%–84%; n=3/6 estimates statistically significant) (**eFigure 4; eTable 19 and 20**).

### By other sub-groups and other outcomes

Due to the limited number of studies reporting time since vaccination (**eTable 21**) and inclusion vs. exclusion of reinfections (**eFigure 5; eTable 22**), no trends were detected. The range of aVE against specific cardiovascular, musculoskeletal, neurological, or endocrine symptoms/conditions (**eFigure 2; eTable 9**) were consistent with the aVE range reported against ≥1 PCC. Other outcomes where VE could not be computed are summarized in **eTable 10**. For example, Strain and colleagues reported that those vaccinated post-PCC onset with mRNA vs. non-mRNA vaccines experienced greater symptom improvement.^42^ Tran and colleagues reported that those vaccinated after infection (vs. unvaccinated) were less likely to report an unacceptable symptom state, more likely to achieve full remission, and experienced less severe long COVID (lower impact on patient quality of life).^43^ Nehme and colleagues reported a higher PCC symptom prevalence among those who received ≥1 dose before infection (vs. unvaccinated).^44^ Taquet and colleagues reported significant differences in the risk of some specific PCC as composite endpoints with death for those vaccinated with 1 dose vs. unvaccinated and 2 doses vs. unvaccinated.^45^

## Discussion

Despite heterogeneity across studies included in this SLR with respect to study design, population, PCC definition, follow-up period and data source, definition of ‘vaccinated’, and predominant variant circulating, there is consistent evidence suggesting that pre-infection COVID-19 vaccination reduces the risk of ≥1 PCC. Regardless of the number of doses or vaccine type received, 77% of adjusted pre-infection VE estimates against ≥1 PCC (vs. unvaccinated) were statistically significant and all statistically significant findings were protective (range: 7%–95%). Evidence was limited for vaccination at other timepoints relative to infection or PCC onset. Collectively, these findings are consistent with guidance provided by the Centers for Disease Control and Prevention^46^, International Coalition of Medicines Regulatory Authorities^47^, World Health Organization^48^, European Medicines Agency^49^ and United Kingdom Health Security Agency^50^.

Among studies where vaccination occurred before the index infection, aVE estimates varied by vaccine type and the number of doses received. For example, the range of protection offered by vaccination (based on comparisons across strata or direct comparisons) was slightly higher for mRNA vaccines than other vaccine types. This is consistent with other studies that reported higher VEs for mRNA than non-mRNA vaccines based on direct comparisons^51,52^ or differences across strata based on vaccine type or manufacturer.^35,53–57^ We also observed a dose response for mRNA-specific aVEs, whereby additional doses offered greater protection (i.e., VE range for 1 dose: 14%–51%; 2 doses: 22%–75%; and 3 doses: 64%–84%); however, only 10 studies reported mRNA-specific estimates. Other studies (including those published after this SLR’s search period) have reported higher VEs for additional doses, regardless of vaccine type.^18,51,52,58–66^ Further, two studies specifically showed the benefit of additional doses during Omicron predominance (4 vs. 3 doses,^67^ and 3 vs. 2 doses).^68^ Additionally, more recently published mRNA-specific and BNT162b2-specific VEs reported by dose fall within the range of those summarized in this SLR (2 mRNA doses vs. unvaccinated [VE=57%],^69^ ≥2 BNT162b2 doses vs. unvaccinated [VE=78%],^54^ 2 doses BNT162b2 vs. unvaccinated [VE=51%],^65^ 2 doses mRNA-1273 vs. unvaccinated (VE=52%),^65^ and vaccinated with BNT162b2 bivalent vs. unvaccinated or not up-to-date [VE range=41%–43%].^70^

Other studies have reported a lower PCC burden during Omicron predominance,^66,71^ however, directly comparing VE estimates across different variant periods is challenging due to other time-varying factors which may impact the likelihood of experiencing PCC.^72^ We present the range of pre-infection aVE against ≥1 PCC for those infected (1) during Omicron predominance and (2) when other variants predominated. The aVE range was slightly higher during periods of Omicron predominance and a greater proportion of estimates were statistically significant; however there were fewer studies conducted during Omicron than periods when other variants predominated. Our findings suggest that pre-infection vaccination offers protection against PCC regardless of the variant causing infection and is consistent with findings reported by Henderson and colleagues^51^ but not Lundberg-Morris and colleagues.^65^ Our findings are also consistent with those demonstrating that 70%–76% of the reduced PCC burden from pre-Omicron to Omicron variant periods was attributed to COVID-19 vaccinations.^66^

Woodrow and colleagues previously reported higher estimates for PCC based on self-report vs. health records.^73^ Consistent with this finding, we observed the highest aVE estimates for PCC at one month post-infection (all patient-reported PCC). Although there is the potential for PCC to be misclassified in both data sources, the consistent trends observed across studies with different PCC data sources suggests that pre-infection COVID-19 vaccination likely reduces the risk of PCC.

Although the PCC burden has been reported to be higher among those hospitalized during the acute phase ^2,34,59,66,74–83^, we did not observe differences in the range of pre-infection aVE against ≥1 PCC based on the proportion of the sample hospitalized during the acute phase. This may reflect (1) heterogeneity in other study design characteristics (e.g., predominant variant, number of vaccine doses, PCC definition, PCC data source), (2) survival bias (those experiencing PCC must have survived to experience PCC), and (3) heterogeneity in disease severity among those hospitalized i.e. admitted to the intensive care unit, requiring supplemental oxygen or mechanical ventilation, etc.). Other SLRs have similarly reported heterogeneity according to PCC definitions and study design.^84^

Findings from this SLR should be interpreted with caution given the heterogeneous study designs, data sources, and definitions for vaccinated and PCC. Given that many studies in this review did not report the time between vaccination and infection, we were unable to assess the impact of time since vaccination on VE against ≥1 PCC. It was consequently difficult to tease apart the impact of changing variants and time since last dose. For example, of 93 aVE estimates against ≥1 PCC (vs. unvaccinated), the majority (86%) operationalized ‘vaccinated’ as receipt of one dose (n=25), ≥1 dose (n=14), or 2 doses or primary series completion (n=41). If VE against PCC wanes over time, as a few studies reported,^65,85,86^ the wide range of aVEs observed for receipt of 2 doses or the primary series (7%–90%) could be explained by varying lengths of time since vaccination. Similarly, there may be heterogeneity in multi-year studies where receipt of the primary series in 2021 conveys a different level of protection than completion of the primary series in 2023. Further, the dose response observed in some studies may reflect more time since primary series completion than a booster dose, for example. Few studies have evaluated the protection offered by adapted vaccines (i.e., bivalent, XBB.1.5, and JN.1) and most lack of information on prior SARS-CoV-2 infections (including time since last infection and variant causing last infection) which can create an imbalance in the immune histories of the comparison groups and has been shown to impact VE against infection^87,88^ and the likelihood of PCC^67^. SLR/meta-analyses^89–91^ have also shown that those treated with antivirals during the acute phase have a reduced risk of PCC; however, antiviral use during the acute phase was not documented or accounted for in the studies summarized here, further contributing to the heterogeneity across studies.

## Conclusion

Our findings provide a comprehensive synthesis of aVE estimates for COVID-19 vaccination against PCC and describe trends in the effectiveness of COVID-19 vaccines for preventing PCC across key factors known to impact the risk of PCC and the effectiveness of vaccines against other COVID-19 endpoints (e.g., acute infection severity, PCC definition and data source, type and number of vaccine doses, and predominant variant). Collectively our findings suggest that COVID-19 vaccination received prior to SARS-CoV-2 infection reduces the subsequent risk of developing PCC regardless of the predominant variant circulating.

## Data Availability

All data supporting the findings of this study are available within the paper and its Supplementary Information.

## Author Contribution Statement

All authors were involved in study conception and design, material preparation, data collection and analysis. The first draft of the manuscript was written by Abby E Rudolph, Isabelle Whittle, Olivia Wright, and Kristen Markus, and all authors provided comments on subsequent drafts. All authors read and approved the final manuscript.

## References

1. World Health Organization. WHO COVID-19 dashboard: Number of COVID-19 cases reported to WHO. Accessed September 11, 2024. https://data.who.int/dashboards/covid19/cases?n=c

2. Davis HE, McCorkell L, Vogel JM, Topol EJ. Long COVID: major findings, mechanisms and recommendations. Nature Reviews Microbiology . 2023/03/01 2023;21(3):133-146. doi:10.1038/s41579-022-00846-2

3. Chen C, Haupert SR, Zimmermann L, Shi X, Fritsche LG, Mukherjee B. Global Prevalence of Post-Coronavirus Disease 2019 (COVID-19) Condition or Long COVID: A Meta-Analysis and Systematic Review. J Infect Dis. Nov 1 2022;226(9):1593–1607. doi:10.1093/infdis/jiac136

4. Yang J, Markus K, Andersen KM, et al. Definition and measurement of post-COVID-19 conditions in real-world practice: a global systematic literature review. BMJ Open . 2024;14(1):e077886. doi:10.1136/bmjopen-2023-077886

5. National Academies of Sciences E, Medicine.,. A Long COVID Definition: A Chronic, Systemic Disease State with Profound Consequences. 2024;

6. Al-Aly Z, Davis H, McCorkell L, et al. Long COVID science, research and policy. Nat Med . Aug 2024;30(8):2148–2164. doi:10.1038/s41591-024-03173-6

7. Sagy YW, Feldhamer I, Brammli-Greenberg S, Lavie G. Estimating the economic burden of long-Covid: the additive cost of healthcare utilisation among COVID-19 recoverees in Israel. BMJ Global Health . 2023;8(7):e012588.

8. Mirin AA. A preliminary estimate of the economic impact of long COVID in the United States. *Fatigue: Biomedicine*, Health & Behavior. 2022;10(4):190–199.

9. Cutler DM. The Costs of Long COVID. JAMA Health Forum. May 6 2022;3(5):e221809. doi:10.1001/jamahealthforum.2022.1809

10. Mohammed I, Nauman A, Paul P, et al. The efficacy and effectiveness of the COVID-19 vaccines in reducing infection, severity, hospitalization, and mortality: a systematic review. Hum Vaccin Immunother . 2022/01/31 2022;18(1):2027160. doi:10.1080/21645515.2022.2027160

11. Feikin DR, Higdon MM, Abu-Raddad LJ, et al. Duration of effectiveness of vaccines against SARS-CoV- 2 infection and COVID-19 disease: results of a systematic review and meta-regression. Lancet. Mar 5 2022;399(10328):924–944. doi:10.1016/s0140-6736(22)00152-0

12. Lin D-Y, Xu Y, Gu Y, et al. Effects of COVID-19 vaccination and previous SARS-CoV-2 infection on omicron infection and severe outcomes in children under 12 years of age in the USA: an observational cohort study. The Lancet Infectious Diseases. 2023;

13. Rudolph AE, Khan FL, Shah A, et al. Effectiveness of BNT162b2 BA. 4/5 bivalent mRNA vaccine against symptomatic COVID-19 among immunocompetent individuals testing at a large US retail pharmacy. The Journal of Infectious Diseases. 2023:jiad474.

14. Link-Gelles R. Effectiveness of bivalent mRNA vaccines in preventing symptomatic SARS-CoV-2 infection—increasing community access to testing program, United States, September–November 2022. MMWR Morbidity and Mortality Weekly Report. 2022;71

15. Gao P, Liu J, Liu M. Effect of COVID-19 Vaccines on Reducing the Risk of Long COVID in the Real World: A Systematic Review and Meta-Analysis. Int J Environ Res Public Health. Sep 29 2022;19(19)doi:10.3390/ijerph191912422

16. Hamad Saied M, van der Griend L, van Straalen JW, Wulffraat NM, Vastert S, Jansen MHA. The protective effect of COVID-19 vaccines on developing multisystem inflammatory syndrome in children (MIS- C): a systematic literature review and meta-analysis. Pediatr Rheumatol Online J. Aug 7 2023;21(1):80. doi:10.1186/s12969-023-00848-1

17. Man MA, Rosca D, Bratosin F, et al. Impact of Pre-Infection COVID-19 Vaccination on the Incidence and Severity of Post-COVID Syndrome: A Systematic Review and Meta-Analysis. Vaccines. 2024;12(2):189.

18. Marra AR, Kobayashi T, Suzuki H, et al. The effectiveness of coronavirus disease 2019 (COVID-19) vaccine in the prevention of post-COVID-19 conditions: A systematic literature review and meta-analysis. Antimicrob Steward Healthc Epidemiol. 2022;2(1):e192. doi:10.1017/ash.2022.336

19. Mumtaz A, Sheikh AAE, Khan AM, et al. COVID-19 Vaccine and Long COVID: A Scoping Review. Life (Basel). Jul 16 2022;12(7)doi:10.3390/life12071066

20. Notarte KI, Catahay JA, Velasco JV, et al. Impact of COVID-19 vaccination on the risk of developing long-COVID and on existing long-COVID symptoms: A systematic review. eClinicalMedicine. November 2022;53 (no pagination)101624. doi:10.1016/j.eclinm.2022.101624

21. Tsampasian V, Elghazaly H, Chattopadhyay R, et al. Risk Factors Associated With Post-COVID-19 Condition: A Systematic Review and Meta-analysis. JAMA Intern Med. Jun 1 2023;183(6):566–580. doi:10.1001/jamainternmed.2023.0750

22. Watanabe A, Iwagami M, Yasuhara J, Takagi H, Kuno T. Protective effect of COVID-19 vaccination against long COVID syndrome: A systematic review and meta-analysis. Vaccine. Mar 10 2023;41(11):1783–1790. doi:10.1016/j.vaccine.2023.02.008

23. Marra AR, Kobayashi T, Callado GY, et al. The effectiveness of COVID-19 vaccine in the prevention of post-COVID conditions: a systematic literature review and meta-analysis of the latest research. Antimicrob Steward Healthc Epidemiol. 2023;3(1):e168. doi:10.1017/ash.2023.447

24. National Institute for Health and Care Excellence. COVID-19 rapid guideline: managing the longterm effects of COVID-19. Accessed April 2023, 2023. https://www.nice.org.uk/guidance/ng188/resources/covid19-rapid-guideline-managing-the-longterm-effects-of-covid19-pdf-51035515742

25. Li J, Zhou Y, Ma J, et al. The long-term health outcomes, pathophysiological mechanisms and multidisciplinary management of long COVID. Signal Transduct Target Ther. Nov 1 2023;8(1):416. doi:10.1038/s41392-023-01640-z

26. Byambasuren O, Stehlik P, Clark J, Alcorn K, Glasziou P. Effect of covid-19 vaccination on long covid: systematic review. BMJ Med. 2023;2(1):e000385. doi:10.1136/bmjmed-2022-000385

27. Jennings S, Corrin T, Waddell L. A systematic review of the evidence on the associations and safety of COVID-19 vaccination and post COVID-19 condition. Epidemiol Infect. Aug 18 2023;151:e145. doi:10.1017/s0950268823001279

28. Higgins JPT TJ, Chandler J, Cumpston M, Li T, Page MJ, Welch VA,. Cochrane Handbook for Systematic Reviews of Interventions version 6.4 (updated August 2023) . Cochrane 2023; 2024. www.training.cochrane.org/handbook

29. Page MJ, McKenzie JE, Bossuyt PM, et al. The PRISMA 2020 statement: an updated guideline for reporting systematic reviews. Bmj. Mar 29 2021;372:n71. doi:10.1136/bmj.n71

30. Joanna Briggs Institute. Critical Appraisal Tools. Accessed February 9, 2024. https://jbi.global/critical-appraisal-tools

31. CoVariants. Overview of Variants in Countries. 2024. https://covariants.org/per-country

32. Luo JZ, J. Tang, H. T. Wong, H. K. Lyu, A. Cheung, C. H. Bian, Z. Prevalence and risk factors of long COVID 6-12 months after infection with the Omicron variant among nonhospitalized patients in Hong Kong. Journal of Medical Virology. June 2023;95(6) (no pagination)e28862. doi:10.1002/jmv.28862

33. Durstenfeld MS, Peluso MJ, Peyser ND, et al. Factors Associated with Long COVID Symptoms in an Online Cohort Study. Open Forum Infectious Diseases. 01 Feb 2023;10(2) (no pagination)ofad047. doi:10.1093/ofid/ofad047

34. Richard SA, Pollett SD, Fries AC, et al. Persistent COVID-19 Symptoms at 6 Months after Onset and the Role of Vaccination before or after SARS-CoV-2 Infection. JAMA Network Open. 18 Jan 2023;6(1):E2251360. doi:10.1001/jamanetworkopen.2022.51360

35. de Arriba Fernandez AB, J. L. A. Frances, A. E. Mora, A. C. Perez, A. G. Barreiros, M. Ad. Evaluation of persistent COVID and SARS-CoV-2 reinfection in a cohort of patients on the island of Gran Canaria, Spain. Semergen. 25 Jan 2023;49(5):101939. doi:10.1016/j.semerg.2023.101939

36. Nascimento TCDCdVC, L. Ruiz, A. D. Ledo, C. B. Fernandes, V. P. L. Cardoso, L. F. Junior, J. M. V. Saretta, R. Kalil-Filho, R. Drager, L. F. Vaccination status and long COVID symptoms in patients discharged from hospital. Scientific reports. 11 Feb 2023;13(1):2481. doi:10.1038/s41598-023-28839-y

37. Xue P, Merikanto I, Chung F, et al. Persistent short nighttime sleep duration is associated with a greater post-COVID risk in fully mRNA-vaccinated individuals. Translational Psychiatry. December 2023;13(1) (no pagination)32. doi:10.1038/s41398-023-02334-4

38. Smits PD, Rodriguez PJ, Gratzl S, et al. Effect of vaccination on time till Long COVID, a comparison of two ways to model effect of vaccination and two outcome definitions. medRxiv. 2023:2023.04.28.23289271. doi:10.1101/2023.04.28.23289271

39. Di Fusco M, Sun X, Moran MM, et al. Impact of COVID-19 and effects of booster vaccination with BNT162b2 on six-month long COVID symptoms, quality of life, work productivity and activity impairment during Omicron. Journal of Patient-Reported Outcomes. 2023/07/24 2023;7(1):77. doi:10.1186/s41687-023-00616-5

40. Català M, Mercadé-Besora N, Kolde R, et al. The Effectiveness of COVID-19 Vaccines to Prevent Long COVID Symptoms: Staggered Cohort Analyses of Data from the UK, Spain, and Estonia. SSRN. 2023;doi:10.2139/ssrn.4474215

41. Al-Aly Z, Bowe B, Xie Y. Long COVID after breakthrough SARS-CoV-2 infection. Nature Medicine. 2022/07/01 2022;28(7):1461-1467. doi:10.1038/s41591-022-01840-0

42. Strain WDS, O. Banerjee, A. Van der Togt, V. Hishmeh, L. Rossman, J. The Impact of COVID Vaccination on Symptoms of Long COVID: An International Survey of People with Lived Experience of Long COVID. Vaccines. May 2022;10(5) (no pagination)652. doi:10.3390/vaccines10050652

43. Tran VTP, E. Saldanha, J. Pane, I. Ravaud, P. Efficacy of first dose of covid-19 vaccine versus no vaccination on symptoms of patients with long covid: target trial emulation based on ComPaRe e-cohort. BMJ Med. 2023;2(1):e000229. doi:10.1136/bmjmed-2022-000229

44. Nehme M, Vetter P, Chappuis F, Kaiser L, Guessous I. Prevalence of Post-Coronavirus Disease Condition 12 Weeks After Omicron Infection Compared With Negative Controls and Association With Vaccination Status. Clin Infect Dis. May 3 2023;76(9):1567–1575. doi:10.1093/cid/ciac947

45. Taquet MD, Q. Harrison, P. J. Six-month sequelae of post-vaccination SARS-CoV-2 infection: A retrospective cohort study of 10,024 breakthrough infections. Brain, Behavior, and Immunity. July 2022;103:154–162. doi:10.1016/j.bbi.2022.04.013

46. Centers for Disease Control and Prevention. Long COVID or Post-COVID Conditions. Accessed February 9, 2024. https://www.cdc.gov/coronavirus/2019-ncov/long-term-effects/index.html#:~:text=Long%20COVID%20is%20broadly%20defined,after%20acute%20COVID%2D19%20infection.

47. Authorities ICoMR. ICMRA statement on the safety of COVID-19 vaccines. Accessed 13 August, 2024. https://www.icmra.info/drupal/strategicinitiatives/vaccines/safety_statement

48. World Health Organization. Coronavirus disease (COVID-19): Post COVID-19 condition. Accessed 13 August, 2024. https://www.who.int/news-room/questions-and-answers/item/coronavirus-disease-(covid-19)-post-covid-19-condition

49. European Medicines Agency. COVID-19 vaccines: key facts. Accessed 13 August, 2024. https://www.ema.europa.eu/en/human-regulatory-overview/public-health-threats/coronavirus-disease-covid-19/covid-19-medicines/covid-19-vaccines-key-facts#vaccine-safety-7220

50. UK Health Security Agency. UKHSA review shows vaccinated less likely to have long COVID than unvaccinated. Accessed 13 August, 2024. https://www.gov.uk/government/news/ukhsa-review-shows-vaccinated-less-likely-to-have-long-covid-than-unvaccinated

51. Henderson AD, Butler-Cole BF, Tazare J, et al. Clinical coding of long COVID in primary care 2020- 2023 in a cohort of 19 million adults: an OpenSAFELY analysis. EClinicalMedicine. Jun 2024;72:102638. doi:10.1016/j.eclinm.2024.102638

52. Song J, Choi S, Jeong S, et al. Protective effect of vaccination on the risk of cardiovascular disease after SARS-CoV-2 infection. Clin Res Cardiol. Jul 31 2024;doi:10.1007/s00392-023-02271-8

53. Català M, Mercadé-Besora N, Kolde R, et al. The effectiveness of COVID-19 vaccines to prevent long COVID symptoms: staggered cohort study of data from the UK, Spain, and Estonia. Lancet Respir Med. Mar 2024;12(3):225–236. doi:10.1016/s2213-2600(23)00414-9

54. Wong MCS, Huang J, Wong YY, et al. Epidemiology, Symptomatology, and Risk Factors for Long COVID Symptoms: Population-Based, Multicenter Study. JMIR public health and surveillance. 07 Mar 2023;9:e42315. doi:10.2196/42315

55. Mercadé-Besora N, Li X, Kolde R, et al. The role of COVID-19 vaccines in preventing post-COVID-19 thromboembolic and cardiovascular complications. Heart. Apr 15 2024;110(9):635–643. doi:10.1136/heartjnl-2023-323483

56. de Arriba Fernandez AAB, J. L. Espineira Frances, A. Cabeza Mora, A. Gutierrez Perez, A. Diaz Barreiros, M. A. Serra Majem, L. Assessment of SARS-CoV-2 Infection According to Previous Metabolic Status and Its Association with Mortality and Post-Acute COVID-19. Nutrients. July 2022;14(14) (no pagination)2925. doi:10.3390/nu14142925

57. Ghosh P, Niesen MJM, Pawlowski C, et al. Severe acute infection and chronic pulmonary disease are risk factors for developing post-COVID-19 conditions. medRxiv. Dec 1 2022;doi:10.1101/2022.11.30.22282831

58. Woldegiorgis M, Cadby G, Ngeh S, et al. Long COVID in a highly vaccinated but largely unexposed Australian population following the 2022 SARS-CoV-2 Omicron wave: a cross-sectional survey. Med J Aust. Apr 1 2024;220(6):323–330. doi:10.5694/mja2.52256

59. Wander PL, Baraff A, Fox A, et al. Rates of ICD-10 Code U09.9 Documentation and Clinical Characteristics of VA Patients With Post-COVID-19 Condition. JAMA Netw Open. Dec 1 2023;6(12):e2346783. doi:10.1001/jamanetworkopen.2023.46783

60. Wan EYF, Mok AHY, Yan VKC, et al. Association between BNT162b2 and CoronaVac vaccination and risk of CVD and mortality after COVID-19 infection: A population-based cohort study. Cell Rep Med. Oct 17 2023;4(10):101195. doi:10.1016/j.xcrm.2023.101195

61. Lam ICH, Zhang R, Man KKC, et al. Persistence in risk and effect of COVID-19 vaccination on long-term health consequences after SARS-CoV-2 infection. Nat Commun. Feb 26 2024;15(1):1716. doi:10.1038/s41467-024-45953-1

62. Lim JT, Liang En W, Tay AT, et al. Long-term Cardiovascular, Cerebrovascular, and Other Thrombotic Complications in COVID-19 Survivors: A Retrospective Cohort Study. Clinical Infectious Diseases. 2023;78(1):70–79. doi:10.1093/cid/ciad469

63. Lukowsky LR, Der-Martirosian C, Northcraft H, Kalantar-Zadeh K, Goldfarb DS, Dobalian A. Predictors of Acute Kidney Injury (AKI) among COVID-19 Patients at the US Department of Veterans Affairs: The Important Role of COVID-19 Vaccinations. Vaccines (Basel). Jan 30 2024;12(2)doi:10.3390/vaccines12020146

64. Hsieh TYJ, Chang R, Yong SB, Liao PL, Hung YM, Wei JC. COVID-19 Vaccination Prior to SARS-CoV-2 Infection Reduced Risk of Subsequent Diabetes Mellitus: A Real-World Investigation Using U.S. Electronic Health Records. Diabetes Care. Dec 1 2023;46(12):2193–2200. doi:10.2337/dc23-0936

65. Lundberg-Morris L, Leach S, Xu Y, et al. Covid-19 vaccine effectiveness against post-covid-19 condition among 5891722 individuals in Sweden: population based cohort study. BMJ. 2023;383:e076990. doi:10.1136/bmj-2023-076990

66. Xie Y, Choi T, Al-Aly Z. Postacute sequelae of SARS-CoV-2 infection in the pre-delta, delta, and omicron eras. New England Journal of Medicine. 2024;

67. Mikolajczyk R, Diexer S, Klee B, et al. Likelihood of Post-COVID Condition in people with hybrid immunity; data from the German National Cohort (NAKO). J Infect. Aug 2024;89(2):106206. doi:10.1016/j.jinf.2024.106206

68. Spiliopoulos L, Sørensen AIV, Bager P, et al. Postacute symptoms 4 months after SARS-CoV-2 infection during the Omicron period: a nationwide Danish questionnaire study. Am J Epidemiol. Aug 5 2024;193(8):1106–1114. doi:10.1093/aje/kwad225

69. Saheb Sharif-Askari F, Ali Hussain Alsayed H, Saheb Sharif-Askari N, Saddik B, Al Sayed Hussain A, Halwani R. Risk factors and early preventive measures for long COVID in non-hospitalized patients: analysis of a large cohort in the United Arab Emirates. Public Health. May 2024;230:198–206. doi:10.1016/j.puhe.2024.02.031

70. Di Fusco M, Sun X, Allen KE, et al. Effectiveness of BNT162b2 BA.4/5 Bivalent COVID-19 Vaccine against Long COVID Symptoms: A US Nationwide Study. Vaccines (Basel). Feb 11 2024;12(2)doi:10.3390/vaccines12020183

71. Foulkes S, Evans J, Neill C, et al. Prevalence and impact of persistent symptoms following SARS-CoV-2 infection among healthcare workers: a cross-sectional survey in the SIREN cohort. J Infect. 2024:106259.

72. Gonçalves BP. Re:“Postacute symptoms 4 months after SARS-CoV-2 infection during the Omicron period: a nationwide Danish questionnaire study”. American Journal of Epidemiology. 2024:kwae021.

73. Woodrow M, Carey C, Ziauddeen N, et al. Systematic Review of the Prevalence of Long COVID. Open Forum Infect Dis. Jul 2023;10(7):ofad233. doi:10.1093/ofid/ofad233

74. Mayor N, Meza-Torres B, Okusi C, et al. Developing a Long COVID Phenotype for Postacute COVID-19 in a National Primary Care Sentinel Cohort: Observational Retrospective Database Analysis. JMIR Public Health Surveill. Aug 11 2022;8(8):e36989. doi:10.2196/36989

75. Sudre CH, Murray B, Varsavsky T, et al. Attributes and predictors of long COVID. Nat Med. Apr 2021;27(4):626–631. doi:10.1038/s41591-021-01292-y

76. Abu Hamdh BN, Z. A prospective cohort study assessing the relationship between long-COVID symptom incidence in COVID-19 patients and COVID-19 vaccination. Scientific reports. 25 Mar 2023;13(1):4896. doi:10.1038/s41598-023-30583-2

77. Chilunga FP, Appelman B, van Vugt M, et al. Differences in incidence, nature of symptoms, and duration of long COVID among hospitalised migrant and non-migrant patients in the Netherlands: a retrospective cohort study. The Lancet Regional Health – Europe. 2023;29doi:10.1016/j.lanepe.2023.100630

78. Emecen AN, Keskin S, Turunc O, et al. The presence of symptoms within 6 months after COVID-19: a single-center longitudinal study. Irish Journal of Medical Science. April 2023;192(2):741–750. doi:10.1007/s11845-022-03072-0

79. Ioannou GNB, A. Fox, A. Shahoumian, T. Hickok, A. O’Hare, A. M. Bohnert, A. S. B. Boyko, E. J. Maciejewski, M. L. Bowling, C. B. Viglianti, E. Iwashyna, T. J. Hynes, D. M. Rates and Factors Associated with Documentation of Diagnostic Codes for Long COVID in the National Veterans Affairs Health Care System. JAMA Network Open. 29 Jul 2022;5(7):E2224359. doi:10.1001/jamanetworkopen.2022.24359

80. Karyakarte RP, Das R, Rajmane MV, et al. The Burden and Characteristics of Post-COVID-19 Conditions Among Laboratory-Confirmed Delta and Omicron COVID-19 Cases: A Preliminary Study From Maharashtra, India. Cureus. Sep 2023;15(9):e44888. doi:10.7759/cureus.44888

81. Modji KKS, McCoy KE, Creswell PD, Morris CR, Tomasallo CD. Long COVID Among Wisconsin Workers in the Workers’ Compensation System: Associations With Sociodemographics, Vaccination, and Predominant Variant Period From March 1, 2020 to July 31, 2022. J Occup Environ Med. Feb 1 2024;66(2):e34-e41. doi:10.1097/jom.0000000000003018

82. Pérez-González A, Araújo-Ameijeiras A, Fernández-Villar A, et al. Long COVID in hospitalized and non-hospitalized patients in a large cohort in Northwest Spain, a prospective cohort study. Scientific Reports. 2022/03/01 2022;12(1):3369. doi:10.1038/s41598-022-07414-x

83. Hedberg P, Granath F, Bruchfeld J, et al. Post COVID-19 condition diagnosis: A population-based cohort study of occurrence, associated factors, and healthcare use by severity of acute infection. Journal of Internal Medicine. 2023;293(2):246–258. 10.1111/joim.13584

84. Chaichana U, Man KKC, Chen A, et al. Definition of Post-COVID-19 Condition Among Published Research Studies. JAMA Netw Open. Apr 3 2023;6(4):e235856. doi:10.1001/jamanetworkopen.2023.5856

85. Qin S, Li Y, Wang L, Zhao X, Ma X, Gao GF. Assessment of vaccinations and breakthrough infections after adjustment of the dynamic zero-COVID-19 strategy in China: an online survey. Emerg Microbes Infect. Dec 2023;12(2):2258232. doi:10.1080/22221751.2023.2258232

86. Herman B, Viwattanakulvanid P, Dzulhadj A, Oo AC, Patricia K, Pongpanich S. Effect of full vaccination and post-covid olfactory dysfunction in recovered covid-19 patient. A retrospective longitudinal study with propensity matching. medRxiv. 2022:2022.01.10.22269007. doi:10.1101/2022.01.10.22269007

87. Auvigne V, Tamandjou Tchuem CR, Schaeffer J, Vaux S, Parent Du Chatelet I. Protection against symptomatic SARS-CoV-2 infection conferred by the Pfizer-BioNTech Original/BA.4-5 bivalent vaccine compared to the mRNA Original monovalent vaccines - A matched cohort study in France. Vaccine. Aug 31 2023;41(38):5490-5493. doi:10.1016/j.vaccine.2023.07.071

88. Shrestha NK, Burke PC, Nowacki AS, Simon JF, Hagen A, Gordon SM. Effectiveness of the Coronavirus Disease 2019 Bivalent Vaccine. Open Forum Infect Dis. Jun 2023;10(6):ofad209. doi:10.1093/ofid/ofad209

89. Choi YJ, Seo YB, Seo J-W, et al. Effectiveness of Antiviral Therapy on Long COVID: A Systematic Review and Meta-Analysis. Journal of Clinical Medicine. 2023;12(23):7375.

90. Jiang J, Li Y, Jiang Q, Jiang Y, Qin H, Li Y. Early use of oral antiviral drugs and the risk of post COVID-19 syndrome: A systematic review and network meta-analysis. J Infect. Aug 2024;89(2):106190. doi:10.1016/j.jinf.2024.106190

91. Sun G, Lin K, Ai J, Zhang W. The efficacy of antivirals, corticosteroids, and monoclonal antibodies as acute COVID-19 treatments in reducing the incidence of long COVID: a systematic review and meta-analysis. Clin Microbiol Infect. Jul 14 2024;doi:10.1016/j.cmi.2024.07.006

92. Antonelli M, Penfold RS, Merino J, et al. Risk factors and disease profile of post-vaccination SARS-CoV-2 infection in UK users of the COVID Symptom Study app: a prospective, community-based, nested, case-control study. The Lancet Infectious Diseases. January 2022;22(1):43–55. doi:10.1016/S1473-3099%2821%2900460-6

93. Antonelli M, Pujol JC, Spector TD, Ourselin S, Steves CJ. Risk of long COVID associated with delta versus omicron variants of SARS-CoV-2. The Lancet. 2022;399(10343):2263–2264. doi:10.1016/S0140-6736(22)00941-2

94. Ayoubkhani D, Bosworth ML, King S, et al. Risk of Long COVID in People Infected With Severe Acute Respiratory Syndrome Coronavirus 2 After 2 Doses of a Coronavirus Disease 2019 Vaccine: Community-Based, Matched Cohort Study. Open Forum Infectious Diseases. September 2022;9(9) (no pagination)ofac464. doi:10.1093/ofid/ofac464

95. Cortellini A, Tabernero J, Mukherjee U, et al. SARS-CoV-2 omicron (B.1.1.529)-related COVID-19 sequelae in vaccinated and unvaccinated patients with cancer: results from the OnCovid registry. The Lancet Oncology. April 2023;24(4):335-346. doi:10.1016/S1470-2045%2823%2900056-6

96. Elmazny A, Magdy R, Hussein M, et al. Neuropsychiatric post-acute sequelae of COVID-19: prevalence, severity, and impact of vaccination. European Archives of Psychiatry and Clinical Neuroscience. 2023;doi:10.1007/s00406-023-01557-2

97. Marra AR, Sampaio VS, Ozahata MC, et al. Risk factors for long coronavirus disease 2019 (long COVID) among healthcare personnel, Brazil, 2020-2022. *Infection Control and Hospital Epidemiology* . 2023;doi:10.1017/ice.2023.95

98. Parodi JB, Indavere A, Bobadilla Jacob P, et al. Impact of COVID-19 vaccination in post-COVID cardiac complications. Vaccine. 17 Feb 2023;41(8):1524–1528. doi:10.1016/j.vaccine.2023.01.052

99. Patel NJ, Cook C, Vanni K, et al. Impact of vaccination on postacute sequelae of SARS CoV-2 infection in patients with rheumatic diseases. Annals of the Rheumatic Diseases. 01 Apr 2023;82(4):565–573. doi:10.1136/ard-2022-223439

100. Ranucci M, Baryshnikova E, Anguissola M, et al. The Very Long COVID: Persistence of Symptoms after 12-18 Months from the Onset of Infection and Hospitalization. J Clin Med. Feb 28 2023;12(5)doi:10.3390/jcm12051915

101. Senjam SS, Balhara YPS, Kumar P, et al. A Comprehensive Assessment of Self-Reported Post COVID- 19 Symptoms Among Beneficiaries of Hospital Employee Scheme at a Tertiary Healthcare Institution in Northern India. Int J Gen Med. 2022;15:7355–7372. doi:10.2147/ijgm.S381070

102. Tannous J, Pan AP, Potter T, et al. Real-world effectiveness of COVID-19 vaccines and anti-SARS-CoV- 2 monoclonal antibodies against postacute sequelae of SARS-CoV-2: Analysis of a COVID-19 observational registry for a diverse US metropolitan population. BMJ Open. 05 Apr 2023;13(4) (no pagination)e067611. doi:10.1136/bmjopen-2022-067611

103. Tosi DT, F.Brito, Y.Sarasua, A.Ruiz, J.Hammel, I. S. Vaccination as a protective factor for PASC in Frail, Pre-frail, and Robust Veterans During the Omicron Wave. Conference Abstract. Journal of the American Geriatrics Society. April 2023;71(Supplement 1):S158–S159. doi:10.1111/jgs.18336

104. Azzolini E, Levi R, Sarti R, et al. Association Between BNT162b2 Vaccination and Long COVID After Infections Not Requiring Hospitalization in Health Care Workers. Jama. Aug 16 2022;328(7):676–678. doi:10.1001/jama.2022.11691

105. Ertesvag NU, Iversen A, Blomberg B, et al. Post COVID-19 condition after delta infection and omicron reinfection in children and adolescents. eBioMedicine. June 2023;92 (no pagination)104599. doi:10.1016/j.ebiom.2023.104599

106. Gallant MR-P, C.Lemaire-Paquette, S.Piche, A. SARS-CoV-2 infection outcomes associated with the Delta variant: A prospective cohort study. Jammi. 2022;8(1):49–56. doi:10.3138/jammi-2022-0022

107. Herman BW, M. C. S. Viwattanakulvanid, P. Vaccination status, favipiravir, and micronutrient supplementation roles in post-COVID symptoms: A longitudinal study. PLoS ONE. July 2022;17(7 July) (no pagination)e0271385. doi:10.1371/journal.pone.0271385

108. Kahlert CR, Strahm C, Güsewell S, et al. Post-Acute Sequelae After Severe Acute Respiratory Syndrome Coronavirus 2 Infection by Viral Variant and Vaccination Status: A Multicenter Cross-Sectional Study. Clinical Infectious Diseases. 2023;77(2):194–202. doi:10.1093/cid/ciad143

109. Mohr NM, Plumb ID, Harland KK, et al. Presence of symptoms 6 weeks after COVID-19 among vaccinated and unvaccinated US healthcare personnel: a prospective cohort study. BMJ Open. 2023;13(2):e063141. doi:10.1136/bmjopen-2022-063141

110. Perlis RH, Santillana M, Ognyanova K, et al. Prevalence and Correlates of Long COVID Symptoms Among US Adults. JAMA Network Open. 27 Oct 2022;5(10):E2238804. doi:10.1001/jamanetworkopen.2022.38804

111. Brannock MD, Chew RF, Preiss AJ, et al. Long COVID Risk and Pre-COVID Vaccination: An EHR-Based Cohort Study from the RECOVER Program. Preprint. medRxiv. Oct 07 2022;07:07. doi:10.1101/2022.10.06.22280795

