## Supplementary Materials for "Factors affecting the impact of COVID-19 vaccination on post COVID-19 conditions among adults: A Systematic Literature Review"

**eTable 1. Embase, Medline and EBM reviews search strategies**

| **Embase search strategy** | | | | | | | |
| --- | --- | --- | --- | --- | --- | --- | --- |
| **#** | | **Description** | **Searches** | | **Results** | | |
| 1 | | ***Long COVID terms*** | ((long or post or persist* or sequela* or complication) adj5 (COVID* coronavirus* or SARS*)).ti,ab. | | 4205 | | |
| 2 | |  | (longCOVID or long COVID or long-COVID or long haul COVID or long-haul COVID or post-acute COVID syndrome or persistent COVID-19 or post-acute COVID19 syndrome or long hauler COVID or post-acute sequelae of SARS-COV-2 infection or chronic COVID syndrome or post COVID-19 syndrome or post COVID-19 condition or post-acute sequelae of COVID).ti,ab. | | 4350 | | |
| 3 | |  | exp long COVID/ | | 4323 | | |
| 4 | |  | or/1-3 | | 9451 | | |
| 5 | | ***Vaccine terms*** | exp SARS-CoV-2 vaccine/ or exp vaccine/ | | 410900 | | |
| 6 | |  | (COVID-19 adj2 (vaccine$ or vaccination$ or innoculation$)).ti,ab. | | 28587 | | |
| 7 | |  | 5 or 6 | | 417089 | | |
| 8 | | ***Outcome terms*** | (symptom$ or post-acute outcomes or long term outcomes or (((long term or persisten$) adj3 symptom) or outcome)).ti,ab. | | 3780633 | | |
| 9 | |  | (persistence or prevalence or incidence or epidemiology).ti,ab. | | 2612260 | | |
| 10 | |  | (symptom duration or symptom resolution).ti,ab. | | 11428 | | |
| 11 | |  | symptom/ | | 162312 | | |
| 12 | |  | (((protect$ or risk) adj2 factor) or protective factor or risk factor).ti,ab,kw. | | 429231 | | |
| 13 | |  | or/8-12 | | 6135128 | | |
| 14 | | ***Total*** | 4 and 7 and 13 | | 695 | | |
| **Medline search strategy** | | | | | | | |
| **#** | **Description** | | **Searches** | | | **Results** | |
| 1 | ***Long COVID terms*** | | ((long or post or persist* or sequela* or complication) adj5 (COVID* coronavirus* or SARS*)).ti,ab. | | | 3120 | |
| 2 |  |  | (longCOVID or long COVID or long-COVID or long haul COVID or long-haul COVID or post-acute COVID syndrome or persistent COVID-19 or post-acute COVID19 syndrome or long hauler COVID or post-acute sequelae of SARS-COV-2 infection or chronic COVID syndrome or post COVID-19 syndrome or post COVID-19 condition or post-acute sequelae of COVID).ti,ab. | | | 3344 | |
| 3 |  |  | exp Post-Acute COVID-19 Syndrome/ | | | 1986 | |
| 4 |  |  | or/1-3 | | | 6250 | |
| 5 | ***Vaccine terms*** | | COVID-19 Vaccines/ or exp vaccines/ | | | 275478 | |
| 6 |  |  | (COVID-19 adj2 (vaccine$ or vaccination$ or innoculation$)).ti,ab. | | | 23298 | |
| 7 |  |  | 5 or 6 | | | 284832 | |
| 8 | ***Outcome terms*** | | (symptom$ or post-acute outcomes or long term outcomes or (((long term or persisten$) adj3 symptom) or outcome)).ti,ab. | | | 2514896 | |
| 9 |  |  | (persistence or prevalence or incidence or epidemiology).ti,ab. | | | 1824346 | |
| 10 |  |  | (symptom duration or symptom resolution or symptom).ti,ab. | | | 256492 | |
| 11 |  |  | (((protect$ or risk) adj2 factor) or protective factor or risk factor).ti,ab,kw. | | | 280961 | |
| 12 |  |  | or/8-11 | | | 4170188 | |
| 13 | ***Total*** | | 4 and 7 and 12 | | | 275 | |
| **EMB review search strategy** | | | | | | | |
| **#** | **Description** | | | **Searches** | | | **Results** |
| 1 | ***Long COVID terms*** | | | ((long or post or persist* or sequela* or complication) adj5 (COVID* coronavirus* or SARS*)).ti,ab. | | | 121 |
| 2 |  |  |  | (longCOVID or long COVID or long-COVID or long haul COVID or long-haul COVID or post-acute COVID syndrome or persistent COVID-19 or post-acute COVID19 syndrome or long hauler COVID or post-acute sequelae of SARS-COV-2 infection or chronic COVID syndrome or post COVID-19 syndrome or post COVID-19 condition or post-acute sequelae of COVID).ti,ab. | | | 268 |
| 3 |  |  |  | or/1-2 | | | 356 |
| 4 | ***Vaccine terms*** | | | COVID-19 Vaccines/ or exp vaccines/ | | | 16600 |
| 5 |  |  |  | (COVID-19 adj2 (vaccine$ or vaccination$ or innoculation$)).ti,ab. | | | 1165 |
| 6 |  |  |  | 4 or 5 | | | 17364 |
| 7 | ***Outcome terms*** | | | (symptom$ or post-acute outcomes or long term outcomes or (((long term or persisten$) adj3 symptom) or outcome)).ti,ab. | | | 622095 |
| 8 |  |  |  | (persistence or prevalence or incidence or epidemiology).ti,ab. | | | 176142 |
| 9 |  |  |  | (symptom duration or symptom resolution or symptom).ti,ab. | | | 61523 |
| 10 |  |  |  | (((protect$ or risk) adj2 factor) or protective factor or risk factor).ti,ab,kw. | | | 34936 |
| 11 |  |  |  | or/7-10 | | | 749053 |
| 12 | ***Total*** | | | 3 and 6 and 11 | | | 13 |

**eTable 2. PICOS study eligibility criteria**

| **Topic** | **Inclusion Criteria** | **Exclusion criteria** |
| --- | --- | --- |
| Population(s) | Persons aged ≥12 years with PCC identified by clinical assessment/diagnosis or self-reported and who received a COVID-19 vaccination at any of the three time points outlined:   - Before SARS-CoV-2 infection - After acute SARS-CoV-2 infection - After PCC | Individuals infected with COVID-19 but not diagnosed with PCC (unless included as a comparator group to one of the inclusion population groups)  Pediatrics |
| Intervention | At least one treatment arm investigating mRNA or adenoviral vector COVID-19 vaccines, or inactivated vaccines administered as a homogeneous or heterogeneous dose, including but not limited to:   - Comirnaty (Pfizer-BioNTech) - Spikevax (Moderna) - Vaxzevria (AstraZeneca) - Nuvaxoid (Novavax) - Jcovden (Johnson & Johnson/Janssen) - VidPrevtyn Beta (Sanofi Pasteur) - Sinovac (Sinovac Biotech Ttd) - Other vaccines to be specified and agreed with Pfizer - Vaccination in combination with other treatment | Studies focusing primarily on alternative COVID-19 treatment/ prevention measures (i.e., antivirals, monoclonal antibodies). |
| Comparison | Any COVID-19 vaccine with alternative dose (e.g., primary series [1, 2 doses], booster series)  Unvaccinated / 0 doses | N/A |
| Outcomes | Below outcomes to be assessed over time:   - Protection/prevention against PCC with vaccination - Persistence/duration of symptoms postacute infection phase/Severity of symptoms - Prevalence/incidence of symptoms post-acute infection phase - Time to symptom resolution post-acute infection phase - Impact of vaccination on individuals with existing PCC (severity, duration, attenuation, resolution) - Complications including worsening of existing medical conditions or development of new condition (cardiopulmonary disease or musculoskeletal condition) | N/A |
| Time | No time horizon, studies only published and relevant from January 1^st^ 2020 to July 18^th^ 2023 | N/A |
| Study design | Study types to be included are:   - Prospective cohort studies - Retrospective cohort studies - Case-control studies - Cross sectional studies - Cross-over studies - Clinical Trials - Target trial emulation - Systematic literature reviews and meta-analyses (bibliography checks only) | Study types to be excluded are:   - Editorials - Case studies - Letters to journals - Non-systematic literature reviews - Study protocols - Conference minutes - Publication pre-prints which have been pre-prints for >6 months. |

N/A: not applicable; mRNA: messenger ribonucleic acid; PCC: post-COVID-19 conditions.

**eTable 3. Summary of studies included in SLR (n=97)**

| Author, year (* indicates low-risk study) | Countries | Study population (e.g., HCPs, community sample, hospitalized patients, etc.) | Data collection period and PCC Follow-up Period | Data sources for SARS-CoV-2, vaccination, and PCC | Variant | Vaccination timing relative to infection or PCC | Vaccination definition |
| --- | --- | --- | --- | --- | --- | --- | --- |
| Abu Hamdh, 2023*^1^ | Palestine | N=669 real time- polymerase chain reaction (RT-PCR) confirmed COVID-19 patients (≥18 years)  Community sample | COVID-19 patients diagnosed: September 2021 – January 2022  Follow-up: semi-structured telephone interviews on days 10, 30, 60, and 90 | SARS-CoV-2: EHR  Vaccination: patient-reported  PCC: medical codes/ claims | Not available on CoVariants | Before infection | Vaccination defined as ≥1 dose. Prior to infection, 41.0% were vaccinated against COVID-19: Pfizer (17.8%), Sputnik light (12.7%), Moderna (3.7%), Sputnik-V (2.4%), Sinopharm (1.9%), AstraZeneca (2.4%) |
| Al-Aly, 2022*^2^ | US | N=13,482,547 from a national healthcare database of the US department of Veterans affair.  N=147,414 included. | March 2020 – January 2021. Follow-up: within 6 months (all outcomes were assessed starting from 30 days after first positive SARS-CoV-2 test.) | SARS-CoV-2, vaccination, and PCC: Veterans Health Administration (VHA) Corporate Data Warehouse | WT, Alpha, Delta | Before infection | Complete vaccination defined following CDC guidelines at 14 days after first Janssen vaccination (11.62%) and 14 days after second Pfizer-BioNTech (47.94%) or Moderna (40.44%) |
| Alghamdi, 2022*^3^ | Saudi Arabia | N=2,218 participants diagnosed with laboratory-confirmed SARS-CoV-2 infection | Participants asked to provide duration of each PCC experienced (<1 to > 6 months.) | SARS-CoV-2, vaccination, and PCC: collected via online questionnaire | NR | NR | Oxford/AstraZeneca or Pfizer |
| Alradhi, 2022^4^ | Iraq | N=756 ≥18 years of age, no risk factors or chronic diseases | 4 October –25 November 2021 | SARS-CoV-2, vaccination, and PCC: survey, patient-reported | Delta | Before infection | Pfizer (70.77%), Sino-harm (17.64%), and AstraZeneca (11.58%) |
| Angarita-Fonseca, 2023^5^ | Multiple - South America | N=2,466 reported having COVID-19 | Survey administered: November–December 2021 | SARS-CoV-2, vaccination, and PCC: patient-reported via questionnaire | Delta | Mixed | NR |
| Antonelli, 2022a*^6^ | UK | N=16,800 ≥18 years old using the COVID Symptom Study App | December 2020–July 2021  Follow-up: ≥14 days after first dose of vaccination and ≥7 days after second dose of vaccination. | SARS-CoV-2, vaccination, and PCC: patient-reported via study app on phone | Alpha, Delta | Before infection | 2 or 1 doses. Pfizer, Moderna, Oxford/AstraZeneca, Unknown |
| Antonelli, 2022b*^7^ | UK | N=97,364, with positive RT-PCR test for COVID-19 after vaccination | June 2021 –March 2022  Follow-up period: 28 days after testing positive. | SARS-CoV-2, vaccination, and PCC: patient-reported data from the COVID symptom study app | Delta, Omicron | Before infection | NR |
| Arjun, 2022a*^8^ | India | N=487 adults (age ≥18 years) diagnosed with COVID-19 via RT PCR test and were either hospitalized or tested as an outpatient. | April 2021 – September 2021  Follow-up at 4 weeks and 6 months after diagnosis  Median follow-up for 4-week interview: 44 days (IQR: 39,47)  Median follow-up for 6m interview: of 223 days (IQR: 195,251) | SARS-CoV-2, vaccination, and PCC: patient-reported | Delta | NR | Mixed (majority Covaxin)  2 doses (58.9%),  1 dose (16.6%), unvaccinated (24.5%) |
| Arjun, 2022b*^9^ | India | N=524 participants (≥18 years) with COVID-19 RT-PCR-confirmed diagnosis from COVID-19 screening Outpatient Department (OPD) of a tertiary care government hospital and research institute | February 2022  Individuals contacted for telephone interviews ≥ 29 days after diagnosis. Median number of days of follow-up was 73 (IQR: 67–83) | SARS-CoV-2, vaccination, and PCC: patient-reported through telephone interview | Omicron | NR | Mixed (majority Covaxin: 74.7%)  3 doses (2.9%), 2 doses (88.9%), 1 dose (3.2%), unvaccinated (5%). |
| Augustin, 2023^10^ | Germany | N=222 outpatients experienced mild symptoms of COVID-19 and had received care at the post-COVID outpatient clinic of the University Hospital of Cologne (UHC) | Received care at post-COVID outpatient clinic: April 2020 – August 2021  PCC: ≤15 months post-infection, including four scheduled visits, defined as T1 (baseline), T2 (6 months), T3 (10 months), and T4 (15 months). Median follow-up was 15 months (IQR: 15-16) | SARS-CoV-2, vaccination, and PCC: patient-reported | Timepoint NR | After infection | ≥1 dose (77.0%) between 10 and 15 months after symptom onset.  59.6% and 40.4% received single and double vaccination, respectively. |
| Ayoubkhani, 2022a*^11^ | UK | N=28,356 participants aged 18─69 years, completing community-based surveys | February–September 2021  Long COVID measured for 12 weeks following positive SARS-CoV-2 test result.  Median follow-up among Vaccinated: 96 days (IQR: 90 to 104)  Median follow-up among Unvaccinated: 98 days (IQR: 89 to 109) | SARS-CoV-2, vaccination, and PCC: patient-reported through telephone interview: patient-reported via UK COVID-19 Infection Survey (CIS), run by the Office for National Statistics (ONS) | Alpha, Delta | After infection | Oxford/AstraZeneca, Pfizer, Moderna |
| Ayoubkhani, 2022b*^12^ | UK | N=13,187 participants aged 18─69 years who tested positive for SARS-CoV-2 | April 2020 – November 2021 | SARS-CoV-2, vaccination, and PCC: patient-reported through telephone interview: patient-reported via UK COVID-19 Infection Survey (CIS), run by the Office for National Statistics (ONS) | Delta | Before infection | ≥2 doses of Oxford/AstraZeneca, Pfizer, or Moderna, ≥14 days before the first test-confirmed infection. |
| Azzolini, 2022*^13^ | Italy | N=2,560 HCPs across 9 Italian healthcare facilities, with documented positive COVID-19 test | March 2020 –April 2022 | SARS-CoV-2, vaccination, and PCC: patient-reported via survey | WT, Alpha, Delta, Omicron | Before infection | Pfizer vaccination ≥14 days prior to infection. |
| Babicki, 2023*^14^ | Poland | N=801 patients who contracted COVID-19 and examined during a personal medical visit after contracting COVID-19 | COVID-19 diagnosis dates not specified.  Follow-up visits: in the 3rd and 12th month after the end of the disease | SARS-CoV-2, vaccination, and PCC: patient-reported, collected during medical visits, part of the STOP-COVID registry | Timepoint NR | Mixed | Pfizer, Moderna, AstraZeneca or Janssen  2 doses of Pfizer, Moderna or AstraZeneca, or 1 dose of Janssen.  9.4% vaccinated before and 73.6% vaccinated after SARS-CoV-2 infection. |
| Ballouz, 2023^15^ | Switzerland | N=1,045 Zurich Cohort; N=305 Corona Immunitas | Follow-up at 6-months | SARS-CoV-2, vaccination, and PCC: patient-reported via survey | Delta, Omicron | Before infection | 74.6% received 1 or 2 doses, and 25.4% received 3 doses.  mRNA: 231 (99.6%); Adenovirus vector: 1 (0.4%) |
| Bello-Chavolla, 2023*^16^ | Mexico | N=11,472 completed interviews, adults aged ≥20 years | July –December 2022 | SARS-CoV-2, vaccination, and PCC: patient-reported via the Mexican National Health and Nutrition Survey (ENSANUT) | Delta, Omicron | NR | ≥1 dose (86.21%),  1 dose (11.62%), 2 doses (29.6%), 3 doses (39.51%), 4 doses (5.48%). |
| Berry, 2023*^17^ | Bonaire | N=47 PCC patients | November – December 2021  SARS-CoV-2 positive cases detected until October 2021.  Telephone interviews conducted: November to December 2021 | SARS-CoV-2, vaccination, and PCC: patient-reported via telephone interview | Delta | After infection | ≥1 dose Pfizer ≥8 weeks after SARS-CoV-2 infection.  After infection, 23% remained unvaccinated; 76% were vaccinated with ≥1 dose of Pfizer vaccine. |
| Bostanci, 2023*^18^ | Cyprus | N=296 patients diagnosed with SARS-CoV-2 infection, non-hospitalized | September 2021 –February 2022 | SARS-CoV-2, vaccination, and PCC: patient-reported via online questionnaire | WT, Alpha, Delta, Omicron | Before infection | Coronavac (30.4%), Pfizer (49.2%), Moderna (1.6%), Janssen (14.7%), Oxford AstraZeneca (4.2%) |
| Brannock, 2023*^19^ | US | Clinic-based: n=47,404, with ICD-10 COVID-19 diagnosis code or positive test, ≥18 years of age.  Model-based: n=198,514. | Patients with evidence of COVID-19 between August 2021 and January 2022  EHRs available 45 days after COVID-19 infection used to establish long COVID.  Minimum follow-up period: 164 days (range: 150-389 days). | SARS-CoV-2, vaccination, and PCC: Medical codes/ claims | Delta, Omicron | Before infection | 2 doses of Pfizer, Moderna, or 1 dose of AstraZeneca, 2 weeks prior to the index were considered vaccinated |
| Brunvoll, 2022*^20^ | Norway | N=1,420 of the Norwegian COVID-19 Cohort | Follow-up surveys administered November 2020, January 2022 (2–15 months after positive SARS-CoV-2 test). | SARS-CoV-2, vaccination, and PCC: patient-reported via questionnaire | WT | Before infection | Full vaccination ≥14 days after 2^nd^ dose |
| Cai, 2023*^21^ | China | N=21,799 laboratory confirmed, or clinician diagnosed with COVID-19 at Huashan Hospital | March–June 2022  1^st^ follow-up: 6 months post discharge  2^nd^ follow-up: for those with Long COVID - additional 12-month follow-up | SARS-CoV-2, vaccination, and PCC: patient-reported via survey | Omicron | NR | 1 dose (5.28%) and ≥2 doses (85.08%). |
| Català, 2023*^22^ | Multiple - Europe | CPRD GOLD (UK): 3,258,766 CPRD AURUM (UK): 11,590,479 SIDIAP (Spain): 5,333,339  CORIVA (Estonia): 379,870 | Cohort 1: January 2021  Cohort 2: January─ February 2021  Cohort 3: March─ April 2021  Cohort 4: April─ July 2021  Follow-up from index date until earliest end of observation (date of data extraction [December 2021 for CPRD GOLD; January, 2022 for CPRD AURUM; June, 2022 for SIDIAP; and December, 2022 for CORIVA], all-cause death, leaving the GP practice or stopping contribution of data to database (for CPRD), or the next vaccination (first vaccination for unvaccinated group, second vaccination for vaccinated group) | SARS-CoV-2, vaccination PCC: linked national/ regional registry data (CPRD GOLD [UK], CPRD AURUM [UK], SIDIAP [Spain] and CORIVA [Estonia]. | Delta, Omicron | Before infection | 1 dose of any vaccine.  AstraZeneca: 59% of CPRD GOLD, 56% of CPRD AURUM, 15% of SIDIAP, 6% of CORIVA.  Pfizer: 38% CPRD GOLD, 40% of CPRD AURUM, 66% of SIDIAP, 75% of CORIVA.  Janssen and Moderna also included. |
| Chelly, 2023*^23^ | Tunisia | N=1,381 infected with COVID-19 ≥1 | Conducted in February 2022 among those infected with COVID-19 between March 2020 and February 2022 | SARS-CoV-2, vaccination, and PCC: patient-reported via online questionnaire | WT, Alpha, Delta, Omicron | Before infection | 3.2% unvaccinated, 7.7% partially vaccinated and 19.1% fully vaccinated |
| Chilunga, 2023*^24^ | Netherlands | Hospitalized migrant and non-migrant patients | January 2021–December 2021  PCC follow-up at 6, 12 weeks and >12 weeks if symptoms persisted. | SARS-CoV-2, vaccination, and PCC: EHR | WT, Alpha, Delta | Before infection | Any vaccination prior to admission. |
| Cortellini, 2023*^25^ | Multiple - Europe | N=1,808 patients aged ≥18 years with a RT-PCR-confirmed diagnosis of COVID-19 and a history of solid or hematological malignancy, either active or in remission, at the time of COVID-19 diagnosis. | February 27, 2020– January 31, 2022, with follow–up update on June 20 2022  Unvaccinated: assessed median 40 days (IQR: 25─67) days since COVID-19 diagnosis; partially vaccinated, 50 days (28─68); double-vaccinated, 38 days (23─68); boosted, 34 days (22─67). | SARS-CoV-2, vaccination and PCC: EHR, Registry data (OnCovid) | Alpha, Delta, Omicron | Before infection | ≥2 doses prior to infection with Pfizer, Moderna, AstraZeneca, or 1 dose of Janssen |
| de Arriba Fernández, 2022*^26^ | Spain | N=110,726 patients ≥12 years of age, and were diagnosed with COVID-19 | COVID-19 diagnosed between June 2021 – February 2022 | SARS-CoV-2, vaccination, and PCC: EHR and registry of vaccination against COVID-19 in Spain (REGVACU). | Delta, Omicron | Mixed | 2 doses separated by ≥19 days if 1^st^ dose was Pfizer, 21 days if AstraZeneca or 25 days mRNA-1273.  Or ≥7 days if the last dose was Pfizer or 14 days if it was AstraZeneca or Moderna.  1 dose of Janssen ≥14 days before infection |
| de Arriba Fernández, 2023*^27^ | Spain | N=110,726 patients ≥12 years of age, and were diagnosed with COVID-19 | COVID-19 diagnosed between June 2021 to February 2022 | SARS-CoV-2, vaccination, and PCC: EHR and registry of vaccination against COVID-19 in Spain (REGVACU). | Delta, Omicron | Before infection | Moderna, Pfizer, AstraZeneca, Janssen |
| de Bruijn, 2023*^28^ | Netherlands | N=15,391 enrolled on the national Dutch prospective LongCOVID-study, aged ≥18 years, with positive COVID-19 test | Cases and controls enrolled between January 2022–May 2022 | SARS-CoV-2, vaccination, and PCC: patient-reported via online questionnaire | Delta, Omicron | Before infection | Moderna, Pfizer, AstraZeneca, Janssen |
| Di Fusco, 2023*^29^ | US | N=430 individuals testing for SARS-CoV-2 at one of ~ 5000 CVS Health test sites across the US | Recruitment: January 2022 –April 2022  Follow-up: occurred through October 2022, survey administered at week 4, month 3 and month 6 after enrolment. | SARS-CoV-2, vaccination, and PCC: patient-reported via online survey | Omicron | Before infection | Immunocompetent participants: full vaccination, 2 doses ≥14 days before SARS-CoV-2 testing; partially vaccinated, 1 dose; boosted, 3 doses.  Immunocompromised: full vaccination, 3 doses; boosted, 4 doses. |
| Durstenfeld, 2023*^30^ | US | N=1,480 participants ≥18 years, who reported a positive SARS-CoV-2 test result | Enrolment: March 2020  Data collection: March 2020─ May 2022  Long-COVID data collection: January or May 2022 | SARS-CoV-2, vaccination, and PCC: patient-reported via online survey | WT, Alpha, Delta, Omicron | Before and after infection depending on outcome | Vaccine NR, 70.9% of participants had 1st vaccine dose |
| El Otmani, 2022*^31^ | Morocco | N=236 HCPs | February 2021 –April 2021 | SARS-CoV-2, vaccination, and PCC: patient-reported via survey | Alpha | After infection | Sinopharm and AstraZeneca |
| Elias, 2023*^32^ | Ethiopia | N=340 severe COVID-19 patients who were discharged alive from Millennium COVID-19 care center (MCCC) | February 1, 2023 and April 30, 2023 | SARS-CoV-2, vaccination and PCC: patient-reported via telephone interview and structured questionnaire adapted from the W.H.O Global COVID-19 Clinical Platform Case Report Form (CRF) for Post COVID condition (Post COVID-19 CRF) questionnaire | Omicron | NR | NR |
| Elmazny, 2023*^33^ | Egypt | 1,638 Egyptian COVID-19 survivors ≥18 years of age (569 males, 1,069 females) ≥2 months after recovery from COVID-19 infection (diagnosis confirmed with PCR testing on nasopharyngeal sample. (Mean [SD] age: 38.28 ± 13.004 years). Participants followed up in outpatient clinics or hospitalized in COVID-19 wards in Beni-Suef University Hospital | Study conducted over 10 months (May 2020–February 2022)  PASC assessment: 3 months after COVID-19 infection | SARS-CoV-2, vaccination, and PCC: patient-reported via interview (telephone and face-to-face in clinic) | WT, Alpha, Delta, Omicron | Mixed (outcomes refer to vaccination before infection) | Fully vaccinated before infection (18.6%); vaccinated after infection (49.9%); not vaccinated (31.5%).  Pfizer (15.2%), AstraZeneca (31.7%), Janssen (3.9%), Sputnik V (1.0%), Vac Sinovac (18.1%), Sinopharm (28.5%), Moderna (1.6%) |
| Elnadi, 2023^34^ | Multiple - African | N=706 healthcare workers | December 2021–January 2022 | SARS-CoV-2, vaccination, and PCC: patient-reported via online questionnaire | Delta, Omicron | NR | 66.4% received a COVID-19 vaccine and 80.6% of those vaccinated received 2 doses. |
| Emecen, 2023^35^ | Turkey | N=5,610 SARS-CoV-2 RNA positive participants aged ≥ 18 years | December 2020– May 2021 | SARS-CoV-2, vaccination, and PCC: patient-reported | WT, Alpha | Before infection | Fully vaccinated: ≥2 weeks after 2^nd^ dose of CoronaVac or Pfizer vaccine. |
| Ertesvag, 2023*^36^ | Norway | Children and adolescents (n = 276) aged 10–20 years with confirmed SARS-CoV-2. | COVID-19 infection: August─ September 2021  Follow-up: 3 (median: 104 days [IQR: 62─109]) and 6 ([median: 245 days [IQR: 205─254]) months post-infection. | SARS-CoV-2, vaccination, and PCC: patient-reported via questionnaire telephone interview of children or their parents (children <16 years) | Delta | Before infection | Received 2^nd^ dose ≥14 days prior to infection.  Pfizer (94%), Moderna (6%) |
| Fernández-de-las-Peñas, 2022*^37^ | Spain | N=210 patients hospitalized for a SARS-CoV-2 infection in an urban hospital in Madrid, Spain who survived. | COVID-19 diagnosis: July – August 2021  Follow-up: 5 months after infection and hospitalization (mean: 6.3 months [SD: 1.0]). | SARS-CoV-2 and vaccination: EMR  PCC: patient-reported via survey | Delta | Before infection | Vaccinated: 2 doses of Pfizer vaccine.  1 dose considered as non-vaccinated. |
| Gaber, 2020^38^ | UK | N=77 healthcare workers with long-COVID. | Vaccination: December 2020─ January 2021  Data collection: February 2021 | SARS-CoV-2, vaccination, and PCC: patient-reported via questionnaire | Alpha | After PCC onset | ≥2 weeks after single vaccination of Pfizer or Moderna |
| Gallant, 2022*^39^ | Canada | N=138 individuals from the Estrie region of Quebec (community sample) | Follow-up interviews: 1-, 3-, 6-, 12-, 18- and 24- months post-infection (August – November 2021) | SARS-CoV-2, vaccination, and PCC: patient-reported | Delta | Before infection | Breakthrough infections defined as infection ≥2 weeks after 2 doses of vaccine (Pfizer or Moderna) |
| Hajjaji, 2022^40^ | France | N=7,281 cancer patients treated at Oscar Lambret Cancer Center during the pandemic. | April 2020–July 2021  Cohort 1: Survey shared at 3 time points (April 2020, July 2020, and August 2021)  Cohort 2: August 2021 | SARS-CoV-2, vaccination, and PCC: patient-reported via online questionnaire | WT, Alpha, Delta | NR | With long-lasting symptoms:  N=46 non-vaccinated  N=78 1 dose  N=44 2 doses |
| Hastie, 2022*^41^ | Scotland | N=96,238 >16 years of age, with positive PCR test for SARS-CoV-2 | May 2021  Follow-up surveys at 6-, 12-, and 18-months | SARS-CoV-2, vaccination, and PCC: patient-reported via online questionnaire | Alpha, Delta | Before infection | Non-vaccinated (94.29%)  1 dose (4.96%)  2 doses (0.64%) |
| Haupert, 2023*^42^ | US | N=44,198 COVID-19-positive survivors, matched with n=160,399 test negative controls. | COVID-19 diagnoses: March 2020 to August 2022 | SARS-CoV-2, vaccination, and PCC: EHR records in the Michigan Medicine health system | WT, Alpha, Delta, Omicron | Before infection | Fully vaccinated: 2 doses of Moderna, Pfizer-BioNTech, or AstraZeneca; 1 dose of Janssen, and ≥21 days had elapsed. |
| Herman, 2022*^43^ | Indonesia | N=923 recovered COVID-19 patients (community sample) | July 2021–February 2022 | SARS-CoV-2, vaccination, and PCC: patient-reported via online questionnaire | Delta, Omicron | Before infection | Fully vaccinated: ≥14 days after 2^nd^ dose. |
| Hirahata, 2022*^44^ | Japan | N=1,989 long-COVID patients | 6 January 2020 – 2 October 2021 | SARS-CoV-2, vaccination, and PCC: EMR and patient-reported | WT, Alpha, Delta | NR | NR |
| Ioannou, 2022*^45^ | US | N=198,601 COVID-19 survivors (3-months post infection) | February 2020–April 2021  Follow-up: mean (SD) follow-up of 13.5 (3.6) months post-acute infection | SARS-CoV-2, vaccination, and PCC: EHR from Veterans Affairs (VA) database | WT, Alpha | Before infection | ≥1 dose of Moderna or Pfizer before infection.  87.5% non-vaccinated  8.8% 1 dose  3.6% 2 doses |
| Jacobs, 2023^46^ | US | N= 257,554 adults | June – October 2022 | SARS-CoV-2, vaccination, and PCC: patient-reported via questionnaire | Omicron | NR | NR |
| Jaramillo, 2023*^47^ | US | N=225 ≥18 years of age | August–September 2022  Duration since testing positive ranged from 6─906 days; only those positive ≥60 days prior to survey date were selected | SARS-CoV-2, vaccination, and PCC: patient-reported via online survey | Omicron | NR | All individuals vaccinated.  57% with long COVID and 55% with COVID reinfection cohort received 2 boosters. |
| Jassat, 2023*^48^ | South Africa | Four cohorts recruited: (1) hospitalized Beta wave; (2) hospitalized Delta wave; (3) hospitalized Omicron wave; and (4) non-hospitalized Delta wave.  N=2,626 hospitalized and n=1,074 non-hospitalized completed survey | Follow-up: 1-, 3-, and 6-months post SARS-CoV-2 infection | SARS-CoV-2, vaccination, and PCC: patient-reported via case report form | Beta, Delta, Omicron | Before and after infection (individual outcomes) | Fully vaccinated: 2 doses of Pfizer or 1 dose of Janssen ≥14 days before infection. |
| Kahlert, 2023*^49^ | Switzerland | N=2,912 healthcare workers | July/August 2020 –May/June 2022 | SARS-CoV-2, vaccination, and PCC: patient-reported via electronic questionnaire | WT, Alpha, Delta, Omicron | Before infection | ≥99% of vaccinated participants received Pfizer or Moderna |
| Kim, 2023*^50^ | South Korea | N=132 patients diagnosed with COVID-19 (age 16–70 years) | February – March 2020  Follow-up: 1^st^ at 5–6 months after onset of COVID-19 symptoms or diagnosis in September 2020.  2^nd^ 11–12 months after COVID-19 symptom onset or diagnosis between May and June, 2021.  3^rd^ at 24 months after COVID-19 symptom onset or diagnosis between May and June, 2022. | SARS-CoV-2, vaccination, and PCC: Patient-reported | WT | After infection | Pfizer, Moderna, AstraZeneca, Novavax or Janssen |
| Kubota, 2023*^51^ | Brazil | N=226 PCC patients, n=177 (78.3%) presented ≥3 months since first COVID-19 symptom | Enrolment: March to November 2021 | SARS-CoV-2, vaccination, and PCC: patient-reported via in-person structured interview and clinical exam | Timepoint NR | NR | Oxford (34.1%), CoronaVac (37.2%), Pfizer (16.8%); AstraZeneca (0.9%), NR (11.1%)  1 dose, 34.1%; 2 doses 54.9% |
| Kuodi, 2022*^52^ | Israel | N=3,572 patients >18 years testing positive for COVID-19 in three major hospitals (analysis restricted to N=951 based on previous infection and vaccination status). | March 2020 – November 2021 | SARS-CoV-2, vaccination, and PCC: patient-reported via online survey | WT, Alpha, Delta | Mixed | Pfizer  1 dose, 36%; ≥2 doses, 31% |
| Kuodi, 2023*^53^ | Israel | N=951 hospitalized patients and community patients, aged ≥18 years, with | March 2020 –June 2022 | SARS-CoV-2, vaccination, and PCC: patient-reported via online questionnaire | WT, Alpha, Delta, Omicron | Before or after infection | Pfizer |
| Lam, 2023*^54^ | Hong Kong and the UK | N=4,259,981 patients identified with a positive RT-PCR for COVID-19 from HKHA databases | HKHA: April 2020 –May 2022. UKB: March 2020 –May 2021 | SARS-CoV-2, vaccination, and PCC: EHR from Hong Kong Hospital Authority (HKHA) and United Kingdom Biobank (UKB) databases | WT, Alpha (UK only) Delta, Omicron | Before infection | Pfizer, AstraZeneca |
| Lippke, 2023^55^ | Germany | N=235 post-/long-COVID patients | March –October 2022  Follow-up surveys: 3-4 weeks and 4-5 weeks later | SARS-CoV-2, vaccination, and PCC: computer assisted telephone interview | Omicron | Before and after infection depending on outcome | Janssen, Unspecified |
| Luo, 2023*^56^ | China | N=6,242 non-hospitalized subjects of all ages with SARS-CoV-2 infection | Diagnosis: December 2021 to March 2022  Follow-up: November 2022–January 2023; median time from positive test to survey was 9 (IQR: 8─10) months | SARS-CoV-2, vaccination, and PCC: EHR and patient-reported symptom survey | Omicron | Before and after infection depending on outcome | Sinovac or Pfizer  <3 doses, 57.5% |
| Marra, 2023*^57^ | Brazil | N=18,340 HCPs ≥18 years | March 2020 – July 2022  Follow-up: 1-, 3- and 6-months after first infection | SARS-CoV-2, vaccination, and PCC: institutional electronic records including EMRs, laboratory reports from the microbiology laboratory, and separate records at the employee health clinic. Patient with positive test were assessed by the employee health clinic via in-person appointment, telephone appointment, or emails. | WT, Gamma, Delta, Omicron | Before infection | Oxford-AstraZeneca, CoronaVac, Pfizer, Janssen vaccines  0 doses: (LC: 60.8%; no LC: 39.5%)  1 dose: (LC: 5.0%; ), (3.6%, no LC)  2 doses: (14.1%, LC), (13.0%, no LC)  3 doses: (20.1% LC), (41.3% no LC)  4 doses: (0.1% LC), (2.6%, no LC) |
| McLaughlin, 2023a*^58^ | Scotland | N=253 patients ≥18 years of age, self-diagnosed of confirmed long-COVID | April 2021 –August 2022 | SARS-CoV-2, vaccination and PCC: patient-reported via online survey | Alpha, Delta, Omicron | NR | NR |
| McLaughlin, 2023b*^59^ | Scotland | N=253 patients ≥18 years of age, self-diagnosed of confirmed long-COVID | April 2021 –August 2022 | SARS-CoV-2, vaccination and PCC: patient-reported via online survey | Alpha, Delta, Omicron | NR | NR |
| Meza-Torres, 2022*^60^ | England | N=416,505 COVID-19 infection cases | March 2020 –April 2021  Long-COVID assessed 1─6 months after index infection | SARS-CoV-2, vaccination and PCC: Primary care sentinel cohort (PCSC) of the Oxford–Royal College of General Practitioners Research and Surveillance Centre | WT, Alpha | Mixed | NR |
| Mizrahi, 2023*^61^ | Israel | N=1,913,234 of Maccabi Healthcare Services members | March 2020 – October 2021  Follow-up for PCC: early (30-180 days) and late (180-360 days) time periods after infection. | SARS-CoV-2, vaccination and PCC: EHR | WT, Alpha, Delta | Before infection | Vaccinated: infection ≥14 days after 2^nd^ dose. |
| Mohr, 2023*^62^ | US | N=419 healthcare personnel with symptomatic acute COVID-19 infection | December 2020 – August 2021  Post-COVID follow-up: 6 weeks after symptom onset. | SARS-CoV-2, vaccination and PCC: patient-reported via survey/ interview | WT, Alpha, Delta | Before infection | Vaccinated: 2^nd^ dose of any mRNA vaccine ≥14 days prior to infection. |
| Munipalli, 2023*^63^ | US | N=132 patients with long-COVID symptoms, ≥18 years of age | NR | SARS-CoV-2 and PCC: patient-reported  Vaccination: EHR | Timepoint NR | Before infection | 89.4% vaccinated  Moderna, Pfizer, Janssen |
| Nascimento, 2023*^64^ | Brazil | N=412 patients admitted to two hospitals with confirmed COVID-19 | May 2021– February 2022  Follow-up: 90 days | SARS-CoV-2 and vaccination: EMR  PCC: patient-reported via survey for post-discharge data | Gamma, Delta, Omicron | Before infection | Fully vaccinated: ≥2 doses of any vaccine (NR) but ≥1 dose for Janssen. |
| Naveed, 2023*^65^ | Canada | N=629,935 participants (aged ≥18 years) who tested positive for SARS-CoV-2 by RT-PCR | January 2020 –December 2021 | SARS-CoV-2, vaccination and PCC: British Columbia COVID-19 cohort platform | WT, Alpha, Delta | Before infection | Fully vaccinated: ≥14 days since 2^nd^ dose  Partially vaccinated: ≥14 days since 1^st^ dose  Non-vaccinated: 0 doses |
| Nehme, 2022a*^66^ | Switzerland | N=2,094 who tested positive for COVID-19 using nasal swab RT-PCR at the Geneva University Hospitals outpatient testing center | April–July 2021 | SARS-CoV-2, vaccination and PCC: patient-reported via online survey | Delta | After infection | Moderna (60.7%), Pfizer (38.5%) |
| Nehme, 2022b*^67^ | Switzerland | N=2,689 individuals previously tested positive for SARS-CoV-2 infection (RT-PCR test) at the outpatient testing center of the Geneva University Hospitals, Switzerland. | December 2021–May 2022 | SARS-CoV-2, vaccination and PCC: patient-reported via online survey | Omicron | Before infection | Non-vaccinated: 14.8%  1 dose: 5.6%  ≥2 doses: 75.5% (fully vaccinated)  Moderna (61.2%), Pfizer (36.1%) |
| Parodi, 2023*^68^ | Argentina | N=1,883 patients who consulted for PCC 15 days after discharge for acute COVID-19 infection | 1st September 2021 – 30th June 2022  Mean time between COVID-19 diagnosis and consultation: 62 days (range: 16–330 days) | SARS-CoV-2, vaccination and PCC: collected via clinical examination/ consultation | Gamma, Delta, Omicron | Before infection | 2 doses ≥14 days before infection. |
| Patel, 2023*^69^ | US | N=280 patient with systemic autoimmune rheumatic diseases | 1 March 2020 – 8 July 2022 | SARS-CoV-2: EHRs  Vaccination and PCC: patient-reported via survey | Delta, Omicron | Before infection | 2 doses of Pfizer or Moderna or 1 dose of Janssen ≥14 days before infection. |
| Peghin, 2022*^70^ | Italy | N=479 diagnosed with COVID-19 at Udine Hospital. | September 2020 –November 2022 | SARS-CoV-2, vaccination and PCC: patient-reported via interview/ questionnaires | WT, Alpha | After infection | AstraZeneca (5.6%), Janssen (0.8%), Pfizer (90.5%), Moderna (3.2%) |
| Peghin, 2023*^71^ | Italy | N=230 in- and out-patients with COVID-19, ≥18 years of age | COVID-19 diagnosis: March 2020 –May 2021  Follow-up surveys:  6-, 12-, and 24-months | SARS-CoV-2, vaccination and PCC: patient-reported via telephone interview | WT, Alpha, Delta, Omicron | NR | AstraZeneca, Janssen, Pfizer, Moderna |
| Perlis, 2022*^72^ | US | N=16,091 individuals aged ≥18 years, inclusive of all 50 states and the District of Columbia | February 2021 and July 2022 (survey conducted every 6 weeks) | SARS-CoV-2, vaccination and PCC: patient-reported via internet survey | Alpha, Delta, Omicron | Before infection | Full vaccination: 2 doses of Janssen prior to the first month of illness, or a single dose . |
| Pinato, 2022^73^ | Multiple - Europe | N=2,090 patients aged ≥ years, with a history of solid or hematological malignancy | February 2020 –December 2021  Follow-up clinical assessments: median time of 40 days from COVID-19 diagnosis | SARS-CoV-2, vaccination and PCC: collected via OnCovid registry and clinical assessments | WT, Alpha, Delta, Omicron | Before infection | Fully vaccinated: 2 doses of Pfizer (59.3%), Moderna (19.8%), AstraZeneca (17.6%), or one dose of Janssen (≥28 days before infection for Janssen). |
| Qasmieh, 2023^74^ | US | N=3,042 adult US residents | June – July 2022 | SARS-CoV-2, vaccination and PCC: patient-reported via survey | Omicron | NR | NR |
| Rahman, 2023^75^ | Pakistan | N=250, diagnosed through PCR testing in the city of Peshawar, Pakistan | June 2022–August 2022 | SARS-CoV-2, vaccination and PCC: patient-reported via in-person questionnaire | Omicron | Before infection | Fully vaccinated: ≥2 doses of Pfizer, Sinopharm, Sinovac, or Moderna |
| Ranucci, 2023*^76^ | Italy | N=131  Conducted at the IRCCS San Donato, a Clinical Research Hospital | COVID-19 diagnosis: January 2021 to July 2022  Recruitment: April to November 2022  Follow-up: April to November 2022 | SARS-CoV-2 and vaccination: EHR  PCC: patient-reported via interview | WT, Alpha, Delta, Omicron | Before infection | Fully vaccinated: 2 doses at time of hospital admission.  Pfizer (63.6%), Moderna (17.4%), AstraZeneca (5.8%), Janssen (1.7%), Unknown (11.5%) |
| Resendez, 2023*^77^ | US | N=367,148 with positive COVID-19 tests | January 1, 2020 and August 18, 2022.  PCC assessed 1─7 months following positive COVID testing. | SARS-CoV-2, vaccination and PCC: EHR of all patients that tested positive for COVID-19 at VA medical facilities nationally | WT, Alpha, Delta, Omicron | Before infection | ≥1 dose, ≥2 weeks and ≤9 months before infection. |
| Richard, 2023*^78^ | US | N=1,832 US military health system beneficiaries, aged ≥18 years | COVID-19 diagnosis: February 2020 – December 2021  Follow-up: 1-, 3-, 6-, 9-, and 12-months after symptom onset. | SARS-CoV-2 and vaccination: EMR  PCC: patient-reported via survey | WT, Alpha, Delta | Before infection and after PCC onset infection (individual outcomes) | 77.1% unvaccinated prior to infection.  Post-symptom onset: 12.5%, 39.0%, 58.0%, 73.2%, and 82.9% received ≥1 dose at 1-, 3-, 6-, 9-, and 12-month follow-up with Janssen, Moderna, or Pfizer |
| Robertson, 2022*^79^ | US | N=3,042 living in the US | June – July 2022 | SARS-CoV-2, vaccination and PCC: patient-reported via survey | Omicron | NR | Pfizer, Moderna, Janssen |
| Scherlinger, 2022*^80^ | France | N=567 adults with confirmed (RT-PCR/serology) or suspected COVID-19 and long-COVID symptoms. | 3 and 17 August 2021 | SARS-CoV-2, vaccination and PCC: patient-reported via online survey | Delta | After PCC onset | 64.4% had a complete vaccination scheme.  Pfizer (78.1%),  Moderna (16.4%),  AstraZeneca (4.3%),  Janssen (0.5%),  mRNA/vector vaccine combination (0.5%),  unknown (0.3%) |
| Senjam, 2022*^81^ | India | N=773 positive report of SARS CoV-2 test | COVID-19 diagnosis: January to April 2021  Follow-up survey: June – July 2021 | SARS-CoV-2, vaccination and PCC: patient-reported via semi-structured questionnaire | Delta | Before infection | 52.7% not vaccinated. 22.6% had one dose and 24.7% had two doses before infection. |
| Smits, 2023*^82^ | US | N=438,431 with first positive COVID-19 laboratory test between October 2021 and November 2022 | COVID-19 diagnosis: October 2021 to November 2022 | SARS-CoV-2 and vaccination: EHR  PCC: EHR with ICD-10 and Systemized Nomenclature of Medicine – Clinical Terms (SNOMED – CT) codes | Omicron | Before (for assessed outcomes) | Primary or primary + booster vaccination schedule ≥28 days from infection, with Moderna, Pfizer, Novavax, or Janssen |
| Strain, 2022*^83^ | Global | N=812 with current or recent symptoms of Long COVID at the time of vaccination, a diagnosis of COVID-19 based on PCR/antibody testing, symptoms and contact with a proven case, or symptoms alone | March 2021 and April 2021  Follow-up: 1-21 weeks post-single vaccination | SARS-CoV-2, vaccination and PCC: patient-reported via semi-structured questionnaire | WT, Alpha, Delta | After PCC onset | AstraZeneca (50%),  Pfizer (40%),  Moderna (8.6%) |
| Tannous, 2023*^84^ | US | N=53,239 participants aged ≥18 years AND with a positive PCR test result for COVID-19 AND who survived beyond 28-days of their initial diagnosis | March 2020 – November 2021 | SARS-CoV-2, vaccination and PCC: EMR | WT, Alpha, Delta | Before infection | 2 doses of mRNA vaccines or 1 dose of Janssen ≥14 prior to infection. |
| Taquet, 2022*^85^ | Global | N=93,981 with confirmed COVID-19 infection occurring ≥14 days since vaccination | January 2021 –August 2021  6-month follow-up | SARS-CoV-2, vaccination and PCC: EHR via TriNetX Analytics | WT, Alpha, Delta | Before infection | Pfizer (65.1%), Moderna (9.0%), Janssen (1.6%), Unknown (24.4%) |
| Tosi, 2023*^86^ | US | N=148,700 US Veterans with positive COVID-19 test, without prior positive tests, who were alive 30 days after infection | January 22 | SARS-CoV-2, vaccination and PCC: EHR via VA COVID-19 Shared Data Resource | Omicron | Before infection | Fully vaccinated: 2 doses of Pfizer or Moderna or 1 dose of Janssen; 7 days after 2nd dose or 1 dose of Janssen.  Booster: additional dose of any vaccine.  Primary series: 39.2%  Booster: 22.5% |
| Tran, 2023*^87^ | France | N=910 adults (≥18 years) with confirmed or highly suspected COVID-19 infection | Long-COVID experienced between November 2020–May 2021 | SARS-CoV-2, vaccination and PCC: patient-reported via online questionnaire | WT, Alpha | After PCC onset | Pfizer (78.9%, Moderna (10.3%), AstraZeneca (10.5%), Janssen (0.2%) |
| Tsuchida, 2022^88^ | Japan | N=42 long COVID-19 patients visiting the outpatient department specializing in Long COVID at the hospital. | Changes in symptoms were evaluated before and 14–21 days after first vaccination | SARS-CoV-2, vaccination and PCC: patient-reported via survey | Timepoint NR | After PCC onset | NR |
| van der Maaden, 2023*^89^ | Netherlands | N=14,572 ≥18 years of age, mostly non-hospitalized | Enrolled: May – December 2021  Follow-up: 3-months | SARS-CoV-2, vaccination and PCC: patient-reported via survey | Delta | Before infection | Pfizer: 67.2%, AstraZeneca: 19.9%, Janssen: 13.0%, Moderna: 7.5%. |
| Wanga, 2021^90^ | US | N=3,135 adults tested for COVID-19 (positive or negative result) | April 2021 | SARS-CoV-2, vaccination and PCC: patient-reported via online survey | Alpha | Mixed | Received ≥1 dose of vaccine |
| Whittaker, 2021*^91^ | England | N=456,002 adult (≥18 years) patients with a COVID-19 diagnosis admitted to hospital within two weeks of diagnosis or managed in the community | COVID-19 diagnosis: August 2020 –February 2021  Data collection: May 2021  Follow-up: Maximum of 9.2 months | SARS-CoV-2, vaccination and PCC: EHR via GP records from the CPRD Aurum database. | WT, Alpha | Mixed | Pfizer, Moderna, Oxford-AstraZeneca |
| Wisnivesky, 2022*^92^ | US | N=453 patients (≥18 years) with laboratory documented infection with SARS-CoV-2, received care at Mount Sinai Health System (MSHS) registry | July 2020 –February 2021  6-month interview: August 2021 | SARS-CoV-2, vaccination and PCC: patient-reported via survey | WT, Alpha | After infection and PCC onset | Pfizer (62%), Moderna (32%) |
| Wong, 2023^93^ | China | N=2,712 community-dwelling COVID-19 patients | June, 2022 | SARS-CoV-2, vaccination and PCC: EHR and survey | Omicron | Before infection | Sinovac, Pfizer, other vaccines unnamed |
| Wynberg, 2022*^94^ | Netherlands | N=316 patients aged 16─85 years, with confirmed COVID-19 infection by RT-PCR | COVID-19 diagnosis May 2020 to June 2021  Recruited: January 2021 to November 2021 | SARS-CoV-2, vaccination and PCC: patient-reported via interview and clinical assessment | WT, Alpha, Delta | After infection | Pfizer (62%), Moderna (3%), AstraZeneca (3%), Janssen (1%), Missing (<1%) |
| Xue, 2023*^95^ | Global | N=9,717 (≥18 years) | May and December 2021 | SARS-CoV-2, vaccination and PCC: patient-reported via survey | Gamma, Alpha, Delta, Omicron | Before infection | Moderna, Pfizer  <2 doses: 38.4%  2 doses: 61.6% |
| Zhang, 2023*^96^ | Hong Kong | N=207 individuals aged ≥55 years, with ≥1 physical condition (e.g., hypertension, diabetes mellitus, chronic pain, sarcopenia), plus ≥1 mental/social condition | November 2019 –May 2021 (pre–5th wave) and from April –May 2022 (during the 5th wave) | SARS-CoV-2 and PCC: patient-reported via face-to-face pre-assessments in a public primary care outpatient  Vaccination: EHR | WT, Delta, Omicron | NR | Sinovac, Pfizer, Mixed  ≥1 dose: 85%  0 doses: 15% |
| Zisis, 2022*^97^ | US | N=1,578,719 ≥18 years with confirmed COVID-19 diagnosis | 3-month post-diagnosis follow-up between September 2020 –December 2021 | SARS-CoV-2, vaccination and PCC: EMR via the TriNetX database | WT, Alpha, Delta | Before infection | 1.6% had complete vaccination |

CDC: Centers for Disease Control and Prevention; CIS: COVID-19 Infection Survey; CPRD: Clinical Practice Research Datalink; CRF: Case Report Form; CT: Clinical terms; EHR: Electronic Health Record; EMR: Electronic Medical Record; HCP: Healthcare professional; HKHA: Hong Kong Hospital Authority; ICD-10: International Classification of Diseases- 10; IQR: Interquartile range; LC: Long-COVID; MCCC: Millennium COVID-19 care center; mRNA: Messenger ribonucleic acid; MSHS: Mount Sinai Health System; NR: Not reported; OCS: Office for National Statistics; OPD: Outpatient Department; PCC: post‑COVID‑19 conditions; PCR: Polymerase Chain Reaction; PCSC: Primary care sentinel cohort; RT-PCR: Real-time Polymerase Chain reaction; SARS: Severe acute respiratory syndrome; SD: Standard deviation; UK: United Kingdom; UKB: United Kingdom Biobank; US: United States; VA: Veterans Affairs; VHA: Veterans Health Administration; WT: Wild-Type.

**eTable 4. Summary of JBI Critical Appraisal ratings: Cohort Study design**

| **Primary author**  **Study Title** | Were the two groups similar and recruited from the same population? | Were the exposures measured similarly to assign people to both exposed and unexposed groups? | Was the exposure measured in a valid and reliable way? | Were confounding factors identified? | Were strategies to deal with confounding factors stated? | Were the groups/participants free of the outcome at the start of the study (or at the moment of exposure)? | Were the outcomes measured in a valid and reliable way? | Was the follow up time reported and sufficient to be long enough for outcomes to occur? | Was follow up complete, and if not, were the reasons to loss to follow up described and explored? | Were strategies to address incomplete follow up utilized? | Was appropriate statistical analysis used? | **Appraisal** | **Rationale for critical appraisal rating** |
| --- | --- | --- | --- | --- | --- | --- | --- | --- | --- | --- | --- | --- | --- |
| Abu-Hamdh, 2023 | Yes | Yes | Yes | NR | NR | Yes | Yes | Yes | Yes | NA | Yes | Low risk of bias |  |
| A prospective cohort study assessing the relationship between long‑COVID symptom incidence in COVID‑19 patients and COVID‑19 vaccination |  |  |  |  |  |  |  |  |  |  |  |  |  |
| Al-Aly, 2022 | Yes | Yes | Yes | Yes | Yes | NR | Yes | Yes | NR | NR | Yes | Low risk of bias |  |
| Long COVID after breakthrough SARS-CoV-2 infection |  |  |  |  |  |  |  |  |  |  |  |  |  |
| De Arriba Fernández, 2023 | Yes | Yes | Yes | Yes | Yes | No | Yes | NR | NR | NR | Yes | Low risk of bias |  |
| Evaluation of persistent COVID and SARS-CoV-2 reinfection in a cohort of patients on the island of Gran Canaria, Spain |  |  |  |  |  |  |  |  |  |  |  |  |  |
| Augustin, 2023 | NR | NR | Yes | NR | NR | Yes | Yes | Yes | NR | NR | NR | Moderate risk of bias | Limitations in extracting information on study design due to paper being a conference abstract. |
| 15-Month attributes of post-COVID syndrome in therapeutically vaccinated outpatients |  |  |  |  |  |  |  |  |  |  |  |  |  |
| Ayoubkhani, 2022 | Yes | Yes | Yes | Yes | Yes | NR | Yes | Yes | Yes | NA | Yes | Low risk of bias |  |
| Trajectory of long covid symptoms after covid-19 vaccination: community based cohort study |  |  |  |  |  |  |  |  |  |  |  |  |  |
| Ayoubkhani, 2022 | Yes | Yes | Yes | Yes | Yes | Yes | Yes | Yes | Yes | NR | Yes | Low risk of bias |  |
| Risk of Long COVID in People Infected With Severe Acute Respiratory Syndrome Coronavirus 2 After 2 Doses of a Coronavirus Disease 2019 Vaccine: Community-Based, Matched Cohort Study |  |  |  |  |  |  |  |  |  |  |  |  |  |
| Azzolini, 2022 | Yes | Yes | Yes | Yes | Yes | Yes | Yes | NR | NR | NR | Yes | Low risk of bias |  |
| Association Between BNT162b2 Vaccination and Long COVID After Infections Not Requiring Hospitalization in Health Care Workers |  |  |  |  |  |  |  |  |  |  |  |  |  |
| Babicki, 2023 | Yes | Yes | Yes | No | NR | Yes | Yes | Yes | Yes | NR | Yes | Low risk of bias |  |
| Do COVID-19 Vaccinations Affect the Most Common Post-COVID Symptoms? Initial Data from the STOP-COVID Register-12-Month Follow-Up |  |  |  |  |  |  |  |  |  |  |  |  |  |
| Ballouz, 2023 | No | No | Yes | Yes | Yes | Yes | Yes | Yes | NR | NR | Yes | Moderate risk of bias | Cohorts recruited from different population samples, demographics varied, attrition rate not reported. |
| Post COVID-19 condition after Wildtype, Delta, and Omicron SARS-CoV-2 infection and prior vaccination: Pooled analysis of two population-based cohorts |  |  |  |  |  |  |  |  |  |  |  |  |  |
| Berry, 2023 | Yes | Yes | Yes | Yes | Yes | No | Yes | NR | NR | NA | Yes | Low risk of bias |  |
| Association between post-infection COVID-19 vaccination and symptom severity of post COVID-19 condition among patients on Bonaire, Caribbean Netherlands: a retrospective cohort study. |  |  |  |  |  |  |  |  |  |  |  |  |  |
| Brannock, 2023 | Yes | Yes | Yes | Yes | Yes | NR | Yes | Yes | NR | NR | Yes | Low risk of bias |  |
| Long COVID risk and pre-COVID vaccination in an EHR-based cohort study from the RECOVER program |  |  |  |  |  |  |  |  |  |  |  |  |  |
| Cai, 2023 | Yes | Yes | Yes | Yes | No | Yes | Yes | Yes | NR | No | Yes | Low risk of bias |  |
| A one-year follow-up study of systematic impact of long COVID symptoms among patients post SARS-CoV-2 omicron variants infection in Shanghai, China |  |  |  |  |  |  |  |  |  |  |  |  |  |
| Catala , 2023 | No | Yes | Yes | Yes | Yes | Yes | Yes | Yes | Yes | NA | Yes | Low risk of bias |  |
| The effectiveness of COVID-19 vaccines to prevent long COVID |  |  |  |  |  |  |  |  |  |  |  |  |  |
| Chilunga, 2023 | Yes | Yes | Yes | Yes | Yes | Yes | Yes | Yes | Yes | No | Yes | Low risk of bias |  |
| Differences in incidence, nature of symptoms, and duration of long COVID among hospitalised migrant and non-migrant patients in the Netherlands: a retrospective cohort study |  |  |  |  |  |  |  |  |  |  |  |  |  |
| Cortellini , 2023 | Yes | Yes | Yes | Yes | Yes | Yes | Yes | NR | Yes | Yes | Yes | Low risk of bias |  |
| SARS-CoV-2 omicron (B.1.1.529)-related COVID-19 sequelae in vaccinated and unvaccinated patients with cancer: results from the OnCovid registry |  |  |  |  |  |  |  |  |  |  |  |  |  |
| de Arriba Fernández, 2022 | Yes | Yes | Yes | Yes | Yes | No | NR | NR | NR | NR | Yes | Low risk of bias | Definition of long-COVID not provided. Unclear on how long patients had had post-acute COVID-19 prior to study |
| Assessment of SARS-CoV-2 Infection According to Previous Metabolic Status and Its Association with Mortality and Post-Acute COVID-19 |  |  |  |  |  |  |  |  |  |  |  |  |  |
| de Bruijn, 2023 | Yes | Yes | Yes | Yes | Yes | No | No | Yes | Yes | No | Yes | Low risk of bias | Pre-print |
| Lower prevalence of Post-Covid-19 Condition following Omicron SARS-CoV-2 infection |  |  |  |  |  |  |  |  |  |  |  |  |  |
| Emecen, 2023 | No | Yes | Yes | Yes | Yes | Yes | Yes | Yes | Yes | NR | Yes | Moderate risk of bias | Single-centre cohort study (no control group), number of participants vaccinated/unvaccinated at each time point not well reported, unclear how study dealt with loss to follow-up |
| The presence of symptoms within 6 months after COVID‑19: a single‑center longitudinal study |  |  |  |  |  |  |  |  |  |  |  |  |  |
| Ertesvag, 2023 | Yes | Yes | Yes | Yes | Yes | No | Yes | Yes | Yes | NR | Yes | Low risk of bias |  |
| Post COVID-19 condition after delta infection and omicron reinfection in children and adolescents |  |  |  |  |  |  |  |  |  |  |  |  |  |
| Fernández-de-las-Peñas, 2022 | Yes | Yes | Yes | NR | NR | Yes | Yes | Yes | Yes | NR | Yes | Low risk of bias | Results from the multivariate analysis not reported; only stated they were not significant |
| Differences in Long-COVID Symptoms between Vaccinated and Non-Vaccinated (BNT162b2 Vaccine) Hospitalized COVID-19 Survivors Infected with the Delta Variant |  |  |  |  |  |  |  |  |  |  |  |  |  |
| Gallant, 2023 | Yes | Yes | Yes | Yes | Yes | No | Yes | Yes | NR | NR | Yes | Low risk of bias |  |
| SARS-CoV-2 infection outcomes associated with the Delta variant: A prospective cohort study |  |  |  |  |  |  |  |  |  |  |  |  |  |
| Hajjaji, 2022 | Yes | Yes | Yes | NR | No | NR | NR | Yes | No | No | Yes | Moderate risk of bias | Missing report of study methodology |
| 16 Months Follow Up of Patients’ Behavior and Mild COVID-19 Patterns in a Large Cohort of Cancer Patients During the Pandemic |  |  |  |  |  |  |  |  |  |  |  |  |  |
| Hastie, 2022 | Yes | Yes | Yes | Yes | Yes | Yes | Yes | Yes | Yes | NA | Yes | Low risk of bias |  |
| Outcomes among confirmed cases and a matched comparison group in the Long-COVID in Scotland study |  |  |  |  |  |  |  |  |  |  |  |  |  |
| Haupert, 2023 | Yes | Yes | Yes | Yes | Yes | Yes | Yes | Yes | NA | NA | Yes | Low risk of bias |  |
| A Case-Crossover Phenome-wide association study (PheWAS) for understanding Post-COVID-19 diagnosis patterns |  |  |  |  |  |  |  |  |  |  |  |  |  |
| Herman, 2022 | Yes | Yes | Yes | Yes | Yes | Yes | Yes | Yes | Yes | NA | Yes | Low risk of bias |  |
| Vaccination status, favipiravir, and micronutrient supplementation roles in post-COVID symptoms: A longitudinal study |  |  |  |  |  |  |  |  |  |  |  |  |  |
| Ioaanou, 2022 | Yes | Yes | Yes | Yes | Yes | Yes | Yes | Yes | NA | NA | Yes | Low risk of bias |  |
| Rates and Factors Associated With Documentation of Diagnostic Codes for Long COVID in the National Veterans Affairs Health Care System |  |  |  |  |  |  |  |  |  |  |  |  |  |
| Jacobs, 2023 | Yes | NR | NR | Yes | Yes | Yes | Yes | NR | NR | NR | Yes | Moderate risk of bias | How COVID-19 was identified was not reported, follow-up/ drop out of study not detailed |
| Racial, ethnic, and sex disparities in the incidence and cognitive symptomology of long COVID-19 |  |  |  |  |  |  |  |  |  |  |  |  |  |
| Jassat, 2023 | Yes | Yes | Yes | Yes | Yes | NR | Yes | Yes | Yes | No | Yes | Low risk of bias |  |
| A cohort study of post-COVID-19 condition across the Beta, Delta, and Omicron waves in South Africa: 6-month follow-up of hospitalized and nonhospitalized participants |  |  |  |  |  |  |  |  |  |  |  |  |  |
| Kim, 2023 | Yes | Yes | Yes | Yes | Yes | No | Yes | Yes | Yes | NA | Yes | Low risk of bias |  |
| Long COVID prevalence and impact on quality of life 2 years after acute COVID‑19 |  |  |  |  |  |  |  |  |  |  |  |  |  |
| Lam, 2023 | Yes | Yes | Yes | Yes | Yes | NR | Yes | Yes | NA | NA | Yes | Low risk of bias | Quality assessment completed for HKHA and UKB analyses separately. |
| Long-term post-acute sequelae of COVID-19 infection: a retrospective, multi-database cohort study in Hong Kong and the UK |  |  |  |  |  |  |  |  |  |  |  |  |  |
| Luo, 2023 | Yes | Yes | Yes | Yes | No | Yes | Yes | Yes | NA | NA | Yes | Low risk of bias |  |
| Prevalence and risk factors of long COVID 6–12 months after infection with the Omicron variant among non-hospitalized patients in Hong Kong |  |  |  |  |  |  |  |  |  |  |  |  |  |
| Mez-Torres, 2022 | Yes | Yes | Yes | Yes | Yes | Yes | No | Yes | NR | NR | Yes | Low risk of bias |  |
| Differences in Clinical Presentation With Long COVID After Community and Hospital Infection and Associations With All-Cause Mortality: English Sentinel Network Database Study |  |  |  |  |  |  |  |  |  |  |  |  |  |
| Mizrahi, 2023 | Yes | Yes | Yes | Yes | Yes | Yes | Yes | Yes | Yes | NA | Yes | Low risk of bias |  |
| Long covid outcomes at one year after mild SARS-CoV-2 infection: nationwide cohort study |  |  |  |  |  |  |  |  |  |  |  |  |  |
| Mohr, 2023 | Yes | Yes | Yes | Yes | Yes | Yes | NR | Yes | Yes | Yes | Yes | Low risk of bias |  |
| Presence of symptoms 6 weeks after COVID-19 among vaccinated and unvaccinated US healthcare personnel: a prospective cohort study |  |  |  |  |  |  |  |  |  |  |  |  |  |
| Munipalli, 2023 | Yes | Yes | Yes | Yes | No | Yes | Yes | NR | Yes | NA | Yes | Low risk of bias |  |
| Clinical differences in symptomology, characteristics, and risk factors in patients with post-acute sequelae of COVID-19: an experience from a tertiary-care academic center |  |  |  |  |  |  |  |  |  |  |  |  |  |
| Nascimento, 2023 | No | Yes | Yes | Yes | NR | Yes | Yes | Yes | Yes | NA | Yes | Low risk of bias |  |
| Vaccination status and long COVID symptoms in patients discharged from hospital |  |  |  |  |  |  |  |  |  |  |  |  |  |
| Naveed, 2023 | Yes | Yes | Yes | Yes | Yes | Yes | Yes | Yes | NR | NR | Yes | Low risk of bias |  |
| Association of COVID-19 Infection With Incident Diabetes |  |  |  |  |  |  |  |  |  |  |  |  |  |
| Parodi, 2023 | Yes | Yes | Yes | Yes | Yes | Yes | Yes | NR | NR | NR | Yes | Low risk of bias | Total population number reported incorrectly throughout the study. |
| Impact of COVID-19 vaccination in post-COVID cardiac complications |  |  |  |  |  |  |  |  |  |  |  |  |  |
| Patel, 2023 | Yes | Yes | Yes | Yes | Yes | Yes | Yes | No | Yes | NR | Yes | Low risk of bias | Time between COVID-19 infection and survey was shorter among those with breakthrough infection |
| Impact of vaccination on postacute sequelae of SARS CoV-2 infection in patients with rheumatic diseases. |  |  |  |  |  |  |  |  |  |  |  |  |  |
| Peghin, 2022 | Yes | Yes | Yes | Yes | Yes | Yes | Yes | Yes | Yes | NR | Yes | Low risk of bias |  |
| Post COVID-19 syndrome and humoral response association after 1 year in vaccinated and unvaccinated patients |  |  |  |  |  |  |  |  |  |  |  |  |  |
| Peghin, 2023 | Yes | Yes | Yes | Yes | Yes | Yes | Yes | Yes | NR | NR | Yes | Low risk of bias |  |
| Post-COVID-19 Syndrome 2 Years After the First Wave: The Role of Humoral Response, Vaccination and Reinfection |  |  |  |  |  |  |  |  |  |  |  |  |  |
| Pinato, 2022 | Yes | Yes | Yes | No | No | No | Yes | NR | Yes | NR | Yes | Moderate risk of bias | Confounding factors identified for other analyses but not for COVID-19 sequelae according to vaccination status. Follow-up period not specified. |
| Vaccination against SARS-CoV-2 protects from morbidity, mortality and sequelae from COVID19 in patients with cancer |  |  |  |  |  |  |  |  |  |  |  |  |  |
| Ranucci , 2023 | Yes | Yes | Yes | Yes | NR | No | Yes | Yes | Yes | NA | Yes | Low risk of bias |  |
| The Very Long COVID: Persistence of Symptoms after 12–18 Months from the Onset of Infection and Hospitalization |  |  |  |  |  |  |  |  |  |  |  |  |  |
| Richard, 2023 | Yes | Yes | Yes | Yes | Yes | Yes | Yes | Yes | No | No | Yes | Low risk of bias | Loss to follow-up not explored but steps taken to minimize risk of incomplete data |
| Persistent COVID-19 Symptoms at 6 Months After Onset and the Role of Vaccination Before or After SARS-CoV-2 Infection |  |  |  |  |  |  |  |  |  |  |  |  |  |
| Smits , 2023 | Yes | Yes | Yes | Yes | Yes | Yes | Yes | NR | Yes | NA | Yes | Low risk of bias |  |
| Effect of vaccination on time till Long COVID, a comparison of two ways to model effect of vaccination and two outcome definitions |  |  |  |  |  |  |  |  |  |  |  |  |  |
| Tannous, 2023 | Yes | Yes | Yes | Yes | Yes | Yes | Yes | Yes | Yes | NA | Yes | Low risk of bias |  |
| Real-world effectiveness of COVID-19 vaccines and anti-SARS-CoV-2 monoclonal antibodies against postacute sequelae of SARS-CoV-2: analysis of a COVID-19 observational registry for a diverse US metropolitan population |  |  |  |  |  |  |  |  |  |  |  |  |  |
| Taquet, 2022 | Yes | Yes | Yes | Yes | Yes | NR | Yes | Yes | NA | NA | Yes | Low risk of bias | Confounders addressed; unavoidable confounding factors stated |
| Six-month sequelae of post-vaccination SARS-CoV-2 infection: A retrospective cohort study of 10,024 breakthrough infections |  |  |  |  |  |  |  |  |  |  |  |  |  |
| Tosi, 2023 | Yes | Yes | Yes | No | No | NR | Yes | Yes | NA | NA | Yes | Low risk of bias |  |
| Vaccination as a protective factor for PASC in Frail, Pre-frail, and Robust Veterans During the Omicron Wave |  |  |  |  |  |  |  |  |  |  |  |  |  |
| Tran, 2023 | Yes | Yes | Yes | Yes | Yes | Yes | Yes | Yes | Yes | NA | Yes | Low risk of bias |  |
| Efficacy of first dose of covid-19 vaccine versus no vaccination on symptoms of patients with long covid: target trial emulation based on ComPaRe e-cohort. |  |  |  |  |  |  |  |  |  |  |  |  |  |
| Tsuchida, 2022 | Yes | Yes | NR | No | No | No | Yes | No | NA | NA | Yes | Moderate risk of bias | Limited description of study methods and included patients. Follow-up was only 2 weeks post vaccination, and no confounding factors were identified or adjusted for. |
| Relationship between changes in symptoms and antibody titers after a single vaccination in patients with Long COVID |  |  |  |  |  |  |  |  |  |  |  |  |  |
| van der Maaden, 2023 | Yes | Yes | Yes | Yes | NR | Yes | NR | Yes | Yes | Yes | Yes | Low risk of bias |  |
| Prevalence and Severity of Symptoms 3 Months After Infection With SARS-CoV-2 Compared to Test-Negative and Population Controls in the Netherlands |  |  |  |  |  |  |  |  |  |  |  |  |  |
| Whittaker, 2021 | Yes | Yes | Yes | Yes | Yes | Yes | Yes | Yes | NR | NR | Yes | Low risk of bias |  |
| GP consultation rates for sequelae after acute covid-19 in patients managed in the community or hospital in the UK: population based study |  |  |  |  |  |  |  |  |  |  |  |  |  |
| Wisnivesky, 2022 | Yes | Yes | Yes | Yes | Yes | NR | Yes | Yes | Yes | No | Yes | Low risk of bias |  |
| Association of Vaccination with the Persistence of Post-COVID Symptoms |  |  |  |  |  |  |  |  |  |  |  |  |  |
| Wynberg, 2023 | Yes | Yes | Yes | NR | NR | No | Yes | Yes | NR | NR | Yes | Low risk of bias |  |
| The effect of SARS-CoV-2 vaccination on post-acute sequelae of COVID-19 (PASC): A prospective cohort study |  |  |  |  |  |  |  |  |  |  |  |  |  |
| Zhang , 2023 | Yes | Yes | Yes | Yes | Yes | Yes | Yes | Yes | Yes | NA | Yes | Low risk of bias |  |
| Determinants of post-COVID-19 symptoms among adults aged 55 or above with chronic conditions in primary care: data from a prospective cohort in Hong Kong |  |  |  |  |  |  |  |  |  |  |  |  |  |
| Zisis, 2022 | Yes | Yes | Yes | Yes | Yes | Yes | Yes | Yes | NR | NR | Yes | Low risk of bias |  |
| The Protective Effect of Coronavirus Disease 2019 (COVID-19) Vaccination on Postacute Sequelae of COVID-19: A Multicenter Study From a Large National Health Research Network |  |  |  |  |  |  |  |  |  |  |  |  |  |

**eTable 5. Summary of JBI Critical Appraisal ratings: Cross-sectional Study design**

| **Primary author**  **Study Title** | Were the criteria for inclusion in the sample clearly defined? | Were the study subjects and the setting described in detail? | Was the exposure measured in a valid and reliable way? | Were objective, standard criteria used for measurement of the condition? | Were confounding factors identified? | Were strategies to deal with confounding factors stated? | Were the outcomes measured in a valid and reliable way? | Was appropriate statistical analysis used? | **Appraisal** | **Rationale for critical appraisal rating** |
| --- | --- | --- | --- | --- | --- | --- | --- | --- | --- | --- |
| Alghamdi, 2022 | Yes | Yes | Yes | Yes | No | NR | Yes | Yes | Low risk of bias | Study setting not described |
| Neuropsychiatric symptoms in post-COVID-19 long haulers |  |  |  |  |  |  |  |  |  |  |
| Alradh, 2022 | Yes | No | NR | NR | No | No | Yes | Yes | Moderate risk of bias | Unclear how the exposure was measured, confounding not explored in analysis |
| A comparative study to the persistence post COVID syndrome and the infection period between vaccinated and non-vaccinated patients in Al Basrah Province |  |  |  |  |  |  |  |  |  |  |
| Angarita-Fonseca, 2023 | Yes | No | NR | No | No | No | NR | Yes | High risk of bias | Participants were able to complete the survey multiple times, no confounding variables were identified. |
| Exploring long COVID condition in Latin America: Its impact on patients' activities and associated healthcare use |  |  |  |  |  |  |  |  |  |  |
| Arjun, 2022 | Yes | Yes | Yes | Yes | Yes | Yes | Yes | Yes | Low risk of bias |  |
| Characteristics and predictors of Long COVID among diagnosed cases of COVID-19 |  |  |  |  |  |  |  |  |  |  |
| Arjun, 2022 | No | Yes | Yes | Yes | Yes | NR | Yes | Yes | Low risk of bias |  |
| Long COVID following Omicron wave in Eastern India—A retrospective cohort study |  |  |  |  |  |  |  |  |  |  |
| Bello-Chavolla , 2023 | No | Yes | Yes | Yes | Yes | Yes | Yes | Yes | Low risk of bias |  |
| Nationally representative prevalence and determinants of post-acute sequelae of SARS-CoV-2 infection (Long COVID) amongst Mexican adults in 2022 |  |  |  |  |  |  |  |  |  |  |
| Bostanci, 2023 | No | Yes | Yes | Yes | Yes | NR | Yes | Yes | Low risk of bias |  |
| Long-COVID-19 in Asymptomatic, Non-Hospitalized, and Hospitalized Populations: A Cross-Sectional Study |  |  |  |  |  |  |  |  |  |  |
| Brunvoll, 2023 | Yes | Yes | Yes | Yes | NR | Yes | Yes | Yes | Low risk of bias |  |
| Post-acute symptoms 3-15 months after COVID-19 among unvaccinated and vaccinated individuals with a breakthrough infection |  |  |  |  |  |  |  |  |  |  |
| Chelly, 2023 | Yes | Yes | No | No | No | No | NR | Yes | Moderate risk of bias | No diagnostic test to confirm acute COVID-19, limited detail of study methodologies |
| Symptoms and risk factors for long COVID in Tunisian population |  |  |  |  |  |  |  |  |  |  |
| Di Fusco, 2023 | Yes | Yes | Yes | Yes | Yes | Yes | Yes | Yes | Low risk of bias |  |
| Impact of COVID-19 and effects of booster vaccination with BNT162b2 on six-month long COVID symptoms, quality of life, work productivity and activity impairment during Omicron |  |  |  |  |  |  |  |  |  |  |
| Durstenfeld, 2022 | No | Yes | No | NR | Yes | Yes | Yes | Yes | Low risk of bias | Study did not describe exclusion criteria and was NR as to how long COVID was defined |
| Factors Associated With Long COVID Symptoms in an Online Cohort Study |  |  |  |  |  |  |  |  |  |  |
| Kuodi, 2023 | Yes | Yes | Yes | Yes | Yes | Yes | Yes | Yes | Low risk of bias |  |
| Association between BNT162b2 vaccination and health-related quality of life up to 18 months post-SARS-CoV-2 infection in Israel: A cross sectional survey |  |  |  |  |  |  |  |  |  |  |
| Elias , 2023 | Yes | No | No | Yes | Yes | Yes | Yes | Yes | Low risk of bias |  |
| Determinants of Post-acute COVID-19 Syndrome among hospitalized severe COVID-19 patients: a 2-year follow up study |  |  |  |  |  |  |  |  |  |  |
| Elmazny, 2023 | Yes | No | Yes | Yes | No | No | NR | Yes | Low risk of bias |  |
| Neuropsychiatric post‑acute sequelae of COVID‑19: prevalence, severity, and impact of vaccination |  |  |  |  |  |  |  |  |  |  |
| Elnadi, 2023 | Yes | No | No | No | Yes | No | No | Yes | High risk of bias | Unclear study sample demographics and inclusion in analyses |
| Prevalence of Post COVID-19 Condition among healthcare workers: self-reported 1 online survey in four African countries |  |  |  |  |  |  |  |  |  |  |
| Gaber, 2020 | No | No | NR | NR | No | No | Yes | NR | High risk of bias | No information on participant characteristics, study does not describe how long COVID was diagnosed. |
| Are mRNA Covid 19 vaccines safe in Long Covid patients? A Health Care Workers perspective |  |  |  |  |  |  |  |  |  |  |
| Hirahata, 2022 | Yes | No | NR | Yes | Yes | Yes | Yes | Yes | Low risk of bias |  |
| Characteristics of Long COVID: Cases from the First to the Fifth Wave in Greater Tokyo, Japan |  |  |  |  |  |  |  |  |  |  |
| Jaramillo, 2023 | Yes | Yes | Yes | Yes | Yes | NR | Yes | Yes | Low risk of bias |  |
| Characteristics of Chemosensory Perception in Long COVID and COVID Reinfection |  |  |  |  |  |  |  |  |  |  |
| Kahlert, 2023 | NR | Yes | Yes | NR | Yes | Yes | Yes | Yes | Low risk of bias |  |
| Post-Acute Sequelae After Severe Acute Respiratory Syndrome Coronavirus 2 Infection by Viral Variant and Vaccination Status: A Multicenter Cross-Sectional Study |  |  |  |  |  |  |  |  |  |  |
| Kubota, 2023 | Yes | Yes | Yes | Yes | No | NR | Yes | NR | Low risk of bias | Adjusted analyses not conducted to account for confounding factors. |
| Pain paths among post-COVID-19 condition subjects: A prospective cross-sectional study with in-person evaluation |  |  |  |  |  |  |  |  |  |  |
| Kuodi, 2022 | Yes | Yes | Yes | No | Yes | Yes | Yes | Yes | Low risk of bias |  |
| Association between BNT162b2 vaccination and reported incidence of post-COVID-19 symptoms: cross-sectional study 2020-21, Israel |  |  |  |  |  |  |  |  |  |  |
| Lippke, 2023 | No | No | Yes | NR | Yes | NR | Yes | Yes | Moderate risk of bias | Criteria reported in previous study. Only age/ sex reported for participant demographics. No comorbidities and clear definition for PASC reported |
| Patients’ Post-/Long-COVID Symptoms, Vaccination and Functional Status—Findings from a State-Wide Online Screening Study |  |  |  |  |  |  |  |  |  |  |
| McLaughlin, 2023a | Yes | Yes | Yes | Yes | Yes | NR | Yes | Yes | Low risk of bias |  |
| A Cross-Sectional Study of Symptom Prevalence, Frequency, Severity, and Impact of Long-COVID in Scotland: Part I |  |  |  |  |  |  |  |  |  |  |
| McLaughlin, 2023b | Yes | Yes | Yes | Yes | Yes | NR | Yes | Yes | Low risk of bias |  |
| A Cross-Sectional Study of Symptom Prevalence, Frequency, Severity, and Impact of Long-COVID in Scotland: Part II |  |  |  |  |  |  |  |  |  |  |
| Nehme, 2022 | Yes | No | Yes | NR | Yes | Yes | NR | Yes | Low risk of bias |  |
| Symptoms After COVID-19 Vaccination in Patients with Post-Acute Sequelae of SARS-CoV-2 |  |  |  |  |  |  |  |  |  |  |
| Nehme, 2022 | Yes | Yes | Yes | Yes | Yes | Yes | Yes | Yes | Low risk of bias |  |
| Prevalence of Post-Coronavirus Disease Condition 12 Weeks After Omicron Infection Compared With Negative Controls and Association With Vaccination Status |  |  |  |  |  |  |  |  |  |  |
| Perlis, 2022 | NR | Yes | Yes | Yes | Yes | Yes | Yes | Yes | Low risk of bias |  |
| Prevalence and Correlates of Long COVID Symptoms Among US Adults |  |  |  |  |  |  |  |  |  |  |
| Qasmieh, 2023 | No | Yes | No | NR | Yes | Yes | NR | Yes | Moderate risk of bias | Lacking detail on study inclusion criteria, demographics and methodology |
| The prevalence of SARS-CoV-2 infection and long COVID in U.S. adults during the BA.4/BA.5 surge, June–July 2022 |  |  |  |  |  |  |  |  |  |  |
| Rahman, 2023 | Yes | No | Yes | Yes | No | No | NR | No | High risk of bias | No description of baseline demographics and limited methodology |
| Prevalence and difference of COVID-19 symptoms, post-COVID conditions and duration of illness among the vaccinated and unvaccinated population: a cross-sectional study in Peshawar |  |  |  |  |  |  |  |  |  |  |
| Robertson, 2022 | No | NR | Yes | Yes | Yes | Yes | Yes | Yes | Low risk of bias |  |
| The Epidemiology of Long Coronavirus Disease in US Adults |  |  |  |  |  |  |  |  |  |  |
| Scherlinger, 2022 | Yes | Yes | Yes | Yes | No | No | Yes | Yes | Low risk of bias |  |
| Effect of SARS-CoV-2 Vaccination on Symptoms from Post-Acute Sequelae of COVID-19: Results from the Nationwide VAXILONG Study |  |  |  |  |  |  |  |  |  |  |
| Senjam , 2022 | No | Yes | Yes | Yes | Yes | Yes | Yes | Yes | Low risk of bias |  |
| A Comprehensive Assessment of Self-Reported Post COVID-19 Symptoms Among Beneficiaries of Hospital Employee Scheme at a Tertiary Healthcare Institution in Northern India |  |  |  |  |  |  |  |  |  |  |
| Strain, 2022 | Yes | No | Yes | Yes | Yes | Yes | Yes | Yes | Low risk of bias |  |
| The Impact of COVID Vaccination on Symptoms of Long COVID: An International Survey of People with Lived Experience of Long COVID |  |  |  |  |  |  |  |  |  |  |
| Wanga, 2021 | Yes | No | No | NR | Yes | NR | Yes | Yes | High risk of bias | Self-reported COVID test status, evidence of specific test (e.g., PCR) not required. Respondent-reported symptoms, no limit on follow-up period nor consideration of circulating SARS-CoV-2 variants. Lacking key baseline characteristics (e.g., comorbidities, risk factors such as smoker status) |
| Long-Term Symptoms Among Adults Tested for SARS-CoV-2 — United States, January 2020–April 2021 |  |  |  |  |  |  |  |  |  |  |
| Wong, 2023 | Yes | Yes | No | NR | Yes | Yes | No | Yes | Moderate risk of bias | Sample was greater than 100% as respondents were allowed to respond multiple times. No diagnostic test confirmed COVID-19. |
| Epidemiology, Symptomatology, and Risk Factors for Long COVID Symptoms: Population-Based, Multicenter Study |  |  |  |  |  |  |  |  |  |  |
| Xue, 2023 | Yes | Yes | Yes | Yes | Yes | Yes | Yes | Yes | Low risk of bias |  |
| Persistent short nighttime sleep duration is associated with a greater post-COVID risk in fully mRNA-vaccinated individuals |  |  |  |  |  |  |  |  |  |  |

**eTable 6. Summary of JBI Critical Appraisal ratings: Case-control Study design**

| **Primary author**  **Study Title** | Were the groups comparable other than the presence of disease in cases or the absence of disease in controls? | Were cases and controls matched appropriately? | Were the same criteria used for identification of cases and controls? | Was exposure measured in a standard, valid and reliable way? | Was exposure measured in the same way for cases and controls? | Were confounding factors identified? | Were strategies to deal with confounding factors stated? | Were outcomes assessed in a standard, valid and reliable way for cases and controls? | Was the exposure period of interest long enough to be meaningful? | Was appropriate statistical analysis used? | Appraisal | **Rationale for critical appraisal rating** |
| --- | --- | --- | --- | --- | --- | --- | --- | --- | --- | --- | --- | --- |
| Antonelli, 2022 | Yes | Yes | Yes | Yes | Yes | Yes | Yes | Yes | Yes | Yes | Low risk of bias |  |
| Risk factors and disease profile of post-vaccination SARS-CoV-2 infection in UK users of the COVID Symptom Study app: a prospective, community-based, nested, case-control study |  |  |  |  |  |  |  |  |  |  |  |  |
| Antonelli, 2022 | Yes | No | Yes | Yes | Yes | Yes | Yes | Yes | Yes | Yes | Low risk of bias |  |
| Risk of long COVID associated with delta versus omicron variants of SARS-CoV-2 |  |  |  |  |  |  |  |  |  |  |  |  |
| El Otmani, 2022 | Yes | Yes | Yes | Yes | Yes | Yes | Yes | Yes | NR | Yes | Low risk of bias |  |
| Demography, comorbidities, severity and duration of the Covid-19 symptoms, and vaccination against SARS CoV-2 |  |  |  |  |  |  |  |  |  |  |  |  |
| Marra, 2023 | Yes | Yes | Yes | Yes | Yes | NR | NR | Yes | Yes | Yes | Low risk of bias |  |
| Risk factors for long coronavirus disease 2019 (long COVID) among healthcare personnel, Brazil, 2020-2022 |  |  |  |  |  |  |  |  |  |  |  |  |
| Resendez, 2023 | NR | Yes | Yes | Yes | Yes | Yes | Yes | Yes | Yes | Yes | Low risk of bias |  |
| Defining the Subtypes of Long COVID and Risk Factors for Prolonged Disease |  |  |  |  |  |  |  |  |  |  |  |  |

**eTable 7. Study characteristics for 97 included studies and the 56 studies with low risk of bias and adjusted estimates for the relationship between vaccination and PCC**

| **Study Characteristics** | **Included Studies (n=97)** | **Studies with low risk of bias and reporting adjusted estimates for the relationship between vaccination and PCC (N=56)** |
| --- | --- | --- |
| **Study design *N (%)*** | | |
| Cross-sectional | 35 (36.1) | 18 (32.1) |
| Retrospective cohort | 29 (29.9) | 20 (35.7) |
| Prospective cohort | 26 (26.8) | 14 (25.0) |
| Case-control | 6 (6.2) | 3 (5.4) |
| Ambidirectional cohort | 1 (1.0) | 1 (1.8) |
| **WHO Region *N (%)*** | | |
| Europe | 41 (42.3) | 22 (39.3) |
| Americas | 31 (32.0) | 21 (37.5) |
| Eastern Mediterranean | 7 (7.2) | 2 (3.6) |
| Western Pacific | 7 (7.2) | 3 (5.4) |
| South-East Asia | 4 (4.1) | 4 (7.1) |
| Africa | 3 (3.1) | 1 (1.8) |
| Global (i.e., multiple WHO regions) | 4 (4.1) | 3 (5.4) |
| **Predominant variant period *N (%)*** | |  |
| Pre-Omicron | 41 (42.3) | 26 (46.4) |
| Omicron | 15 (15.5) | 8 (14.3) |
| Both | 34 (35.1) | 21 (37.5) |
| Could not be determined | 7 (7.2) | 1 (1.8) |
| **PCC data source *N (%)*** | | |
| Patient-reported | 67 (69.1) | 37 (66.1) |
| Medical diagnostic codes/ claims | 29 (29.9) | 18 (32.1) |
| Both | 1 (1.0) | 1 (1.8) |
| **Age group *N (%)*** | | |
| Adults | 78 (80.4) | 46 (82.1) |
| Adults and adolescents (≥12 years) | 15 (15.5) | 8 (14.3) |
| Children and adolescents (≥12 years) | 1 (1.0) | 1 (1.8) |
| NR | 3 (3.1) | 1 (1.8) |
| **Vaccination type *N (%)*** | | |
| Mixed mRNA and non-mRNA | 48 (49.5) | 27 (48.2) |
| mRNA only | 18 (18.6) | 15 (26.8) |
| Mixed (NR) | 4 (4.1) | 3 (5.4) |
| Non-mRNA only | 3 (3.1) | 2 (3.6) |
| NR | 24 (24.7) | 9 (16.1) |
| **Risk of bias assessment *N (%)*** | | |
| Low risk of bias | 80 (82.5) | 56 (100.0) |
| Moderate risk of bias | 12 (12.4) | 0 (0.0) |
| High risk of bias | 5 (5.2) | 0 (0.0) |
| **Vaccination timing relative to infection^a^** | | |
| Before infection | 55 (56.7) | 39 (68.4) |
| After infection | 10 (10.3) | 4 (7.1) |
| After PCC onset | 6 (6.2) | 3 (5.3) |
| Mixed vaccination timing | 11 (11.3) | 7 (10.5) |
| Not Reported | 18 (18.6) | 6 (10.5) |

mRNA: messenger ribonucleic acid; NR: not reported; PCC: post‑COVID‑19 conditions; WHO: World Health Organization.

^a^Durstenfeld, 2022 and Luo, 2023 reported estimates for both before infection and after infection; Richard, 2023 reported estimates for both before infection and after PCC onset. Therefore, the total number of studies here exceeds that included in the overall SLR (n=97) and those at low risk of bias reporting adjusted VE (n=56).

**eFigure 1. Comparative aVE against ≥1 PCC (reference group other than unvaccinated), by groups being compared and timing of vaccination relative to infection or PCC onset**

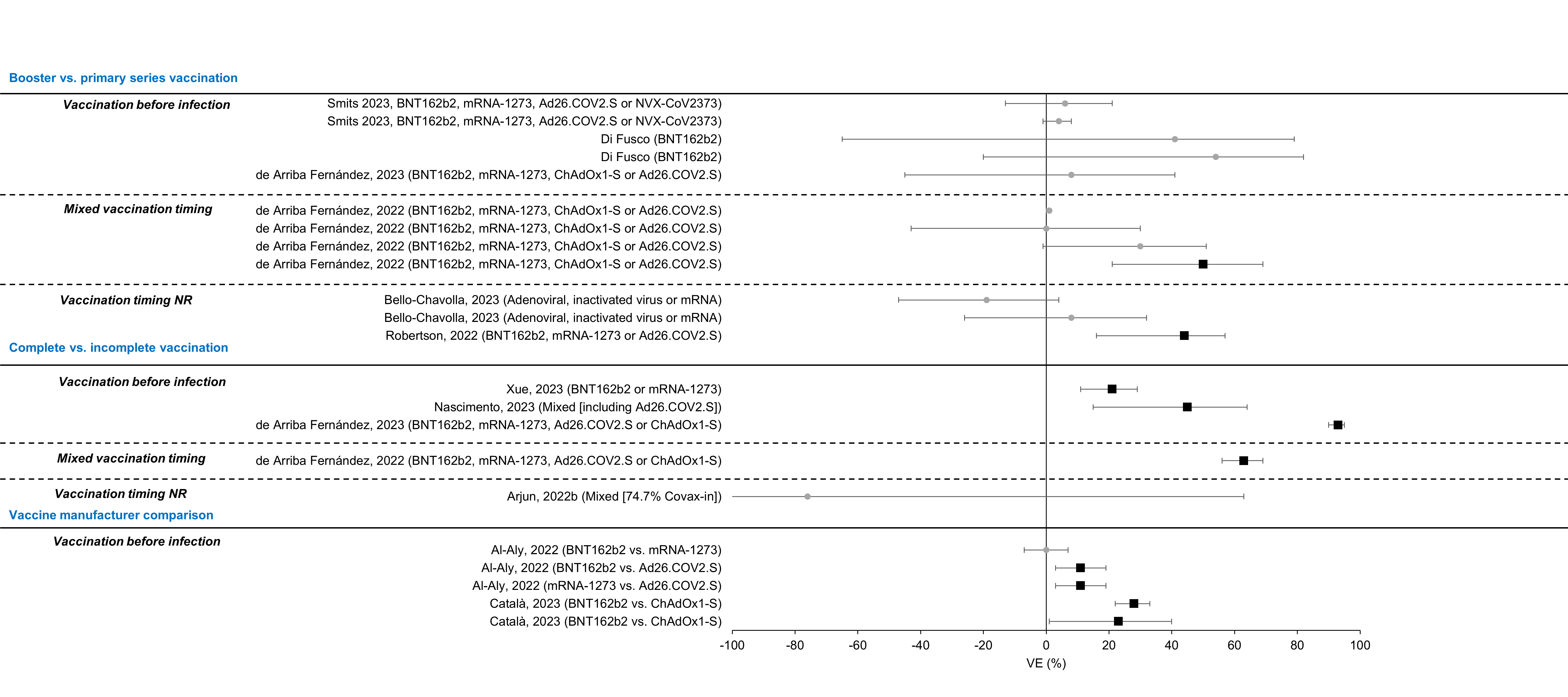

Of 22 VE estimates, 10 (45.5%) reported statistically significant (based on VE 95% CI) protective effect against ≥1 PCC.

CI: confidence interval; NR: not reported; PCC: post‑COVID‑19 conditions; VE: vaccine effectiveness.

References: Al-Aly, 2022^2^; Arjun, 2022b^9^; Bello-Chavolla, 2023^16^; Català, 2023^22^; de Arriba Fernández, 2022^26^; de Arriba Fernández, 2023^27^; Di Fusco^29^; Nascimento, 2023^64^; Robertson, 2022^79^; Smits 2023^82^; Xue, 2023^95^

**eTable 8. Summary of publications where adjusted comparative VE against ≥1 PCC (reference group other than unvaccinated) could be computed, by groups being compared and timing of vaccination relative to infection or PCC onset (N=11 studies with 22 VE estimates)**

| **Author, year (Vaccine, dose)** | **Comparison** | **Vaccination timepoints** | **Variant** | **Subgroup** | **Subgroup details** | **Adjustments** | **VE** | **Lower 95% CI** | **Upper 95% CI** |
| --- | --- | --- | --- | --- | --- | --- | --- | --- | --- |
| Smits 2023, BNT162b2, mRNA-1273, Ad26.COV2.S or NVX-CoV2373, booster vs. primary series) | Booster vs primary | Before infection | Omicron | Vaccination status | ICD-10 code for long COVID diagnosis (CDC definition) | Adjusted for vaccination status, comorbidities, sex, race, ethnicity, age, timing of infection, smoking status, influenza vaccination status, inpatient and outpatient encounters in previous two years | 6 | -13 | 21 |
| Smits 2023, BNT162b2, mRNA-1273, Ad26.COV2.S or NVX-CoV2373, booster vs. primary series) | Booster vs primary | Before infection | Omicron | Vaccination status | ICD-10 codes for any long COVID symptom (CDC definition) | Adjusted for vaccination status, comorbidities, sex, race, ethnicity, age, timing of infection, smoking status, influenza vaccination status, inpatient and outpatient encounters in previous two years | 4 | -1 | 8 |
| Di Fusco (BNT162b2, booster vs. primary) | Booster vs primary | Before infection | Omicron | Number of symptoms | ≥3 symptoms | Adjusted for timepoint, vaccination status and interaction of time by vaccination status, participant pre-COVID-19 symptom onset score, sociodemographic characteristics (age, sex, regions, social vulnerability, race/ethnicity, high risk occupations), previously tested positive for COVID-19, severity of acute illness and immunocompromised status | 41 | -65 | 79 |
| Di Fusco (BNT162b2, booster vs. primary series) | Booster vs primary | Before infection | Omicron | Number of symptoms | ≥2 symptoms | Adjusted for timepoint, vaccination status and interaction of time by vaccination status, participant pre-COVID-19 symptom onset score, sociodemographic characteristics (age, sex, regions, social vulnerability, race/ethnicity, high risk occupations), previously tested positive for COVID-19, severity of acute illness and immunocompromised status | 54 | -20 | 82 |
| de Arriba Fernández, 2023 (BNT162b2, mRNA-1273, ChAdOx1-S or Ad26.COV2.S, booster vs. no booster) | Booster vs no booster | Before infection | Delta, Omicron | NA | NA | Adjusted for age, sex, comorbidity history, body mass index, date of first, second and booster doses of a COVID-19 vaccine, type of COVID-19 vaccine, death, immunosuppressive treatment, diagnosis of persistent COVID-19, and diagnostic test results | 8 | -45 | 41 |
| de Arriba Fernández, 2022 (BNT162b2, mRNA-1273, ChAdOx1-S or Ad26.COV2.S, booster [3rd dose] vs. no booster [≤2 doses]) | Booster (3rd dose) vs no booster (≤2 doses) | Mixed | Delta, Omicron | Age group | 12-17 years old | Adjusted for age, sex, immunosuppressive treatment, analytical determinations (high and low density lipoprotein cholesterol, total cholesterol, triglycerides and fasting glucose levels), COVID-19 vaccine, complete vaccination schedule, booster dose (3rd dose) by age and comorbidities | 1 | 0 | 1 |
| de Arriba Fernández, 2022 (BNT162b2, mRNA-1273, ChAdOx1-S or Ad26.COV2.S, booster [3rd dose] vs. no booster [≤2 doses]) | Booster (3rd dose) vs no booster (≤2 doses) | Mixed | Delta, Omicron | Age group | 18-49 years old | Adjusted for age, sex, immunosuppressive treatment, analytical determinations (high and low density lipoprotein cholesterol, total cholesterol, triglycerides and fasting glucose levels), COVID-19 vaccine, complete vaccination schedule, booster dose (3rd dose) by age and comorbidities | 0 | -43 | 30 |
| de Arriba Fernández, 2022 (BNT162b2, mRNA-1273, ChAdOx1-S or Ad26.COV2.S, booster [3rd dose] vs. no booster [≤2 doses]) | Booster (3rd dose) vs no booster (≤2 doses) | Mixed | Delta, Omicron | Age group | 50-69 years old | Adjusted for age, sex, immunosuppressive treatment, analytical determinations (high and low density lipoprotein cholesterol, total cholesterol, triglycerides and fasting glucose levels), COVID-19 vaccine, complete vaccination schedule, booster dose (3rd dose) by age and comorbidities | 30 | -1 | 51 |
| de Arriba Fernández, 2022 (BNT162b2, mRNA-1273, ChAdOx1-S or Ad26.COV2.S, booster [3rd dose] vs. no booster [≤2 doses]) | Booster (3rd dose) vs no booster (≤2 doses) | Mixed | Delta, Omicron | Age group | ≥70 years old | Adjusted for age, sex, immunosuppressive treatment, analytical determinations (high and low density lipoprotein cholesterol, total cholesterol, triglycerides and fasting glucose levels), COVID-19 vaccine, complete vaccination schedule, booster dose (3rd dose) by age and comorbidities | 50 | 21 | 69 |
| Bello-Chavolla, 2023 (Adenoviral, inactivated virus or mRNA, booster vs. primary series) | Booster vs primary series | NR | Delta, Omicron | Long COVID definition | WHO definition | Adjusted by survey weights | -19 | -47 | 4 |
| Bello-Chavolla, 2023 (Adenoviral, inactivated virus or mRNA, booster vs. primary series) | Booster vs primary series | NR | Delta, Omicron | Long COVID definition | PASC defined by PASC score ≥12 points | Adjusted by survey weights | 8 | -26 | 32 |
| Robertson, 2022 (BNT162b2, mRNA-1273 or Ad26.COV2.S, booster vs. complete vaccination) | Booster vs complete vaccination (inverse) | NR | Omicron | NA | NA | Adjusted for gender and age | 44 | 16 | 57 |
| Xue, 2023 (BNT162b2 or mRNA-1273, 2 doses vs. 0 or 1 dose) | 2 doses vs 0 or 1 dose | Before infection | Alpha, Gamma, Delta, Omicron | NA | NA | Adjusted for severity of COVID-19, Chronic medication use, presence of chronic conditions, self-perceived health status, monthly household income, smoking status, living status, educational level, current occupation, marital status, gender and age | 21 | 11 | 29 |
| Nascimento, 2023 (Mixed [including Ad26.COV2.S], complete vs. no or incomplete vaccination) | Complete vs no or incomplete vaccination | Before infection | Gamma, Delta, Omicron | NA | NA | Adjusted for vaccination status, age, sex, diabetes, previous cardiovascular disease, chronic kidney disease, ICU admission, pulmonary embolism or deep vein thrombosis, and corticosteroid use | 45 | 15 | 64 |
| de Arriba Fernández, 2023 (BNT162b2, mRNA-1273, Ad26.COV2.S or ChAdOx1-S, complete vs. incomplete vaccination) | Complete vs incomplete vaccination | Before infection | Delta, Omicron | NA | NA | Adjusted for age, sex, comorbidity history, body mass index, date of first, second and booster doses of a COVID-19 vaccine, type of COVID-19 vaccine, death, immunosuppressive treatment, diagnosis of persistent COVID-19, and diagnostic test results | 93 | 90 | 95 |
| de Arriba Fernández, 2022 (BNT162b2, mRNA-1273, Ad26.COV2.S or ChAdOx1-S, complete vs. no or incomplete vaccination) | Complete vs incomplete or no vaccination | Mixed | Delta, Omicron | NA | NA | Adjusted for age, sex, immunosuppressive treatment, analytical determinations (high and low density lipoprotein cholesterol, total cholesterol, triglycerides and fasting glucose levels), COVID-19 vaccine, complete vaccination schedule, booster dose (3rd dose) by age and comorbidities | 63 | 56 | 69 |
| Arjun, 2022b (Mixed [74.7% Covax-in], 2 doses vs. 0 or 1 dose) | 2 doses vs 0 or 1 dose | NR | Omicron (assumed, based on population variant level of >92%) | NA | NA | Adjusted for age, sex, education, occupation, BMI, history of substance use, COVID-19 history, pre-existing medical conditions, acute COVID-19 symptoms and severity, and level of care received for acute COVID-19 | -76 | -728 | 63 |
| Al-Aly, 2022 (BNT162b2 vs. mRNA-1273, primary series) | BNT162b2 vs mRNA-1273 | Before infection | Wild-type, Alpha, Delta | NA | NA | Adjusted for age, race, ADI, the number in the two years prior to enrolment of outpatient visits, inpatient visits, unique medication prescriptions, and routine blood chemistry panels, BMI, history of influenza vaccination, smoking status, comorbidities, blood pressure, estimated glomerular filtration rate, and hospital system average hospital bed occupancy, total hospital bed capacity, positivity rate of tests, number of tests, and HHS region SARS-CoV-2 variant proportion in the week prior to enrolment, geographic region of VISN, calendar week of vaccination and enrolment, algorithmically selected high dimensional variables | 0 | -7 | 7 |
| Al-Aly, 2022 (BNT162b2 vs. Ad26.COV2.S, primary series) | BNT162b2 vs Ad26.COV2.S | Before infection | Wild-type, Alpha, Delta | NA | NA | Adjusted for age, race, ADI, the number in the two years prior to enrolment of outpatient visits, inpatient visits, unique medication prescriptions, and routine blood chemistry panels, BMI, history of influenza vaccination, smoking status, comorbidities, blood pressure, estimated glomerular filtration rate, and hospital system average hospital bed occupancy, total hospital bed capacity, positivity rate of tests, number of tests, and HHS region SARS-CoV-2 variant proportion in the week prior to enrolment, geographic region of VISN, calendar week of vaccination and enrolment, algorithmically selected high dimensional variables | 11 | 3 | 19 |
| Al-Aly, 2022 (mRNA-1273 vs. Ad26.COV2.S, primary series) | mRNA-1273 vs Ad26.COV2.S | Before infection | Wild-type, Alpha, Delta | NA | NA | Adjusted for age, race, ADI, the number in the two years prior to enrolment of outpatient visits, inpatient visits, unique medication prescriptions, and routine blood chemistry panels, BMI, history of influenza vaccination, smoking status, comorbidities, blood pressure, estimated glomerular filtration rate, and hospital system average hospital bed occupancy, total hospital bed capacity, positivity rate of tests, number of tests, and HHS region SARS-CoV-2 variant proportion in the week prior to enrolment, geographic region of VISN, calendar week of vaccination and enrolment, algorithmically selected high dimensional variables | 11 | 3 | 19 |
| Català, 2023 (BNT162b2 vs. ChAdOx1-S, 1 dose) | Pfizer versus AstraZeneca (1 dose) | Before infection | Delta, Omicron | Cohort | CPRD AURUM | Propensity score overlap weighting for age (5-year age bands for people aged 18 years and older); location (primary care practice, health-care region, and nation); index date; previous observation years; number of previous GP visits; number of previous SARS-CoV-2 PCR tests; and regional vaccination, testing, and COVID-19 incidence rates | 28 | 22 | 33 |
| Català, 2023 (BNT162b2 vs. ChAdOx1-S, 1 dose) | Pfizer versus AstraZeneca (1 dose) | Before infection | Delta, Omicron | Cohort | CPRD GOLD | Propensity score overlap weighting for age (5-year age bands for people aged 18 years and older); location (primary care practice, health-care region, and nation); index date; previous observation years; number of previous GP visits; number of previous SARS-CoV-2 PCR tests; and regional vaccination, testing, and COVID-19 incidence rates | 23 | 1 | 40 |

BMI: Body mass index; CI: Confidence interval; CDC: Centers for Disease Control and Prevention; CPRD: Clinical Practice Research Datalink; GP: General Practitioner; HHS: Health and Human Services; ICD-10: International Classification of Diseases- 10; ICU: Intensive care units; mRNA: Messenger ribonucleic acid; NA: Not applicable; NR: Not reported; PASC: Post-acute sequelae of COVID-19; PCR: Polymerase Chain Reaction; VE: Vaccine effectiveness; VISN: Veterans Integrated Services Network; WHO: World Health Organization.

**eFigure 2. Adjusted VE (vs. unvaccinated) against ≥1 specific Cardiovascular, Musculoskeletal, Neurological, and Endocrine endpoints, sorted by organ system and timing of vaccination relative to infection or PCC onset**

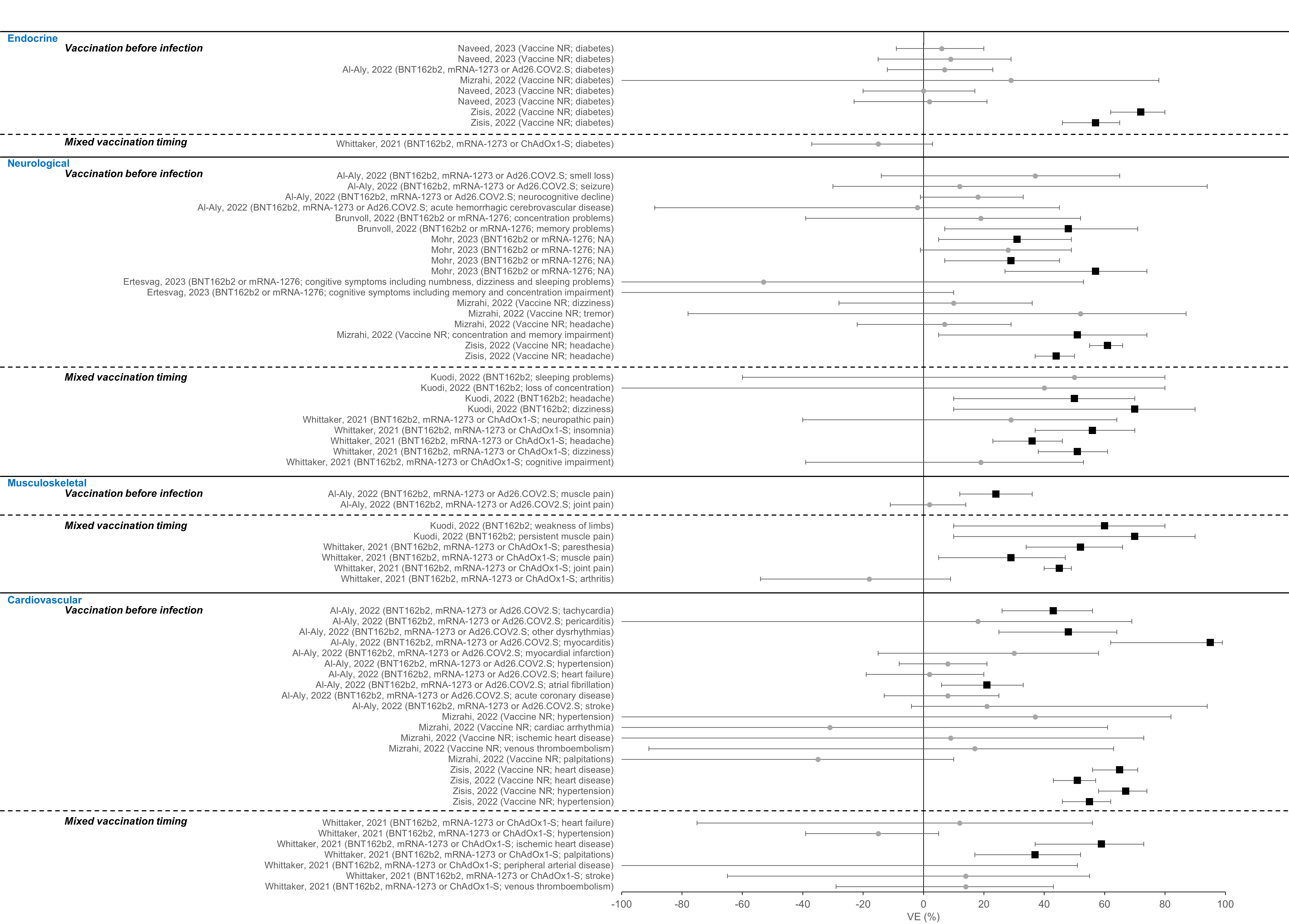

Of 71 VE estimates, 30 (42.3%) reported statistically significant (based on VE 95% CI) protective effect against ≥1 PCC.

CI: confidence interval; NR: not reported; PCC: post‑COVID‑19 conditions; VE: vaccine effectiveness.

References: Al-Aly, 2022^2^; Brunvoll, 2022^20^; Ertesvag, 2023^36^; Kuodi, 2022^52^; Mizrahi, 2023^61^; Mohr, 2023^62^; Naveed, 2023^65^; Whittaker, 2021^91^; Zisis, 2022^97^

**eTable 9. Summary of publications where adjusted VE (vs. unvaccinated) against ≥1 against specific Cardiovascular, Musculoskeletal, Neurological, and Endocrine PCC could be computed, sorted by organ system and timing of vaccination relative to infection or PCC onset and PCC (N=9 studies with 70 VE estimates)**

| **Author, year (Vaccine, dose)** | **Vaccination timepoint** | **PCC Organ System** | **Symptom** | **Variant** | **Subgroup** | **Subgroup details** | **Adjustments** | **VE** | **Lower 95% CI** | **Upper 95% CI** |
| --- | --- | --- | --- | --- | --- | --- | --- | --- | --- | --- |
| Naveed, 2023 (Vaccine NR, 2 doses) | Before infection | Endocrine | Diabetes | WT, Alpha, Delta | Timepoint | >90 days post-acute diagnosis | Adjusted for age, sex, and material deprivation index | 6 | -9 | 20 |
| Naveed, 2023 (Vaccine NR, 2 doses) | Before infection | Endocrine | Diabetes | WT, Alpha, Delta | Timepoint | >30 days post-acute diagnosis | Adjusted for age, sex (overall), acute myocardial infarction, asthma, chronic kidney disease, depression, hypertension, glucocorticoid use, vaccination status, and material deprivation index score | 9 | -15 | 29 |
| Al-Aly, 2022 (BNT162b2, mRNA-1273 or Ad26.COV2.S, primary series) | Before infection | Endocrine | Diabetes | WT, Alpha, Delta | NA | NA | Adjusted for age, race, area deprivation index, the number in the year prior to testing positive of outpatient visits, inpatient visits, unique medication prescriptions, and routine blood chemistry panels, body mass index, prior history of receiving an influenza vaccination, smoking status, comorbidities, blood pressure, estimated glomerular filtration rate, and hospital system average hospital bed occupancy, total hospital bed capacity, positivity rate of tests, number of tests, and US HHS region SARS-CoV-2 variant proportion in the week prior to enrolment, geographic region of VISN, and calendar week of enrolment | 7 | -12 | 23 |
| Mizrahi, 2023 (Vaccine NR, ≥1 dose) | Before infection | Endocrine | Diabetes | Alpha, Delta | NA | NA | Adjusted using inverse propensity score weighting Cox regression. | 29 | -134 | 78 |
| Naveed, 2023 (Vaccine NR, 1 dose) | Before infection | Endocrine | Diabetes | WT, Alpha, Delta | Timepoint | >90 days post-acute diagnosis | Adjusted for age, sex, and material deprivation index | 0 | -20 | 17 |
| Naveed, 2023 (Vaccine NR, 1 dose) | Before infection | Endocrine | Diabetes | WT, Alpha, Delta | Timepoint | >30 days post-acute diagnosis | Adjusted for age, sex (overall), acute myocardial infarction, asthma, chronic kidney disease, depression, hypertension, glucocorticoid use, vaccination status, and material deprivation index score | 2 | -23 | 21 |
| Zisis, 2022 (Vaccine NR, dose NR) | Before infection | Endocrine | Diabetes | WT, Alpha, Delta | Timepoint | 90 days post-acute diagnosis | Adjusted using propensity score matching (1:1) to balance the 2 cohorts on age, sex, race, and comorbidities | 72 | 62 | 80 |
| Zisis, 2022 (Vaccine NR, dose NR) | Before infection | Endocrine | Diabetes | WT, Alpha, Delta | Timepoint | 28 days post-acute diagnosis | Adjusted using propensity score matching (1:1) to balance the 2 cohorts on age, sex, race, and comorbidities | 57 | 46 | 65 |
| Whittaker, 2021 (BNT162b2, mRNA-1273 or ChAdOx1-S, ≥1 dose) | Mixed (before and after PCC diagnosis [post-acute COVID-19 diagnosis]) | Endocrine | Diabetes | WT, Alpha | NA | NA | Adjusted for age, sex, smoking status, BMI, and CCI with the exception of nausea, VTE and liver disease which were adjusted for age, sex, smoking status and CCI due to model convergence | -15 | -37 | 3 |
| Al-Aly, 2022 (BNT162b2, mRNA-1273 or Ad26.COV2.S, primary series) | Before infection | Neurological | Smell loss | WT, Alpha, Delta | NA | NA | Adjusted for age, race, area deprivation index, the number in the year prior to testing positive of outpatient visits, inpatient visits, unique medication prescriptions, and routine blood chemistry panels, body mass index, prior history of receiving an influenza vaccination, smoking status, comorbidities, blood pressure, estimated glomerular filtration rate, and hospital system average hospital bed occupancy, total hospital bed capacity, positivity rate of tests, number of tests, and US HHS region SARS-CoV-2 variant proportion in the week prior to enrolment, geographic region of VISN, and calendar week of enrolment | 37 | -14 | 65 |
| Al-Aly, 2022 (BNT162b2, mRNA-1273 or Ad26.COV2.S, primary series) | Before infection | Neurological | Seizure | WT, Alpha, Delta | NA | NA | Adjusted for age, race, area deprivation index, the number in the year prior to testing positive of outpatient visits, inpatient visits, unique medication prescriptions, and routine blood chemistry panels, body mass index, prior history of receiving an influenza vaccination, smoking status, comorbidities, blood pressure, estimated glomerular filtration rate, and hospital system average hospital bed occupancy, total hospital bed capacity, positivity rate of tests, number of tests, and US HHS region SARS-CoV-2 variant proportion in the week prior to enrolment, geographic region of VISN, and calendar week of enrolment | 12 | -30 | 94 |
| Al-Aly, 2022 (BNT162b2, mRNA-1273 or Ad26.COV2.S, primary series) | Before infection | Neurological | Neurocognitive decline | WT, Alpha, Delta | NA | NA | Adjusted for age, race, area deprivation index, the number in the year prior to testing positive of outpatient visits, inpatient visits, unique medication prescriptions, and routine blood chemistry panels, body mass index, prior history of receiving an influenza vaccination, smoking status, comorbidities, blood pressure, estimated glomerular filtration rate, and hospital system average hospital bed occupancy, total hospital bed capacity, positivity rate of tests, number of tests, and US HHS region SARS-CoV-2 variant proportion in the week prior to enrolment, geographic region of VISN, and calendar week of enrolment | 18 | -1 | 33 |
| Al-Aly, 2022 (BNT162b2, mRNA-1273 or Ad26.COV2.S, primary series) | Before infection | Neurological | Acute haemorrhagic cerebrovascular disease | WT, Alpha, Delta | NA | NA | Adjusted for age, race, area deprivation index, the number in the year prior to testing positive of outpatient visits, inpatient visits, unique medication prescriptions, and routine blood chemistry panels, body mass index, prior history of receiving an influenza vaccination, smoking status, comorbidities, blood pressure, estimated glomerular filtration rate, and hospital system average hospital bed occupancy, total hospital bed capacity, positivity rate of tests, number of tests, and US HHS region SARS-CoV-2 variant proportion in the week prior to enrolment, geographic region of VISN, and calendar week of enrolment | -2 | -89 | 45 |
| Brunvoll, 2022 (BNT162b2 or mRNA-1276, 2 doses) | Before infection | Neurological | Concentration problems | WT | NA | NA | Adjusted for gender, age, body mass index, chronic disease, and days since COVID-19 | 19 | -39 | 52 |
| Brunvoll, 2022 (BNT162b2 or mRNA-1276, 2 doses) | Before infection | Neurological | Memory problems | WT | NA | NA | Adjusted for gender, age, body mass index, chronic disease, and days since COVID-19 | 48 | 7 | 71 |
| Mohr, 2023 (BNT162b2 or mRNA-1276, 2 doses) | Before infection | Neurological | NA | WT, Alpha, Delta | Timepoint | >16 weeks post-vaccination | Adjusted for age, race, ethnicity, comorbidities, calendar month of diagnosis, and weeks since symptoms started. | 31 | 5 | 49 |
| Mohr, 2023 (BNT162b2 or mRNA-1276, 2 doses) | Before infection | Neurological | NA | WT, Alpha, Delta | Timepoint | ≤16 weeks post-vaccination | Adjusted for age, race, ethnicity, comorbidities, calendar month of diagnosis, and weeks since symptoms started | 28 | -1 | 49 |
| Mohr, 2023 (BNT162b2 or mRNA-1276, 2 doses) | Before infection | Neurological | NA | WT, Alpha, Delta | Timepoint | 6 weeks | Adjusted for age, race, ethnicity, comorbidities, calendar month of diagnosis, and weeks since symptoms started. | 29 | 7 | 45 |
| Mohr, 2023 (BNT162b2 or mRNA-1276, 2 doses) | Before infection | Neurological | NA | WT, Alpha, Delta | NA | Symptoms self-reported as moderate or severe (as opposed to mild) | Adjusted for age, race and ethnicity, comorbidities, calendar month of diagnosis, and weeks since symptoms started | 57 | 27 | 74 |
| Ertesvag, 2023 (BNT162b2 or mRNA-1276, ≥1 dose) | Before infection | Neurological | Including numbness, dizziness and sleeping problems | Delta | Population | Adolescents (≥16 years) | Adjusted for female sex, age, reinfection, and headache at baseline | -53 | -515 | 53 |
| Ertesvag, 2023 (BNT162b2 or mRNA-1276, ≥1 dose) | Before infection | Neurological | Cognitive symptoms including memory and concentration impairment | Delta | Population | Adolescents (≥16 years) | Adjusted for female sex, age, reinfection, and headache at baseline | -167 | -798 | 10 |
| Mizrahi, 2023 (Vaccine NR, ≥1 dose) | Before infection | Neurological | Dizziness | Alpha, Delta | NA | NA | Adjusted using inverse propensity score weighting Cox regression. | 10 | -28 | 36 |
| Mizrahi, 2023 (Vaccine NR, ≥1 dose) | Before infection | Neurological | Tremor | Alpha, Delta | NA | NA | Adjusted using inverse propensity score weighting Cox regression. | 52 | -78 | 87 |
| Mizrahi, 2023 (Vaccine NR, ≥1 dose) | Before infection | Neurological | Headache | Alpha, Delta | NA | NA | Adjusted using inverse propensity score weighting Cox regression. | 7 | -22 | 29 |
| Mizrahi, 2023 (Vaccine NR, ≥1 dose) | Before infection | Neurological | Concentration and memory impairment | Alpha, Delta | NA | NA | Adjusted using inverse propensity score weighting Cox regression. | 51 | 5 | 74 |
| Zisis, 2022 (Vaccine NR, dose NR) | Before infection | Neurological | Headache | WT, Alpha, Delta | Timepoint | 90 days post-acute diagnosis | Adjusted using propensity score matching (1:1) to balance the 2 cohorts on age, sex, race, and comorbidities | 61 | 55 | 66 |
| Zisis, 2022 (Vaccine NR, dose NR) | Before infection | Neurological | Headache | WT, Alpha, Delta | Timepoint | 28 days post-acute diagnosis | Adjusted using propensity score matching (1:1) to balance the 2 cohorts on age, sex, race, and comorbidities | 44 | 37 | 50 |
| Kuodi, 2022 (BNT162b2, 2 doses) | Mixed | Neurological | Sleeping problems | WT, Alpha, Delta | NA | NA | Adjusted for time to from beginning of symptoms to survey response, age, and existing symptoms | 50 | -60 | 80 |
| Kuodi, 2022 (BNT162b2, 2 doses) | Mixed | Neurological | Loss of concentration | WT, Alpha, Delta | NA | NA | Adjusted for time to from beginning of symptoms to survey response, age, and existing symptoms | 40 | -110 | 80 |
| Kuodi, 2022 (BNT162b2, 2 doses) | Mixed | Neurological | Headache | WT, Alpha, Delta | NA | NA | Adjusted for time to from beginning of symptoms to survey response, age, and existing symptoms | 50 | 10 | 70 |
| Kuodi, 2022 (BNT162b2, 2 doses) | Mixed | Neurological | Dizziness | WT, Alpha, Delta | NA | NA | Adjusted for time to from beginning of symptoms to survey response, age, and existing symptoms | 70 | 10 | 90 |
| Whittaker, 2021 (BNT162b2, mRNA-1273 or ChAdOx1-S, ≥1 dose) | Mixed (before and after PCC diagnosis [post-acute COVID-19 diagnosis]) | Neurological | Neuropathic pain | WT, Alpha | NA | NA | Adjusted for age, sex, smoking status, BMI, and CCI with the exception of nausea, VTE and liver disease which were adjusted for age, sex, smoking status and CCI due to model convergence | 29 | -40 | 64 |
| Whittaker, 2021 (BNT162b2, mRNA-1273 or ChAdOx1-S, ≥1 dose) | Mixed (before and after PCC diagnosis [post-acute COVID-19 diagnosis]) | Neurological | Insomnia | WT, Alpha | NA | NA | Adjusted for age, sex, smoking status, BMI, and CCI with the exception of nausea, VTE and liver disease which were adjusted for age, sex, smoking status and CCI due to model convergence | 56 | 37 | 70 |
| Whittaker, 2021 (BNT162b2, mRNA-1273 or ChAdOx1-S, ≥1 dose) | Mixed (before and after PCC diagnosis [post-acute COVID-19 diagnosis]) | Neurological | Headache | WT, Alpha | NA | NA | Adjusted for age, sex, smoking status, BMI, and CCI with the exception of nausea, VTE and liver disease which were adjusted for age, sex, smoking status and CCI due to model convergence | 36 | 23 | 46 |
| Whittaker, 2021 (BNT162b2, mRNA-1273 or ChAdOx1-S, ≥1 dose) | Mixed (before and after PCC diagnosis [post-acute COVID-19 diagnosis]) | Neurological | Dizziness | WT, Alpha | NA | NA | Adjusted for age, sex, smoking status, BMI, and CCI with the exception of nausea, VTE and liver disease which were adjusted for age, sex, smoking status and CCI due to model convergence | 51 | 38 | 61 |
| Whittaker, 2021 (BNT162b2, mRNA-1273 or ChAdOx1-S, ≥1 dose) | Mixed (before and after PCC diagnosis [post-acute COVID-19 diagnosis]) | Neurological | Cognitive impairment | WT, Alpha | NA | NA | Adjusted for age, sex, smoking status, BMI, and CCI with the exception of nausea, VTE and liver disease which were adjusted for age, sex, smoking status and CCI due to model convergence | 19 | -39 | 53 |
| Al-Aly, 2022 (BNT162b2, mRNA-1273 or Ad26.COV2.S, primary series) | Before infection | Musculoskeletal | Muscle pain | WT, Alpha, Delta | NA | NA | Adjusted for age, race, area deprivation index, the number in the year prior to testing positive of outpatient visits, inpatient visits, unique medication prescriptions, and routine blood chemistry panels, body mass index, prior history of receiving an influenza vaccination, smoking status, comorbidities, blood pressure, estimated glomerular filtration rate, and hospital system average hospital bed occupancy, total hospital bed capacity, positivity rate of tests, number of tests, and US HHS region SARS-CoV-2 variant proportion in the week prior to enrolment, geographic region of VISN, and calendar week of enrolment | 24 | 12 | 36 |
| Al-Aly, 2022 (BNT162b2, mRNA-1273 or Ad26.COV2.S, primary series) | Before infection | Musculoskeletal | Joint pain | WT, Alpha, Delta | NA | NA | Adjusted for age, race, area deprivation index, the number in the year prior to testing positive of outpatient visits, inpatient visits, unique medication prescriptions, and routine blood chemistry panels, body mass index, prior history of receiving an influenza vaccination, smoking status, comorbidities, blood pressure, estimated glomerular filtration rate, and hospital system average hospital bed occupancy, total hospital bed capacity, positivity rate of tests, number of tests, and US HHS region SARS-CoV-2 variant proportion in the week prior to enrolment, geographic region of VISN, and calendar week of enrolment | 2 | -11 | 14 |
| Kuodi, 2022 (BNT162b2, 2 doses) | Mixed | Musculoskeletal | Weakness of limbs | WT, Alpha, Delta | NA | NA | Adjusted for time to from beginning of symptoms to survey response, age, and existing symptoms | 60 | 10 | 80 |
| Kuodi, 2022 (BNT162b2, 2 doses) | Mixed | Musculoskeletal | Persistent muscle pain | WT, Alpha, Delta | NA | NA | Adjusted for time to from beginning of symptoms to survey response, age, and existing symptoms | 70 | 10 | 90 |
| Whittaker, 2021 (BNT162b2, mRNA-1273 or ChAdOx1-S, ≥1 dose) | Mixed (before and after PCC diagnosis [post-acute COVID-19 diagnosis]) | Musculoskeletal | Paraesthesia | WT, Alpha | NA | NA | Adjusted for age, sex, smoking status, BMI, and CCI with the exception of nausea, VTE and liver disease which were adjusted for age, sex, smoking status and CCI due to model convergence | 52 | 34 | 66 |
| Whittaker, 2021 (BNT162b2, mRNA-1273 or ChAdOx1-S, ≥1 dose) | Mixed (before and after PCC diagnosis [post-acute COVID-19 diagnosis]) | Musculoskeletal | Muscle pain | WT, Alpha | NA | NA | Adjusted for age, sex, smoking status, BMI, and CCI with the exception of nausea, VTE and liver disease which were adjusted for age, sex, smoking status and CCI due to model convergence | 29 | 5 | 47 |
| Whittaker, 2021 (BNT162b2, mRNA-1273 or ChAdOx1-S, ≥1 dose) | Mixed (before and after PCC diagnosis [post-acute COVID-19 diagnosis]) | Musculoskeletal | Joint pain | WT, Alpha | NA | NA | Adjusted for age, sex, smoking status, BMI, and CCI with the exception of nausea, VTE and liver disease which were adjusted for age, sex, smoking status and CCI due to model convergence | 45 | 40 | 49 |
| Whittaker, 2021 (BNT162b2, mRNA-1273 or ChAdOx1-S, ≥1 dose) | Mixed (before and after PCC diagnosis [post-acute COVID-19 diagnosis]) | Musculoskeletal | Arthritis | WT, Alpha | NA | NA | Adjusted for age, sex, smoking status, BMI, and CCI with the exception of nausea, VTE and liver disease which were adjusted for age, sex, smoking status and CCI due to model convergence | -18 | -54 | 9 |
| Al-Aly, 2022 (BNT162b2, mRNA-1273 or Ad26.COV2.S, primary series) | Before infection | Cardiovascular | Tachycardia | WT, Alpha, Delta | NA | NA | Adjusted for age, race, area deprivation index, the number in the year prior to testing positive of outpatient visits, inpatient visits, unique medication prescriptions, and routine blood chemistry panels, body mass index, prior history of receiving an influenza vaccination, smoking status, comorbidities, blood pressure, estimated glomerular filtration rate, and hospital system average hospital bed occupancy, total hospital bed capacity, positivity rate of tests, number of tests, and US HHS region SARS-CoV-2 variant proportion in the week prior to enrolment, geographic region of VISN, and calendar week of enrolment | 43 | 26 | 56 |
| Al-Aly, 2022 (BNT162b2, mRNA-1273 or Ad26.COV2.S, primary series) | Before infection | Cardiovascular | Stroke | WT, Alpha, Delta | NA | NA | Adjusted for age, race, area deprivation index, the number in the year prior to testing positive of outpatient visits, inpatient visits, unique medication prescriptions, and routine blood chemistry panels, body mass index, prior history of receiving an influenza vaccination, smoking status, comorbidities, blood pressure, estimated glomerular filtration rate, and hospital system average hospital bed occupancy, total hospital bed capacity, positivity rate of tests, number of tests, and US HHS region SARS-CoV-2 variant proportion in the week prior to enrolment, geographic region of VISN, and calendar week of enrolment | 21 | -4 | 94 |
| Al-Aly, 2022 (BNT162b2, mRNA-1273 or Ad26.COV2.S, primary series) | Before infection | Cardiovascular | Pericarditis | WT, Alpha, Delta | NA | NA | Adjusted for age, race, area deprivation index, the number in the year prior to testing positive of outpatient visits, inpatient visits, unique medication prescriptions, and routine blood chemistry panels, body mass index, prior history of receiving an influenza vaccination, smoking status, comorbidities, blood pressure, estimated glomerular filtration rate, and hospital system average hospital bed occupancy, total hospital bed capacity, positivity rate of tests, number of tests, and US HHS region SARS-CoV-2 variant proportion in the week prior to enrolment, geographic region of VISN, and calendar week of enrolment | 18 | -119 | 69 |
| Al-Aly, 2022 (BNT162b2, mRNA-1273 or Ad26.COV2.S, primary series) | Before infection | Cardiovascular | Other dysrhythmias | WT, Alpha, Delta | NA | NA | Adjusted for age, race, area deprivation index, the number in the year prior to testing positive of outpatient visits, inpatient visits, unique medication prescriptions, and routine blood chemistry panels, body mass index, prior history of receiving an influenza vaccination, smoking status, comorbidities, blood pressure, estimated glomerular filtration rate, and hospital system average hospital bed occupancy, total hospital bed capacity, positivity rate of tests, number of tests, and US HHS region SARS-CoV-2 variant proportion in the week prior to enrolment, geographic region of VISN, and calendar week of enrolment | 48 | 25 | 64 |
| Al-Aly, 2022 (BNT162b2, mRNA-1273 or Ad26.COV2.S, primary series) | Before infection | Cardiovascular | Myocarditis | WT, Alpha, Delta | NA | NA | Adjusted for age, race, area deprivation index, the number in the year prior to testing positive of outpatient visits, inpatient visits, unique medication prescriptions, and routine blood chemistry panels, body mass index, prior history of receiving an influenza vaccination, smoking status, comorbidities, blood pressure, estimated glomerular filtration rate, and hospital system average hospital bed occupancy, total hospital bed capacity, positivity rate of tests, number of tests, and US HHS region SARS-CoV-2 variant proportion in the week prior to enrolment, geographic region of VISN, and calendar week of enrolment | 95 | 62 | 99 |
| Al-Aly, 2022 (BNT162b2, mRNA-1273 or Ad26.COV2.S, primary series) | Before infection | Cardiovascular | Myocardial infarction | WT, Alpha, Delta | NA | NA | Adjusted for age, race, area deprivation index, the number in the year prior to testing positive of outpatient visits, inpatient visits, unique medication prescriptions, and routine blood chemistry panels, body mass index, prior history of receiving an influenza vaccination, smoking status, comorbidities, blood pressure, estimated glomerular filtration rate, and hospital system average hospital bed occupancy, total hospital bed capacity, positivity rate of tests, number of tests, and US HHS region SARS-CoV-2 variant proportion in the week prior to enrolment, geographic region of VISN, and calendar week of enrolment | 30 | -15 | 58 |
| Al-Aly, 2022 (BNT162b2, mRNA-1273 or Ad26.COV2.S, primary series) | Before infection | Cardiovascular | Hypertension | WT, Alpha, Delta | NA | NA | Adjusted for age, race, area deprivation index, the number in the year prior to testing positive of outpatient visits, inpatient visits, unique medication prescriptions, and routine blood chemistry panels, body mass index, prior history of receiving an influenza vaccination, smoking status, comorbidities, blood pressure, estimated glomerular filtration rate, and hospital system average hospital bed occupancy, total hospital bed capacity, positivity rate of tests, number of tests, and US HHS region SARS-CoV-2 variant proportion in the week prior to enrolment, geographic region of VISN, and calendar week of enrolment | 8 | -8 | 21 |
| Al-Aly, 2022 (BNT162b2, mRNA-1273 or Ad26.COV2.S, primary series) | Before infection | Cardiovascular | Heart failure | WT, Alpha, Delta | NA | NA | Adjusted for age, race, area deprivation index, the number in the year prior to testing positive of outpatient visits, inpatient visits, unique medication prescriptions, and routine blood chemistry panels, body mass index, prior history of receiving an influenza vaccination, smoking status, comorbidities, blood pressure, estimated glomerular filtration rate, and hospital system average hospital bed occupancy, total hospital bed capacity, positivity rate of tests, number of tests, and US HHS region SARS-CoV-2 variant proportion in the week prior to enrolment, geographic region of VISN, and calendar week of enrolment | 2 | -19 | 20 |
| Al-Aly, 2022 (BNT162b2, mRNA-1273 or Ad26.COV2.S, primary series) | Before infection | Cardiovascular | Atrial fibrillation | WT, Alpha, Delta | NA | NA | Adjusted for age, race, area deprivation index, the number in the year prior to testing positive of outpatient visits, inpatient visits, unique medication prescriptions, and routine blood chemistry panels, body mass index, prior history of receiving an influenza vaccination, smoking status, comorbidities, blood pressure, estimated glomerular filtration rate, and hospital system average hospital bed occupancy, total hospital bed capacity, positivity rate of tests, number of tests, and US HHS region SARS-CoV-2 variant proportion in the week prior to enrolment, geographic region of VISN, and calendar week of enrolment | 21 | 6 | 33 |
| Al-Aly, 2022 (BNT162b2, mRNA-1273 or Ad26.COV2.S, primary series) | Before infection | Cardiovascular | Acute coronary disease | WT, Alpha, Delta | NA | NA | Adjusted for age, race, area deprivation index, the number in the year prior to testing positive of outpatient visits, inpatient visits, unique medication prescriptions, and routine blood chemistry panels, body mass index, prior history of receiving an influenza vaccination, smoking status, comorbidities, blood pressure, estimated glomerular filtration rate, and hospital system average hospital bed occupancy, total hospital bed capacity, positivity rate of tests, number of tests, and US HHS region SARS-CoV-2 variant proportion in the week prior to enrolment, geographic region of VISN, and calendar week of enrolment | 8 | -13 | 25 |
| Mizrahi, 2023 (Vaccine NR, ≥1 dose) | Before infection | Cardiovascular | Hypertension | Alpha, Delta | NA | NA | Adjusted using inverse propensity score weighting Cox regression. | 37 | -122 | 82 |
| Mizrahi, 2023 (Vaccine NR, ≥1 dose) | Before infection | Cardiovascular | Cardiac arrhythmia | Alpha, Delta | NA | NA | Adjusted using inverse propensity score weighting Cox regression. | -31 | -337 | 61 |
| Mizrahi, 2023 (Vaccine NR, ≥1 dose) | Before infection | Cardiovascular | Ischemic heart disease | Alpha, Delta | NA | NA | Adjusted using inverse propensity score weighting Cox regression. | 9 | -201 | 73 |
| Mizrahi, 2023 (Vaccine NR, ≥1 dose) | Before infection | Cardiovascular | Venous thromboembolism | Alpha, Delta | NA | NA | Adjusted using inverse propensity score weighting Cox regression. | 17 | -91 | 63 |
| Mizrahi, 2023 (Vaccine NR, ≥1 dose) | Before infection | Cardiovascular | Palpitations | Alpha, Delta | NA | NA | Adjusted using inverse propensity score weighting Cox regression. | -35 | -104 | 10 |
| Zisis, 2022 (Vaccine NR, dose NR) | Before infection | Cardiovascular | Heart disease | WT, Alpha, Delta | Timepoint | 90 days post-acute diagnosis | Adjusted using propensity score matching (1:1) to balance the 2 cohorts on age, sex, race, and comorbidities | 65 | 56 | 71 |
| Zisis, 2022 (Vaccine NR, dose NR) | Before infection | Cardiovascular | Heart disease | WT, Alpha, Delta | Timepoint | 28 days post-acute diagnosis | Adjusted using propensity score matching (1:1) to balance the 2 cohorts on age, sex, race, and comorbidities | 51 | 43 | 57 |
| Zisis, 2022 (Vaccine NR, dose NR) | Before infection | Cardiovascular | Hypertension | WT, Alpha, Delta | Timepoint | 90 days post-acute diagnosis | Adjusted using propensity score matching (1:1) to balance the 2 cohorts on age, sex, race, and comorbidities | 67 | 58 | 74 |
| Zisis, 2022 (Vaccine NR, dose NR) | Before infection | Cardiovascular | Hypertension | WT, Alpha, Delta | Timepoint | 28 days post-acute diagnosis | Adjusted using propensity score matching (1:1) to balance the 2 cohorts on age, sex, race, and comorbidities | 55 | 46 | 62 |
| Whittaker, 2021 (BNT162b2, mRNA-1273 or ChAdOx1-S, ≥1 dose) | Mixed (before and after PCC diagnosis [post-acute COVID-19 diagnosis]) | Cardiovascular | Heart failure | WT, Alpha | NA | NA | Adjusted for age, sex, smoking status, BMI, and CCI with the exception of nausea, VTE and liver disease which were adjusted for age, sex, smoking status and CCI due to model convergence | 12 | -75 | 56 |
| Whittaker, 2021 (BNT162b2, mRNA-1273 or ChAdOx1-S, ≥1 dose) | Mixed (before and after PCC diagnosis [post-acute COVID-19 diagnosis]) | Cardiovascular | Hypertension | WT, Alpha | NA | NA | Adjusted for age, sex, smoking status, BMI, and CCI with the exception of nausea, VTE and liver disease which were adjusted for age, sex, smoking status and CCI due to model convergence | -15 | -39 | 5 |
| Whittaker, 2021 (BNT162b2, mRNA-1273 or ChAdOx1-S, ≥1 dose) | Mixed (before and after PCC diagnosis [post-acute COVID-19 diagnosis]) | Cardiovascular | Ischemic heart disease | WT, Alpha | NA | NA | Adjusted for age, sex, smoking status, BMI, and CCI with the exception of nausea, VTE and liver disease which were adjusted for age, sex, smoking status and CCI due to model convergence | 59 | 37 | 73 |
| Whittaker, 2021 (BNT162b2, mRNA-1273 or ChAdOx1-S, ≥1 dose) | Mixed (before and after PCC diagnosis [post-acute COVID-19 diagnosis]) | Cardiovascular | Palpitations | WT, Alpha | NA | NA | Adjusted for age, sex, smoking status, BMI, and CCI with the exception of nausea, VTE and liver disease which were adjusted for age, sex, smoking status and CCI due to model convergence | 37 | 17 | 52 |
| Whittaker, 2021 (BNT162b2, mRNA-1273 or ChAdOx1-S, ≥1 dose) | Mixed (before and after PCC diagnosis [post-acute COVID-19 diagnosis]) | Cardiovascular/ neurological | Peripheral arterial disease* | WT, Alpha | NA | NA | Adjusted for age, sex, smoking status, BMI, and CCI with the exception of nausea, VTE and liver disease which were adjusted for age, sex, smoking status and CCI due to model convergence | -101 | -724 | 51 |
| Whittaker, 2021 (BNT162b2, mRNA-1273 or ChAdOx1-S, ≥1 dose) | Mixed (before and after PCC diagnosis [post-acute COVID-19 diagnosis]) | Cardiovascular/ neurological | Stroke | WT, Alpha | NA | NA | Adjusted for age, sex, smoking status, BMI, and CCI with the exception of nausea, VTE and liver disease which were adjusted for age, sex, smoking status and CCI due to model convergence | 14 | -65 | 55 |
| Whittaker, 2021 (BNT162b2, mRNA-1273 or ChAdOx1-S, ≥1 dose) | Mixed (before and after PCC diagnosis [post-acute COVID-19 diagnosis]) | Cardiovascular/ neurological | Venous thromboembolism | WT, Alpha | NA | NA | Adjusted for age, sex, smoking status, BMI, and CCI with the exception of nausea, VTE and liver disease which were adjusted for age, sex, smoking status and CCI due to model convergence | 14 | -29 | 43 |

BMI: Body mass index; CI: Confidence interval; CCI: Charlson Comorbidity Index; HHS: Health and Human Services; mRNA: Messenger ribonucleic acid; NA: Not applicable; NR: Not reported; PCC: Post- COVID condition; US: United States; VE: Vaccine effectiveness; VISN: Veterans Integrated Services Network; VTE: Venous thromboembolism; WT: Wild-type.

**eTable 10. Summary of adjusted associations between vaccination and PCC that cannot be converted to VE (N=10 studies)**

| Author | Vaccination timepoint (dose) | Outcome definition | Measure | Adjusted covariates | Subgroup | Outcome value | Protective effect |
| --- | --- | --- | --- | --- | --- | --- | --- |
| Strain, 2022 | After PCC onset (≥1 dose) | Overall improvement fatigue post-vaccination | Mean (95% CI); p-value | Adjusted for baseline symptom score, sex, age group and duration of long COVID symptoms in those with symptoms at baseline and/or who developed symptoms as a result of vaccination | ChAdOx1 (reference) | 13.7 (9.7–17.8) | N/A |
|  |  |  |  |  | BNT162b2 (p-value vs. ChAdOx1) | 17.8 | No Difference |
|  |  |  |  |  |  | (13.2–22.4); p= 0.08 |  |
|  |  |  |  |  | mRNA-1273 (p-value vs. ChAdOx1) | 26.5 | Yes |
|  |  |  |  |  |  | (16.9–36.1); p=0.009 |  |
|  |  |  | p-value |  | p-value for mRNA-1273 vs. BNT162b2 | P= 0.08 | No Difference |
|  |  | Overall improvement brain fog post-vaccination | Mean (95% CI); p-value | Adjusted for baseline symptom score, sex, age group and duration of long COVID symptoms in those with symptoms at baseline and/or who developed symptoms as a result of vaccination | ChAdOx1 (reference) | 22.2 | N/A |
|  |  |  |  |  |  | (17.4–27.0) |  |
|  |  |  |  |  | BNT162b2 (p-value vs. ChAdOx1) | 21.7 | No Difference |
|  |  |  |  |  |  | (16.1–27.2); p=0.9 |  |
|  |  |  |  |  | mRNA-1273 (p-value vs. ChAdOx1) | 31.7 | Yes |
|  |  |  |  |  |  | (20.6–42.9); p=0.01 |  |
|  |  |  | p-value |  | p-value for mRNA-1273 vs. BNT162b2 | P=0.02 | Yes |
|  |  | Overall improvement myalgia post-vaccination | Mean (95% CI); p-value | Adjusted for baseline symptom score, sex, age group and duration of long COVID symptoms in those with symptoms at baseline and/or who developed symptoms as a result of vaccination | ChAdOx1 (reference) | 11.5 | N/a |
|  |  |  |  |  |  | (6.1–16.8) |  |
|  |  |  |  |  | BNT162b2 (p-value vs. ChAdOx1) | 16.8 | No Difference |
|  |  |  |  |  |  | (10.4–23.1); p=0.3 |  |
|  |  |  |  |  | mRNA-1273 (p-value vs. ChAdOx1) | 30.7 | Yes |
|  |  |  |  |  |  | (18.3–43.0); p=0.006 |  |
|  |  |  | p-value |  | p-value for mRNA-1273 vs. BNT162b2 | p=0.04 | Yes |
|  |  | Overall improvement shortness of breath post-vaccination | Mean (95% CI); p-value | Adjusted for baseline symptom score, sex, age group and duration of long COVID symptoms in those with symptoms at baseline and/or who developed symptoms as a result of vaccination | ChAdOx1 (reference) | 23.2 | N/A |
|  |  |  |  |  |  | (18.1–28.3) |  |
|  |  |  |  |  | BNT162b2 (p-value vs. ChAdOx1) | 24.5 | No Difference |
|  |  |  |  |  |  | (18.6–30.3); p=0.5 |  |
|  |  |  |  |  | mRNA-1273 (p-value vs. ChAdOx1) | 33.7 | No Difference |
|  |  |  |  |  |  | (21.6–45.8); p=0.07 |  |
|  |  |  | p-value |  | p-value for mRNA-1273 vs. BNT162b2 | P=0.2 | No Difference |
|  |  | Overall improvement insomnia post-vaccination | Mean (95% CI); p-value | Adjusted for baseline symptom score, sex, age group and duration of long COVID symptoms in those with symptoms at baseline and/or who developed symptoms as a result of vaccination | ChAdOx1 (reference) | 23.6 | N/A |
|  |  |  |  |  |  | (17.2–30.1) |  |
|  |  |  |  |  | BNT162b2 (p-value vs. ChAdOx1) | 28.5 | No Difference |
|  |  |  |  |  |  | (21.3–35.7); p=0.8 |  |
|  |  |  |  |  | mRNA-1273 (p-value vs. ChAdOx1) | 30.2 | No Difference |
|  |  |  |  |  |  | (16.2–44.2); p=0.1 |  |
|  |  |  | p-value |  | p-value for mRNA-1273 vs. BNT162b2 | P=0.2 | No Difference |
|  |  | Overall improvement chest pain/palpitations post-vaccination | Mean (95% CI); p-value | Adjusted for baseline symptom score, sex, age group and duration of long COVID symptoms in those with symptoms at baseline and/or who developed symptoms as a result of vaccination | ChAdOx1 (reference) | 25.6 | N/A |
|  |  |  |  |  |  | (20.0–31.1) |  |
|  |  |  |  |  | BNT162b2 (p-value vs. ChAdOx1) | 26.1 | No Difference |
|  |  |  |  |  |  | (19.8–32.3); p=0.98 |  |
|  |  |  |  |  | mRNA-1273 (p-value vs. ChAdOx1) | 34.8 | No Difference |
|  |  |  |  |  |  | (22.1–47.5); p=0.2 |  |
|  |  |  | p-value |  | p-value for mRNA-1273 vs. BNT162b2 | P=0.2 | No Difference |
|  |  | Overall improvement gastro-intestinal symptoms post-vaccination | Mean (95% CI); p-value | Adjusted for baseline symptom score, sex, age group and duration of long COVID symptoms in those with symptoms at baseline and/or who developed symptoms as a result of vaccination | ChAdOx1 (reference) | 24.7 | N/A |
|  |  |  |  |  |  | (18.5–31.0) |  |
|  |  |  |  |  | BNT162b2 (p-value vs. ChAdOx1) | 24.6 | No Difference |
|  |  |  |  |  |  | (17.4–31.8); p=0.98 |  |
|  |  |  |  |  | mRNA-1273 (p-value vs. ChAdOx1) | 41.7 | Yes |
|  |  |  |  |  |  | (27.8–55.6); p=0.002 |  |
|  |  |  | p-value |  | p-value for mRNA-1273 vs. BNT162b2 | p<0.001 | Yes |
|  |  | Overall improvement anosmia post-vaccination | Mean (95% CI); p-value | Adjusted for baseline symptom score, sex, age group and duration of long COVID symptoms in those with symptoms at baseline and/or who developed symptoms as a result of vaccination | ChAdOx1 (reference) | 33.5 | N/A |
|  |  |  |  |  |  | (26.7–40.2) |  |
|  |  |  |  |  | BNT162b2 (p-value vs. ChAdOx1) | 31.5 | No Difference |
|  |  |  |  |  |  | (24.1–38.9); p=0.7 |  |
|  |  |  |  |  | mRNA-1273 (p-value vs. ChAdOx1) | 46.3 | No Difference |
|  |  |  |  |  |  | (26.1–46.3); p=0.4 |  |
|  |  |  | p-value |  | p-value for mRNA-1273 vs. BNT162b2 | P=0.2 | No Difference |
|  |  | Overall improvement autonomic dysfunction post-vaccination | Mean (95% CI); p-value | Adjusted for baseline symptom score, sex, age group and duration of long COVID symptoms in those with symptoms at baseline and/or who developed symptoms as a result of vaccination | ChAdOx1 (reference) | 23.9 | N/A |
|  |  |  |  |  |  | (16.2–31.7) |  |
|  |  |  |  |  | BNT162b2 (p-value vs. ChAdOx1) | 28.9 | Yes |
|  |  |  |  |  |  | (13.6–44.3); p=0.01 |  |
|  |  |  |  |  | mRNA-1273 (p-value vs. ChAdOx1) | 33.7 | Yes |
|  |  |  |  |  |  | (25.4–41.9); p=0.004 |  |
|  |  |  | p-value |  | p-value for mRNA-1273 vs. BNT162b2 | P=0.1 | No Difference |
|  |  | Overall improvement Postural Orthostatic Tachycardia symptoms post-vaccination | Mean (95% CI); p-value | Adjusted for baseline symptom score, sex, age group and duration of long COVID symptoms in those with symptoms at baseline and/or who developed symptoms as a result of vaccination | ChAdOx1 (reference) | 23 | N/A |
|  |  |  |  |  |  | (15.3–30.7) |  |
|  |  |  |  |  | BNT162b2 (p-value vs. ChAdOx1) | 25.4 | No Difference |
|  |  |  |  |  |  | (17.0–33.7); p=0.7 |  |
|  |  |  |  |  | mRNA-1273 (p-value vs. ChAdOx1) | 27.7 | No Difference |
|  |  |  |  |  |  | (11.7–43.7); p=0.1 |  |
|  |  |  | p-value |  | p-value for mRNA-1273 vs. BNT162b2 | P=0.2 | No Difference |
|  |  | Overall improvement persistent cough post-vaccination | Mean (95% CI); p-value | Adjusted for baseline symptom score, sex, age group and duration of long COVID symptoms in those with symptoms at baseline and/or who developed symptoms as a result of vaccination | ChAdOx1 (reference) | 24.1 | N/A |
|  |  |  |  |  |  | (16.4–31.8) |  |
|  |  |  |  |  | BNT162b2 (p-value vs. ChAdOx1) | 26.1 | No Difference |
|  |  |  |  |  |  | (17.1–35.1); p=0.5 |  |
|  |  |  |  |  | mRNA-1273 (p-value vs. ChAdOx1) | 32.4 | No Difference |
|  |  |  |  |  |  | (15.5–49.3); p=0.3 |  |
|  |  |  | p-value |  | p-value for mRNA-1273 vs. BNT162b2 | P=0.2 | No Difference |
|  |  | Overall improvement fever post-vaccination | Mean (95% CI); p-value | Adjusted for baseline symptom score, sex, age group and duration of long COVID symptoms in those with symptoms at baseline and/or who developed symptoms as a result of vaccination | ChAdOx1 (reference) | 1.7 | N/A |
|  |  |  |  |  |  | (−10.5–13.9) |  |
|  |  |  |  |  | BNT162b2 (p-value vs. ChAdOx1) | 22.4 | No Difference |
|  |  |  |  |  |  | (8.6–36.3); p=0.4 |  |
|  |  |  |  |  | mRNA-1273 (p-value vs. ChAdOx1) | 36.2 | No Difference |
|  |  |  |  |  |  | (10.1–62.2); p=0.1 |  |
|  |  |  | p-value |  | p-value for mRNA-1273 vs. BNT162b2 | P=0.1 | No Difference |
|  |  | Overall improvement rash (including COVID toes) post-vaccination | Mean (95% CI); p-value | Adjusted for baseline symptom score, sex, age group and duration of long COVID symptoms in those with symptoms at baseline and/or who developed symptoms as a result of vaccination | ChAdOx1 (reference) | 29.7 | N/A |
|  |  |  |  |  |  | (12.7–37.6) |  |
|  |  |  |  |  | BNT162b2 (p-value vs. ChAdOx1) | 32.4 | No Difference |
|  |  |  |  |  |  | (23.6–41.1); p=0.7 |  |
|  |  |  |  |  | mRNA-1273 (p-value vs. ChAdOx1) | 33.9 | No Difference |
|  |  |  |  |  |  | (17.0–50.9); p=0.6 |  |
|  |  |  | p-value |  | p-value for mRNA-1273 vs. BNT162b2 | P=0.6 | No Difference |
|  |  | Overall improvement vascular complications post-vaccination | Mean (95% CI); p-value | Adjusted for baseline symptom score, sex, age group and duration of long COVID symptoms in those with symptoms at baseline and/or who developed symptoms as a result of vaccination | ChAdOx1 (reference) | 25.3 | N/A |
|  |  |  |  |  |  | (15.7–34) |  |
|  |  |  |  |  | BNT162b2 (p-value vs. ChAdOx1) | 29.8 | No Difference |
|  |  |  |  |  |  | (19.1–40.4); P=0.7 |  |
|  |  |  |  |  | mRNA-1273 (p-value vs. ChAdOx1) | 31.4 | No Difference |
|  |  |  |  |  |  | (10.8–52.0); P=0.8 |  |
|  |  |  | p-value |  | p-value for mRNA-1273 vs. BNT162b2 | P=0.6 | No Difference |
| Hirahata, 2022 | NR | Performance status score in Long-COVID | Regression coefficient | Adjusted for age, sex, employment status, infection waves and vaccination | Vaccinated vs unvaccinated | 0.14 (-0.41–0.69) | Yes, better performance status score in long-COVID when vaccinated |
|  |  |  |  | Adjusted for age, sex, employment status, infection waves and vaccination, fatigue, depressive symptom, brain fog, headache, dyspnoea, insomnia, palpitation, body pain, loss of smell, loss of appetite, loss of taste, hair loss, fever, and cough. | Vaccinated vs unvaccinated | -0.15 (-0.64–0.34) |  |
| Taquet, 2022 | Before infection (primary series) | Composite risk of death and abdominal symptoms | Matched HR | Matched with the same number of unvaccinated cases (who had received an influenza vaccine), accounting for age, sex, ethnicity, obesity, socioeconomic deprivation and specific health conditions, and date of infection. | Any vaccination (1 or 2 doses) | 1.02 (0.93-1.12) | No difference |
|  |  | Composite risk of death and abnormal breathing |  |  |  | 0.96 (0.89-1.04) | No difference |
|  |  | Composite risk of death and anosmia |  |  |  | 0.82 (0.69-0.98) | Yes |
|  |  | Composite risk of death and anxiety disorder |  |  |  | 1.06 (0.97-1.15) | No difference |
|  |  | Composite risk of death and anxiety/depression |  |  |  | 1.07 (1.00-1.15) | No |
|  |  | Composite risk of death and arrhythmia |  |  |  | 0.93 (0.86-1.01) | No difference |
|  |  | Composite risk of death and cardiac failure |  |  |  | 0.95 (0.86-1.05) | No difference |
|  |  | Composite risk of death and cardiomyopathy |  |  |  | 0.92 (0.80-1.05) | No difference |
|  |  | Composite risk of death and cerebral haemorrhage |  |  |  | 0.82 (0.68-0.97) | Yes |
|  |  | Composite risk of death and chest/throat pain |  |  |  | 0.98 (0.88-1.08) | No difference |
|  |  | Composite risk of death and cognitive symptoms |  |  |  | 0.93 (0.83-1.04) | No difference |
|  |  | Composite risk of death and coronary disease |  |  |  | 0.87 (0.75-1.00) | No difference |
|  |  | Risk of death |  |  |  | 0.75 (0.62-0.91) | Yes |
|  |  | Composite risk of death and fatigue |  |  |  | 0.89 (0.81-0.97) | Yes |
|  |  | Composite risk of death and GORD |  |  |  | 1.02 (0.94-1.11) | No difference |
|  |  | Composite risk of death and hair loss |  |  |  | 0.75 (0.64-0.88) | Yes |
|  |  | Composite risk of death and headache |  |  |  | 1.07 (0.96-1.18) | No difference |
|  |  | Composite risk of death and hospitalization |  |  |  | 0.90 (0.84–0.97) | Yes |
|  |  | Composite risk of death and hypercoagulopathy/DVT/PE |  |  |  | 0.81 (0.72-0.91) | Yes |
|  |  | Composite risk of death and hyperlipidaemia |  |  |  | 1.02 (0.96-1.08) | No difference |
|  |  | Composite risk of death and hypertension |  |  |  | 0.98 (0.93-1.04) | No difference |
|  |  | Composite risk of death and hypoxaemia |  |  |  | 0.72 (0.65-0.80) | Yes |
|  |  | Composite risk of death and ICU admission |  |  |  | 0.75 (0.65–0.85) | Yes |
|  |  | Composite risk of death and interstitial lung disease |  |  |  | 0.84 (0.73-0.98 | Yes |
|  |  | Composite risk of death and intubation/ ventilation |  |  |  | 0.72 (0.61–0.84) | Yes |
|  |  | Composite risk of death and ischaemic stroke |  |  |  | 0.84 (0.73-0.97) | Yes |
|  |  | Composite risk of death and joint pain |  |  |  | 0.98 (0.89-1.08) | No difference |
|  |  | Composite risk of death and kidney disease |  |  |  | 0.94 (0.86-1.02) | No difference |
|  |  | Composite risk of death and liver disease |  |  |  | 0.94 (0.84-1.06) | No difference |
|  |  | Composite risk of death and any long-COVID feature |  |  |  | 1.01 (0.96–1.05) | No difference |
|  |  | Composite risk of death and mood disorder |  |  |  | 1.05 (0.96-1.15) | No difference |
|  |  | Composite risk of death and myalgia |  |  |  | 0.78 (0.67-0.91) | Yes |
|  |  | Composite risk of death and myocarditis |  |  |  | 0.77 (0.64-0.92) | Yes |
|  |  | Composite risk of death and myoneural junction/muscle disease |  |  |  | 0.78 (0.66-0.92) | Yes |
|  |  | Composite risk of death and nerve/nerve root/plexus disorder |  |  |  | 0.82 (0.72-0.95) | Yes |
|  |  | Composite risk of death and obesity |  |  |  | 0.97 (0.91-1.04) | No difference |
|  |  | Composite risk of death and other pain |  |  |  | 0.90 (0.81-0.99) | Yes |
|  |  | Composite risk of death and oxygen requirement |  |  |  | 0.83 (0.75–0.92 | Yes |
|  |  | Composite risk of death and peripheral neuropathy |  |  |  | 0.93 (0.81-1.07) | No difference |
|  |  | Composite risk of death and psychotic disorder |  |  |  | 0.75 (0.63-0.89) | Yes |
|  |  | Composite risk of death and respiratory failure |  |  |  | 0.70 (0.63-0.78) | Yes |
|  |  | Composite risk of death and seizures |  |  |  | 0.73 (0.62-0.86) | Yes |
|  |  | Composite risk of death and sleep disorders |  |  |  | 0.95 (0.88-1.04) | No difference |
|  |  | Composite risk of death and type 2 diabetes mellitus |  |  |  | 0.97 (0.90-1.05) | No difference |
|  |  | Composite risk of death and urticaria |  |  |  | 0.77 (0.65-0.92) | Yes |
|  |  | Composite risk of death and abdominal symptoms |  |  | Two doses | 1.00 (0.89-1.12) | No difference |
|  |  | Composite risk of death and abnormal breathing |  |  |  | 0.89 (0.81-0.98) | Yes |
|  |  | Composite risk of death and anosmia |  |  |  | 0.68 (0.55-0.84) | Yes |
|  |  | Composite risk of death and anxiety disorder |  |  |  | 1.00 (0.90-1.10) | No difference |
|  |  | Composite risk of death and anxiety/depression |  |  |  | 1.03 (0.94-1.12) | No difference |
|  |  | Composite risk of death and arrhythmia |  |  |  | 0.90 (0.81-0.99) | Yes |
|  |  | Composite risk of death and cardiac failure |  |  |  | 0.95 (0.84-1.07) | No difference |
|  |  | Composite risk of death and cardiomyopathy |  |  |  | 0.85 (0.72-0.99) | Yes |
|  |  | Composite risk of death and cerebral haemorrhage |  |  |  | 0.69 (0.56-0.85) | Yes |
|  |  | Composite risk of death and chest/throat pain |  |  |  | 0.92 (0.82-1.04) | No difference |
|  |  | Composite risk of death and cognitive symptoms |  |  |  | 0.87 (0.76-0.99) | Yes |
|  |  | Composite risk of death and coronary disease |  |  |  | 0.85 (0.72-1.01) | No difference |
|  |  | Risk of death |  |  |  | 0.62 (0.49-0.78) | Yes |
|  |  | Composite risk of death and fatigue |  |  |  | 0.86 (0.77-0.96) | Yes |
|  |  | Composite risk of death and GORD |  |  |  | 1.02 (0.93-1.13) | No difference |
|  |  | Composite risk of death and hair loss |  |  |  | 0.66 (0.54-0.81) | Yes |
|  |  | Composite risk of death and headache |  |  |  | 1.00 (0.88-1.13) | No difference |
|  |  | Composite risk of death and hospitalization |  |  |  | 0.91 (0.83-1.00) | Yes |
|  |  | Composite risk of death and hypercoagulopathy/DVT/PE |  |  |  | 0.80 (0.70-0.92) | Yes |
|  |  | Composite risk of death and hyperlipidaemia |  |  |  | 1.05 (0.98-1.13) | No difference |
|  |  | Composite risk of death and hypertension |  |  |  | 1.05 (0.98-1.12) | No difference |
|  |  | Composite risk of death and hypoxaemia |  |  |  | 0.66 (0.58-0.74) | Yes |
|  |  | Composite risk of death and ICU admission |  |  |  | 0.72 (0.61-0.85) | Yes |
|  |  | Composite risk of death and interstitial lung disease |  |  |  | 0.74 (0.62-0.88) | Yes |
|  |  | Composite risk of death and intubation/ ventilation |  |  |  | 0.63 (0.52-0.76) | Yes |
|  |  | Composite risk of death and ischaemic stroke |  |  |  | 0.77 (0.64-0.91) | Yes |
|  |  | Composite risk of death and joint pain |  |  |  | 0.91 (0.81-1.02) | No difference |
|  |  | Composite risk of death and kidney disease |  |  |  | 0.97 (0.87-1.07) | No difference |
|  |  | Composite risk of death and liver disease |  |  |  | 0.93 (0.81-1.07) | No difference |
|  |  | Composite risk of death and any long-COVID feature |  |  |  | 1.00 (0.95-1.06) | No difference |
|  |  | Composite risk of death and mood disorder |  |  |  | 1.03 (0.92-1.14) | No difference |
|  |  | Composite risk of death and myalgia |  |  |  | 0.70 (0.59-0.84) | Yes |
|  |  | Composite risk of death and myocarditis |  |  |  | 0.64 (0.51-0.80) | Yes |
|  |  | Composite risk of death and myoneural junction/muscle disease |  |  |  | 0.67 (0.54-0.82) | Yes |
|  |  | Composite risk of death and nerve/nerve root/plexus disorder |  |  |  | 0.78 (0.66-0.92) | Yes |
|  |  | Composite risk of death and obesity |  |  |  | 0.97 (0.89-1.04) | No difference |
|  |  | Composite risk of death and other pain |  |  |  | 0.85 (0.76-0.96) | Yes |
|  |  | Composite risk of death and oxygen requirement |  |  |  | 0.75 (0.67-0.85) | Yes |
|  |  | Composite risk of death and peripheral neuropathy |  |  |  | 0.82 (0.70-0.97) | Yes |
|  |  | Composite risk of death and psychotic disorder |  |  |  | 0.65 (0.52-0.79) | Yes |
|  |  | Composite risk of death and respiratory failure |  |  |  | 0.63 (0.56-0.71) | Yes |
|  |  | Composite risk of death and seizures |  |  |  | 0.67 (0.55-0.82) | Yes |
|  |  | Composite risk of death and sleep disorders |  |  |  | 0.96 (0.86-1.06) | No difference |
|  |  | Composite risk of death and type 2 diabetes mellitus |  |  |  | 1.03 (0.94-1.12) | No difference |
|  |  | Composite risk of death and urticaria |  |  |  | 0.65 (0.52-0.80) | Yes |
|  |  | Composite risk of death and abdominal symptoms |  |  | One dose | 0.97 (0.83-1.12) | No difference |
|  |  | Composite risk of death and abnormal breathing |  |  |  | 1.00 (0.88-1.13) | No difference |
|  |  | Composite risk of death and anosmia |  |  |  | 0.82 (0.62-1.09) | No difference |
|  |  | Composite risk of death and anxiety disorder |  |  |  | 0.95 (0.84-1.08) | No difference |
|  |  | Composite risk of death and anxiety/depression |  |  |  | 0.96 (0.85-1.07) | No difference |
|  |  | Composite risk of death and arrhythmia |  |  |  | 0.96 (0.84-1.09) | No difference |
|  |  | Composite risk of death and cardiac failure |  |  |  | 1.01 (0.86-1.20) | No difference |
|  |  | Composite risk of death and cardiomyopathy |  |  |  | 1.02 (0.81-1.27) | No difference |
|  |  | Composite risk of death and cerebral haemorrhage |  |  |  | 0.82 (0.61-1.09) | No difference |
|  |  | Composite risk of death and chest/throat pain |  |  |  | 1.03 (0.87-1.21) | No difference |
|  |  | Composite risk of death and cognitive symptoms |  |  |  | 0.81 (0.68-0.97) | Yes |
|  |  | Composite risk of death and coronary disease |  |  |  | 0.95 (0.75-1.20) | No difference |
|  |  | Risk of death |  |  |  | 0.72 (0.53-0.98) | Yes |
|  |  | Composite risk of death and fatigue |  |  |  | 0.87 (0.76-1.01) | No difference |
|  |  | Composite risk of death and GORD |  |  |  | 0.99 (0.87-1.12) | No difference |
|  |  | Composite risk of death and hair loss |  |  |  | 0.74 (0.57-0.97) | Yes |
|  |  | Composite risk of death and headache |  |  |  | 1.12 (0.95-1.32) | No difference |
|  |  | Composite risk of death and hospitalization |  |  |  | 0.90 (0.80-1.02) | No difference |
|  |  | Composite risk of death and hypercoagulopathy/DVT/PE |  |  |  | 0.88 (0.73-1.07) | No difference |
|  |  | Composite risk of death and hyperlipidaemia |  |  |  | 0.98 (0.89-1.08) | No difference |
|  |  | Composite risk of death and hypertension |  |  |  | 0.98 (0.90-1.07) | No difference |
|  |  | Composite risk of death and hypoxaemia |  |  |  | 0.78 (0.66-0.92) | Yes |
|  |  | Composite risk of death and ICU admission |  |  |  | 0.72 (0.58-0.90) | Yes |
|  |  | Composite risk of death and interstitial lung disease |  |  |  | 0.79 (0.62-1.00) | No difference |
|  |  | Composite risk of death and intubation/ ventilation |  |  |  | 0.71 (0.54-0.93) | Yes |
|  |  | Composite risk of death and ischaemic stroke |  |  |  | 0.99 (0.78-1.25) | No difference |
|  |  | Composite risk of death and joint pain |  |  |  | 0.95 (0.82-1.10) | No difference |
|  |  | Composite risk of death and kidney disease |  |  |  | 0.86 (0.74-0.99) | Yes |
|  |  | Composite risk of death and liver disease |  |  |  | 0.91 (0.75-1.10) | No difference |
|  |  | Composite risk of death and any long-COVID feature |  |  |  | 0.96 (0.89-1.03) | No difference |
|  |  | Composite risk of death and mood disorder |  |  |  | 0.93 (0.81-1.07) | No difference |
|  |  | Composite risk of death and myalgia |  |  |  | 0.75 (0.59-0.97) | Yes |
|  |  | Composite risk of death and myocarditis |  |  |  | 0.77 (0.57-1.04) | No difference |
|  |  | Composite risk of death and myoneural junction/muscle disease |  |  |  | 0.77 (0.57-1.02) | No difference |
|  |  | Composite risk of death and nerve/nerve root/plexus disorder |  |  |  | 0.81 (0.64-1.01) | No difference |
|  |  | Composite risk of death and obesity |  |  |  | 0.98 (0.89-1.08) | No difference |
|  |  | Composite risk of death and other pain |  |  |  | 0.87 (0.75-1.01) | No difference |
|  |  | Composite risk of death and oxygen requirement |  |  |  | 0.84 (0.71-0.99) | Yes |
|  |  | Composite risk of death and peripheral neuropathy |  |  |  | 1.05 (0.84-1.31) | No difference |
|  |  | Composite risk of death and psychotic disorder |  |  |  | 0.65 (0.49-0.85) | Yes |
|  |  | Composite risk of death and respiratory failure |  |  |  | 0.75 (0.64-0.89) | Yes |
|  |  | Composite risk of death and seizures |  |  |  | 0.71 (0.55-0.92) | Yes |
|  |  | Composite risk of death and sleep disorders |  |  |  | 0.96 (0.84-1.10) | No difference |
|  |  | Composite risk of death and type 2 diabetes mellitus |  |  |  | 0.95 (0.85-1.07) | No difference |
|  |  | Composite risk of death and urticaria |  |  |  | 0.81 (0.60-1.08) | No difference |
|  |  | Composite risk of death and abdominal symptoms |  |  | Any vaccination (1 or 2 doses), <60 years | 0.99 (0.85-1.15) | No difference |
|  |  | Composite risk of death and abnormal breathing |  |  |  | 0.88 (0.77-1.00) | No difference |
|  |  | Composite risk of death and anosmia |  |  |  | 0.62 (0.42-0.92) | Yes |
|  |  | Composite risk of death and anxiety disorder |  |  |  | 1.12 (1.00-1.27) | No |
|  |  | Composite risk of death and anxiety/depression |  |  |  | 1.09 (0.97-1.21) | No difference |
|  |  | Composite risk of death and arrhythmia |  |  |  | 0.91 (0.77-1.08) | No difference |
|  |  | Composite risk of death and cardiac failure |  |  |  | 0.85 (0.67-1.08) | No difference |
|  |  | Composite risk of death and cardiomyopathy |  |  |  | 1.00 (0.75-1.32) | No difference |
|  |  | Composite risk of death and cerebral haemorrhage |  |  |  | 0.47 (0.31-0.73) | Yes |
|  |  | Composite risk of death and chest/throat pain |  |  |  | 0.91 (0.77-1.08) | No difference |
|  |  | Composite risk of death and cognitive symptoms |  |  |  | 0.82 (0.64-1.06) | No difference |
|  |  | Composite risk of death and coronary disease |  |  |  | 0.60 (0.42-0.86) | Yes |
|  |  | Risk of death |  |  |  | 0.43 (0.26-0.68) | Yes |
|  |  | Composite risk of death and fatigue |  |  |  | 0.96 (0.81-1.13) | No difference |
|  |  | Composite risk of death and GORD |  |  |  | 1.08 (0.94-1.25) | No difference |
|  |  | Composite risk of death and hair loss |  |  |  | 0.59 (0.42-0.81) | Yes |
|  |  | Composite risk of death and headache |  |  |  | 1.03 (0.89-1.20) | No difference |
|  |  | Composite risk of death and hospitalization |  |  |  | 0.81 (0.70-0.94) | Yes |
|  |  | Composite risk of death and hypercoagulopathy/DVT/PE |  |  |  | 0.72 (0.57-0.90) | Yes |
|  |  | Composite risk of death and hyperlipidaemia |  |  |  | 1.01 (0.90-1.14) | No difference |
|  |  | Composite risk of death and hypertension |  |  |  | 0.97 (0.88-1.08) | No difference |
|  |  | Composite risk of death and hypoxaemia |  |  |  | 0.52 (0.42-0.64) | Yes |
|  |  | Composite risk of death and ICU admission |  |  |  | 0.50 (0.38-0.68) | Yes |
|  |  | Composite risk of death and interstitial lung disease |  |  |  | 0.55 (0.40-0.75) | Yes |
|  |  | Composite risk of death and intubation/ ventilation |  |  |  | 0.48 (0.33-0.69) | Yes |
|  |  | Composite risk of death and ischaemic stroke |  |  |  | 0.62 (0.44-0.89) | Yes |
|  |  | Composite risk of death and joint pain |  |  |  | 1.01 (0.86-1.18) | No difference |
|  |  | Composite risk of death and kidney disease |  |  |  | 0.86 (0.71-1.02) | No difference |
|  |  | Composite risk of death and liver disease |  |  |  | 0.93 (0.75-1.15) | No difference |
|  |  | Composite risk of death and any long-COVID feature |  |  |  | 0.97 (0.90-1.05) | No difference |
|  |  | Composite risk of death and mood disorder |  |  |  | 1.15 (1.00-1.33) | No |
|  |  | Composite risk of death and myalgia |  |  |  | 0.65 (0.49-0.86) | Yes |
|  |  | Composite risk of death and myocarditis |  |  |  | 0.49 (0.32-0.76) | Yes |
|  |  | Composite risk of death and myoneural junction/muscle disease |  |  |  | 0.55 (0.37-0.82) | Yes |
|  |  | Composite risk of death and nerve/nerve root/plexus disorder |  |  |  | 0.68 (0.52-0.89) | Yes |
|  |  | Composite risk of death and obesity |  |  |  | 0.95 (0.87-1.05) | No difference |
|  |  | Composite risk of death and other pain |  |  |  | 0.97 (0.82-1.14) | No difference |
|  |  | Composite risk of death and oxygen requirement |  |  |  | 0.67 (0.54-0.82) | Yes |
|  |  | Composite risk of death and peripheral neuropathy |  |  |  | 0.86 (0.63-1.18) | No difference |
|  |  | Composite risk of death and psychotic disorder |  |  |  | 0.50 (0.35-0.72) | Yes |
|  |  | Composite risk of death and respiratory failure |  |  |  | 0.48 (0.39-0.60) | Yes |
|  |  | Composite risk of death and seizures |  |  |  | 0.45 (0.32-0.64) | Yes |
|  |  | Composite risk of death and sleep disorders |  |  |  | 1.06 (0.92-1.23) | No difference |
|  |  | Composite risk of death and type 2 diabetes mellitus |  |  |  | 1.00 (0.87-1.14) | No difference |
|  |  | Composite risk of death and urticaria |  |  |  | 0.51 (0.34-0.77) | Yes |
|  |  | Composite risk of death and abdominal symptoms |  |  | Any vaccination (1 or 2 doses), ≥60 years | 1.13 (1.00-1.27) | No |
|  |  | Composite risk of death and abnormal breathing |  |  |  | 1.01 (0.91-1.11) | No difference |
|  |  | Composite risk of death and anosmia |  |  |  | 0.93 (0.76-1.13) | No difference |
|  |  | Composite risk of death and anxiety disorder |  |  |  | 1.11 (0.99-1.25) | No difference |
|  |  | Composite risk of death and anxiety/depression |  |  |  | 1.12 (1.02-1.23) | No |
|  |  | Composite risk of death and arrhythmia |  |  |  | 1.04 (0.94-1.14) | No difference |
|  |  | Composite risk of death and cardiac failure |  |  |  | 1.09 (0.97-1.22) | No difference |
|  |  | Composite risk of death and cardiomyopathy |  |  |  | 0.96 (0.82-1.12) | No difference |
|  |  | Composite risk of death and cerebral haemorrhage |  |  |  | 0.97 (0.80-1.18) | No difference |
|  |  | Composite risk of death and chest/throat pain |  |  |  | 1.04 (0.91-1.18) | No difference |
|  |  | Composite risk of death and cognitive symptoms |  |  |  | 1.00 (0.89-1.14) | No difference |
|  |  | Composite risk of death and coronary disease |  |  |  | 1.10 (0.94-1.29) | No difference |
|  |  | Risk of death |  |  |  | 0.91 (0.74-1.12) | No difference |
|  |  | Composite risk of death and fatigue |  |  |  | 0.92 (0.83-1.03) | No difference |
|  |  | Composite risk of death and GORD |  |  |  | 1.02 (0.93-1.13) | No difference |
|  |  | Composite risk of death and hair loss |  |  |  | 0.91 (0.75-1.10) | No difference |
|  |  | Composite risk of death and headache |  |  |  | 0.98 (0.85-1.13) | No difference |
|  |  | Composite risk of death and hospitalization |  |  |  | 1.03 (0.94-1.13) | No difference |
|  |  | Composite risk of death and hypercoagulopathy/DVT/PE |  |  |  | 0.93 (0.81-1.07) | No difference |
|  |  | Composite risk of death and hyperlipidaemia |  |  |  | 1.10 (1.03-1.18) | No |
|  |  | Composite risk of death and hypertension |  |  |  | 1.10 (1.03-1.17) | No |
|  |  | Composite risk of death and hypoxaemia |  |  |  | 0.90 (0.80-1.02) | No difference |
|  |  | Composite risk of death and ICU admission |  |  |  | 0.92 (0.79-1.08) | No difference |
|  |  | Composite risk of death and interstitial lung disease |  |  |  | 0.88 (0.75-1.04) | No difference |
|  |  | Composite risk of death and intubation/ ventilation |  |  |  | 0.85 (0.71-1.03) | No difference |
|  |  | Composite risk of death and ischaemic stroke |  |  |  | 0.98 (0.83-1.15) | No difference |
|  |  | Composite risk of death and joint pain |  |  |  | 0.99 (0.88-1.12) | No difference |
|  |  | Composite risk of death and kidney disease |  |  |  | 1.05 (0.95-1.16) | No difference |
|  |  | Composite risk of death and liver disease |  |  |  | 0.95 (0.82-1.10) | No difference |
|  |  | Composite risk of death and any long-COVID feature |  |  |  | 1.02 (0.95-1.08) | No difference |
|  |  | Composite risk of death and mood disorder |  |  |  | 1.07 (0.95-1.20) | No difference |
|  |  | Composite risk of death and myalgia |  |  |  | 0.95 (0.80-1.14) | No difference |
|  |  | Composite risk of death and myocarditis |  |  |  | 0.90 (0.73-1.10) | No difference |
|  |  | Composite risk of death and myoneural junction/muscle disease |  |  |  | 0.87 (0.72-1.05) | No difference |
|  |  | Composite risk of death and nerve/nerve root/plexus disorder |  |  |  | 0.95 (0.81-1.12) | No difference |
|  |  | Composite risk of death and obesity |  |  |  | 1.03 (0.95-1.12) | No difference |
|  |  | Composite risk of death and other pain |  |  |  | 0.91 (0.81-1.02) | No difference |
|  |  | Composite risk of death and oxygen requirement |  |  |  | 1.02 (0.91-1.15) | No difference |
|  |  | Composite risk of death and peripheral neuropathy |  |  |  | 1.04 (0.89-1.21) | No difference |
|  |  | Composite risk of death and psychotic disorder |  |  |  | 0.86 (0.71-1.05) | No difference |
|  |  | Composite risk of death and respiratory failure |  |  |  | 0.85 (0.76-0.96) | Yes |
|  |  | Composite risk of death and seizures |  |  |  | 0.88 (0.73-1.06) | No difference |
|  |  | Composite risk of death and sleep disorders |  |  |  | 1.00 (0.90-1.11) | No difference |
|  |  | Composite risk of death and type 2 diabetes mellitus |  |  |  | 1.07 (0.98-1.17) | No difference |
|  |  | Composite risk of death and urticaria |  |  |  | 0.93 (0.76-1.14) | No difference |
| Tran, 2023 | After PCC onset (primary series) | Proportion of patients reporting an unacceptable symptom state | Risk Difference (95% CI) | Vaccinated patients propensity score matched 1:1 with unvaccinated controls (propensity scores accounting for sex, age, educational level (≥2 years post-secondary education higher education v lower), number of comorbidities, SARS-CoV-2 infection confirmed by laboratory analysis (yes for patients reporting a positive test result for SARS-CoV-2 infection by PCR test or serological assay, or both, and otherwise no); interval from the start of covid-19 symptoms; history of admission to hospital for covid-19 during the acute phase; score on long covid symptom tool at baseline; and score on long covid impact tool at baseline) | Vaccinated vs. unvaccinated | -7.4 (-14.5, -0.3) | Yes, vaccinated group reported fewer unacceptable symptoms |
|  |  | Long COVID Symptom Tool score at 120 days | Mean Difference in scores (95% CI) |  | Vaccinated vs. unvaccinated | -1.8 (-3.0, -0.5) | Yes, long COVID was less severe in the vaccinated group |
|  |  |  |  |  | Vaccinated with BNT162b2 vs. unvaccinated | -1.9 (-3.4, -0.5) | Yes |
|  |  |  |  |  | Vaccinated with mRNA-1273 vs. unvaccinated | 1.5 (−2.0 to 5.1) | No |
|  |  |  |  |  | Vaccinated with ChAdOx1 vs. unvaccinated | −3.9 (−7.7 to −0.1) | Yes |
|  |  |  |  |  | Vaccinated vs. unvaccinated (among patients with confirmed COVID-19) | -1.8 (-3.4, ‑0.2) | Yes |
|  |  |  |  |  | Vaccinated vs. unvaccinated (among patients whose time since symptom onset was ≤ 12 months) | -1.9 (-3.5, ‑0.3) | Yes |
|  |  |  |  |  | Vaccinated vs. unvaccinated (among patients whose time since symptom onset was > 12 months) | -1.7 (-4.0, 0.6) | No |
|  |  |  |  |  | Vaccinated vs. unvaccinated (vaccinated could not be used as controls in observation periods before they were vaccinated) | -1.6 (-3.2, -0.1) | Yes |
|  |  |  |  |  | Vaccinated vs. unvaccinated (trial 1) | -1.9 (-4.1, 0.4) | No |
|  |  |  |  |  | Vaccinated vs. unvaccinated (trial 2) | -0.9 (-2.7, -0.9) | Yes |
|  |  |  |  |  | Vaccinated vs. unvaccinated (trial 3) | -3.0 (-5.6, -0.5) | Yes |
|  |  |  |  |  | Vaccinated vs. unvaccinated (meta-analysis) | -1.7 (-2.9, -0.5) | Yes |
|  |  |  |  |  | Using inverse probability of treatment weighting using standardized mortality ration weights | -1.4 (-2.6, -0.3) | Yes |
|  |  | Long COVID Impact Tool score at 120 days | Mean Difference in scores (95% CI) |  | Vaccinated vs. unvaccinated | -3.3 (-5.7, -1.0) | Yes, vaccination reduced the effect of long COVID on patients' lives |
|  |  |  |  |  | Vaccinated vs. unvaccinated (among patients with confirmed COVID-19) | -3.8 (‑6.7, ‑0.8) | Yes |
|  |  |  |  |  | Vaccinated with BNT162b2 vs. unvaccinated | -3.6 (-6.1, -1.0) | Yes |
|  |  |  |  |  | Vaccinated with mRNA-1273 vs. unvaccinated | 2.6 (−4.8, 10.1) | No |
|  |  |  |  |  | Vaccinated with ChAdOx1 vs. unvaccinated | −7.8 (−14.7, −0.9) | Yes |
|  |  |  |  |  | Vaccinated vs. unvaccinated (among patients whose time since symptom onset was ≤ 12 months) | -2.9 (-5.7, ‑0.1) | Yes |
|  |  |  |  |  | Vaccinated vs. unvaccinated (among patients whose time since symptom onset was > 12 months) | -2.8 (-6.7, 1.1) | No |
|  |  |  |  |  | Vaccinated vs. unvaccinated (vaccinated could not be used as controls in observation periods before they were vaccinated) | -3.1 (-5.9, -0.2) | Yes |
|  |  |  |  |  | Vaccinated vs. unvaccinated (trial 1) | -4.9 (-8.9, -1.0) | Yes |
|  |  |  |  |  | Vaccinated vs. unvaccinated (trial 2) | -1.9 (-5.2, 1.3) | No |
|  |  |  |  |  | Vaccinated vs. unvaccinated (trial 3) | -3.9 (-8.1, 0.3) | No |
|  |  |  |  |  | Vaccinated vs. unvaccinated (meta-analysis) | -3.3 (-5.5, -1.2) | Yes |
|  |  | Remission of all symptoms from long COVID | HR (95% CI) |  | Vaccinated vs. unvaccinated | 1.93 (1.18, 3.14) | Yes, vaccinated more likely to experience remission of all Long COVID symptoms |
|  |  |  |  |  |  |  | Yes, vaccinated participants more likely to report remission of Long COVID symptoms |
|  |  |  | Risk Difference (95% CI) |  |  | 9.1% (5.0, 13.2) |  |
| Meza-Torres, 2022 | Mixed (primary series) | Association of all-cause mortality in long-COVID | AOR (95%CI) | Adjusted for age, sex, geographic location, Cambridge Multimorbidity Scale and whether vaccine was received before or after COVID-19 diagnosis. | Any vaccine dose, at any time | 0.1 (0.03–0.35) | Yes |
| Kuodi, 2023 | Before or after infection (0-, 1-, 2- or 3- doses) | Change in EQ-5D-5L utility index with post-COVID symptoms | % points (95% CI) | Adjusted for age, ethnicity, hypertension, hospitalization (as a proxy for severity), and duration since testing positive for SARS-CoV-2) by elapsed time testing | 2 doses BNT162b2 received at time of infection, 3–6 months since testing | -0.01 (-0.13–0.1) | Yes, smaller change in EQ-5D-5L with increased number of doses |
|  |  |  |  |  | 3 doses BNT162b2 received at time of infection, 3–6 months since testing | 0.05 (-0.05–0.14) |  |
|  |  |  |  |  | 2 or 3 doses BNT162b2 received at time of infection, 3–6 months since testing | 0.03 (-0.06–0.12) |  |
|  |  |  |  |  | 2 doses BNT162b2 received at time of infection, 7–12 months since testing | 0.17 (0.03–0.31) |  |
|  |  |  |  |  | 3 doses BNT162b2 received at time of infection, 7–12 months since testing | 0.06 (-0.17–0.30) |  |
|  |  |  |  |  | 2 or 3 doses BNT162b2 received at time of infection, 7–12 months since testing | 0.15 (0.02–0.29) |  |
|  |  |  |  |  | 2 doses BNT162b2 received at time of infection, >12 months since testing | 0.05 (-0.07–0.16) |  |
|  |  |  |  |  | 3 doses BNT162b2 received at time of infection, >12 months since testing | 0.04 (-0.15–0.23) |  |
|  |  |  |  |  | 2 or 3 doses BNT162b2 received at time of infection, >12 months since testing | 0.05 (-0.06–0.16) |  |
| Wisnivesky, 2022 | After infection and PASC | Difference in mean change between baseline and 6-month follow-up for vaccinated vs. unvaccinated | Mean change (95% CI) | Propensity score (based on propensity score model including age, gender, race, ethnicity, marital status, income, smoking history, comorbidities, and severity of acute COVID-19) adjusted | Anosmia (1 or 2 doses) | − 0.02 (− 0.35 to 0.31) | No difference |
|  |  |  |  |  | Anosmia (1 dose) | − 0.17 (− 0.62 to 0.28) | No difference |
|  |  |  |  |  | Anosmia (2 doses) | − 0.13 (− 0.42 to 0.15) | No difference |
|  |  |  |  |  | Dyspnoea (1 or 2 doses) | 0.05 (− 0.15 to 0.25) | No difference |
|  |  |  |  |  | Dyspnoea (1 dose) | 0.10 (− 0.20 to 0.39) | No difference |
|  |  |  |  |  | Dyspnoea (2 doses) | − 0.06 (− 0.29 to 0.16) | No difference |
|  |  |  |  |  | Cough (1 or 2 doses) | − 0.17 (− 0.55 to 0.22) | No difference |
|  |  |  |  |  | Cough (1 dose) | 0.22 (− 0.27 to 0.72) | No difference |
|  |  |  |  |  | Cough (2 doses) | − 0.32 (− 0.73 to 0.08) | No difference |
|  |  |  |  |  | Phlegm (1 or 2 doses) | − 0.47 (− 0.87 to 0.10) | No difference |
|  |  |  |  |  | Phlegm (1 dose) | − 0.28 (− 0.92to 0.36) | No difference |
|  |  |  |  |  | Phlegm (2 doses) | − 0.27 (− 0.76 to 0.21) | No difference |
|  |  |  |  |  | Wheezing (1 or 2 doses) | − 0.16 (− 0.83 to 0.50) | No difference |
|  |  |  |  |  | Wheezing (1 dose) | − 0.30 (− 1.17 to 0.56) | No difference |
|  |  |  |  |  | Wheezing (2 doses) | − 0.28 (− 0.98 to 0.42) | No difference |
|  |  |  |  |  | Depression (1 or 2 doses) | 0.02 (− 1.18 to 1.22) | No difference |
|  |  |  |  |  | Depression (1 dose) | 0.39 (− 1.15 to 1.95) | No difference |
|  |  |  |  |  | Depression (2 doses) | 0.31 (− 0.95 to 1.56) | No difference |
|  |  |  |  |  | Anxiety (1 or 2 doses) | 0.51 (− 0.93 to 0.04) | No difference |
|  |  |  |  |  | Anxiety (1 doses) | 1.87 (− 0.01 to 3.76) | No difference |
|  |  |  |  |  | Anxiety (2 doses) | 0.62 (− 0.71 to 1.95) | No difference |
|  |  |  |  |  | COVID-19-related PTSD (1 or 2 doses) | 2.53 (− 3.06 to 8.12) | No difference |
|  |  |  |  |  | COVID-19-related PTSD (1 dose) | 7.58 (− 0.21 to 15.37) | No difference |
|  |  |  |  |  | COVID-19-related PTSD (2 doses) | 2.90 (− 2.13 to 7.93) | No difference |
|  |  |  |  |  | PTSD due to other trauma (1 or 2 doses) | − 2.53 (− 12.11 to 7.04) | No difference |
|  |  |  |  |  | PTSD due to other trauma (1 dose) | − 11.62 (− 26.04 to 2.80) | No difference |
|  |  |  |  |  | PTSD due to other trauma (2 doses) | − 1.75 (− 11.85 to 8.35) | No difference |
|  |  |  |  |  | Qol, Physical function (1 or 2 doses) | − 1.16 (− 3.35 to 1.02) | No difference |
|  |  |  |  |  | Qol, Physical function (1 dose) | − 0.30 (− 2.95 to 2.36) | No difference |
|  |  |  |  |  | Qol, Physical function (2 doses) | − 0.87 (− 3.04 to 1.30) | No difference |
|  |  |  |  |  | Qol, Anxiety (1 or 2 doses) | − 0.29 (− 2.84 to 2.27) | No difference |
|  |  |  |  |  | Qol, Anxiety (1 dose) | − 0.26 (− 3.24 to 2.72) | No difference |
|  |  |  |  |  | Qol, Anxiety (2 doses) | − 0.69 (− 3.06 to 1.68) | No difference |
|  |  |  |  |  | QoL, Depression (1 or 2 doses) | − 1.12 (− 3.80 to 1.56) | No difference |
|  |  |  |  |  | QoL, Depression (1 dose) | − 1.53 (− 5.09 to 2.04) | No difference |
|  |  |  |  |  | QoL, Depression (2 doses) | 1.53 (− 1.22 to 4.28) | No difference |
|  |  |  |  |  | QoL, Fatigue (1 or 2 doses) | − 1.42 (− 4.15 to 1.32) | No difference |
|  |  |  |  |  | QoL, Fatigue (1 dose) | − 1.15 (− 4.29 to 2.00) | No difference |
|  |  |  |  |  | QoL, Fatigue (2 doses) | − 1.39 (− 3.93 to 1.15) | No difference |
|  |  |  |  |  | QoL, Social Roles (1 or 2 doses) | − 0.17 (− 3.18 to 2.83) | No difference |
|  |  |  |  |  | QoL, Social Roles (1 dose) | − 0.23 (− 3.67 to 4.13) | No difference |
|  |  |  |  |  | QoL, Social Roles (2 doses) | − 1.99 (− 5.11 to 1.14) | No difference |
|  |  |  |  |  | Qol, Sleep (1 or 2 doses) | 1.51 (− 0.86 to 3.87) | No difference |
|  |  |  |  |  | Qol, Sleep (1 dose) | 1.00 (− 1.72 to 3.74) | No difference |
|  |  |  |  |  | Qol, Sleep (2 doses) | 1.95 (− 0.21 to 4.10) | No difference |
|  |  |  |  |  | QoL, Pain (1 or 2 doses) | − 0.02 (− 2.74 to 2.70) | No difference |
|  |  |  |  |  | QoL, Pain (1 dose) | − 1.91 (− 4.74 to 0.92) | No difference |
|  |  |  |  |  | QoL, Pain (2 doses) | − 0.40 (− 2.69 to 1.89) | No difference |
| Jassat, 2023 | Mixed (primary series) | Symptoms persistent at 6 months after COVID-19 infection | IRR (95% CI) | Adjusted for, age, sex, ethnicity, presence of comorbid conditions (asthma, diabetes, hypertension, chronic cardiac disease, chronic kidney disease, malignancy, tuberculosis, HIV, and obesity), number of symptoms during acute infection, acute COVID-19 severity, COVID-19 vaccination status, and wave period | Unvaccinated (vs vaccination before infection) | 0.97 (0.80–1.18) | No difference |
|  |  |  |  |  | Vaccinated after infection (vs before infection) | 0.91 (0.75–1.1) |  |
| Nehme, 2022 | Before infection (0-, 1-, or ≥2 doses; reference= unvaccinated or 1 dose) | Any PCC symptom prevalence | Adjusted prevalence (95% CI) | Adjusted for age, sex, vaccination status, monoclonal antibody treatment during the acute phase, time from infection, and the following pre-existing comorbidities: obesity or overweight, hypertension, diabetes, respiratory disease, cardiovascular disease, headache disorders, cognitive disorders, sleep disorders, depression, anxiety, hypothyroidism, rheumatologic disease, anemia, chronic pain or fibromyalgia, chronic fatigue syndrome, and irritable bowel syndrome | Vaccinated | 9.7 (9.5–1.0) | Yes; p-value for comparison p<0.001 |
|  |  |  |  |  | Unvaccinated | 18.1 (17.5–18.8) | Similar protective effect observed for vaccinated vs. not vaccinated when PCC split by symptom: fatigue, concentration, headache, insomnia, smell, taste, dyspnoea, cough, myalgia, parenthesis, chest pain, palpitations, digestive, and hair loss |
| Hastie, 2022 | Vaccinated Before infection (at least one dose) | No recovery vs. Full recovery | AOR (95%CI) | Adjusted for socioeconomic factors (age, sex, ethnic group, deprivation) and stage of follow-up (6, 12 or 18 months); pre-existing health conditions (count, respiratory and CHD, depression, diabetes); vaccination status; dominant SARS-CoV-2 variant | At least one dose vs. unvaccinated | 0.92 (0.68, 1.26) | No difference |
|  |  | Partial recovery vs. Full recovery |  |  |  | 0.90 (0.78, 1.04) |  |

AOR: Adjusted odds ratio; BMI: Body mass index; CHD: Coronary heart disease; CI: Confidence interval; DVT: Deep vein thrombosis; EQ-5D-5L : EuroQol-5 Dimension 5-Levels; HIV: Human immunodeficiency virus; HR: Hazard ratio; ICU: Intensive care units; IRR: Incidence rate ratio; mRNA: Messenger ribonucleic acid; NA: Not applicable; NR: Not reported; PASC: Post-acute sequelae of COVID-19; PCC: Post- COVID condition; PCR: Polymerase Chain Reaction; PE: Pulmonary embolism; PTSD: Post-traumatic stress disorder; QoL: Quality of life; SD: Standard deviation; VE: Vaccine effectiveness; WHO: World Health Organization.

**eTable 11. Matrix representing the number of included studies with low risk of bias (n=80) reporting each type of adjusted estimate (n=56)**

|  | **aVE against ≥1 PCC (vs. unvaccinated) (Fig 2, Fig 3, or eFig 3)** | **Adjusted comparative VE against ≥1 PCC (vs. other ref than unvaccinated) (eFigure 1)** | **aVE against a specific PCC vs. unvaccinated) (eFig 2)** | **Adjusted estimates that could not be converted to VE (eTable 10)** | **Unadjusted estimates only (not reported)** |
| --- | --- | --- | --- | --- | --- |
| **aVE against ≥1 PCC (vs. unvaccinated) (Fig 2, Fig 3, or eFig 3)** | **39** | 9 | 3 | 1 | N/A |
| **Adjusted comparative VE against ≥1 PCC (vs. other ref than unvaccinated) (eFigure 1)** | 9 | **11** | 1 | 0 | N/A |
| **aVE against a specific PCC vs. unvaccinated) (eFig 2)** | 3 | 1 | **9** | 0 | N/A |
| **Adjusted estimates that could not be converted to VE (eTable 10)** | 1 | 0 | 0 | **10** | N/A |
| **Unadjusted estimates only (not reported)** | N/A | N/A | N/A | N/A | **24** |

PCC: post‑COVID‑19 conditions; VE: vaccine effectiveness.

Of the 80 studies with low risk of bias, 56 studies reported adjusted estimates for the relationship between vaccination and PCC and 24 studies reported only unadjusted estimates. This matrix documents the number of publications reporting each of the 4 types of adjusted estimates (bolded numbers in the diagonals) and the number of publications that report adjusted estimates corresponding with both the row title and the column title (numbers in off-diagonals that are not bolded). For example, 39 publications reported adjusted VEs (vs. unvaccinated) against ≥1 PCC, 11 studies reported adjusted comparative VEs (vs. reference group other than unvaccinated) against ≥1 PCC, 9 studies reported aVE against ≥1 specific PCC (vs. unvaccinated), and 10 publications reported adjusted estimates for the relationship between vaccination and PCC that could not be converted to VE. Of note, many publications reported estimates corresponding with more than one of the four outcomes. For example, Of the 39 studies reporting aVE (vs. unvaccinated) against ≥1 PCC, 9 also reported adjusted comparative VE against ≥1 PCC (reference group other than unvaccinated), 3 also reported aVE against a specific PCC (vs. unvaccinated), and one also reported an adjusted estimate that could not be convered to VE. In total, 13 studies reported adjusted estimates corresponding with more than one type of outcome. The remaining 43 studies reported adjusted estimates for only one type of outcome. For example, 27 studies reported only aVE against ≥1 PCC (vs. unvaccinated), 2 studies reported only adjusted comparative VE against ≥1 PCC (vs. reference group other than unvaccinated), 6 studies reported only aVE against specific PCC (vs. unvaccinated), and 9 studies reported only adjusted estimates that could not be converted to aVE.

Publications reporting adjusted estimates corresponding with two outcome types are: Bello-Chavolla , 2023; Catala , 2023; de Arriba Fernández, 2022; de Arriba Fernández, 2023; Di Fusco, 2023; Ertesvag, 2023; Jassat, 2023; Mohr, 2023; Robertson, 2022; Smits, 2023; Xue, 2023. Publications reporting adjusted estimates for three outcome types are: Al-Aly, 2022.

**eFigure 3.** **Adjusted VE (vs. unvaccinated) against ≥1 PCC for vaccination timepoints other than before infection, sorted by timing of vaccination relative to infection or PCC onset, vaccine type, and number of doses**

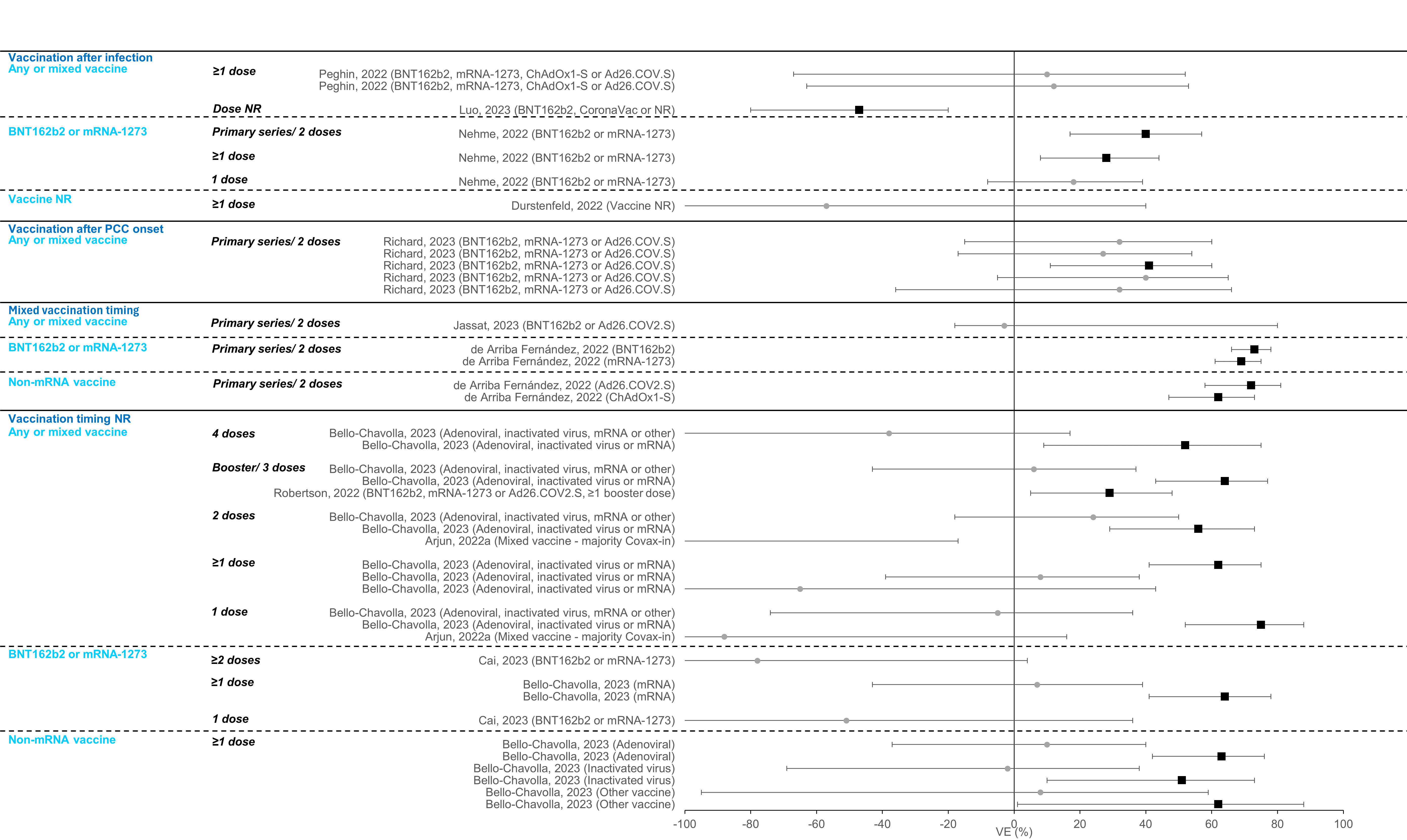

Of 42 estimates, 18 (42.9%) reported statistically significant (based on VE 95% CI) protective effect against ≥1 PCC.

CI: confidence interval; NR: not reported; PCC: post‑COVID‑19 conditions; VE: vaccine effectiveness.

References: Arjun, 2022a^8^; Bello-Chavolla, 2023^16^; Cai, 2023^21^; de Arriba Fernández, 2022^26^; Durstenfeld, 2022^30^; Jassat, 2023^48^; Luo, 2023^56^; Nehme, 2022^66^; Peghin, 2022^70^; Richard, 2023^78^; Robertson, 2022^79^

**eTable 12. Summary of publications where adjusted pre-infection VE against ≥1 PCC (vs. unvaccinated) could be computed for any or unspecified vaccine type or manufacturer, sorted by vaccine type and number of doses (data corresponding to Figure 2) (N=21 studies with 62 VE estimates)**

| **Author, year (Vaccine brand, dose)** | **Variant** | **Subgroup** | **Subgroup detail** | **Adjustments** | **VE** | **Lower 95% CI** | **Upper 95% CI** |
| --- | --- | --- | --- | --- | --- | --- | --- |
| Marra, 2023 (BNT162b2, CoronaVac, ChAdOx1-S or Ad26.COV2.S, 4 doses) | Wild-type, Gamma, Delta, Omicron | Number of doses | 4 | Adjusted for sex, age, BMI, any comorbidity, the number of SARS-CoV-2 infections, and SARS-CoV-2 variant | 95 | 81 | 99 |
| Luo, 2023 (BNT162b2, CoronaVac or NR, ≥3 doses) | Omicron | Number of doses | ≥3 doses | Adjusted for age, gender, BMI, infection days, vaccination status, vaccination after infection, comorbidities, infection days, treatment for the acute stage of illness, and patient's satisfaction of treatment | -11 | -24 | 2 |
| Smits, 2023 (BNT162b2, mRNA-1273, NVX-CoV2373 or Ad26.COV2.S, Booster) | Omicron | Vaccination status | Long COVID diagnosis (ICD-10 codes; 28-365 days post-positive COVID-19 test) | Adjusted for vaccination status, comorbidities, sex, race, ethnicity, age, timing of infection, smoking status, influenza vaccination status, inpatient and outpatient encounters in previous two years | 19 | 5 | 31 |
| Smits, 2023 (BNT162b2, mRNA-1273, NVX-CoV2373 or Ad26.COV2.S, Booster) | Omicron | Vaccination status | Long-COVID symptoms (28-365 days post-positive COVID-19 test) | Adjusted for vaccination status, comorbidities, sex, race, ethnicity, age, timing of infection, smoking status, influenza vaccination status, inpatient and outpatient encounters in previous two years | 13 | 9 | 17 |

| **Author, year (Vaccine brand, dose)** | **Variant** | **Subgroup** | **Subgroup detail** | **Adjustments** | **VE** | **Lower 95% CI** | **Upper 95% CI** |
| --- | --- | --- | --- | --- | --- | --- | --- |
| Tosi, 2023 (BNT162b2, mRNA-1273 or Ad26.COV2.S, Booster) | Omicron | Vaccination, patient frailty | Boosted, robust patient | Adjusted for age, BMI, race, ethnicity, smoking, active patient in the past 24 months, and rurality | 26 | 18 | 33 |
| Tosi, 2023 (BNT162b2, mRNA-1273 or Ad26.COV2.S, Booster) | Omicron | Vaccination, patient frailty | Boosted, pre-frail patient | Adjusted for age, BMI, race, ethnicity, smoking, active patient in the past 24 months, and rurality | 35 | 28 | 41 |
| Tosi, 2023 (BNT162b2, mRNA-1273 or Ad26.COV2.S, Booster) | Omicron | Vaccination, patient frailty | Boosted, frail patient | Adjusted for age, BMI, race, ethnicity, smoking, active patient in the past 24 months, and rurality | 30 | 23 | 36 |
| Cortellini, 2023 (BNT162b2, mRNA-1273, ChAdOx1-S or Ad26.COV2.S, 3 doses) | Alpha, Delta, Omicron | Number of doses | 3 | Inverse probability of treatment weighting applied | 53 | -18 | 82 |
| Marra, 2023 (BNT162b2, CoronaVac, ChAdOx1-S or Ad26.COV2.S, 3 doses) | Wild-type, Gamma, Delta, Omicron | Number of doses | 3 | Adjusted for sex, age, BMI, any comorbidity, the number of SARS-CoV-2 infections, and SARS-CoV-2 variant | 37 | -2 | 61 |
| Tannous, 2023 (mRNA or adenoviral, Primary series) | Wild-type, Alpha, Delta | NA | NA | Adjusted for age, sex, race, ethnicity, Charlson Comorbidity Index, COVID-19 illness severity, and Texas state area deprivation index | 42 | 34 | 48 |
| Elmazny, 2023 (BNT162b2, Ad26.COV2.S, ChAdOx1-S, Sputnik V, CoronaVac, BBIBP-CorV, Primary series) | WT, Alpha, Delta, Omicron | NA | NA | Adjusted for age, sex, and COVID-19 severity | 40 | 20 | 55 |
| Smits, 2023 (BNT162b2, mRNA-1273, NVX-CoV2373 or Ad26.COV2.S, Primary series) | Omicron | Vaccination status | Long COVID diagnosis (ICD-10 codes; 28–365 days post-positive COVID-19 test) | Adjusted for vaccination status, comorbidities, sex, race, ethnicity, age, timing of infection, smoking status, influenza vaccination status, inpatient and outpatient encounters in previous two years | 14 | 1 | 26 |
| Smits, 2023 (BNT162b2, mRNA-1273, NVX-CoV2373 or Ad26.COV2.S, Primary series) | Omicron | Vaccination status | Long-COVID symptoms (28-365 days post-positive COVID-19 test) | Adjusted for vaccination status, comorbidities, sex, race, ethnicity, age, timing of infection, smoking status, influenza vaccination status, inpatient and outpatient encounters in previous two years | 10 | 6 | 13 |
| Brannock, 2023 (BNT162b2, mRNA-1273 or ChAdOx1-S, Primary series) | Delta, Omicron | Methodology - proportional hazards | Model-based cohort IPTW-adjusted HR | Inverse probability of treatment weighting applied | 37 | 31 | 43 |
| Brannock, 2023 (BNT162b2, mRNA-1273 or ChAdOx1-S, Primary series) | Delta, Omicron | Methodology - logistic regression analysis | Model-based cohort IPTW-adjusted OR | Inverse probability of treatment weighting applied | 30 | 25 | 35 |
| Brannock, 2023 (BNT162b2, mRNA-1273 or ChAdOx1-S, Primary series) | Delta, Omicron | Methodology - proportional hazards | Clinic-based cohort IPTW-adjusted HR | Inverse probability of treatment weighting applied | 33 | 21 | 44 |
| Brannock, 2023 (BNT162b2, mRNA-1273 or ChAdOx1-S, Primary series) | Delta, Omicron | Methodology - logistic regression analysis | Clinic-based cohort IPTW-adjusted OR | Inverse probability of treatment weighting applied | 30 | 19 | 40 |
| Al-Aly, 2022 (BNT162b2, mRNA-1273 or Ad26.COV2.S, Primary series) | Wild-type, Alpha, Delta | NA | Risk of incident post-acute sequelae | Adjusted for age, race, area deprivation index, outpatient visits, inpatient visits, and unique medication prescriptions in previous year, routine blood chemistry panels, BMI, history of influenza vaccination, smoking status, comorbidities, systolic blood pressure, diastolic blood pressure, estimated glomerular filtration rate, and hospital system average hospital bed occupancy, total hospital bed capacity, positivity rate of tests, number of tests, and HHS region SARS-CoV-2 variant proportion in the week prior to enrolment, geographic region of VISN, and calendar week of enrolment | 15 | 11 | 18 |
| Al-Aly, 2022 (BNT162b2, mRNA-1273 or Ad26.COV2.S, Primary series) | Wild-type, Alpha, Delta | Setting | Non-hospitalized | Adjusted for age, race, area deprivation index, outpatient visits, inpatient visits, and unique medication prescriptions in previous year, routine blood chemistry panels, BMI, history of influenza vaccination, smoking status, comorbidities, systolic blood pressure, diastolic blood pressure, estimated glomerular filtration rate, and hospital system average hospital bed occupancy, total hospital bed capacity, positivity rate of tests, number of tests, and HHS region SARS-CoV-2 variant proportion in the week prior to enrolment, geographic region of VISN, and calendar week of enrolment | 7 | 4 | 11 |
| Al-Aly, 2022 (BNT162b2, mRNA-1273 or Ad26.COV2.S, Primary series) | Wild-type, Alpha, Delta | Setting | Hospitalized | Adjusted for age, race, area deprivation index, outpatient visits, inpatient visits, and unique medication prescriptions in previous year, routine blood chemistry panels, BMI, history of influenza vaccination, smoking status, comorbidities, systolic blood pressure, diastolic blood pressure, estimated glomerular filtration rate, and hospital system average hospital bed occupancy, total hospital bed capacity, positivity rate of tests, number of tests, and HHS region SARS-CoV-2 variant proportion in the week prior to enrolment, geographic region of VISN, and calendar week of enrolment | 12 | 4 | 19 |
| Al-Aly, 2022 (BNT162b2, mRNA-1273 or Ad26.COV2.S, Primary series) | Wild-type, Alpha, Delta | Setting | ICU | Adjusted for age, race, area deprivation index, outpatient visits, inpatient visits, and unique medication prescriptions in previous year, routine blood chemistry panels, BMI, history of influenza vaccination, smoking status, comorbidities, systolic blood pressure, diastolic blood pressure, estimated glomerular filtration rate, and hospital system average hospital bed occupancy, total hospital bed capacity, positivity rate of tests, number of tests, and HHS region SARS-CoV-2 variant proportion in the week prior to enrolment, geographic region of VISN, and calendar week of enrolment | 22 | 10 | 33 |
| Patel, 2023 (BNT162b2, mRNA-1273 or Ad26.COV2.S, Primary series) | Delta, Omicron | PASC definition | WHO; ≥90 days post-infection | Adjusted for age, sex, race, comorbidity count and use of any one of the following medications: anti-CD20 monoclonal antibodies, methotrexate, mycophenolate or glucocorticoids. | 90 | 78 | 96 |
| Patel, 2023 (BNT162b2, mRNA-1273 or Ad26.COV2.S, Primary series) | Delta, Omicron | PASC definition | CDC; ≥28 days | Adjusted for age, sex, race, comorbidity count and use of any one of the following medications: anti-CD20 monoclonal antibodies, methotrexate, mycophenolate or glucocorticoids. | 51 | 17 | 71 |
| Richard, 2023 (BNT162b2, mRNA-1273 or Ad26.COV2.S, Primary series) | Wild-type, Alpha, Delta | Timepoint | ≥90 days | Adjusted for Delta variant period, maximum symptom severity during acute illness, days hospitalized for COVID-19, comorbidities, acute respiratory symptoms, age, sex and BMI category | 27 | -14 | 53 |
| Richard, 2023 (BNT162b2, mRNA-1273 or Ad26.COV2.S, Primary series) | Wild-type, Alpha, Delta | Timepoint | ≥28 days | Adjusted for Delta variant period, maximum symptom severity during acute illness, days hospitalized for COVID-19, comorbidities, acute respiratory symptoms, age, sex and BMI category | 28 | 4 | 46 |
| Tosi, 2023 (BNT162b2, mRNA-1273 or Ad26.COV2.S, Primary series) | Omicron | Vaccination, patient frailty | Vaccinated, robust patient | Adjusted for age, BMI, race, ethnicity, smoking, active patient in the past 24 months, and rurality | 14 | 7 | 19 |
| Tosi, 2023 (BNT162b2, mRNA-1273 or Ad26.COV2.S, Primary series) | Omicron | Vaccination, patient frailty | Vaccinated, pre-frail patient | Adjusted for age, BMI, race, ethnicity, smoking, active patient in the past 24 months, and rurality | 20 | 13 | 26 |
| Tosi, 2023 (BNT162b2, mRNA-1273 or Ad26.COV2.S, Primary series) | Omicron | Vaccination, patient frailty | Vaccinated, frail patient | Adjusted for age, BMI, race, ethnicity, smoking, active patient in the past 24 months, and rurality | 20 | 13 | 27 |
| Parodi, 2023 (mRNA or inactivated viral, 2 doses) | Gamma, Delta, Omicron | NA | NA | Adjusted for age, sex, hypertension and smoking | 48 | 31 | 60 |
| Ranucci, 2023 (BNT162b2, mRNA-1273, ChAdOx1-S or Ad26.COV2.S or NR, 2 doses) | Wild-type, Alpha, Delta, Omicron | Symptom | Persistent major physical symptoms | Adjusted for age, severity of disease | 69 | 40 | 84 |
| Ranucci, 2023 (BNT162b2, mRNA-1273, ChAdOx1-S or Ad26.COV2.S or NR, 2 doses) | Wild-type, Alpha, Delta, Omicron | Symptom | Persistent major neuropsychological symptoms | Adjusted for previous neurological symptoms | 80 | 57 | 90 |
| Cortellini, 2023 (BNT162b2, mRNA-1273, ChAdOx1-S or Ad26.COV2.S, 2 doses) | Alpha, Delta, Omicron | Number of doses | 2 | Inverse probability of treatment weighting applied | 59 | 30 | 76 |
| Marra, 2023 (BNT162b2, CoronaVac, ChAdOx1-S or Ad26.COV2.S, 2 doses) | Wild-type, Gamma, Delta, Omicron | Number of doses | 2 | Adjusted for sex, age, BMI, any comorbidity, the number of SARS-CoV-2 infections, and SARS-CoV-2 variant | -17 | -76 | 21 |
| Antonelli, 2022 (BNT162b2, mRNA-1273, ChAdOx1-S or NR, 2 doses) | Alpha, Delta | Number of doses | 2 | Adjusted for age, BMI, sex | 49 | 18 | 68 |
| Ayoubkhani, 2022 (BNT162b2, ChAdOx1-S or Ad26.COV2.S, 2 doses) | Delta | Vaccine and long COVID severity | Adenoviral and mRNA; Long COVID of any severity | Adjusted for age, sex, white or non-white ethnicity, country/region of residence, area deprivation quintile group, and self-reported, pre-existing health/disability status and time from infection to follow-up for Long Covid | 41 | 31 | 50 |
| Ayoubkhani, 2022 (BNT162b2, ChAdOx1-S or Ad26.COV2.S, 2 doses) | Delta | Vaccine and long COVID severity | Adenoviral and mRNA; activity-limiting Long COVID | Adjusted for age, sex, white or non-white ethnicity, country/region of residence, area deprivation quintile group, and self-reported, pre-existing health/disability status and time from infection to follow-up for Long Covid | 41 | 27 | 52 |
| Abu Hamdh, 2023 (BNT162b2, mRNA-1273, Sputnik V, Sputnik Light or ChAdOx1-S, ≥1 dose) | NR | NA | NA | Adjusted for sex, age, comorbidities, hospital admission, dyspnoea, previous COVID-19 infection, fatigue | 86 | 76 | 91 |
| Català, 2023 (BNT162b2, mRNA-1273, ChAdOx1-S or Ad26.COV2.S, 1 dose) | Delta, Omicron | Cohort | CPRD AURUM | Propensity score overlap weighting for age (5-year age bands for people aged ≥18 years), location, index date, previous observation years, number of previous GP visits, number of previous SARS-CoV-2 PCR tests, regional vaccination, testing, COVID-19 incidence rates | 36 | 26 | 45 |
| Català, 2023 (BNT162b2, mRNA-1273, ChAdOx1-S or Ad26.COV2.S, 1 dose) | Delta, Omicron | Cohort | CPRD AURUM, ≥28 days | Propensity score overlap weighting for age (5-year age bands for people aged ≥18 years), location, index date, previous observation years, number of previous GP visits, number of previous SARS-CoV-2 PCR tests, regional vaccination, testing, COVID-19 incidence rates | 30 | 16 | 42 |
| Català, 2023 (BNT162b2, mRNA-1273, ChAdOx1-S or Ad26.COV2.S, 1 dose) | Delta, Omicron | Cohort | CPRD ARUM, GP diagnosis | Propensity score overlap weighting for age (5-year age bands for people aged ≥18 years), location, index date, previous observation years, number of previous GP visits, number of previous SARS-CoV-2 PCR tests, regional vaccination, testing, COVID-19 incidence rates | 36 | 23 | 46 |
| Català, 2023 (BNT162b2, mRNA-1273, ChAdOx1-S or Ad26.COV2.S, 1 dose) | Delta, Omicron | Cohort | CORIVA | Propensity score overlap weighting for age (5-year age bands for people aged ≥18 years), location, index date, previous observation years, number of previous GP visits, number of previous SARS-CoV-2 PCR tests, regional vaccination, testing | 38 | 22 | 51 |
| Català, 2023 (BNT162b2, mRNA-1273, ChAdOx1-S or Ad26.COV2.S, 1 dose) | Delta, Omicron | Cohort | CORIVA, ≥28 days | Propensity score overlap weighting for age (5-year age bands for people aged ≥18 years), location, index date, previous observation years, number of previous GP visits, number of previous SARS-CoV-2 PCR tests, regional vaccination, testing | 38 | 23 | 50 |
| Català, 2023 (BNT162b2, mRNA-1273, ChAdOx1-S or Ad26.COV2.S, 1 dose) | Delta, Omicron | Cohort | CPRD GOLD | Propensity score overlap weighting for age (5-year age bands for people aged ≥18 years), location, index date, previous observation years, number of previous GP visits, number of previous SARS-CoV-2 PCR tests, regional vaccination, testing, COVID-19 incidence rates | 45 | 35 | 54 |
| Català, 2023 (BNT162b2, mRNA-1273, ChAdOx1-S or Ad26.COV2.S, 1 dose) | Delta, Omicron | Cohort | CPRD GOLD, ≥28 days | Propensity score overlap weighting for age (5-year age bands for people aged ≥18 years), location, index date, previous observation years, number of previous GP visits, number of previous SARS-CoV-2 PCR tests, regional vaccination, testing, COVID-19 incidence rates | 34 | 17 | 47 |
| Català, 2023 (BNT162b2, mRNA-1273, ChAdOx1-S or Ad26.COV2.S, 1 dose) | Delta, Omicron | Cohort | SIDIAP | Propensity score overlap weighting for age (5-year age bands for people aged ≥18 years), location, index date, previous observation years, number of previous GP visits, number of previous SARS-CoV-2 PCR tests, regional vaccination, testing | 16 | 7 | 25 |
| Català, 2023 (BNT162b2, mRNA-1273, ChAdOx1-S or Ad26.COV2.S, 1 dose) | Delta, Omicron | Cohort | SIDIAP, ≥28 days | Propensity score overlap weighting for age (5-year age bands for people aged ≥18 years), location, index date, previous observation years, number of previous GP visits, number of previous SARS-CoV-2 PCR tests, regional vaccination, testing | 12 | 2 | 21 |
| Català, 2023 (BNT162b2, mRNA-1273, ChAdOx1-S or Ad26.COV2.S, 1 dose) | Delta, Omicron | Cohort | SIDIAP, GP diagnosis | Propensity score overlap weighting for age (5-year age bands for people aged ≥18 years), location, index date, previous observation years, number of previous GP visits, number of previous SARS-CoV-2 PCR tests, regional vaccination, testing | 40 | 33 | 48 |
| de Arriba Fernández, 2023 (BNT162b2, mRNA-1273, ChAdOx1-S or Ad26.COV2.S, 1 dose) | Delta, Omicron | Vaccination status | Incomplete | Adjusted for age, sex, comorbidity history, body mass index, date of first, second and booster doses of a COVID-19 vaccine, type of COVID-19 vaccine, death, immunosuppressive treatment, diagnosis of persistent COVID-19, and diagnostic test results | 82 | 71 | 89 |
| Marra, 2023 (BNT162b2, CoronaVac, ChAdOx1-S or Ad26.COV2.S, 1 dose) | Wild-type, Gamma, Delta, Omicron | Number of doses | 1 | Adjusted for sex, age, BMI, any comorbidity, the number of SARS-CoV-2 infections, and SARS-CoV-2 variant | 9 | -39 | 40 |
| Antonelli, 2022 (BNT162b2, mRNA-1273, ChAdOx1-S or NR, 1 dose) | Alpha, Delta | Number of doses | 1 | Adjusted for age, BMI, sex | -3 | -24 | 15 |
| Senjam, 2022 (Vaccine NR, 2 doses) | Delta | NA | NA | Adjusted for severity of COVID-19, chronic medication use, presence of chronic conditions, self-perceived health status, monthly household income, smoking status, living status, educational level, current occupation, marital status, gender and age | 39 | 4 | 60 |
| Antonelli, 2022b (Vaccine NR, ≥1 dose) | Delta, Omicron | Time since vaccination | Vaccinated >6 months prior to infection | Adjusted by age, sex, BMI, index of multiple deprivation, presence of comorbidities and vaccination status | 74 | 68 | 80 |
| Antonelli, 2022b (Vaccine NR, ≥1 dose) | Delta, Omicron | Time since vaccination | Vaccinated 3–6 months prior to infection | Adjusted by age, sex, BMI, index of multiple deprivation, presence of comorbidities and vaccination status | 76 | 68 | 81 |
| Antonelli, 2022b (Vaccine NR, ≥1 dose) | Delta, Omicron | Time since vaccination | Vaccinated <3 months prior to infection | Adjusted by age, sex, BMI, index of multiple deprivation, presence of comorbidities and vaccination status | 50 | 41 | 57 |
| Antonelli, 2022b (Vaccine NR, ≥1 dose) | Delta, Omicron | Age; Time since vaccination | 18–59; vaccinated >6 months prior to infection | Adjusted by age, sex, BMI, index of multiple deprivation, presence of comorbidities and vaccination status | 77 | 68 | 84 |
| Antonelli, 2022b (Vaccine NR, ≥1 dose) | Delta, Omicron | Age; Time since vaccination | 18–59; vaccinated 3–6 months prior to infection | Adjusted by age, sex, BMI, index of multiple deprivation, presence of comorbidities and vaccination status | 67 | 56 | 76 |
| Antonelli, 2022b (Vaccine NR, ≥1 dose) | Delta, Omicron | Age; Time since vaccination | 18–59; vaccinated <3 months prior to infection | Adjusted by age, sex, BMI, index of multiple deprivation, presence of comorbidities and vaccination status | 59 | 47 | 68 |
| Antonelli, 2022b (Vaccine NR, ≥1 dose) | Delta, Omicron | Age; Time since vaccination | ≥60; vaccinated >6 months prior to infection | Adjusted by age, sex, BMI, index of multiple deprivation, presence of comorbidities and vaccination status | 72 | 61 | 80 |
| Antonelli, 2022b (Vaccine NR, ≥1 dose) | Delta, Omicron | Age; Time since vaccination | ≥60; vaccinated 3–6 months prior to infection | Adjusted by age, sex, BMI, index of multiple deprivation, presence of comorbidities and vaccination status | 54 | -3 | 80 |
| Antonelli, 2022b (Vaccine NR, ≥1 dose) | Delta, Omicron | Age; Time since vaccination | ≥60; vaccinated <3 months prior to infection | Adjusted by age, sex, BMI, index of multiple deprivation, presence of comorbidities and vaccination status | 46 | 32 | 57 |
| Durstenfeld, 2022 (Vaccine NR, ≥1 dose) | Wild-type, Alpha, Delta, Omicron | NA | NA | Adjusted for age, COVID-19 wave, number of initial symptoms, sex, myocardial infarction, stroke, atrial fibrillation, sleep apnoea, COPD or asthma, and immunodeficiency, plus receipt of COVID-19 vaccination before or after COVID-19 diagnosis, ethnicity, subjective socioeconomic status, highest level of education, primary employment, pre-COVID depressive symptoms, anxiety symptoms, and financial security | 19 | -49 | 56 |
| Senjam, 2022 (Vaccine NR, 1 dose) | Delta | NA | NA | Adjusted for age, gender, educational level, BMI, workplace, blood group, smoking, alcohol consumption, presence of comorbidities, oxygen requirement, severity of COVID-19 disease, overall health status following COVUD-19 illness | 9 | -40 | 41 |

BMI: Body mass index; CDC: Centers for Disease Control and Prevention; CI: confidence interval; CPRD: Clinical Practice Research Datalink; GP: General Practitioner; HHS: Health and Human Services; HR: Hazard ratio; ICD-10: International Classification of Diseases- 10; IPTW: Inverse probability of treatment weighting; mRNA: Messenger ribonucleic acid; NA: Not applicable; NR: Not reported; OR: Odds ratio; PASC: Post-acute sequelae of COVID-19; PCR: Polymerase Chain Reaction; VE: vaccine effectiveness; VISN: Veterans Integrated Services Network; WHO: World Health Organization*.*

**eTable 13. Summary of publications where adjusted pre-infection VE against ≥1 PCC (vs. unvaccinated) could be computed for mRNA and non-mRNA vaccines, by Vaccine Type, Manufacturer, and Dose (data corresponding to Figure 3) (N=12 studies with 31 VE estimates)**

| **Author, year (Vaccine, dose)** | **Variant** | **Subgroup** | **Subgroup detail** | **Adjustments** | **VE** | **Lower 95% CI** | **Upper 95% CI** |
| --- | --- | --- | --- | --- | --- | --- | --- |
| Azzolini, 2022 (BNT162b2, 3 doses) | Wild-type, Alpha, Delta, Omicron | Number of doses | 3 | Adjusted for sex, vaccine dose, allergies and comorbidities | 84 | 16 | 97 |
| Di Fusco, 2023 (BNT162b2, 3 doses) | Omicron | Vaccination status | Boosted vs unvaccinated (≥3 symptoms) | Adjusted for timepoint, vaccination status and interaction of time by vaccination status, participant pre-COVID-19 symptom onset score, sociodemographic characteristics (age, sex, regions, social vulnerability, race/ethnicity, high risk occupations), previously tested positive for COVID-19, severity of acute illness and immunocompromised status | 64 | 13 | 85 |
| Di Fusco, 2023 (BNT162b2, 3 doses) | Omicron | Vaccination status | Boosted vs unvaccinated (≥2 symptoms) | Adjusted for timepoint, vaccination status and interaction of time by vaccination status, participant pre-COVID-19 symptom onset score, sociodemographic characteristics (age, sex, regions, social vulnerability, race/ethnicity, high risk occupations), previously tested positive for COVID-19, severity of acute illness and immunocompromised status | 70 | 30 | 87 |

| **Author, year (Vaccine, dose)** | **Variant** | **Subgroup** | **Subgroup detail** | **Adjustments** | **VE** | **Lower 95% CI** | **Upper 95% CI** |
| --- | --- | --- | --- | --- | --- | --- | --- |
| Azzolini, 2022 (BNT162b2, 2 doses) | Wild-type, Alpha, Delta, Omicron | Number of doses | 2 | Adjusted for sex, vaccine dose, allergies and comorbidities | 75 | 13 | 93 |
| Di Fusco, 2023 (BNT162b2, 2 doses) | Omicron | Vaccination status | Primed vs unvaccinated (≥3 symptoms) | Adjusted for timepoint, vaccination status and interaction of time by vaccination status, participant pre-COVID-19 symptom onset score, sociodemographic characteristics (age, sex, regions, social vulnerability, race/ethnicity, high risk occupations), previously tested positive for COVID-19, severity of acute illness and immunocompromised status | 40 | -34 | 73 |
| Di Fusco, 2023 (BNT162b2, 2 doses) | Omicron | Vaccination status | Primed vs unvaccinated (≥2 symptoms) | Adjusted for timepoint, vaccination status and interaction of time by vaccination status, participant pre-COVID-19 symptom onset score, sociodemographic characteristics (age, sex, regions, social vulnerability, race/ethnicity, high risk occupations), previously tested positive for COVID-19, severity of acute illness and immunocompromised status | 36 | -39 | 70 |
| Azzolini, 2022 (BNT162b2, 1 dose) | Wild-type, Alpha, Delta, Omicron | Number of doses | 1 | Adjusted for sex, vaccine dose, allergies and comorbidities | 14 | -249 | 79 |
| Català, 2023 (BNT162b2, 1 dose) | Delta, Omicron | Vaccination group | CPRD AURUM (Pfizer vaccine) | Propensity score overlap weighting for age (5-year age bands for people aged ≥18 years), location, index date, previous observation years, number of previous GP visits, number of previous SARS-CoV-2 PCR tests, regional vaccination, testing, COVID-19 incidence rates | 47 | 38 | 55 |
| Català, 2023 (BNT162b2, 1 dose) | Delta, Omicron | Vaccination group | CORIVA (Pfizer vaccine) | Propensity score overlap weighting for age (5-year age bands for people aged ≥18 years), location, index date, previous observation years, number of previous GP visits, number of previous SARS-CoV-2 PCR tests, regional vaccination, testing | 37 | 20 | 50 |
| Català, 2023 (BNT162b2, 1 dose) | Delta, Omicron | Vaccination group | CPRD GOLD (Pfizer vaccine) | Propensity score overlap weighting for age (5-year age bands for people aged ≥18 years), location, index date, previous observation years, number of previous GP visits, number of previous SARS-CoV-2 PCR tests, regional vaccination, testing, COVID-19 incidence rates | 51 | 37 | 61 |
| Català, 2023 (BNT162b2, 1 dose) | Delta, Omicron | Vaccination group | SIDIAP (Pfizer vaccine) | Propensity score overlap weighting for age (5-year age bands for people aged ≥18 years), location, index date, previous observation years, number of previous GP visits, number of previous SARS-CoV-2 PCR tests, regional vaccination, testing | 14 | 5 | 23 |
| Gallant, 2022 (BNT162b2 or mRNA-1273, ≥2 doses) | Delta | NA | NA | Adjusted for age, sex, number of symptoms during acute COVID-19, and number of comorbidities. | -3 | -18 | 10 |
| Ioannou, 2022 (BNT162b2 or mRNA-1273, 2 doses) | Wild-type, Alpha | NA | NA | Adjusted for age, sex, race, ethnicity, urban vs rural residence, Charlson Comorbidity Index score, VA Integrated Service Network, time period of infection/pandemic wave, number of primary care, mental health and specialty care encounters in the 2 years before infection | 22 | 10 | 32 |
| Ayoubkhani 2022 (BNT162b2 or mRNA-1273, 2 doses) | Delta | Vaccine and long COVID severity | mRNA vaccine platform; activity-limiting Long COVID | Adjusted for age, sex, white or non-white ethnicity, country/region of residence, area deprivation quintile group, and self-reported, pre-existing health/disability status and time from infection to follow-up for Long Covid | 50 | 25 | 56 |
| Ayoubkhani 2022 (BNT162b2 or mRNA-1273, 2 doses) | Delta | Vaccine and long COVID severity | mRNA vaccine platform; Long COVID of any severity | Adjusted for age, sex, white or non-white ethnicity, country/region of residence, area deprivation quintile group, and self-reported, pre-existing health/disability status and time from infection to follow-up for Long Covid | 50 | 31 | 63 |
| Mohr, 2023 (BNT162b2 or mRNA-1273, 2 doses) | Wild-type, Alpha, Delta | Timepoint | ≤16 weeks since vaccination; COVID-like symptoms | Adjusted for age, race and ethnicity, comorbidities, calendar month of diagnosis, and weeks since symptoms started | 33 | 13 | 49 |
| Mohr, 2023 (BNT162b2 or mRNA-1273, 2 doses) | Wild-type, Alpha, Delta | Timepoint | >16 weeks since vaccination; COVID-like symptoms | Adjusted for age, race and ethnicity, comorbidities, calendar month of diagnosis, and weeks since symptoms started | 28 | 12 | 41 |
| Mohr, 2023 (BNT162b2 or mRNA-1273, 2 doses) | Wild-type, Alpha, Delta | Timepoint | 6 weeks | Adjusted for age, race and ethnicity, comorbidities, calendar month of diagnosis and weeks since symptoms started | 30 | 16 | 42 |
| Kahlert, 2023 (BNT162b2 or mRNA-1273, ≥1 dose) | Wild-type, Alpha, Delta, Omicron | NA | NA | Adjusted for age, sex, BMI, ethnicity, children in household, comorbidity, medication, smoker status, alcohol intake, medical profession, COVID-19 patient contact, ICU work, full-time work, SARS-CoV-2 variant | -27 | -94 | 18 |
| Ertesvag, 2023 (BNT162b2 or mRNA-1273, ≥1 dose) | Delta | Timepoint and population | Vaccination as a predictor of any persisting symptoms among adolescents (≥16 years) 8-months post-Delta infection | Adjusted for female sex, age, reinfection, and any symptoms at baseline | -98 | -457 | 30 |
| Ertesvag, 2023 (BNT162b2 or mRNA-1273, ≥1 dose) | Delta | Timepoint and population | Vaccination as a predictor of any persisting symptoms 3-months post-Delta infection | Adjusted by female sex, age, reinfection, any symptoms at baseline. | -123 | -904 | 45 |
| Ioannou, 2022 (BNT162b2 or mRNA-1273, 1 dose) | Wild-type, Alpha | NA | NA | Adjusted for age, sex, race, ethnicity, urban vs rural residence, Charlson Comorbidity Index score, VA Integrated Service Network, time period of infection/pandemic wave, number of primary cares, mental health and specialty care encounters in the 2 years before infection | -3 | -10 | 5 |
| Xue, 2023 (BNT162b2 or mRNA-1273, 1 dose) | Alpha, Gamma, Delta, Omicron | NA | NA | Adjusted for age, gender, ethnicity, smoking status, alcohol consumption, educational level, since first vaccination, marital status, living area, level of physical activity, weight status, comorbidities. | -13 | -34 | 5 |
| Perlis, 2022 (Ad26.COV2.S, 2 doses) | Alpha, Delta, Omicron | NA | NA | Adjusted for age, gender, race and ethnicity, educational level, urbanicity, region and predominant variant | 28 | 14 | 40 |
| Perlis, 2022 (Ad26.COV2.S, 1 dose) | Alpha, Delta, Omicron | NA | NA | Adjusted for age, gender, race and ethnicity, educational level, urbanicity, region and predominant variant | 7 | -25 | 31 |
| Ayoubkhani, 2022 (ChAdOx1-S, 2 doses) | Delta | Vaccine and long COVID severity | AstraZeneca; activity limiting Long COVID | Adjusted for age, sex, white or non-white ethnicity, country/region of residence, area deprivation quintile group, and self-reported, pre-existing health/disability status and time from infection to follow-up for Long Covid | 37 | 20 | 51 |
| Ayoubkhani, 2022 (ChAdOx1-S, 2 doses) | Delta | Vaccine and long COVID severity | AstraZeneca; Long COVID of any severity | Adjusted for age, sex, white or non-white ethnicity, country/region of residence, area deprivation quintile group, and self-reported, pre-existing health/disability status and time from infection to follow-up for Long Covid | 38 | 25 | 49 |
| Català, 2023 (ChAdOx1-S, 1 dose) | Delta, Omicron | Vaccination group | CPRD AURUM (AstraZeneca vaccine) | Propensity score overlap weighting for age (5-year age bands for people aged ≥18 years), location, index date, previous observation years, number of previous GP visits, number of previous SARS-CoV-2 PCR tests, regional vaccination, testing, COVID-19 incidence rates | 29 | 21 | 37 |
| Català, 2023 (ChAdOx1-S, 1 dose) | Delta, Omicron | Vaccination group | CPRD GOLD (AstraZeneca vaccine) | Propensity score overlap weighting for age (5-year age bands for people aged ≥18 years), location, index date, previous observation years, number of previous GP visits, number of previous SARS-CoV-2 PCR tests, regional vaccination, testing, COVID-19 incidence rates | 37 | 23 | 48 |
| Català, 2023 (ChAdOx1-S, 1 dose) | Delta, Omicron | Vaccination group | SIDIAP (AstraZeneca vaccine) | Propensity score overlap weighting for age (5-year age bands for people aged ≥18 years), location, index date, previous observation years, number of previous GP visits, number of previous SARS-CoV-2 PCR tests, regional vaccination, testing | 16 | 4 | 27 |
| Herman, 2022 (inactivated viral vaccine, 2 doses) | Delta, Omicron | NA | NA | Adjusted by shortness of breath | 26 | -8 | 49 |

BMI: Body mass index; CI: Confidence interval; CPRD: Clinical Practice Research Datalink; GP: General Practitioner; ICU: Intensive care unit; mRNA: Messenger ribonucleic acid; NA: Not applicable; NR: Not reported; PCR: Polymerase Chain Reaction; VA: Veterans Affairs; VE: Vaccine effectiveness.

**eTable 14. Summary of publications where adjusted VE (vs. unvaccinated) against ≥1 PCC could be computed for vaccination timepoints other than before infection, sorted by timing of vaccination relative to infection or PCC onset, vaccine type (any, mRNA, non-mRNA), and number of doses (N=11 studies with 41 VE estimates)**

| **Author year (Vaccine brand, dose)** | **Vaccination timepoint** | **Variant** | **Subgroup** | **Subgroup details** | **Adjustments** | **VE** | **Lower 95% CI** | **Upper 95% CI** |
| --- | --- | --- | --- | --- | --- | --- | --- | --- |
| Peghin, 2022 (BNT162b2, mRNA-1273, ChAdOx1-S or Ad26.COV.S) | After infection | Wild-type, Alpha | NA | NA | Adjusted for gender, age, presence of symptoms at acute COVID-19 onset, acute COVID-19 management and IgG antibodies | 10 | -67 | 52 |
| Peghin, 2022 (BNT162b2, mRNA-1273, ChAdOx1-S or Ad26.COV.S) | After infection | Wild-type, Alpha | NA | NA | Adjusted for gender, age, acute COVID-19 severity and IgG antibodies | 12 | -63 | 53 |
| Luo, 2023 (BNT162b2, CoronaVac or NR) | After infection | Omicron | NA | NA | Adjusted for age, gender, BMI, infection days, vaccination status, vaccination after infection, comorbidities, infection days, treatment for the acute stage of illness, and patient's satisfaction of treatment | -47 | -80 | -20 |
| Nehme, 2022 (BNT162b2 or mRNA-1273) | After infection | Delta | Number of doses | 2 | Adjusted for time for infection, age, sex, smoking, and the following comorbidities present prior to the infection: overweight or obese, hypertension, respiratory disease, cardiovascular disease, diabetes, migraine, tension headache, sleeping disorder, anxiety, depression, hypothyroidism, anemia, and chronic fatigue. | 40 | 17 | 57 |
| Nehme, 2022 (BNT162b2 or mRNA-1273) | After infection | Delta | Number of doses | 1 or 2 | Adjusted for time for infection, age, sex, smoking, and the following comorbidities present prior to the infection: overweight or obese, hypertension, respiratory disease, cardiovascular disease, diabetes, migraine, tension headache, sleeping disorder, anxiety, depression, hypothyroidism, anemia, and chronic fatigue. | 28 | 8 | 44 |
| Nehme, 2022 (BNT162b2 or mRNA-1273) | After infection | Delta | Number of doses | 1 | Adjusted for time for infection, age, sex, smoking, and the following comorbidities present prior to the infection: overweight or obese, hypertension, respiratory disease, cardiovascular disease, diabetes, migraine, tension headache, sleeping disorder, anxiety, depression, hypothyroidism, anemia, and chronic fatigue. | 18 | -8 | 39 |
| Durstenfeld, 2022 (Vaccine NR) | After infection | Wild-type, Alpha, Delta, Omicron | NA | NA | Adjusted for age, COVID-19 wave, number of initial symptoms, sex, myocardial infarction, stroke, atrial fibrillation, sleep apnoea, COPD or asthma, and immunodeficiency, plus receipt of COVID-19 vaccination before or after COVID-19 diagnosis, ethnicity, subjective socioeconomic status, highest level of education, primary employment, pre-COVID depressive symptoms, anxiety symptoms, and financial security | -57 | -313 | 40 |
| Richard, 2023 (BNT162b2, mRNA-1273 or Ad26.COV.S) | After PCC onset | Wild-type, Alpha, Delta | Timepoint | 12 months | Adjusted for Delta variant period, maximum symptom severity during acute illness, days hospitalized for COVID-19, comorbidities, acute respiratory symptoms, age, sex and BMI category | 32 | -15 | 60 |
| Richard, 2023 (BNT162b2, mRNA-1273 or Ad26.COV.S) | After PCC onset | Wild-type, Alpha, Delta | Timepoint | 9 months | Adjusted for Delta variant period, maximum symptom severity during acute illness, days hospitalized for COVID-19, comorbidities, acute respiratory symptoms, age, sex and BMI category | 27 | -17 | 54 |
| Richard, 2023 (BNT162b2, mRNA-1273 or Ad26.COV.S) | After PCC onset | Wild-type, Alpha, Delta | Timepoint | 6 months | Adjusted for Delta variant period, maximum symptom severity during acute illness, days hospitalized for COVID-19, comorbidities, acute respiratory symptoms, age, sex and BMI category | 41 | 11 | 60 |
| Richard, 2023 (BNT162b2, mRNA-1273 or Ad26.COV.S) | After PCC onset | Wild-type, Alpha, Delta | Timepoint | 3 months | Adjusted for Delta variant period, maximum symptom severity during acute illness, days hospitalized for COVID-19, comorbidities, acute respiratory symptoms, age, sex and BMI category | 40 | -5 | 65 |
| Richard, 2023 (BNT162b2, mRNA-1273 or Ad26.COV.S) | After PCC onset | Wild-type, Alpha, Delta | Timepoint | 1 month | Adjusted for Delta variant period, maximum symptom severity during acute illness, days hospitalized for COVID-19, comorbidities, acute respiratory symptoms, age, sex and BMI category | 32 | -36 | 66 |
| Jassat, 2023 (BNT162b2 or Ad26.COV2.S) | Mixed | Beta, Delta, Omicron | NA | NA | Adjusted for age, sex, race/ethnicity, comorbidities, number of symptoms during acute infection, acute COVID-19 severity, COVID-19 vaccination status, and wave period | -3 | -18 | 80 |
| de Arriba Fernández, 2022 (BNT162b2) | Mixed | Delta, Omicron | Vaccine type | Pfizer | Adjusted for age, sex, immunosuppressive treatment, analytical determinations (high and low density lipoprotein cholesterol, total cholesterol, triglycerides and fasting glucose levels), COVID-19 vaccine, complete vaccination schedule, booster dose (3rd dose), and comorbidities | 73 | 66 | 78 |
| de Arriba Fernández, 2022 (mRNA-1273) | Mixed | Delta, Omicron | Vaccine type | Moderna | Adjusted for age, sex, immunosuppressive treatment, analytical determinations (high and low density lipoprotein cholesterol, total cholesterol, triglycerides and fasting glucose levels), COVID-19 vaccine, complete vaccination schedule, booster dose (3rd dose), and comorbidities | 69 | 61 | 75 |
| de Arriba Fernández, 2022 (Ad26.COV2.S) | Mixed | Delta, Omicron | Vaccine type | Janssen | Adjusted for age, sex, immunosuppressive treatment, analytical determinations (high and low density lipoprotein cholesterol, total cholesterol, triglycerides and fasting glucose levels), COVID-19 vaccine, complete vaccination schedule, booster dose (3rd dose), and comorbidities | 72 | 58 | 81 |
| de Arriba Fernández, 2022 (ChAdOx1-S) | Mixed | Delta, Omicron | Vaccine type | AstraZeneca | Adjusted for age, sex, immunosuppressive treatment, analytical determinations (high and low density lipoprotein cholesterol, total cholesterol, triglycerides and fasting glucose levels), COVID-19 vaccine, complete vaccination schedule, booster dose (3rd dose), and comorbidities | 62 | 47 | 73 |
| Bello-Chavolla, 2023 (Adenoviral, inactivated virus, mRNA or other) | NR | Delta, Omicron | Dose and long COVID definition | 4, WHO definition | Adjusted by survey weights | -38 | -132 | 17 |
| Bello-Chavolla, 2023 (Adenoviral, inactivated virus or mRNA) | NR | Delta, Omicron | Dose and long COVID definition | 4, PASC score ≥12 | Adjusted by survey weights | 52 | 9 | 75 |
| Bello-Chavolla, 2023 (Adenoviral, inactivated virus, mRNA or other) | NR | Delta, Omicron | Dose and long COVID definition | 3, WHO definition | Adjusted by survey weights | 6 | -43 | 37 |
| Bello-Chavolla, 2023 (Adenoviral, inactivated virus or mRNA) | NR | Delta, Omicron | Dose and long COVID definition | 3, PASC score ≥12 | Adjusted by survey weights | 64 | 43 | 77 |
| Robertson, 2022 (BNT162b2, mRNA-1273 or Ad26.COV2.S, ≥1 booster dose) | NR | Omicron | Dose | ≥1 Booster dose | Adjusted for age and gender | 29 | 5 | 48 |
| Bello-Chavolla, 2023 (Adenoviral, inactivated virus, mRNA or other) | NR | Delta, Omicron | Dose and long COVID definition | 2, WHO definition | Adjusted by survey weights | 24 | -18 | 50 |
| Bello-Chavolla, 2023 (Adenoviral, inactivated virus or mRNA) | NR | Delta, Omicron | Dose and long COVID definition | 2, PASC score ≥12 | Adjusted by survey weights | 56 | 29 | 73 |
| Arjun, 2022a (Mixed vaccine - majority Covax-in) | NR | Delta | Number of doses | 2 | Adjusted for age, sex, occupation, BMI, medical history (including related to acute COVID-19 disease), and COVID-19 vaccination | -132 | -358 | -17 |
| Bello-Chavolla, 2023 (Adenoviral, inactivated virus or mRNA) | NR | Delta, Omicron | Long COVID definition | Presence of any persistent COVID-19 symptom | Adjusted by survey weights | 62 | 41 | 75 |
| Bello-Chavolla, 2023 (Adenoviral, inactivated virus or mRNA) | NR | Delta, Omicron | Long COVID definition | PASC defined by PASC score ≥12 points | Adjusted by survey weights | 8 | -39 | 38 |
| Bello-Chavolla, 2023 (Adenoviral, inactivated virus or mRNA) | NR | Delta, Omicron | Long COVID definition | Incapacitating persistent post COVID-19 sequelae disaggregated by PASC scores using a 12-point threshold | Adjusted by survey weights | -65 | -503 | 43 |
| Bello-Chavolla, 2023 (Adenoviral, inactivated virus, mRNA or other) | NR | Delta, Omicron | Dose and long COVID definition | 1, WHO definition | Adjusted by survey weights | -5 | -74 | 36 |
| Bello-Chavolla, 2023 (Adenoviral, inactivated virus or mRNA) | NR | Delta, Omicron | Dose and long COVID definition | 1, PASC score ≥12 | Adjusted by survey weights | 75 | 52 | 88 |
| Arjun, 2022a (Mixed vaccine - majority Covax-in) | NR | Delta | Number of doses | 1 | Adjusted for age, sex, occupation, BMI, medical history (including related to acute COVID-19 disease), and COVID-19 vaccination | -88 | -322 | 16 |
| Cai, 2023 (BNT162b2 or mRNA-1273) | NR | Omicron | Number of doses | ≥2 | Adjusted for: age, number of comorbidities, severity of acute infection | -78 | -228 | 4 |
| Bello-Chavolla, 2023 (mRNA) | NR | Delta, Omicron | Vaccine and long COVID definition | mRNA; WHO definition | Adjusted by survey weights | 7 | -43 | 39 |
| Bello-Chavolla, 2023 (mRNA) | NR | Delta, Omicron | Vaccine and long COVID definition | mRNA; PASC defined by PASC score ≥12 points | Adjusted by survey weights | 64 | 41 | 78 |
| Cai, 2023 (BNT162b2 or mRNA-1273) | NR | Omicron | Number of doses | 1 | Adjusted for: age, number of comorbidities, severity of acute infection | -51 | -259 | 36 |
| Bello-Chavolla, 2023 (Adenoviral) | NR | Delta, Omicron | Vaccine and long COVID definition | Adenovirus vector; WHO definition | Adjusted by survey weights | 10 | -37 | 40 |
| Bello-Chavolla, 2023 (Adenoviral) | NR | Delta, Omicron | Vaccine and long COVID definition | Adenovirus vector; PASC defined by PASC score ≥12 points | Adjusted by survey weights | 63 | 42 | 76 |
| Bello-Chavolla, 2023 (Inactivated virus) | NR | Delta, Omicron | Vaccine and long COVID definition | Inactivated virus; WHO definition | Adjusted by survey weights | -2 | -69 | 38 |
| Bello-Chavolla, 2023 (Inactivated virus) | NR | Delta, Omicron | Vaccine and long COVID definition | Inactivated virus; PASC defined by PASC score ≥12 points | Adjusted by survey weights | 51 | 10 | 73 |
| Bello-Chavolla, 2023 (Other vaccine) | NR | Delta, Omicron | Vaccine and long COVID definition | Other; WHO definition | Adjusted by survey weights | 8 | -95 | 59 |
| Bello-Chavolla, 2023 (Other vaccine) | NR | Delta, Omicron | Vaccine and long COVID definition | Other; PASC defined by PASC score ≥12 points | Adjusted by survey weights | 62 | 1 | 88 |

BMI: Body mass index; CI: Confidence interval; mRNA: Messenger ribonucleic acid; NA: Not applicable; NR: Not reported; PASC: Post-acute sequelae of COVID-19; PCC: post‑COVID‑19 conditions; SD: Standard deviation; VE: Vaccine effectiveness; WHO: World Health Organization.

**eTable 15. Summary of pre-infection aVE against ≥1 PCC (vs. unvaccinated) by Vaccine Type, Manufacturer, and Number of Doses Received**

|  | Number of estimates | Number of statistically significant estimates | Proportion of estimates that are statistically significant (%) | Minimum statistically significant VE | Maximum statistically significant VE |
| --- | --- | --- | --- | --- | --- |
| Any or mixed vaccine | | | | | |
| Overall | 50 | 43 | 86 | 7 | 95 |
| 1 dose | 13 | 11 | 85 | 12 | 82 |
| ≥1 dose | 1 | 1 | 100 | 86 | 86 |
| Primary series/ 2 doses | 27 | 25 | 93 | 7 | 90 |
| Booster/ 3 doses | 7 | 5 | 71 | 13 | 35 |
| ≥3 doses | 1 | 0 | 0 | n/a | n/a |
| 4 doses | 1 | 1 | 100 | 95 | 95 |
| Any mRNA | | | | | |
| Overall | 23 | 14 | 61 | 14 | 84 |
| 1 dose | 7 | 4 | 57 | 14 | 51 |
| ≥1 dose | 3 | 0 | 0 | n/a | n/a |
| Primary series/ 2 doses | 9 | 7 | 78 | 22 | 75 |
| ≥2 doses | 1 | 0 | 0 | n/a | n/a |
| 4 doses | 3 | 3 | 100 | 64 | 84 |
| BNT162b2 | | | | | |
| Overall | 11 | 8 | 73 | 14 | 84 |
| 1 dose | 5 | 4 | 80 | 14 | 51 |
| 2 doses | 3 | 1 | 33 | 75 | 75 |
| 3 doses | 3 | 3 | 100 | 64 | 84 |
| Non-mRNA | | | | | |
| Overall | 8 | 6 | 75 | 16 | 38 |
| 1 dose | 4 | 3 | 75 | 16 | 37 |
| 2 doses | 4 | 3 | 75 | 28 | 38 |

mRNA: messenger ribonucleic acid; VE: vaccine effectiveness

Overall, aVE estimates for non-mRNA vaccines were available in 4 different studies, aVE estimates for mRNA vaccines were available in 10 different studies, aVE estimates for any/mixed vaccines were available in 18 different studies, aVE estimates for BNT162b2 vaccines were available in 3 different studies, aVE estimates when vaccine type and manufacturer were not reported were available in 2 different studies.

**eTable 16. Summary of publications where adjusted pre-infection VE against ≥1 PCC (vs. unvaccinated) could be computed, by predominant variant circulating during acute infection (data corresponding to Figure 4) (N=31 studies with 93 VE estimates)**

| **Author, year (Vaccine, dose)** | **Variant** | **Predominant variant period** | **Subgroup** | **Subgroup detail** | **Adjustments** | **VE** | **Lower 95% CI** | **Upper 95% CI** |
| --- | --- | --- | --- | --- | --- | --- | --- | --- |
| Al-Aly, 2022 (BNT162b2, mRNA-1273 or Ad26.COV2.S, Primary series) | Wild-type, Alpha, Delta | Pre-omicron | NA | Risk of incident post-acute sequelae | Adjusted for age, race, area deprivation index, outpatient visits, inpatient visits, and unique medication prescriptions in previous year, routine blood chemistry panels, BMI, history of influenza vaccination, smoking status, comorbidities, systolic blood pressure, diastolic blood pressure, estimated glomerular filtration rate, and hospital system average hospital bed occupancy, total hospital bed capacity, positivity rate of tests, number of tests, and HHS region SARS-CoV-2 variant proportion in the week prior to enrolment, geographic region of VISN, and calendar week of enrolment | 15 | 11 | 18 |

| **Author, year (Vaccine, dose)** | **Variant** | **Omicron period** | **Subgroup** | **Subgroup detail** | **Adjustments** | **VE** | **Lower 95% CI** | **Upper 95% CI** |
| --- | --- | --- | --- | --- | --- | --- | --- | --- |
| Al-Aly, 2022 (BNT162b2, mRNA-1273 or Ad26.COV2.S, Primary series) | Wild-type, Alpha, Delta | Pre-omicron | Setting | Non-hospitalized | Adjusted for age, race, area deprivation index, outpatient visits, inpatient visits, and unique medication prescriptions in previous year, routine blood chemistry panels, BMI, history of influenza vaccination, smoking status, comorbidities, systolic blood pressure, diastolic blood pressure, estimated glomerular filtration rate, and hospital system average hospital bed occupancy, total hospital bed capacity, positivity rate of tests, number of tests, and HHS region SARS-CoV-2 variant proportion in the week prior to enrolment, geographic region of VISN, and calendar week of enrolment | 7 | 4 | 11 |
| Al-Aly, 2022 (BNT162b2, mRNA-1273 or Ad26.COV2.S, Primary series) | Wild-type, Alpha, Delta | Pre-omicron | Setting | Hospitalized | Adjusted for age, race, area deprivation index, outpatient visits, inpatient visits, and unique medication prescriptions in previous year, routine blood chemistry panels, BMI, history of influenza vaccination, smoking status, comorbidities, systolic blood pressure, diastolic blood pressure, estimated glomerular filtration rate, and hospital system average hospital bed occupancy, total hospital bed capacity, positivity rate of tests, number of tests, and HHS region SARS-CoV-2 variant proportion in the week prior to enrolment, geographic region of VISN, and calendar week of enrolment | 12 | 4 | 19 |
| Al-Aly, 2022 (BNT162b2, mRNA-1273 or Ad26.COV2.S, Primary series) | Wild-type, Alpha, Delta | Pre-omicron | Setting | ICU | Adjusted for age, race, area deprivation index, outpatient visits, inpatient visits, and unique medication prescriptions in previous year, routine blood chemistry panels, BMI, history of influenza vaccination, smoking status, comorbidities, systolic blood pressure, diastolic blood pressure, estimated glomerular filtration rate, and hospital system average hospital bed occupancy, total hospital bed capacity, positivity rate of tests, number of tests, and HHS region SARS-CoV-2 variant proportion in the week prior to enrolment, geographic region of VISN, and calendar week of enrolment | 22 | 10 | 33 |
| Antonelli, 2022 (BNT162b2, mRNA-1273, ChAdOx1-S or NR, 2 doses) | Alpha, Delta | Pre-omicron | Number of doses | 2 | Adjusted for age, BMI, sex | 49 | 18 | 68 |
| Antonelli, 2022 (BNT162b2, mRNA-1273, ChAdOx1-S or NR, 1 dose) | Alpha, Delta | Pre-omicron | Number of doses | 1 | Adjusted for age, BMI, sex | -3 | -24 | 15 |
| Ayoubkhani, 2022 (BNT162b2, ChAdOx1-S or Ad26.COV2.S, 2 doses) | Delta | Pre-omicron | Vaccine and long COVID severity | Adenoviral and mRNA; Long COVID of any severity | Adjusted for age, sex, white or non-white ethnicity, country/region of residence, area deprivation quintile group, and self-reported, pre-existing health/disability status and time from infection to follow-up for Long Covid | 41 | 31 | 50 |
| Ayoubkhani, 2022 (BNT162b2, ChAdOx1-S or Ad26.COV2.S, 2 doses) | Delta | Pre-omicron | Vaccine and long COVID severity | Adenoviral and mRNA; activity-limiting Long COVID | Adjusted for age, sex, white or non-white ethnicity, country/region of residence, area deprivation quintile group, and self-reported, pre-existing health/disability status and time from infection to follow-up for Long Covid | 41 | 27 | 52 |
| Ayoubkhani 2022 (BNT162b2 or mRNA-1273, 2 doses) | Delta | Pre-omicron | Vaccine and long COVID severity | mRNA vaccine platform; Long COVID of any severity | Adjusted for age, sex, white or non-white ethnicity, country/region of residence, area deprivation quintile group, and self-reported, pre-existing health/disability status and time from infection to follow-up for Long Covid | 50 | 31 | 63 |
| Ayoubkhani 2022 (BNT162b2 or mRNA-1273, 2 doses) | Delta | Pre-omicron | Vaccine and long COVID severity | mRNA vaccine platform; activity-limiting Long COVID | Adjusted for age, sex, white or non-white ethnicity, country/region of residence, area deprivation quintile group, and self-reported, pre-existing health/disability status and time from infection to follow-up for Long Covid | 50 | 25 | 56 |
| Ayoubkhani 2022 (ChAdOx1-S, 2 doses) | Delta | Pre-omicron | Vaccine and long COVID severity | AstraZeneca; Long COVID of any severity | Adjusted for age, sex, white or non-white ethnicity, country/region of residence, area deprivation quintile group, and self-reported, pre-existing health/disability status and time from infection to follow-up for Long Covid | 38 | 25 | 49 |
| Ayoubkhani 2022 (ChAdOx1-S, 2 doses) | Delta | Pre-omicron | Vaccine and long COVID severity | AstraZeneca; activity limiting Long COVID | Adjusted for age, sex, white or non-white ethnicity, country/region of residence, area deprivation quintile group, and self-reported, pre-existing health/disability status and time from infection to follow-up for Long Covid | 37 | 20 | 51 |
| Ertesvag, 2023 (BNT162b2 or mRNA-1273, ≥1 dose) | Delta | Pre-omicron | Timepoint and population | Vaccination as a predictor of any persisting symptoms 3-months post-Delta infection | Adjusted by female sex, age, reinfection, any symptoms at baseline. | -123 | -904 | 45 |
| Ertesvag, 2023 (BNT162b2 or mRNA-1273, ≥1 dose) | Delta | Pre-omicron | Timepoint and population | Vaccination as a predictor of any persisting symptoms among adolescents (≥16 years) 8-months post-Delta infection | Adjusted for female sex, age, reinfection, and any symptoms at baseline | -98 | -457 | 30 |
| Gallant, 2022 (BNT162b2 or mRNA-1273, ≥2 doses) | Delta | Pre-omicron | NA | NA | Adjusted for age, sex, number of symptoms during acute COVID-19, and number of comorbidities. | -3 | -18 | 10 |
| Ioannou, 2022 (BNT162b2 or mRNA-1273, 2 doses) | Wild-type, Alpha | Pre-omicron | NA | NA | Adjusted for age, sex, race, ethnicity, urban vs rural residence, Charlson Comorbidity Index score, VA Integrated Service Network, time period of infection/pandemic wave, number of primary care, mental health and specialty care encounters in the 2 years before infection | 22 | 10 | 32 |
| Ioannou, 2022 (BNT162b2 or mRNA-1273, 1 dose) | Wild-type, Alpha | Pre-omicron | NA | NA | Adjusted for age, sex, race, ethnicity, urban vs rural residence, Charlson Comorbidity Index score, VA Integrated Service Network, time period of infection/pandemic wave, number of primary care, mental health and specialty care encounters in the 2 years before infection | -3 | -10 | 5 |
| Mohr, 2023 (BNT162b2 or mRNA-1273, 2 doses) | Wild-type, Alpha, Delta | Pre-omicron | Timepoint | >16 weeks since vaccination | Adjusted for age, race and ethnicity, comorbidities, calendar month of diagnosis, and weeks since symptoms started | 28 | 12 | 41 |
| Mohr, 2023 (BNT162b2 or mRNA-1273, 2 doses) | Wild-type, Alpha, Delta | Pre-omicron | Timepoint | ≤16 weeks since vaccination symptoms | Adjusted for age, race and ethnicity, comorbidities, calendar month of diagnosis, and weeks since symptoms started | 33 | 13 | 49 |
| Mohr, 2023 (BNT162b2 or mRNA-1273, 2 doses) | Wild-type, Alpha, Delta | Pre-omicron | Timepoint | 6 weeks since vaccination | Adjusted for age, race and ethnicity, comorbidities, calendar month of diagnosis and weeks since symptoms started | 30 | 16 | 42 |
| Richard, 2023 (BNT162b2, mRNA-1273 or Ad26.COV2.S, Primary series) | Wild-type, Alpha, Delta | Pre-omicron | Timepoint | ≥90 days | Adjusted for Delta variant period, maximum symptom severity during acute illness, days hospitalized for COVID-19, comorbidities, acute respiratory symptoms, age, sex and BMI category | 27 | -14 | 53 |
| Richard, 2023 (BNT162b2, mRNA-1273 or Ad26.COV2.S, Primary series) | Wild-type, Alpha, Delta | Pre-omicron | Timepoint | ≥28 days | Adjusted for Delta variant period, maximum symptom severity during acute illness, days hospitalized for COVID-19, comorbidities, acute respiratory symptoms, age, sex and BMI category | 28 | 4 | 46 |
| Senjam, 2022 (Vaccine NR, 2 doses) | Delta | Pre-omicron | NA | NA | Adjusted for severity of COVID-19, chronic medication use, presence of chronic conditions, self-perceived health status, monthly household income, smoking status, living status, educational level, current occupation, marital status, gender and age | 39 | 4 | 60 |
| Senjam, 2022 (Vaccine NR, 1 dose) | Delta | Pre-omicron | NA | NA | Adjusted for age, gender, educational level, BMI, workplace, blood group, smoking, alcohol consumption, presence of comorbidities, oxygen requirement, severity of COVID-19 disease, overall health status following COVUD-19 illness | 9 | -40 | 41 |
| Tannous, 2023 (mRNA or adenoviral, Primary series) | Wild-type, Alpha, Delta | Pre-omicron | NA | NA | Adjusted for age, sex, race, ethnicity, Charlson Comorbidity Index, COVID-19 illness severity, and Texas state area deprivation index | 42 | 34 | 48 |
| Di Fusco, 2023 (BNT162b2, 3 doses) | Omicron | Omicron | Vaccination and number of symptoms | Boosted vs unvaccinated (≥3 symptoms) | Adjusted for timepoint, vaccination status and interaction of time by vaccination status, participant pre-COVID-19 symptom onset score, sociodemographic characteristics (age, sex, regions, social vulnerability, race/ethnicity, high risk occupations), previously tested positive for COVID-19, severity of acute illness and immunocompromised status | 64 | 13 | 85 |
| Di Fusco, 2023 (BNT162b2, 3 doses) | Omicron | Omicron | Vaccination and number of symptoms | Boosted vs unvaccinated (≥2 symptoms) | Adjusted for timepoint, vaccination status and interaction of time by vaccination status, participant pre-COVID-19 symptom onset score, sociodemographic characteristics (age, sex, regions, social vulnerability, race/ethnicity, high risk occupations), previously tested positive for COVID-19, severity of acute illness and immunocompromised status | 70 | 30 | 87 |
| Di Fusco, 2023 (BNT162b2, 2 doses) | Omicron | Omicron | Vaccination and number of symptoms | Primed vs unvaccinated (≥3 symptoms) | Adjusted for timepoint, vaccination status and interaction of time by vaccination status, participant pre-COVID-19 symptom onset score, sociodemographic characteristics (age, sex, regions, social vulnerability, race/ethnicity, high risk occupations), previously tested positive for COVID-19, severity of acute illness and immunocompromised status | 40 | -34 | 73 |
| Di Fusco, 2023 (BNT162b2, 2 doses) | Omicron | Omicron | Vaccination and number of symptoms | Primed vs unvaccinated (≥2 symptoms) | Adjusted for timepoint, vaccination status and interaction of time by vaccination status, participant pre-COVID-19 symptom onset score, sociodemographic characteristics (age, sex, regions, social vulnerability, race/ethnicity, high risk occupations), previously tested positive for COVID-19, severity of acute illness and immunocompromised status | 36 | -39 | 70 |
| Luo, 2023 (BNT162b2, CoronaVac or NR, ≥3 doses) | Omicron | Omicron | NA | NA | Adjusted for age, gender, BMI, infection days, vaccination status, vaccination after infection, comorbidities, infection days, treatment for the acute stage of illness, and patient's satisfaction of treatment | -11 | -24 | 2 |
| Smits, 2023 (BNT162b2, mRNA-1273, NVX-CoV2373 or Ad26.COV2.S, Booster) | Omicron | Omicron | Vaccination status | Long COVID diagnosis (ICD-10 codes; 28-365 days post-positive COVID-19 test) | Adjusted for vaccination status, comorbidities, sex, race, ethnicity, age, timing of infection, smoking status, influenza vaccination status, inpatient and outpatient encounters in previous two years | 19 | 5 | 31 |
| Smits, 2023 (BNT162b2, mRNA-1273, NVX-CoV2373 or Ad26.COV2.S, Booster) | Omicron | Omicron | Vaccination status | Long-COVID symptoms (28-365 days post-positive COVID-19 test) | Adjusted for vaccination status, comorbidities, sex, race, ethnicity, age, timing of infection, smoking status, influenza vaccination status, inpatient and outpatient encounters in previous two years | 13 | 9 | 17 |
| Smits, 2023 (BNT162b2, mRNA-1273, NVX-CoV2373 or Ad26.COV2.S, Primary series) | Omicron | Omicron | Vaccination status | Long COVID diagnosis (ICD-10 codes; 28–365 days post-positive COVID-19 test) | Adjusted for vaccination status, comorbidities, sex, race, ethnicity, age, timing of infection, smoking status, influenza vaccination status, inpatient and outpatient encounters in previous two years | 14 | 1 | 26 |
| Smits, 2023 (BNT162b2, mRNA-1273, NVX-CoV2373 or Ad26.COV2.S, Primary series) | Omicron | Omicron | Vaccination status | Long-COVID symptoms (28-365 days post-positive COVID-19 test) | Adjusted for vaccination status, comorbidities, sex, race, ethnicity, age, timing of infection, smoking status, influenza vaccination status, inpatient and outpatient encounters in previous two years | 10 | 6 | 13 |
| Tosi, 2023 (BNT162b2, mRNA-1273 or Ad26.COV2.S, Booster) | Omicron | Omicron | Vaccination and patient frailty | Boosted, robust patient | Adjusted for age, BMI, race, ethnicity, smoking, active patient in the past 24 months, and rurality | 26 | 18 | 33 |
| Tosi, 2023 (BNT162b2, mRNA-1273 or Ad26.COV2.S, Booster) | Omicron | Omicron | Vaccination and patient frailty | Boosted, pre-frail patient | Adjusted for age, BMI, race, ethnicity, smoking, active patient in the past 24 months, and rurality | 35 | 28 | 41 |
| Tosi, 2023 (BNT162b2, mRNA-1273 or Ad26.COV2.S, Booster) | Omicron | Omicron | Vaccination and patient frailty | Boosted, frail patient | Adjusted for age, BMI, race, ethnicity, smoking, active patient in the past 24 months, and rurality | 30 | 23 | 36 |
| Tosi, 2023 (BNT162b2, mRNA-1273 or Ad26.COV2.S, Primary series) | Omicron | Omicron | Vaccination and patient frailty | Vaccinated, robust patient | Adjusted for age, BMI, race, ethnicity, smoking, active patient in the past 24 months, and rurality | 14 | 7 | 19 |
| Tosi, 2023 (BNT162b2, mRNA-1273 or Ad26.COV2.S, Primary series) | Omicron | Omicron | Vaccination and patient frailty | Vaccinated, pre-frail patient | Adjusted for age, BMI, race, ethnicity, smoking, active patient in the past 24 months, and rurality | 20 | 13 | 26 |
| Tosi, 2023 (BNT162b2, mRNA-1273 or Ad26.COV2.S, Primary series) | Omicron | Omicron | Vaccination and patient frailty | Vaccinated, frail patient | Adjusted for age, BMI, race, ethnicity, smoking, active patient in the past 24 months, and rurality | 20 | 13 | 27 |
| Antonelli, 2022b (Vaccine NR, ≥1 dose) | Delta, Omicron | Both | Time since vaccination | Vaccinated >6 months prior to infection | Adjusted by age, sex, BMI, index of multiple deprivation, presence of comorbidities and vaccination status | 74 | 68 | 80 |
| Antonelli, 2022b (Vaccine NR, ≥1 dose) | Delta, Omicron | Both | Time since vaccination | Vaccinated 3–6 months prior to infection | Adjusted by age, sex, BMI, index of multiple deprivation, presence of comorbidities and vaccination status | 76 | 68 | 81 |
| Antonelli, 2022b (Vaccine NR, ≥1 dose) | Delta, Omicron | Both | Time since vaccination | Vaccinated <3 months prior to infection | Adjusted by age, sex, BMI, index of multiple deprivation, presence of comorbidities and vaccination status | 50 | 41 | 57 |
| Antonelli, 2022b (Vaccine NR, ≥1 dose) | Delta, Omicron | Both | Age; Time since vaccination | 18–59; vaccinated >6 months prior to infection | Adjusted by age, sex, BMI, index of multiple deprivation, presence of comorbidities and vaccination status | 77 | 68 | 84 |
| Antonelli, 2022b (Vaccine NR, ≥1 dose) | Delta, Omicron | Both | Age; Time since vaccination | 18–59; vaccinated 3–6 months prior to infection | Adjusted by age, sex, BMI, index of multiple deprivation, presence of comorbidities and vaccination status | 67 | 56 | 76 |
| Antonelli, 2022b (Vaccine NR, ≥1 dose) | Delta, Omicron | Both | Age; Time since vaccination | 18–59; vaccinated <3 months prior to infection | Adjusted by age, sex, BMI, index of multiple deprivation, presence of comorbidities and vaccination status | 59 | 47 | 68 |
| Antonelli, 2022b (Vaccine NR, ≥1 dose) | Delta, Omicron | Both | Age; Time since vaccination | ≥60; vaccinated >6 months prior to infection | Adjusted by age, sex, BMI, index of multiple deprivation, presence of comorbidities and vaccination status | 72 | 61 | 80 |
| Antonelli, 2022b (Vaccine NR, ≥1 dose) | Delta, Omicron | Both | Age; Time since vaccination | ≥60; vaccinated 3–6 months prior to infection | Adjusted by age, sex, BMI, index of multiple deprivation, presence of comorbidities and vaccination status | 54 | -3 | 80 |
| Antonelli, 2022b (Vaccine NR, ≥1 dose) | Delta, Omicron | Both | Age; Time since vaccination | ≥60; vaccinated <3 months prior to infection | Adjusted by age, sex, BMI, index of multiple deprivation, presence of comorbidities and vaccination status | 46 | 32 | 57 |
| Azzolini, 2022 (BNT162b2, 3 doses) | Wild-type, Alpha, Delta, Omicron | Both | NA | NA | Adjusted for sex, vaccine dose, allergies and comorbidities | 84 | 16 | 97 |
| Azzolini, 2022 (BNT162b2, 2 doses) | Wild-type, Alpha, Delta, Omicron | Both | NA | NA | Adjusted for sex, vaccine dose, allergies and comorbidities | 75 | 13 | 93 |
| Azzolini, 2022 (BNT162b2, 1 dose) | Wild-type, Alpha, Delta, Omicron | Both | NA | NA | Adjusted for sex, vaccine dose, allergies and comorbidities | 14 | -249 | 79 |
| Brannock, 2023 (BNT162b2, mRNA-1273 or ChAdOx1-S, Primary series) | Delta, Omicron | Both | Methodology - proportional hazards | Model-based cohort IPTW-adjusted HR | Inverse probability of treatment weighting applied | 37 | 31 | 43 |
| Brannock, 2023 (BNT162b2, mRNA-1273 or ChAdOx1-S, Primary series) | Delta, Omicron | Both | Methodology - logistic regression analysis | Model-based cohort IPTW-adjusted OR | Inverse probability of treatment weighting applied | 30 | 25 | 35 |
| Brannock, 2023 (BNT162b2, mRNA-1273 or ChAdOx1-S, Primary series) | Delta, Omicron | Both | Methodology - proportional hazards | Clinic-based cohort IPTW-adjusted HR | Inverse probability of treatment weighting applied | 33 | 21 | 44 |
| Brannock, 2023 (BNT162b2, mRNA-1273 or ChAdOx1-S, Primary series) | Delta, Omicron | Both | Methodology - logistic regression analysis | Clinic-based cohort IPTW-adjusted OR | Inverse probability of treatment weighting applied | 30 | 19 | 40 |
| Català, 2023 (BNT162b2, mRNA-1273, ChAdOx1-S or Ad26.COV2.S, 1 dose) | Delta, Omicron | Both | Cohort | CPRD AURUM | Propensity score overlap weighting for age (5-year age bands for people aged ≥18 years), location, index date, previous observation years, number of previous GP visits, number of previous SARS-CoV-2 PCR tests, regional vaccination, testing, COVID-19 incidence rates | 36 | 26 | 45 |
| Català, 2023 (BNT162b2, mRNA-1273, ChAdOx1-S or Ad26.COV2.S, 1 dose) | Delta, Omicron | Both | Cohort | CPRD AURUM, ≥28 days | Propensity score overlap weighting for age (5-year age bands for people aged ≥18 years), location, index date, previous observation years, number of previous GP visits, number of previous SARS-CoV-2 PCR tests, regional vaccination, testing, COVID-19 incidence rates | 30 | 16 | 42 |
| Català, 2023 (BNT162b2, mRNA-1273, ChAdOx1-S or Ad26.COV2.S, 1 dose) | Delta, Omicron | Both | Cohort | CPRD ARUM, GP diagnosis | Propensity score overlap weighting for age (5-year age bands for people aged ≥18 years), location, index date, previous observation years, number of previous GP visits, number of previous SARS-CoV-2 PCR tests, regional vaccination, testing, COVID-19 incidence rates | 36 | 23 | 46 |
| Català, 2023 (BNT162b2, mRNA-1273, ChAdOx1-S or Ad26.COV2.S, 1 dose) | Delta, Omicron | Both | Cohort | CORIVA | Propensity score overlap weighting for age (5-year age bands for people aged ≥18 years), location, index date, previous observation years, number of previous GP visits, number of previous SARS-CoV-2 PCR tests, regional vaccination, testing | 38 | 22 | 51 |
| Català, 2023 (BNT162b2, mRNA-1273, ChAdOx1-S or Ad26.COV2.S, 1 dose) | Delta, Omicron | Both | Cohort | CORIVA, ≥28 days | Propensity score overlap weighting for age (5-year age bands for people aged ≥18 years), location, index date, previous observation years, number of previous GP visits, number of previous SARS-CoV-2 PCR tests, regional vaccination, testing | 38 | 23 | 50 |
| Català, 2023 (BNT162b2, mRNA-1273, ChAdOx1-S or Ad26.COV2.S, 1 dose) | Delta, Omicron | Both | Cohort | CPRD GOLD | Propensity score overlap weighting for age (5-year age bands for people aged ≥18 years), location, index date, previous observation years, number of previous GP visits, number of previous SARS-CoV-2 PCR tests, regional vaccination, testing, COVID-19 incidence rates | 45 | 35 | 54 |
| Català, 2023 (BNT162b2, mRNA-1273, ChAdOx1-S or Ad26.COV2.S, 1 dose) | Delta, Omicron | Both | Cohort | CPRD GOLD, ≥28 days | Propensity score overlap weighting for age (5-year age bands for people aged ≥18 years), location, index date, previous observation years, number of previous GP visits, number of previous SARS-CoV-2 PCR tests, regional vaccination, testing, COVID-19 incidence rates | 34 | 17 | 47 |
| Català, 2023 (BNT162b2, mRNA-1273, ChAdOx1-S or Ad26.COV2.S, 1 dose) | Delta, Omicron | Both | Cohort | SIDIAP | Propensity score overlap weighting for age (5-year age bands for people aged ≥18 years), location, index date, previous observation years, number of previous GP visits, number of previous SARS-CoV-2 PCR tests, regional vaccination, testing | 16 | 7 | 25 |
| Català, 2023 (BNT162b2, mRNA-1273, ChAdOx1-S or Ad26.COV2.S, 1 dose) | Delta, Omicron | Both | Cohort | SIDIAP, ≥28 days | Propensity score overlap weighting for age (5-year age bands for people aged ≥18 years), location, index date, previous observation years, number of previous GP visits, number of previous SARS-CoV-2 PCR tests, regional vaccination, testing | 12 | 2 | 21 |
| Català, 2023 (BNT162b2, mRNA-1273, ChAdOx1-S or Ad26.COV2.S, 1 dose) | Delta, Omicron | Both | Cohort | SIDIAP, GP diagnosis | Propensity score overlap weighting for age (5-year age bands for people aged ≥18 years), location, index date, previous observation years, number of previous GP visits, number of previous SARS-CoV-2 PCR tests, regional vaccination, testing | 40 | 33 | 48 |
| Català, 2023 (BNT162b2, 1 dose) | Delta, Omicron | Both | Vaccination group | CPRD AURUM (Pfizer vaccine) | Propensity score overlap weighting for age (5-year age bands for people aged ≥18 years), location, index date, previous observation years, number of previous GP visits, number of previous SARS-CoV-2 PCR tests, regional vaccination, testing, COVID-19 incidence rates | 47 | 38 | 55 |
| Català, 2023 (BNT162b2, 1 dose) | Delta, Omicron | Both | Vaccination group | CORIVA (Pfizer vaccine) | Propensity score overlap weighting for age (5-year age bands for people aged ≥18 years), location, index date, previous observation years, number of previous GP visits, number of previous SARS-CoV-2 PCR tests, regional vaccination, testing | 37 | 20 | 50 |
| Català, 2023 (BNT162b2, 1 dose) | Delta, Omicron | Both | Vaccination group | CPRD GOLD (Pfizer vaccine) | Propensity score overlap weighting for age (5-year age bands for people aged ≥18 years), location, index date, previous observation years, number of previous GP visits, number of previous SARS-CoV-2 PCR tests, regional vaccination, testing, COVID-19 incidence rates | 51 | 37 | 61 |
| Català, 2023 (BNT162b2, 1 dose) | Delta, Omicron | Both | Vaccination group | SIDIAP (Pfizer vaccine) | Propensity score overlap weighting for age (5-year age bands for people aged ≥18 years), location, index date, previous observation years, number of previous GP visits, number of previous SARS-CoV-2 PCR tests, regional vaccination, testing | 14 | 5 | 23 |
| Català, 2023 (ChAdOx1-S, 1 dose) | Delta, Omicron | Both | Vaccination group | CPRD AURUM (AstraZeneca vaccine) | Propensity score overlap weighting for age (5-year age bands for people aged ≥18 years), location, index date, previous observation years, number of previous GP visits, number of previous SARS-CoV-2 PCR tests, regional vaccination, testing, COVID-19 incidence rates | 29 | 21 | 37 |
| Català, 2023 (ChAdOx1-S, 1 dose) | Delta, Omicron | Both | Vaccination group | CPRD GOLD (AstraZeneca vaccine) | Propensity score overlap weighting for age (5-year age bands for people aged ≥18 years), location, index date, previous observation years, number of previous GP visits, number of previous SARS-CoV-2 PCR tests, regional vaccination, testing, COVID-19 incidence rates | 37 | 23 | 48 |
| Català, 2023 (ChAdOx1-S, 1 dose) | Delta, Omicron | Both | Vaccination group | SIDIAP (AstraZeneca vaccine) | Propensity score overlap weighting for age (5-year age bands for people aged ≥18 years), location, index date, previous observation years, number of previous GP visits, number of previous SARS-CoV-2 PCR tests, regional vaccination, testing | 16 | 4 | 27 |
| Cortellini, 2023 (BNT162b2, mRNA-1273, ChAdOx1-S or Ad26.COV2.S, 3 doses) | Alpha, Delta, Omicron | Both | NA | NA | Inverse probability of treatment weighting applied | 53 | -18 | 82 |
| Cortellini, 2023 (BNT162b2, mRNA-1273, ChAdOx1-S or Ad26.COV2.S, 2 doses) | Alpha, Delta, Omicron | Both | NA | NA | Inverse probability of treatment weighting applied | 59 | 30 | 76 |
| de Arriba Fernández, 2023 (BNT162b2, mRNA-1273, ChAdOx1-S or Ad26.COV2.S, 1 dose) | Delta, Omicron | Both | NA | NA | Adjusted for age, sex, comorbidity history, body mass index, date of first, second and booster doses of a COVID-19 vaccine, type of COVID-19 vaccine, death, immunosuppressive treatment, diagnosis of persistent COVID-19, and diagnostic test results | 82 | 71 | 89 |
| Durstenfeld, 2022 (Vaccine NR, ≥1 dose) | Wild-type, Alpha, Delta, Omicron | Both | NA | NA | Adjusted for age, COVID-19 wave, number of initial symptoms, sex, myocardial infarction, stroke, atrial fibrillation, sleep apnoea, COPD or asthma, and immunodeficiency, plus receipt of COVID-19 vaccination before or after COVID-19 diagnosis, ethnicity, subjective socioeconomic status, highest level of education, primary employment, pre-COVID depressive symptoms, anxiety symptoms, and financial security | 19 | -49 | 56 |
| Elmazny, 2023 (BNT162b2, Ad26.COV2.S, ChAdOx1-S, Sputnik V, CoronaVac, BBIBP-CorV, Primary series) | WT, Alpha, Delta, Omicron | Both | NA | NA | Adjusted for age, sex, and COVID-19 severity | 40 | 20 | 55 |
| Herman, 2022 (inactivated viral vaccine, 2 doses) | Delta, Omicron | Both | NA | NA | Adjusted by shortness of breath | 26 | -8 | 49 |
| Kahlert, 2023 (BNT162b2 or mRNA-1273, ≥1 dose) | Wild-type, Alpha, Delta, Omicron | Both | NA | NA | Adjusted for age, sex, BMI, ethnicity, children in household, comorbidity, medication, smoker status, alcohol intake, medical profession, COVID-19 patient contact, ICU work, full-time work, SARS-CoV-2 variant | -27 | -94 | 18 |
| Marra, 2023 (BNT162b2, CoronaVac, ChAdOx1-S or Ad26.COV2.S, 4 doses) | Wild-type, Gamma, Delta, Omicron | Both | NA | NA | Adjusted for sex, age, BMI, any comorbidity, the number of SARS-CoV-2 infections, and SARS-CoV-2 variant | 95 | 81 | 99 |
| Marra, 2023 (BNT162b2, CoronaVac, ChAdOx1-S or Ad26.COV2.S, 3 doses) | Wild-type, Gamma, Delta, Omicron | Both | NA | NA | Adjusted for sex, age, BMI, any comorbidity, the number of SARS-CoV-2 infections, and SARS-CoV-2 variant | 37 | -2 | 61 |
| Marra, 2023 (BNT162b2, CoronaVac, ChAdOx1-S or Ad26.COV2.S, 2 doses) | Wild-type, Gamma, Delta, Omicron | Both | NA | NA | Adjusted for sex, age, BMI, any comorbidity, the number of SARS-CoV-2 infections, and SARS-CoV-2 variant | -17 | -76 | 21 |
| Marra, 2023 (BNT162b2, CoronaVac, ChAdOx1-S or Ad26.COV2.S, 1 dose) | Wild-type, Gamma, Delta, Omicron | Both | NA | NA | Adjusted for sex, age, BMI, any comorbidity, the number of SARS-CoV-2 infections, and SARS-CoV-2 variant | 9 | -39 | 40 |
| Parodi, 2023 (mRNA or inactivated viral, 2 doses) | Gamma, Delta, Omicron | Both | NA | NA | Adjusted for age, sex, hypertension and smoking | 48 | 31 | 60 |
| Patel, 2023 (BNT162b2, mRNA-1273 or Ad26.COV2.S, Primary series) | Delta, Omicron | Both | PASC definition | WHO; ≥90 days post-infection | Adjusted for age, sex, race, comorbidity count and use of any one of the following medications: anti-CD20 monoclonal antibodies, methotrexate, mycophenolate or glucocorticoids. | 90 | 78 | 96 |
| Patel, 2023 (BNT162b2, mRNA-1273 or Ad26.COV2.S, Primary series) | Delta, Omicron | Both | PASC definition | CDC; ≥28 days | Adjusted for age, sex, race, comorbidity count and use of any one of the following medications: anti-CD20 monoclonal antibodies, methotrexate, mycophenolate or glucocorticoids. | 51 | 17 | 71 |
| Perlis, 2022 (Ad26.COV2.S, 2 doses) | Alpha, Delta, Omicron | Both | NA | NA | Adjusted for age, gender, race and ethnicity, educational level, urbanicity, region and predominant variant | 28 | 14 | 40 |
| Perlis, 2022 (Ad26.COV2.S, 1 dose) | Alpha, Delta, Omicron | Both | NA | NA | Adjusted for age, gender, race and ethnicity, educational level, urbanicity, region and predominant variant | 7 | -25 | 31 |
| Ranucci, 2023 (BNT162b2, mRNA-1273, ChAdOx1-S or Ad26.COV2.S or NR, 2 doses) | Wild-type, Alpha, Delta, Omicron | Both | Symptom | Persistent major physical symptoms | Adjusted for age, severity of disease | 69 | 40 | 84 |
| Ranucci, 2023 (BNT162b2, mRNA-1273, ChAdOx1-S or Ad26.COV2.S or NR, 2 doses) | Wild-type, Alpha, Delta, Omicron | Both | Symptom | Persistent major neuropsychological symptoms | Adjusted for previous neurological symptoms | 80 | 57 | 90 |
| Xue, 2023 (BNT162b2 or mRNA-1273, 1 dose) | Alpha, Gamma, Delta, Omicron | Both | NA | NA | Adjusted for age, gender, ethnicity, smoking status, alcohol consumption, educational level, since first vaccination, martial status, living area, level of physical activity, weight status, comorbidities. | -13 | -34 | 5 |
| Abu Hamdh, 2023 (BNT162b2, mRNA-1273, Sputnik V, Sputnik Light or ChAdOx1-S, ≥1 dose) | NR | NR | NA | NA | Adjusted for sex, age, comorbidities, hospital admission, dyspnoea, previous COVID-19 infection, fatigue | 86 | 76 | 91 |

BMI: Body mass index; CDC: Centers for Disease Control and Prevention; CI: Confidence interval; CPRD: Clinical Practice Research Datalink; GP: General Practitioner; HHS: Health and Human Services; HR: Hazard ratio; ICD-10: International Classification of Diseases- 10; ICU: Intensive care units; IPTW: Inverse probability of treatment weighting; mRNA: Messenger ribonucleic acid; NA: Not applicable; NR: Not reported; OR: Odds ratio; PASC: Post-acute sequelae of COVID-19; PCR: Polymerase Chain Reaction; VE: Vaccine effectiveness; VISN: Veterans Integrated Services Network; WHO: World Health Organization.

**eTable 17. Summary of publications where adjusted pre-infection VE against ≥1 PCC (vs. unvaccinated) could be computed, sorted by PCC data source (N=31 studies with 93 VE estimates)**

| **Author, year (Vaccine brand, dose)** | **Follow-up time** | **Variant** | **Subgroup** | **Subgroup details** | **Adjustments** | | **VE** | | **Lower 95% CI** | **Upper 95% CI** | |
| --- | --- | --- | --- | --- | --- | --- | --- | --- | --- | --- | --- |
| **Medical codes/claims and patient-reported** | | | | | | | | | | | |
| Luo, 2023 (BNT162b2, CoronaVac or NR; ≥3 doses) | 6-12 months post acute infection | Omicron | NA | NA | Adjusted for age, gender, BMI, infection days, vaccination status, vaccination after infection, comorbidities, infection days, treatment for the acute stage of illness, and patient's satisfaction of treatment | -11 | | -24 | | | 2 |
| **Medical codes/ claims** | | | | | | | | | | | |
| Marra, 2023 (BNT162b2, CoronaVac, ChAdOx1-S or Ad26.COV2.S; 4 doses) | 6 months post acute infection | Wild-type, Gamma, Delta, Omicron | NA | NA | Adjusted for sex, age, BMI, any comorbidity, the number of SARS-CoV-2 infections, and SARS-CoV-2 variant | | 95 | | 81 | 99 | |
| Smits, 2023 (BNT162b2, mRNA-1273, NVX-CoV2373 or Ad26.COV2.S; booster) | 28–365 days post-positive COVID-19 test | Omicron | Vaccination status | Long COVID diagnosis (ICD-10 codes; 28-365 days post-positive COVID-19 test) | Adjusted for vaccination status, comorbidities, sex, race, ethnicity, age, timing of infection, smoking status, influenza vaccination status, inpatient and outpatient encounters in previous two years | | 19 | | 5 | 31 | |

| **Author, year (Vaccine brand, dose)** | **Follow-up time** | **Variant** | **Subgroup** | **Subgroup details** | **Adjustments** | **VE** | **Lower 95% CI** | **Upper 95% CI** |
| --- | --- | --- | --- | --- | --- | --- | --- | --- |
| Smits, 2023 (BNT162b2, mRNA-1273, NVX-CoV2373 or Ad26.COV2.S; booster) | 28–365 days post-positive COVID-19 test | Omicron | Vaccination status | Long-COVID symptoms (28-365 days post-positive COVID-19 test) | Adjusted for vaccination status, comorbidities, sex, race, ethnicity, age, timing of infection, smoking status, influenza vaccination status, inpatient and outpatient encounters in previous two years | 13 | 9 | 17 |
| Tosi, 2023 (BNT162b2, mRNA-1273 or Ad26.COV2.S; booster) | NR (specified survival at ≥30 days after acute infection) | Omicron | Vaccine, patient frailty | Boosted, robust patient | Adjusted for age, BMI, race, ethnicity, smoking, active patient in the past 24 months, and rurality | 26 | 18 | 33 |
| Tosi, 2023 (BNT162b2, mRNA-1273 or Ad26.COV2.S; booster) | NR (specified survival at ≥30 days after acute infection) | Omicron | Vaccine, patient frailty | Boosted, pre-frail patient | Adjusted for age, BMI, race, ethnicity, smoking, active patient in the past 24 months, and rurality | 35 | 28 | 41 |
| Tosi, 2023 (BNT162b2, mRNA-1273 or Ad26.COV2.S; booster) | NR (specified survival at ≥30 days after acute infection) | Omicron | Vaccine, patient frailty | Boosted, frail patient | Adjusted for age, BMI, race, ethnicity, smoking, active patient in the past 24 months, and rurality | 30 | 23 | 36 |
| Marra, 2023 (BNT162b2, CoronaVac, ChAdOx1-S or Ad26.COV2.S; 3 doses) | 6 months post acute infection | Wild-type, Gamma, Delta, Omicron | NA | NA | Adjusted for sex, age, BMI, any comorbidity, the number of SARS-CoV-2 infections, and SARS-CoV-2 variant | 37 | -2 | 61 |
| Cortellini, 2023 (BNT162b2, mRNA-1273, ChAdOx1-S or Ad26.COV2.S; 3 doses) | 4–12 weeks post acute infection (Unvaccinated patients were assessed at a median of 40 days (IQR 25–67) and doubly vaccinated patients 38 days (IQR: 23–68)) | Alpha, Delta, Omicron | NA | NA | Inverse probability of treatment weighting applied | 53 | -18 | 82 |
| Al-Aly, 2022 (BNT162b2, mRNA-1273 or Ad26.COV2.S; primary series) | 6 months | Wild-type, Alpha, Delta | NA | Risk of incident post-acute sequelae | Adjusted for age, race, area deprivation index, outpatient visits, inpatient visits, and unique medication prescriptions in previous year, routine blood chemistry panels, BMI, history of influenza vaccination, smoking status, comorbidities, systolic blood pressure, diastolic blood pressure, estimated glomerular filtration rate, and hospital system average hospital bed occupancy, total hospital bed capacity, positivity rate of tests, number of tests, and HHS region SARS-CoV-2 variant proportion in the week prior to enrollment, geographic region of VISN, and calendar week of enrollment | 15 | 11 | 18 |
| Al-Aly, 2022 (BNT162b2, mRNA-1273 or Ad26.COV2.S; primary series) | 6 months | Wild-type, Alpha, Delta | Setting | Non-hospitalized | Adjusted for age, race, area deprivation index, outpatient visits, inpatient visits, and unique medication prescriptions in previous year, routine blood chemistry panels, BMI, history of influenza vaccination, smoking status, comorbidities, systolic blood pressure, diastolic blood pressure, estimated glomerular filtration rate, and hospital system average hospital bed occupancy, total hospital bed capacity, positivity rate of tests, number of tests, and HHS region SARS-CoV-2 variant proportion in the week prior to enrollment, geographic region of VISN, and calendar week of enrollment | 7 | 4 | 11 |
| Al-Aly, 2022 (BNT162b2, mRNA-1273 or Ad26.COV2.S; primary series) | 6 months | Wild-type, Alpha, Delta | Setting | Hospitalized | Adjusted for age, race, area deprivation index, outpatient visits, inpatient visits, and unique medication prescriptions in previous year, routine blood chemistry panels, BMI, history of influenza vaccination, smoking status, comorbidities, systolic blood pressure, diastolic blood pressure, estimated glomerular filtration rate, and hospital system average hospital bed occupancy, total hospital bed capacity, positivity rate of tests, number of tests, and HHS region SARS-CoV-2 variant proportion in the week prior to enrollment, geographic region of VISN, and calendar week of enrollment | 12 | 4 | 19 |
| Al-Aly, 2022 (BNT162b2, mRNA-1273 or Ad26.COV2.S; primary series) | 6 months | Wild-type, Alpha, Delta | Setting | ICU | Adjusted for age, race, area deprivation index, outpatient visits, inpatient visits, and unique medication prescriptions in previous year, routine blood chemistry panels, BMI, history of influenza vaccination, smoking status, comorbidities, systolic blood pressure, diastolic blood pressure, estimated glomerular filtration rate, and hospital system average hospital bed occupancy, total hospital bed capacity, positivity rate of tests, number of tests, and HHS region SARS-CoV-2 variant proportion in the week prior to enrollment, geographic region of VISN, and calendar week of enrollment | 22 | 10 | 33 |
| Brannock, 2023 (BNT162b2, mRNA-1273 or ChAdOx1-S; primary series) | >45 days post COVID-19 infection (minimum of 164 days) | Delta, Omicron | Methodology - proportional hazards | Model-based cohort IPTW-adjusted HR | Inverse probability of treatment weighting applied | 37 | 31 | 43 |
| Brannock, 2023 (BNT162b2, mRNA-1273 or ChAdOx1-S; primary series) | >45 days post COVID-19 infection (minimum of 164 days) | Delta, Omicron | Methodology - logistic regression anlayses | Model-based cohort IPTW-adjusted OR | Inverse probability of treatment weighting applied | 30 | 25 | 35 |
| Brannock, 2023 (BNT162b2, mRNA-1273 or ChAdOx1-S; primary series) | >45 days post COVID-19 infection (minimum of 164 days) | Delta, Omicron | Methodology - proportional hazards | Clinic-based cohort IPTW-adjusted HR | Inverse probability of treatment weighting applied | 33 | 21 | 44 |
| Brannock, 2023 (BNT162b2, mRNA-1273 or ChAdOx1-S; primary series) | >45 days post COVID-19 infection (minimum of 164 days) | Delta, Omicron | Methodology - logistic regression anlayses | Clinic-based cohort IPTW-adjusted OR | Inverse probability of treatment weighting applied | 30 | 19 | 40 |
| Smits, 2023 (BNT162b2, mRNA-1273, NVX-CoV2373 or Ad26.COV2.S; primary series) | 28–365 days post-positive COVID-19 test | Omicron | Vaccination status | Long COVID diagnosis (ICD-10 codes; 28–365 days post-positive COVID-19 test) | Adjusted for vaccination status, comorbidities, sex, race, ethnicity, age, timing of infection, smoking status, influenza vaccination status, inpatient and outpatient encounters in previous two years | 14 | 1 | 26 |
| Smits, 2023 (BNT162b2, mRNA-1273, NVX-CoV2373 or Ad26.COV2.S; primary series) | 28–365 days post-positive COVID-19 test | Omicron | Vaccination status | Long-COVID symptoms (28-365 days post-positive COVID-19 test) | Adjusted for vaccination status, comorbidities, sex, race, ethnicity, age, timing of infection, smoking status, influenza vaccination status, inpatient and outpatient encounters in previous two years | 10 | 6 | 13 |
| Tannous, 2023 (mRNA or adenoviral; primary series) | >28 days post acute infection | Wild-type, Alpha, Delta | Vaccination status | 2 doses mRNA or 1 dose Ad26.COV2.S | Adjusted for age, sex, race, ethnicity, Charlson Comorbidity Index, COVID-19 illness severity, and Texas state area deprivation index | 42 | 34 | 48 |
| Tosi, 2023 (BNT162b2, mRNA-1273 or Ad26.COV2.S; primary series) | NR (specified survival ≥30 days after acute infection) | Omicron | Vaccine, patient frailty | Vaccinated, robust patient | Adjusted for age, BMI, race, ethnicity, smoking, active patient in the past 24 months, and rurality | 14 | 7 | 19 |
| Tosi, 2023 (BNT162b2, mRNA-1273 or Ad26.COV2.S; primary series) | NR (specified survival at ≥30 days after acute infection) | Omicron | Vaccine, patient frailty | Vaccinated, pre-frail patient | Adjusted for age, BMI, race, ethnicity, smoking, active patient in the past 24 months, and rurality | 20 | 13 | 26 |
| Tosi, 2023 (BNT162b2, mRNA-1273 or Ad26.COV2.S; primary series) | NR (specified survival at ≥30 days after acute infection) | Omicron | Vaccine, patient frailty | Vaccinated, frail patient | Adjusted for age, BMI, race, ethnicity, smoking, active patient in the past 24 months, and rurality | 20 | 13 | 27 |
| Ioannou, 2022 (BNT162b2 or mRNA-1273; 2 doses) | Mean: 13.5 months (range: 8–23) | Wild-type, Alpha | Vaccination status and number of doses | 2 doses of mRNA vaccine vs nonvaccinated | Adjusted for age, sex, race, ethnicity, urban vs rural residence, Charlson Comorbidity Index score, VA Integrated Service Network, time period of infection/pandemic wave, number of primary care, mental health and specialty care encounters in the 2 years before infection | 22 | 10 | 32 |
| Marra, 2023 (BNT162b2, CoronaVac, ChAdOx1-S or Ad26.COV2.S; 2 doses) | 6 months post acute infection | Wild-type, Gamma, Delta, Omicron | NA | NA | Adjusted for sex, age, BMI, any comorbidity, the number of SARS-CoV-2 infections, and SARS-CoV-2 variant | -17 | -76 | 21 |
| Cortellini, 2023 (BNT162b2, mRNA-1273, ChAdOx1-S or Ad26.COV2.S; 2 doses) | 4–12 weeks post acute infection (Unvaccinated patients were assessed at a median of 40 days (IQR 25–67) and doubly vaccinated patients 38 days (IQR: 23–68)) | Alpha, Delta, Omicron | NA | NA | Inverse probability of treatment weighting applied | 59 | 30 | 76 |
| Abu Hamdh, 2023 (BNT162b2, mRNA-1273, Sputnik V, Sputnik Light or ChAdOx1-S; ≥1 dose) | 90 days | NR | NA | NA | Adjusted for sex, age, comorbidities, hospital admission, dyspnea, previous COVID-19 infection, fatigue | 86 | 76 | 91 |
| Ioannou, 2022 (BNT162b2 or mRNA-1273; 1 dose) | Mean: 13.5 months (range: 8–23) | Wild-type, Alpha | Vaccination status and number of doses | 1 dose of mRNA vaccine vs nonvaccinated | Adjusted for age, sex, race, ethnicity, urban vs rural residence, Charlson Comorbidity Index score, VA Integrated Service Network, time period of infection/pandemic wave, number of primary care, mental health and specialty care encounters in the 2 years before infection | -3 | -10 | 5 |
| Marra, 2023 (BNT162b2, CoronaVac, ChAdOx1-S or Ad26.COV2.S; 1 dose) | 6 months post acute infection | Wild-type, Gamma, Delta, Omicron | NA | NA | Adjusted for sex, age, BMI, any comorbidity, the number of SARS-CoV-2 infections, and SARS-CoV-2 variant | 9 | -39 | 40 |
| Català, 2023 (BNT162b2, mRNA-1273, ChAdOx1-S or Ad26.COV2.S; 1 dose) | 90–365 days post-positive test or diagnosis of COVID-19 | Delta, Omicron | Cohort | CPRD AURUM | Propensity score overlap weighting for age (5-year age bands for people aged ≥18 years), location, index date, previous observation years, number of previous GP visits, number of previous SARS-CoV-2 PCR tests, regional vaccination, testing, COVID-19 incidence rates | 36 | 26 | 45 |
| Català, 2023 (BNT162b2, mRNA-1273, ChAdOx1-S or Ad26.COV2.S; 1 dose) | 90–365 days post-positive test or diagnosis of COVID-19 | Delta, Omicron | Cohort | CPRD AURUM, GP diagnosis | Propensity score overlap weighting for age (5-year age bands for people aged ≥18 years), location, index date, previous observation years, number of previous GP visits, number of previous SARS-CoV-2 PCR tests, regional vaccination, testing, COVID-19 incidence rates | 36 | 23 | 46 |
| Català, 2023 (BNT162b2, mRNA-1273, ChAdOx1-S or Ad26.COV2.S; 1 dose) | 90–365 days post-positive test or diagnosis of COVID-19 | Delta, Omicron | Cohort | CORIVA | Propensity score overlap weighting for age (5-year age bands for people aged ≥18 years), location, index date, previous observation years, number of previous GP visits, number of previous SARS-CoV-2 PCR tests, regional vaccination, testing | 38 | 22 | 51 |
| Català, 2023 (BNT162b2, mRNA-1273, ChAdOx1-S or Ad26.COV2.S; 1 dose) | 90–365 days post-positive test or diagnosis of COVID-19 | Delta, Omicron | Cohort | CPRD GOLD | Propensity score overlap weighting for age (5-year age bands for people aged ≥18 years), location, index date, previous observation years, number of previous GP visits, number of previous SARS-CoV-2 PCR tests, regional vaccination, testing, COVID-19 incidence rates | 45 | 35 | 54 |
| Català, 2023 (BNT162b2, mRNA-1273, ChAdOx1-S or Ad26.COV2.S; 1 dose) | 90–365 days post-positive test or diagnosis of COVID-19 | Delta, Omicron | Cohort | SIDIAP | Propensity score overlap weighting for age (5-year age bands for people aged ≥18 years), location, index date, previous observation years, number of previous GP visits, number of previous SARS-CoV-2 PCR tests, regional vaccination, testing | 16 | 7 | 25 |
| Català, 2023 (BNT162b2, mRNA-1273, ChAdOx1-S or Ad26.COV2.S; 1 dose) | 90–365 days post-positive test or diagnosis of COVID-19 | Delta, Omicron | Cohort | SIDIAP, GP diagnosis | Propensity score overlap weighting for age (5-year age bands for people aged ≥18 years), location, index date, previous observation years, number of previous GP visits, number of previous SARS-CoV-2 PCR tests, regional vaccination, testing | 40 | 33 | 48 |
| Català, 2023 (BNT162b2; 1 dose) | 90–365 days post-positive test or diagnosis of COVID-19 | Delta, Omicron | Cohort | CPRD AURUM (Pfizer vaccine) | Propensity score overlap weighting for age (5-year age bands for people aged ≥18 years), location, index date, previous observation years, number of previous GP visits, number of previous SARS-CoV-2 PCR tests, regional vaccination, testing, COVID-19 incidence rates | 47 | 38 | 55 |
| Català, 2023 (BNT162b2; 1 dose) | 90–365 days post-positive test or diagnosis of COVID-19 | Delta, Omicron | Cohort | CORIVA (Pfizer vaccine) | Propensity score overlap weighting for age (5-year age bands for people aged ≥18 years), location, index date, previous observation years, number of previous GP visits, number of previous SARS-CoV-2 PCR tests, regional vaccination, testing | 37 | 20 | 50 |
| Català, 2023 (BNT162b2; 1 dose) | 90–365 days post-positive test or diagnosis of COVID-19 | Delta, Omicron | Cohort | CPRD GOLD (Pfizer vaccine) | Propensity score overlap weighting for age (5-year age bands for people aged ≥18 years), location, index date, previous observation years, number of previous GP visits, number of previous SARS-CoV-2 PCR tests, regional vaccination, testing, COVID-19 incidence rates | 51 | 37 | 61 |
| Català, 2023 (BNT162b2; 1 dose) | 90–365 days post-positive test or diagnosis of COVID-19 | Delta, Omicron | Cohort | SIDIAP (Pfizer vaccine) | Propensity score overlap weighting for age (5-year age bands for people aged ≥18 years), location, index date, previous observation years, number of previous GP visits, number of previous SARS-CoV-2 PCR tests, regional vaccination, testing | 14 | 5 | 23 |
| Català, 2023 (ChAdOx1-S; 1 dose) | 90–365 days post-positive test or diagnosis of COVID-19 | Delta, Omicron | Cohort | CPRD AURUM (Astrazeneca vaccine) | Propensity score overlap weighting for age (5-year age bands for people aged ≥18 years), location, index date, previous observation years, number of previous GP visits, number of previous SARS-CoV-2 PCR tests, regional vaccination, testing, COVID-19 incidence rates | 29 | 21 | 37 |
| Català, 2023 (ChAdOx1-S; 1 dose) | 90–365 days post-positive test or diagnosis of COVID-19 | Delta, Omicron | Cohort | CPRD GOLD (Astrazeneca vaccine) | Propensity score overlap weighting for age (5-year age bands for people aged ≥18 years), location, index date, previous observation years, number of previous GP visits, number of previous SARS-CoV-2 PCR tests, regional vaccination, testing, COVID-19 incidence rates | 37 | 23 | 48 |
| Català, 2023 (ChAdOx1-S; 1 dose) | 90–365 days post-positive test or diagnosis of COVID-19 | Delta, Omicron | Cohort | SIDIAP (Astrazeneca vaccine) | Propensity score overlap weighting for age (5-year age bands for people aged ≥18 years), location, index date, previous observation years, number of previous GP visits, number of previous SARS-CoV-2 PCR tests, regional vaccination, testing | 16 | 4 | 27 |
| Català, 2023 (BNT162b2, mRNA-1273, ChAdOx1-S or Ad26.COV2.S; 1 dose) | 28-365 days | Delta, Omicron | Cohort | CPRD AURUM, ≥28 days | Propensity score overlap weighting for age (5-year age bands for people aged ≥18 years), location, index date, previous observation years, number of previous GP visits, number of previous SARS-CoV-2 PCR tests, regional vaccination, testing, COVID-19 incidence rates | 30 | 16 | 42 |
| Català, 2023 (BNT162b2, mRNA-1273, ChAdOx1-S or Ad26.COV2.S; 1 dose) | 28-365 days | Delta, Omicron | Cohort | CORIVA, ≥28 days | Propensity score overlap weighting for age (5-year age bands for people aged ≥18 years), location, index date, previous observation years, number of previous GP visits, number of previous SARS-CoV-2 PCR tests, regional vaccination, testing | 38 | 23 | 50 |
| Català, 2023 (BNT162b2, mRNA-1273, ChAdOx1-S or Ad26.COV2.S; 1 dose) | 28-365 days | Delta, Omicron | Cohort | CPRD GOLD, ≥28 days | Propensity score overlap weighting for age (5-year age bands for people aged ≥18 years), location, index date, previous observation years, number of previous GP visits, number of previous SARS-CoV-2 PCR tests, regional vaccination, testing, COVID-19 incidence rates | 34 | 17 | 47 |
| Català, 2023 (BNT162b2, mRNA-1273, ChAdOx1-S or Ad26.COV2.S; 1 dose) | 28-365 days | Delta, Omicron | Cohort | SIDIAP, ≥28 days | Propensity score overlap weighting for age (5-year age bands for people aged ≥18 years), location, index date, previous observation years, number of previous GP visits, number of previous SARS-CoV-2 PCR tests, regional vaccination, testing | 12 | 2 | 21 |
| de Arriba Fernández, 2023 (BNT162b2, mRNA-1273, ChAdOx1-S or Ad26.COV2.S; 1 dose) | NR | Delta, Omicron | Vaccination status | Incomplete | Adjusted for age, sex, comorbidity history, body mass index, date of first, second and booster doses of a COVID-19 vaccine, type of COVID-19 vaccine, death, immunosuppressive treatment, diagnosis of persistent COVID-19, and diagnostic test results | 82 | 71 | 89 |
| **Patient-reported** | | | | | | | | |
| Di Fusco, 2023 (BNT162b2; 3 doses) | 6 months | Omicron | Vaccination status | Boosted vs unvaccinated (≥3 symptoms) | Adjusted for timepoint, vaccination status and interaction of time by vaccination status, participant pre-COVID-19 symptom onset score, sociodemographic characteristics (age, sex, regions, social vulnerability, race/ethnicity, high risk occupations), previously tested positive for COVID-19, severity of acute illness and immunocompromised status | 64 | 13 | 85 |
| Di Fusco, 2023 (BNT162b2; 3 doses) | 6 months | Omicron | Vaccination status | Boosted vs unvaccinated (≥2 symptoms) | Adjusted for timepoint, vaccination status and interaction of time by vaccination status, participant pre-COVID-19 symptom onset score, sociodemographic characteristics (age, sex, regions, social vulnerability, race/ethnicity, high risk occupations), previously tested positive for COVID-19, severity of acute illness and immunocompromised status | 70 | 30 | 87 |
| Azzolini, 2022 (BNT162b2; 3 doses) | ≥4 weeks post-COVID-19 infection | Wild-type, Alpha, Delta, Omicron | NA | NA | Adjusted for sex, vaccine dose, allergies and comorbidities | 84 | 16 | 97 |
| Gallant, 2022 (BNT162b2 or mRNA-1273; ≥2 doses) | >30 days | Delta | NA | NA | Adjusted for age, sex, number of symptoms during acute COVID-19, and number of comorbidities. | -3 | -18 | 10 |
| Richard, 2023 (BNT162b2, mRNA-1273 or Ad26.COV2.S; primary series) | ≥90 days | Wild-type, Alpha, Delta | Timepoint | ≥90 days | Adjusted for Delta variant period, maximum symptom severity during acute illness, days hospitalized for COVID-19, comorbidities, acute respiratory symptoms, age, sex and BMI category | 27 | -14 | 53 |
| Elmazny, 2023 (BNT162b2, Ad26.COV2.S, ChAdOx1-S, Sputnik V, CoronaVac, BBIBP-CorV; primary series) | 3 months post-acute infection | WT, Alpha, Delta, Omicron | NA | NA | Adjusted for age, sex, and COVID-19 severity | 40 | 20 | 55 |
| Richard, 2023 (BNT162b2, mRNA-1273 or Ad26.COV2.S; primary series) | ≥28 days | Wild-type, Alpha, Delta | Timepoint | ≥28 days | Adjusted for Delta variant period, maximum symptom severity during acute illness, days hospitalized for COVID-19, comorbidities, acute respiratory symptoms, age, sex and BMI category | 28 | 4 | 46 |
| Di Fusco, 2023 (BNT162b2; 2 doses) | 6 months | Omicron | Vaccination status | Primed vs unvaccinated (≥3 symptoms) | Adjusted for timepoint, vaccination status and interaction of time by vaccination status, participant pre-COVID-19 symptom onset score, sociodemographic characteristics (age, sex, regions, social vulnerability, race/ethnicity, high risk occupations), previously tested positive for COVID-19, severity of acute illness and immunocompromised status | 40 | -34 | 73 |
| Di Fusco, 2023 (BNT162b2; 2 doses) | 6 months | Omicron | Vaccination status | Primed vs unvaccinated (≥2 symptoms) | Adjusted for timepoint, vaccination status and interaction of time by vaccination status, participant pre-COVID-19 symptom onset score, sociodemographic characteristics (age, sex, regions, social vulnerability, race/ethnicity, high risk occupations), previously tested positive for COVID-19, severity of acute illness and immunocompromised status | 36 | -39 | 70 |
| Perlis, 2022 (Ad26.COV2.S; 2 doses) | >3 months post-COVID-19 diagnosis | Alpha, Delta, Omicron | NA | NA | Adjusted for age, gender, race and ethnicity, educational level, urbanicity, region and predominant variant | 28 | 14 | 40 |
| Ayoubkhani, 2022 (BNT162b2, ChAdOx1-S or Ad26.COV2.S; 2 doses) | ≥12 weeks | Delta | Vaccine, long COVID definition | Adenoviral and mRNA; Long COVID of any severity | Adjusted for age, sex, white or non-white ethnicity, country/region of residence, area deprivation quintile group, and self-reported, pre-existing health/disability status and time from infection to follow-up for Long Covid | 41 | 31 | 50 |
| Ayoubkhani, 2022 (BNT162b2, ChAdOx1-S or Ad26.COV2.S; 2 doses) | ≥12 weeks | Delta | Vaccine, long COVID definition | Adenoviral and mRNA; activity-limiting Long COVID | Adjusted for age, sex, white or non-white ethnicity, country/region of residence, area deprivation quintile group, and self-reported, pre-existing health/disability status and time from infection to follow-up for Long Covid | 41 | 27 | 52 |
| Ayoubkhani 2022 (BNT162b2 or mRNA-1273; 2 doses) | ≥12 weeks | Delta | Vaccine, long COVID definition | mRNA vaccine platform; Long COVID of any severity | Adjusted for age, sex, white or non-white ethnicity, country/region of residence, area deprivation quintile group, and self-reported, pre-existing health/disability status and time from infection to follow-up for Long Covid | 50 | 31 | 63 |
| Ayoubkhani 2022 (BNT162b2 or mRNA-1273; 2 doses) | ≥12 weeks | Delta | Vaccine, long COVID definition | mRNA vaccine platform; activity-limiting Long COVID | Adjusted for age, sex, white or non-white ethnicity, country/region of residence, area deprivation quintile group, and self-reported, pre-existing health/disability status and time from infection to follow-up for Long Covid | 50 | 25 | 56 |
| Ayoubkhani 2022 (ChAdOx1-S; 2 doses) | ≥12 weeks | Delta | Vaccine, long COVID definition | Astrazeneca; Long COVID of any severity | Adjusted for age, sex, white or non-white ethnicity, country/region of residence, area deprivation quintile group, and self-reported, pre-existing health/disability status and time from infection to follow-up for Long Covid | 38 | 25 | 49 |
| Ayoubkhani 2022 (ChAdOx1-S; 2 doses) | ≥12 weeks | Delta | Vaccine, long COVID definition | Astrazeneca; activity limiting Long COVID | Adjusted for age, sex, white or non-white ethnicity, country/region of residence, area deprivation quintile group, and self-reported, pre-existing health/disability status and time from infection to follow-up for Long Covid | 37 | 20 | 51 |
| Azzolini, 2022 (BNT162b2; 2 doses) | ≥4 weeks post-COVID-19 infection | Wild-type, Alpha, Delta, Omicron | NA | NA | Adjusted for sex, vaccine dose, allergies and comorbidities | 75 | 13 | 93 |
| Herman, 2022 (inactivated viral vaccine; 2 doses) | <90 days | Delta, Omicron | NA | NA | Adjusted by shortness of breath | 26 | -8 | 49 |
| Parodi, 2023 (mRNA or inactivated viral; 2 doses) | Mean time from acute diagnosis to consultation: 62 days (16–330) | Gamma, Delta, Omicron | NA | NA | Adjusted for age, sex, hypertension and smoking | 48 | 31 | 60 |
| Senjam, 2022 (Vaccine NR; 2 doses) | >4 weeks post-positive COVID-19 test (lasting ≤12 weeks) | Delta | NA | NA | Adjusted for severity of COVID-19, chronic medication use, presence of chronic conditions, self-perceived health status, monthly household income, smoking status, living status, educational level, current occupation, marital status, gender and age | 39 | 4 | 60 |
| Antonelli, 2022 (BNT162b2, mRNA-1273, ChAdOx1-S or NR; 2 doses) | 1 month (≥28 days) | Alpha, Delta | NA | NA | Adjusted for age, BMI, sex | 49 | 18 | 68 |
| Ertesvag, 2023 (BNT162b2 or mRNA-1273; ≥1 dose) | 8 months post-infection | Delta | Timepoint and population | Vaccination as a predictor of any persisting symptoms among adolescents (≥16 years) 8-months post-Delta infection | Adjusted for female sex, age, reinfection, and any symptoms at baseline | -98 | -457 | 30 |
| Kahlert, 2023 (BNT162b2 or mRNA-1273; ≥1 dose) | ≥3 months post-COVID-19 infection (Median time since infection was 18.3 (IQR: 17.5–19.2) months for wild-type, 6.5 (IQR: 6.0–9.0) months for Alpha/Delta, and 3.1 (IQR: 2.6–4.0) months for Omicron BA.1 infection) | Wild-type, Alpha, Delta, Omicron | NA | NA | Adjusted for age, sex, BMI, ethnicity, children in household, comorbidity, medication, smoker status, alcohol intake, medical profession, COVID-19 patient contact, ICU work, full-time work, SARS-CoV-2 variant | -27 | -94 | 18 |
| Ertesvag, 2023 (BNT162b2 or mRNA-1273; ≥1 dose) | 3 months | Delta | Timepoint | Vaccination as a predictor of any persisting symptoms 3-months post-Delta infection | Adjusted by female sex, age, reinfection, any symptoms at baseline. | -123 | -904 | 45 |
| Durstenfeld, 2022 (Vaccine NR; ≥1 dose) | >1 month post acute infection | Wild-type, Alpha, Delta, Omicron | NA | NA | Adjusted for age, COVID-19 wave, number of initial symptoms, sex, myocardial infarction, stroke, atrial fibrillation, sleep apnoea, COPD or asthma, and immunodeficiency, plus receipt of COVID-19 vaccination before or after COVID-19 diagnosis, ethnicity, subjective socioeconomic status, highest level of education, primary employment, pre-COVID depressive symptoms, anxiety symptoms, and financial security | 19 | -49 | 56 |
| Antonelli, 2022b (Vaccine NR; ≥1 dose) | 28 days post infection | Delta, Omicron | Time since vaccination | Vaccinated >6 months prior to infection | Adjusted by age, sex, BMI, index of multiple deprivation, presence of comorbidities and vaccination status | 74 | 68 | 80 |
| Antonelli, 2022b (Vaccine NR; ≥1 dose) | 28 days post infection | Delta, Omicron | Time since vaccination | Vaccinated 3–6 months prior to infection | Adjusted by age, sex, BMI, index of multiple deprivation, presence of comorbidities and vaccination status | 76 | 68 | 81 |
| Antonelli, 2022b (Vaccine NR; ≥1 dose) | 28 days post infection | Delta, Omicron | Time since vaccination | Vaccinated <3 months prior to infection | Adjusted by age, sex, BMI, index of multiple deprivation, presence of comorbidities and vaccination status | 50 | 41 | 57 |
| Antonelli, 2022b (Vaccine NR; ≥1 dose) | 28 days post infection | Delta, Omicron | Age; Time since vaccination | 18–59; vaccinated >6 months prior to infection | Adjusted by age, sex, BMI, index of multiple deprivation, presence of comorbidities and vaccination status | 77 | 68 | 84 |
| Antonelli, 2022b (Vaccine NR; ≥1 dose) | 28 days post infection | Delta, Omicron | Age; Time since vaccination | 18–59; vaccinated 3–6 months prior to infection | Adjusted by age, sex, BMI, index of multiple deprivation, presence of comorbidities and vaccination status | 67 | 56 | 76 |
| Antonelli, 2022b (Vaccine NR; ≥1 dose) | 28 days post infection | Delta, Omicron | Age; Time since vaccination | 18–59; vaccinated <3 months prior to infection | Adjusted by age, sex, BMI, index of multiple deprivation, presence of comorbidities and vaccination status | 59 | 47 | 68 |
| Antonelli, 2022b (Vaccine NR; ≥1 dose) | 28 days post infection | Delta, Omicron | Age; Time since vaccination | ≥60; vaccinated >6 months prior to infection | Adjusted by age, sex, BMI, index of multiple deprivation, presence of comorbidities and vaccination status | 72 | 61 | 80 |
| Antonelli, 2022b (Vaccine NR; ≥1 dose) | 28 days post infection | Delta, Omicron | Age; Time since vaccination | ≥60; vaccinated 3–6 months prior to infection | Adjusted by age, sex, BMI, index of multiple deprivation, presence of comorbidities and vaccination status | 54 | -3 | 80 |
| Antonelli, 2022b (Vaccine NR; ≥1 dose) | 28 days post infection | Delta, Omicron | Age; Time since vaccination | ≥60; vaccinated <3 months prior to infection | Adjusted by age, sex, BMI, index of multiple deprivation, presence of comorbidities and vaccination status | 46 | 32 | 57 |
| Perlis, 2022 (Ad26.COV2.S; 1 dose) | >3 months post-COVID-19 diagnosis | Alpha, Delta, Omicron | NA | NA | Adjusted for age, gender, race and ethnicity, educational level, urbanicity, region and predominant variant | 7 | -25 | 31 |
| Xue, 2023 (BNT162b2 or mRNA-1273; 1 dose) | ≥3 months post-COVID-19 infection | Alpha, Gamma, Delta, Omicron | NA | NA | Adjusted for age, gender, ethnicity, smoking status, alcohol consumption, educational level, since since first vaccination, martial status, living area, level of physical activity, weight status, comorbidities. | -13 | -34 | 5 |
| Senjam, 2022 (Vaccine NR; 1 dose) | >4 weeks post-positive COVID-19 test (lasting ≤12 weeks) | Delta | NA | NA | Adjusted for age, gender, educational level, BMI, workplace, blood group, smoking, alcohol consumption, presence of comorbidities, oxygen requirement, severity of COVID-19 disease, overall health status following COVUD-19 illness | 9 | -40 | 41 |
| Azzolini, 2022 (BNT162b2; 1 dose) | ≥4 weeks post-COVID-19 infection | Wild-type, Alpha, Delta, Omicron | NA | NA | Adjusted for sex, vaccine dose, allergies and comorbidities | 14 | -249 | 79 |
| Antonelli, 2022 (BNT162b2, mRNA-1273, ChAdOx1-S or NR; 1 dose) | 1 month (≥28 days) | Alpha, Delta | NA | NA | Adjusted for age, BMI, sex | -3 | -24 | 15 |
| Patel, 2023 (BNT162b2, mRNA-1273 or Ad26.COV2.S; primary series) | ≥90 days post-acute infection | Delta, Omicron | PASC definition | WHO; ≥90 days post-infection | Adjusted for age, sex, race, comorbidity count and use of any one of the following medications: anti-CD20 monoclonal antibodies, methotrexate, mycophenolate or glucocorticoids. | 90 | 78 | 96 |
| Patel, 2023 (BNT162b2, mRNA-1273 or Ad26.COV2.S; primary series) | ≥28 days post-acute infection | Delta, Omicron | PASC definition | CDC; ≥28 days | Adjusted for age, sex, race, comorbidity count and use of any one of the following medications: anti-CD20 monoclonal antibodies, methotrexate, mycophenolate or glucocorticoids. | 51 | 17 | 71 |
| Ranucci, 2023 (BNT162b2, mRNA-1273, ChAdOx1-S or Ad26.COV2.S or NR; 2 doses) | 3–20 months post-COVID-19 infection | Wild-type, Alpha, Delta, Omicron | Outcome | Persistent major physical symptoms | Adjusted for age, severity of disease | 69 | 40 | 84 |
| Ranucci, 2023 (BNT162b2, mRNA-1273, ChAdOx1-S or Ad26.COV2.S or NR; 2 doses) | 3–20 months post-COVID-19 infection | Wild-type, Alpha, Delta, Omicron | Outcome | Persistent major neuropsychological symptoms | Adjusted for previous neurological symptoms | 80 | 57 | 90 |
| Mohr, 2023 (BNT162b2 or mRNA-1273; 2 doses) | 6 weeks | Wild-type, Alpha, Delta | Timepoint | >16 weeks since vaccination; COVID-like symptoms | Adjusted for age, race and ethnicity, comorbidities, calendar month of diagnosis, and weeks since symptoms started | 28 | 12 | 41 |
| Mohr, 2023 (BNT162b2 or mRNA-1273; 2 doses) | 6 weeks | Wild-type, Alpha, Delta | Timepoint | ≤16 weeks since vaccination; COVID-like symptoms | Adjusted for age, race and ethnicity, comorbidities, calendar month of diagnosis, and weeks since symptoms started | 33 | 13 | 49 |
| Mohr, 2023 (BNT162b2 or mRNA-1273; 2 doses) | 6 weeks | Wild-type, Alpha, Delta | Timepoint | 6 weeks | Adjusted for age, race and ethnicity, comorbidities, calendar month of diagnosis and weeks since symptoms started | 30 | 16 | 42 |

BMI: Body mass index; CDC: Centers for Disease Control and Prevention; CI: Confidence interval; CPRD: Clinical Practice Research Datalink; GP: General Practitioner; HHS: Health and Human Services; HR: Hazard ratio; ICD-10: International Classification of Diseases- 10; ICU: Intensive care unit; IPTW: Inverse probability of treatment weighting; IQR: Interquartile range; mRNA: Messenger ribonucleic acid; NA: Not applicable; NR: Not reported; OR: Odds ratio; PASC: Post-acute sequelae of COVID-19; PCR: Polymerase Chain Reaction; VA: Veterans Affairs; VE: Vaccine effectiveness; VISN: Veterans Integrated Services Network.

**eTable 18. Summary of publications where adjusted VE (vs. unvaccinated) against ≥1 PCC could be computed, by PCC follow-up time (N=31 studies with 93 VE estimates)**

| **Author, year (vaccine and dose)** | **PCC Follow-up timepoint** | **Variant** | **Subgroup** | **Subgroup details** | **Adjustments** | **VE (%)** | **Lower 95% CI** | **Upper 95% CI** |
| --- | --- | --- | --- | --- | --- | --- | --- | --- |
| Antonelli, 2022b (Vaccine NR, ≥1 dose) | 28 days post infection | Delta, Omicron | Time since vaccination | Vaccinated >6 months prior to infection | Adjusted by age, sex, BMI, index of multiple deprivation, presence of comorbidities and vaccination status | 74 | 68 | 80 |
| Antonelli, 2022b (Vaccine NR, ≥1 dose) | 28 days post infection | Delta, Omicron | Time since vaccination | Vaccinated 3–6 months prior to infection | Adjusted by age, sex, BMI, index of multiple deprivation, presence of comorbidities and vaccination status | 76 | 68 | 81 |
| Antonelli, 2022b (Vaccine NR, ≥1 dose) | 28 days post infection | Delta, Omicron | Time since vaccination | Vaccinated <3 months prior to infection | Adjusted by age, sex, BMI, index of multiple deprivation, presence of comorbidities and vaccination status | 50 | 41 | 57 |
| Antonelli, 2022b (Vaccine NR, ≥1 dose) | 28 days post infection | Delta, Omicron | Age; Time since vaccination | ≥60; vaccinated >6 months prior to infection | Adjusted by age, sex, BMI, index of multiple deprivation, presence of comorbidities and vaccination status | 72 | 61 | 80 |
| Antonelli, 2022b (Vaccine NR, ≥1 dose) | 28 days post infection | Delta, Omicron | Age; Time since vaccination | ≥60; vaccinated 3–6 months prior to infection | Adjusted by age, sex, BMI, index of multiple deprivation, presence of comorbidities and vaccination status | 54 | -3 | 80 |
| Antonelli, 2022b (Vaccine NR, ≥1 dose) | 28 days post infection | Delta, Omicron | Age; Time since vaccination | ≥60; vaccinated <3 months prior to infection | Adjusted by age, sex, BMI, index of multiple deprivation, presence of comorbidities and vaccination status | 46 | 32 | 57 |
| Antonelli, 2022b (Vaccine NR, ≥1 dose) | 28 days post infection | Delta, Omicron | Age; Time since vaccination | 18–59; vaccinated >6 months prior to infection | Adjusted by age, sex, BMI, index of multiple deprivation, presence of comorbidities and vaccination status | 77 | 68 | 84 |
| Antonelli, 2022b (Vaccine NR, ≥1 dose) | 28 days post infection | Delta, Omicron | Age; Time since vaccination | 18–59; vaccinated 3–6 months prior to infection | Adjusted by age, sex, BMI, index of multiple deprivation, presence of comorbidities and vaccination status | 67 | 56 | 76 |
| Antonelli, 2022b (Vaccine NR, ≥1 dose) | 28 days post infection | Delta, Omicron | Age; Time since vaccination | 18–59; vaccinated <3 months prior to infection | Adjusted by age, sex, BMI, index of multiple deprivation, presence of comorbidities and vaccination status | 59 | 47 | 68 |
| Durstenfeld, 2022 (Vaccine NR, ≥1 dose) | >1 month post-acute infection | Wild-type, Alpha, Delta, Omicron | NA | NA | Adjusted for age, COVID-19 wave, number of initial symptoms, sex, myocardial infarction, stroke, atrial fibrillation, sleep apnoea, COPD or asthma, and immunodeficiency, plus receipt of COVID-19 vaccination before or after COVID-19 diagnosis, ethnicity, subjective socioeconomic status, highest level of education, primary employment, pre-COVID depressive symptoms, anxiety symptoms, and financial security | 19 | -49 | 56 |
| Antonelli, 2022a (BNT162b2, mRNA-1273, ChAdOx1-S or NR, 1 dose) | 1 month (≥28 days) | Alpha, Delta | Number of doses | 1 | Adjusted for age, BMI, sex | -3 | -24 | 15 |
| Antonelli, 2022a (BNT162b2, mRNA-1273, ChAdOx1-S or NR, 1 dose) | 1 month (≥28 days) | Alpha, Delta | Number of doses | 2 | Adjusted for age, BMI, sex | 49 | 18 | 68 |
| Tannous, 2023 (mRNA or adenoviral, primary series) | >28 days post-acute infection | Wild-type, Alpha, Delta | NA | NA | Adjusted for age, sex, race, ethnicity, Charlson Comorbidity Index, COVID-19 illness severity, and Texas state area deprivation index | 42 | 34 | 48 |
| Richard, 2023 (BNT162b2, mRNA-1273 or Ad26.COV.S, primary series) | ≥28 days | Wild-type, Alpha, Delta | NA | NA | Adjusted for Delta variant period, maximum symptom severity during acute illness, days hospitalized for COVID-19, comorbidities, acute respiratory symptoms, age, sex and BMI category | 28 | 4 | 46 |
| Patel, 2023 (BNT162b2, mRNA-1273 or Ad26.COV2.S, primary series) | ≥28 days post-acute infection | Delta, Omicron | PASC definition | CDC; ≥28 days | Adjusted for age, sex, race, comorbidity count and use of any one of the following medications: anti-CD20 monoclonal antibodies, methotrexate, mycophenolate or glucocorticoids. | 51 | 17 | 71 |
| Gallant, 2022 (BNT162b2 or mRNA-1273, ≥2 doses) | >30 days | Delta | NA | NA | Adjusted for age, sex, number of symptoms during acute COVID-19, and number of comorbidities. | -3 | -18 | 10 |
| Tosi, 2023 (BNT162b2, mRNA-1273 or Ad26.COV2.S, primary series) | NR (specified survival ≥30 days after acute infection) | Omicron | Vaccinated, robust patient | NA | Adjusted for age, BMI, race, ethnicity, smoking, active patient in the past 24 months, and rurality | 14 | 7 | 19 |
| Tosi, 2023 (BNT162b2, mRNA-1273 or Ad26.COV2.S, primary series) | NR (specified survival at ≥30 days after acute infection) | Omicron | Vaccinated, pre-frail patient | NA | Adjusted for age, BMI, race, ethnicity, smoking, active patient in the past 24 months, and rurality | 20 | 13 | 26 |
| Tosi, 2023 (BNT162b2, mRNA-1273 or Ad26.COV2.S, primary series) | NR (specified survival at ≥30 days after acute infection) | Omicron | Vaccinated, frail patient | NA | Adjusted for age, BMI, race, ethnicity, smoking, active patient in the past 24 months, and rurality | 20 | 13 | 27 |
| Tosi, 2023 (BNT162b2, mRNA-1273 or Ad26.COV2.S, primary series) | NR (specified survival at ≥30 days after acute infection) | Omicron | Boosted, robust patient | NA | Adjusted for age, BMI, race, ethnicity, smoking, active patient in the past 24 months, and rurality | 26 | 18 | 33 |
| Tosi, 2023 (BNT162b2, mRNA-1273 or Ad26.COV2.S, primary series) | NR (specified survival at ≥30 days after acute infection) | Omicron | Boosted, pre-frail patient | NA | Adjusted for age, BMI, race, ethnicity, smoking, active patient in the past 24 months, and rurality | 35 | 28 | 41 |
| Tosi, 2023 (BNT162b2, mRNA-1273 or Ad26.COV2.S, primary series) | NR (specified survival at ≥30 days after acute infection) | Omicron | Boosted, frail patient | NA | Adjusted for age, BMI, race, ethnicity, smoking, active patient in the past 24 months, and rurality | 30 | 23 | 36 |
| Senjam, 2022 (Vaccine NR, 1 dose) | >4 weeks post-positive COVID-19 test (lasting ≤12 weeks) | Delta | Number of doses | 1 | Adjusted for age, gender, educational level, BMI, workplace, blood group, smoking, alcohol consumption, presence of comorbidities, oxygen requirement, severity of COVID-19 disease, overall health status following COVUD-19 illness | 9 | -40 | 41 |
| Senjam, 2022 (Vaccine NR, 1 dose) | >4 weeks post-positive COVID-19 test (lasting ≤12 weeks) | Delta | Number of doses | 2 | Adjusted for severity of COVID-19, chronic medication use, presence of chronic conditions, self-perceived health status, monthly household income, smoking status, living status, educational level, current occupation, marital status, gender and age | 39 | 4 | 60 |
| Azzolini, 2022 (BNT162b2, 1 dose) | ≥4 weeks post-COVID-19 infection | Wild-type, Alpha, Delta, Omicron | Number of doses | 1 | Adjusted for sex, vaccine dose, allergies and comorbidities | 14 | -249 | 79 |
| Azzolini, 2022 (BNT162b2, 2 doses) | ≥4 weeks post-COVID-19 infection | Wild-type, Alpha, Delta, Omicron | Number of doses | 2 | Adjusted for sex, vaccine dose, allergies and comorbidities | 75 | 13 | 93 |
| Azzolini, 2022 (BNT162b2, 3 doses) | ≥4 weeks post-COVID-19 infection | Wild-type, Alpha, Delta, Omicron | Number of doses | 3 | Adjusted for sex, vaccine dose, allergies and comorbidities | 84 | 16 | 97 |
| Cortellini, 2023 (BNT162b2, mRNA-1273, Ad26.COV2.S or ChAdOx1-S, 2 doses) | 4–12 weeks post-acute infection (Unvaccinated patients were assessed at a median of 40 days (IQR 25–67) and doubly vaccinated patients 38 days (IQR: 23–68)) | Alpha, Delta, Omicron | Number of doses | 2 versus 0 | Inverse probability of treatment weighting applied | 59 | 30 | 76 |
| Cortellini, 2023 (BNT162b2, mRNA-1273, Ad26.COV2.S or ChAdOx1-S, 3 doses) | 4–12 weeks post-acute infection (Unvaccinated patients were assessed at a median of 40 days (IQR 25–67) and doubly vaccinated patients 38 days (IQR: 23–68)) | Alpha, Delta, Omicron | Number of doses | 3 versus 0 | Inverse probability of treatment weighting applied | 53 | -18 | 82 |
| Català, 2023 (ChAdOx1-S, BNT162b2, Ad26.COV2.S or mRNA-1273, 1 dose) | 28-365 days | Delta, Omicron | Cohort | SIDIAP, ≥28 days | Propensity score overlap weighting for age (5-year age bands for people aged ≥18 years), location, index date, previous observation years, number of previous GP visits, number of previous SARS-CoV-2 PCR tests, regional vaccination, testing | 12 | 2 | 21 |
| Català, 2023 (ChAdOx1-S, BNT162b2, Ad26.COV2.S or mRNA-1273, 1 dose) | 28-365 days | Delta, Omicron | Cohort | CPRD AURUM, ≥28 days | Propensity score overlap weighting for age (5-year age bands for people aged ≥18 years), location, index date, previous observation years, number of previous GP visits, number of previous SARS-CoV-2 PCR tests, regional vaccination, testing, COVID-19 incidence rates | 30 | 16 | 42 |
| Català, 2023 (ChAdOx1-S, BNT162b2, Ad26.COV2.S or mRNA-1273, 1 dose) | 28-365 days | Delta, Omicron | Cohort | CPRD GOLD, ≥28 days | Propensity score overlap weighting for age (5-year age bands for people aged ≥18 years), location, index date, previous observation years, number of previous GP visits, number of previous SARS-CoV-2 PCR tests, regional vaccination, testing, COVID-19 incidence rates | 34 | 17 | 47 |
| Català, 2023 (ChAdOx1-S, BNT162b2, Ad26.COV2.S or mRNA-1273, 1 dose) | 28-365 days | Delta, Omicron | Cohort | CORIVA, ≥28 days | Propensity score overlap weighting for age (5-year age bands for people aged ≥18 years), location, index date, previous observation years, number of previous GP visits, number of previous SARS-CoV-2 PCR tests, regional vaccination, testing | 38 | 23 | 50 |
| Smits, 2023 (BNT162b2, mRNA-1273, NVX-CoV2373 or Ad26.COV2.S, primary series) | 28–365 days post-positive COVID-19 test | Omicron | Vaccination status | Long COVID diagnosis (ICD-10 codes; 28–365 days post-positive COVID-19 test) | Adjusted for vaccination status, comorbidities, sex, race, ethnicity, age, timing of infection, smoking status, influenza vaccination status, inpatient and outpatient encounters in previous two years | 14 | 1 | 26 |
| Smits, 2023 (BNT162b2, mRNA-1273, NVX-CoV2373 or Ad26.COV2.S, primary series) | 28–365 days post-positive COVID-19 test | Omicron | Vaccination status | Long-COVID symptoms (28-365 days post-positive COVID-19 test) | Adjusted for vaccination status, comorbidities, sex, race, ethnicity, age, timing of infection, smoking status, influenza vaccination status, inpatient and outpatient encounters in previous two years | 10 | 6 | 13 |
| Smits, 2023 (BNT162b2, mRNA-1273, NVX-CoV2373 or Ad26.COV2.S, primary series) | 28–365 days post-positive COVID-19 test | Omicron | Vaccination status | Long COVID diagnosis (ICD-10 codes; 28-365 days post-positive COVID-19 test) | Adjusted for vaccination status, comorbidities, sex, race, ethnicity, age, timing of infection, smoking status, influenza vaccination status, inpatient and outpatient encounters in previous two years | 19 | 5 | 31 |
| Smits, 2023 (BNT162b2, mRNA-1273, NVX-CoV2373 or Ad26.COV2.S, primary series) | 28–365 days post-positive COVID-19 test | Omicron | Vaccination status | Long-COVID symptoms (28-365 days post-positive COVID-19 test) | Adjusted for vaccination status, comorbidities, sex, race, ethnicity, age, timing of infection, smoking status, influenza vaccination status, inpatient and outpatient encounters in previous two years | 13 | 9 | 17 |
| Mohr, 2023 (BNT162b2 or mRNA-1273, 2 doses) | >16 weeks | Wild-type, Alpha, Delta | Timepoint | >16 weeks since vaccination; COVID-like symptoms | Adjusted for age, race and ethnicity, comorbidities, calendar month of diagnosis, and weeks since symptoms started | 28 | 12 | 41 |
| Mohr, 2023 (BNT162b2 or mRNA-1273, 2 doses) | ≤16 weeks | Wild-type, Alpha, Delta | Timepoint | ≤16 weeks since vaccination; COVID-like symptoms | Adjusted for age, race and ethnicity, comorbidities, calendar month of diagnosis, and weeks since symptoms started | 33 | 13 | 49 |
| Mohr, 2023 (BNT162b2 or mRNA-1273, 2 doses) | 6 weeks | Wild-type, Alpha, Delta | Timepoint | 6 weeks | Adjusted for age, race and ethnicity, comorbidities, calendar month of diagnosis and weeks since symptoms started | 30 | 16 | 42 |
| Brannock, 2023 (BNT162b2, mRNA-1273 or ChAdOx1-S, primary series) | >45 days post COVID-19 infection (minimum of 164 days) | Delta, Omicron | Methodology - logistic regression analysis | Clinic-based cohort IPTW-adjusted OR | Inverse probability of treatment weighting applied | 30 | 19 | 40 |
| Brannock, 2023 (BNT162b2, mRNA-1273 or ChAdOx1-S, primary series) | >45 days post COVID-19 infection (minimum of 164 days) | Delta, Omicron | Methodology - proportional hazards | Clinic-based cohort IPTW-adjusted HR | Inverse probability of treatment weighting applied | 33 | 21 | 44 |
| Brannock, 2023 (BNT162b2, mRNA-1273 or ChAdOx1-S, primary series) | >45 days post COVID-19 infection (minimum of 164 days) | Delta, Omicron | Methodology - logistic regression analysis | Model-based cohort IPTW-adjusted OR | Inverse probability of treatment weighting applied | 30 | 25 | 35 |
| Brannock, 2023 (BNT162b2, mRNA-1273 or ChAdOx1-S, primary series) | >45 days post COVID-19 infection (minimum of 164 days) | Delta, Omicron | Methodology - proportional hazards | Model-based cohort IPTW-adjusted HR | Inverse probability of treatment weighting applied | 37 | 31 | 43 |
| Parodi, 2023 (mRNA or inactivated viral, 2 doses) | Mean time from acute diagnosis to consultation: 62 days (16–330) | Gamma, Delta, Omicron | NA | NA | Adjusted for age, sex, hypertension and smoking | 48 | 31 | 60 |
| Herman, 2022 (Inactivated viral vaccine, 2 doses) | <90 days | Delta, Omicron | NA | NA | Adjusted by shortness of breath | 26 | -8 | 49 |
| Ertesvag, 2023 (BNT162b2 or mRNA-1273, ≥1 dose [95% had 1 dose]) | 3 months | Delta | Timepoint | Vaccination as a predictor of any persisting symptoms 3-months post-Delta infection | Adjusted by female sex, age, reinfection, any symptoms at baseline. | -123 | -904 | 45 |
| Abu Hamdh, 2023 (BNT162b2, mRNA-1273, Sputnik V, Sputnik Light or ChAdOx1-S, ≥1 dose) | 90 days | NR | NA | NA | Adjusted for sex, age, comorbidities, hospital admission, dyspnoea, previous COVID-19 infection, fatigue | 86 | 76 | 91 |
| Ayoubkhani, 2022 (ChAdOx1-S, 2 doses) | ≥12 weeks | Delta | Vaccine and long COVID severity | AstraZeneca; activity limiting Long COVID | Adjusted for age, sex, white or non-white ethnicity, country/region of residence, area deprivation quintile group, and self-reported, pre-existing health/disability status and time from infection to follow-up for Long Covid | 37 | 20 | 51 |
| Ayoubkhani, 2022 (ChAdOx1-S, 2 doses) | ≥12 weeks | Delta | Vaccine and long COVID severity | AstraZeneca; Long COVID of any severity | Adjusted for age, sex, white or non-white ethnicity, country/region of residence, area deprivation quintile group, and self-reported, pre-existing health/disability status and time from infection to follow-up for Long Covid | 38 | 25 | 49 |
| Ayoubkhani, 2022 (BNT162b2 or mRNA-1273, 2 doses) | ≥12 weeks | Delta | Vaccine and long COVID severity | mRNA vaccine platform; activity-limiting Long COVID | Adjusted for age, sex, white or non-white ethnicity, country/region of residence, area deprivation quintile group, and self-reported, pre-existing health/disability status and time from infection to follow-up for Long Covid | 50 | 25 | 56 |
| Ayoubkhani, 2022 (BNT162b2, ChAdOx1-S or Ad26.COV2.S, 2 doses) | ≥12 weeks | Delta | Vaccine and long COVID severity | mRNA vaccine platform; Long COVID of any severity | Adjusted for age, sex, white or non-white ethnicity, country/region of residence, area deprivation quintile group, and self-reported, pre-existing health/disability status and time from infection to follow-up for Long Covid | 50 | 31 | 63 |
| Ayoubkhani, 2022 (BNT162b2 or mRNA-1273, 2 doses) | ≥12 weeks | Delta | Vaccine and long COVID severity | Adenoviral and mRNA; activity-limiting Long COVID | Adjusted for age, sex, white or non-white ethnicity, country/region of residence, area deprivation quintile group, and self-reported, pre-existing health/disability status and time from infection to follow-up for Long Covid | 41 | 27 | 52 |
| Ayoubkhani, 2022 (BNT162b2, ChAdOx1-S or Ad26.COV2.S, 2 doses) | ≥12 weeks | Delta | Vaccine and long COVID severity | Adenoviral and mRNA; Long COVID of any severity | Adjusted for age, sex, white or non-white ethnicity, country/region of residence, area deprivation quintile group, and self-reported, pre-existing health/disability status and time from infection to follow-up for Long Covid | 41 | 31 | 50 |
| Xue, 2023 (BNT162b2 or mRNA-1273, 1 dose) | ≥3 months post-COVID-19 infection | Alpha, Gamma, Delta, Omicron | NA | NA | Adjusted for age, gender, ethnicity, smoking status, alcohol consumption, educational level, since first vaccination, martial status, living area, level of physical activity, weight status, comorbidities. | -13 | -34 | 5 |
| Kahlert, 2023 (BNT162b2 or mRNA-1273, ≥1 dose) | ≥3 months post-COVID-19 infection (Median time since infection was 18.3 (IQR: 17.5–19.2) months for wild-type, 6.5 (IQR: 6.0–9.0) months for Alpha/Delta, and 3.1 (IQR: 2.6–4.0) months for Omicron BA.1 infection) | Wild-type, Alpha, Delta, Omicron | NA | NA | Adjusted for age, sex, BMI, ethnicity, children in household, comorbidity, medication, smoker status, alcohol intake, medical profession, COVID-19 patient contact, ICU work, full-time work, SARS-CoV-2 variant | -27 | -94 | 18 |
| Richard, 2023 (BNT162b2, mRNA-1273 or Ad26.COV.S, primary series) | ≥90 days | Wild-type, Alpha, Delta | Timepoint | ≥90 days | Adjusted for Delta variant period, maximum symptom severity during acute illness, days hospitalized for COVID-19, comorbidities, acute respiratory symptoms, age, sex and BMI category | 27 | -14 | 53 |
| Patel, 2023 (BNT162b2, mRNA-1273 or Ad26.COV2.S, primary series) | ≥90 days post-acute infection | Delta, Omicron | PASC definition | WHO; ≥90 days post-infection | Adjusted for age, sex, race, comorbidity count and use of any one of the following medications: anti-CD20 monoclonal antibodies, methotrexate, mycophenolate or glucocorticoids. | 90 | 78 | 96 |
| Perlis, 2022 (Ad26.COV2.S, 1 dose) | >3 months post-COVID-19 diagnosis | Alpha, Delta, Omicron | NA | NA | Adjusted for age, gender, race and ethnicity, educational level, urbanicity, region and predominant variant | 7 | -25 | 31 |
| Perlis, 2022 (Ad26.COV2.S, 2 doses) | >3 months post-COVID-19 diagnosis | Alpha, Delta, Omicron | NA | NA | Adjusted for age, gender, race and ethnicity, educational level, urbanicity, region and predominant variant | 28 | 14 | 40 |
| Ranucci, 2023 (BNT162b2, mRNA-1273, Ad26.COV2.S, ChAdOx1-S or NR, 2 doses) | 3–20 months post-COVID-19 infection | Wild-type, Alpha, Delta, Omicron | NA | Persistent major physical symptoms | Adjusted for age, severity of disease | 69 | 40 | 84 |
| Ranucci, 2023 (BNT162b2, mRNA-1273, Ad26.COV2.S, ChAdOx1-S or NR, 2 doses) | 3–20 months post-COVID-19 infection | Wild-type, Alpha, Delta, Omicron | Outcome | Persistent major neuropsychological symptoms | Adjusted for previous neurological symptoms | 80 | 57 | 90 |
| Elmazny, 2023 (BNT162b2, Ad26.COV2.S, ChAdOx1-S, Sputnik V, CoronaVac orBBIBP-CorV, primary series) | 3 months post-acute infection | WT, Alpha, Delta, Omicron | NA | NA | Adjusted for age, sex, and COVID-19 severity | 40 | 20 | 55 |
| Català, 2023 (ChAdOx1-S, 1 dose) | 90–365 days post-positive test or diagnosis of COVID-19 | Delta, Omicron | Vaccination group | SIDIAP (AstraZeneca vaccine) | Propensity score overlap weighting for age (5-year age bands for people aged ≥18 years), location, index date, previous observation years, number of previous GP visits, number of previous SARS-CoV-2 PCR tests, regional vaccination, testing | 16 | 4 | 27 |
| Català, 2023 (ChAdOx1-S, 1 dose) | 90–365 days post-positive test or diagnosis of COVID-19 | Delta, Omicron | Vaccination group | CPRD GOLD (AstraZeneca vaccine) | Propensity score overlap weighting for age (5-year age bands for people aged ≥18 years), location, index date, previous observation years, number of previous GP visits, number of previous SARS-CoV-2 PCR tests, regional vaccination, testing, COVID-19 incidence rates | 37 | 23 | 48 |
| Català, 2023 (ChAdOx1-S, 1 dose) | 90–365 days post-positive test or diagnosis of COVID-19 | Delta, Omicron | Vaccination group | CPRD AURUM (AstraZeneca vaccine) | Propensity score overlap weighting for age (5-year age bands for people aged ≥18 years), location, index date, previous observation years, number of previous GP visits, number of previous SARS-CoV-2 PCR tests, regional vaccination, testing, COVID-19 incidence rates | 29 | 21 | 37 |
| Català, 2023 (BNT162b2, 1 dose) | 90–365 days post-positive test or diagnosis of COVID-19 | Delta, Omicron | Vaccination group | SIDIAP (Pfizer vaccine) | Propensity score overlap weighting for age (5-year age bands for people aged ≥18 years), location, index date, previous observation years, number of previous GP visits, number of previous SARS-CoV-2 PCR tests, regional vaccination, testing | 14 | 5 | 23 |
| Català, 2023 (BNT162b2, 1 dose) | 90–365 days post-positive test or diagnosis of COVID-19 | Delta, Omicron | Vaccination group | CPRD GOLD (Pfizer vaccine) | Propensity score overlap weighting for age (5-year age bands for people aged ≥18 years), location, index date, previous observation years, number of previous GP visits, number of previous SARS-CoV-2 PCR tests, regional vaccination, testing, COVID-19 incidence rates | 51 | 37 | 61 |
| Català, 2023 (BNT162b2, 1 dose) | 90–365 days post-positive test or diagnosis of COVID-19 | Delta, Omicron | Vaccination group | CORIVA (Pfizer vaccine) | Propensity score overlap weighting for age (5-year age bands for people aged ≥18 years), location, index date, previous observation years, number of previous GP visits, number of previous SARS-CoV-2 PCR tests, regional vaccination, testing | 37 | 20 | 50 |
| Català, 2023 (BNT162b2, 1 dose) | 90–365 days post-positive test or diagnosis of COVID-19 | Delta, Omicron | Vaccination group | CPRD AURUM (Pfizer vaccine) | Propensity score overlap weighting for age (5-year age bands for people aged ≥18 years), location, index date, previous observation years, number of previous GP visits, number of previous SARS-CoV-2 PCR tests, regional vaccination, testing, COVID-19 incidence rates | 47 | 38 | 55 |
| Català, 2023 (BNT162b2, mRNA-1273, ChAdOx1-S or Ad26.COV2.S, 1 dose) | 90–365 days post-positive test or diagnosis of COVID-19 | Delta, Omicron | Cohort | SIDIAP, GP diagnosis | Propensity score overlap weighting for age (5-year age bands for people aged ≥18 years), location, index date, previous observation years, number of previous GP visits, number of previous SARS-CoV-2 PCR tests, regional vaccination, testing | 40 | 33 | 48 |
| Català, 2023 (BNT162b2, mRNA-1273, ChAdOx1-S or Ad26.COV2.S, 1 dose) | 90–365 days post-positive test or diagnosis of COVID-19 | Delta, Omicron | Vaccination group | SIDIAP | Propensity score overlap weighting for age (5-year age bands for people aged ≥18 years), location, index date, previous observation years, number of previous GP visits, number of previous SARS-CoV-2 PCR tests, regional vaccination, testing | 16 | 7 | 25 |
| Català, 2023 (BNT162b2, mRNA-1273, ChAdOx1-S or Ad26.COV2.S, 1 dose) | 90–365 days post-positive test or diagnosis of COVID-19 | Delta, Omicron | Cohort | CPRD GOLD | Propensity score overlap weighting for age (5-year age bands for people aged ≥18 years), location, index date, previous observation years, number of previous GP visits, number of previous SARS-CoV-2 PCR tests, regional vaccination, testing, COVID-19 incidence rates | 45 | 35 | 54 |
| Català, 2023 (BNT162b2, mRNA-1273, ChAdOx1-S or Ad26.COV2.S, 1 dose) | 90–365 days post-positive test or diagnosis of COVID-19 | Delta, Omicron | Vaccination group | CORIVA | Propensity score overlap weighting for age (5-year age bands for people aged ≥18 years), location, index date, previous observation years, number of previous GP visits, number of previous SARS-CoV-2 PCR tests, regional vaccination, testing | 38 | 22 | 51 |
| Català, 2023 (BNT162b2, mRNA-1273, ChAdOx1-S or Ad26.COV2.S, 1 dose) | 90–365 days post-positive test or diagnosis of COVID-19 | Delta, Omicron | Vaccination group | CPRD AURUM, GP diagnosis | Propensity score overlap weighting for age (5-year age bands for people aged ≥18 years), location, index date, previous observation years, number of previous GP visits, number of previous SARS-CoV-2 PCR tests, regional vaccination, testing, COVID-19 incidence rates | 36 | 23 | 46 |
| Català, 2023 (BNT162b2, mRNA-1273, ChAdOx1-S or Ad26.COV2.S, 1 dose) | 90–365 days post-positive test or diagnosis of COVID-19 | Delta, Omicron | Vaccination group | CPRD AURUM | Propensity score overlap weighting for age (5-year age bands for people aged ≥18 years), location, index date, previous observation years, number of previous GP visits, number of previous SARS-CoV-2 PCR tests, regional vaccination, testing, COVID-19 incidence rates | 36 | 26 | 45 |
| Ioannou, 2022 (BNT162b2 or mRNA-1273, 1 dose) | Mean: 13.5 months (range: 8–23) | Wild-type, Alpha | NA | NA | Adjusted for age, sex, race, ethnicity, urban vs rural residence, Charlson Comorbidity Index score, VA Integrated Service Network, time period of infection/pandemic wave, number of primary care, mental health and specialty care encounters in the 2 years before infection | -3 | -10 | 5 |
| Ioannou, 2022 (BNT162b2 or mRNA-1273, 2 doses) | Mean: 13.5 months (range: 8–23) | Wild-type, Alpha | NA | NA | Adjusted for age, sex, race, ethnicity, urban vs rural residence, Charlson Comorbidity Index score, VA Integrated Service Network, time period of infection/pandemic wave, number of primary care, mental health and specialty care encounters in the 2 years before infection | 22 | 10 | 32 |
| Al-Aly, 2022 (BNT162b2, mRNA-1273 or Ad26.COV2.S, primary series) | 6 months | Wild-type, Alpha, Delta | Setting | Non-hospitalized | Adjusted for age, race, area deprivation index, outpatient visits, inpatient visits, and unique medication prescriptions in previous year, routine blood chemistry panels, BMI, history of influenza vaccination, smoking status, comorbidities, systolic blood pressure, diastolic blood pressure, estimated glomerular filtration rate, and hospital system average hospital bed occupancy, total hospital bed capacity, positivity rate of tests, number of tests, and HHS region SARS-CoV-2 variant proportion in the week prior to enrolment, geographic region of VISN, and calendar week of enrolment | 7 | 4 | 11 |
| Al-Aly, 2022 (BNT162b2, mRNA-1273 or Ad26.COV2.S, primary series) | 6 months | Wild-type, Alpha, Delta | Setting | Hospitalized | Adjusted for age, race, area deprivation index, outpatient visits, inpatient visits, and unique medication prescriptions in previous year, routine blood chemistry panels, BMI, history of influenza vaccination, smoking status, comorbidities, systolic blood pressure, diastolic blood pressure, estimated glomerular filtration rate, and hospital system average hospital bed occupancy, total hospital bed capacity, positivity rate of tests, number of tests, and HHS region SARS-CoV-2 variant proportion in the week prior to enrolment, geographic region of VISN, and calendar week of enrolment | 12 | 4 | 19 |
| Al-Aly, 2022 (BNT162b2, mRNA-1273 or Ad26.COV2.S, primary series) | 6 months | Wild-type, Alpha, Delta | Setting | ICU | Adjusted for age, race, area deprivation index, outpatient visits, inpatient visits, and unique medication prescriptions in previous year, routine blood chemistry panels, BMI, history of influenza vaccination, smoking status, comorbidities, systolic blood pressure, diastolic blood pressure, estimated glomerular filtration rate, and hospital system average hospital bed occupancy, total hospital bed capacity, positivity rate of tests, number of tests, and HHS region SARS-CoV-2 variant proportion in the week prior to enrolment, geographic region of VISN, and calendar week of enrolment | 22 | 10 | 33 |
| Al-Aly, 2022 (BNT162b2, mRNA-1273 or Ad26.COV2.S, primary series) | 6 months | Wild-type, Alpha, Delta | NA | NA | Adjusted for age, race, area deprivation index, outpatient visits, inpatient visits, and unique medication prescriptions in previous year, routine blood chemistry panels, BMI, history of influenza vaccination, smoking status, comorbidities, systolic blood pressure, diastolic blood pressure, estimated glomerular filtration rate, and hospital system average hospital bed occupancy, total hospital bed capacity, positivity rate of tests, number of tests, and HHS region SARS-CoV-2 variant proportion in the week prior to enrolment, geographic region of VISN, and calendar week of enrolment | 15 | 11 | 18 |
| Di Fusco, 2023 (BNT162b2, 2 doses) | 6 months | Omicron | Number of symptoms | ≥2 symptoms | Adjusted for timepoint, vaccination status and interaction of time by vaccination status, participant pre-COVID-19 symptom onset score, sociodemographic characteristics (age, sex, regions, social vulnerability, race/ethnicity, high risk occupations), previously tested positive for COVID-19, severity of acute illness and immunocompromised status | 36 | -39 | 70 |
| Di Fusco, 2023 (BNT162b2, 2 doses) | 6 months | Omicron | Number of symptoms | ≥3 symptoms | Adjusted for timepoint, vaccination status and interaction of time by vaccination status, participant pre-COVID-19 symptom onset score, sociodemographic characteristics (age, sex, regions, social vulnerability, race/ethnicity, high risk occupations), previously tested positive for COVID-19, severity of acute illness and immunocompromised status | 40 | -34 | 73 |
| Di Fusco, 2023 (BNT162b2, 3 doses) | 6 months | Omicron | Number of symptoms | ≥3 symptoms | Adjusted for timepoint, vaccination status and interaction of time by vaccination status, participant pre-COVID-19 symptom onset score, sociodemographic characteristics (age, sex, regions, social vulnerability, race/ethnicity, high risk occupations), previously tested positive for COVID-19, severity of acute illness and immunocompromised status | 64 | 13 | 85 |
| Di Fusco, 2023 (BNT162b2, 3 doses) | 6 months | Omicron | Number of symptoms | ≥2 symptoms | Adjusted for timepoint, vaccination status and interaction of time by vaccination status, participant pre-COVID-19 symptom onset score, sociodemographic characteristics (age, sex, regions, social vulnerability, race/ethnicity, high risk occupations), previously tested positive for COVID-19, severity of acute illness and immunocompromised status | 70 | 30 | 87 |
| Marra, 2023 (BNT162b2, CoronaVac, Ad26.COV2.S or ChAdOx1-S, 1 dose) | 6 months post-acute infection | Wild-type, Gamma, Delta, Omicron | NA | NA | Adjusted for sex, age, BMI, any comorbidity, the number of SARS-CoV-2 infections, and SARS-CoV-2 variant | 9 | -39 | 40 |
| Marra, 2023 (BNT162b2, CoronaVac, Ad26.COV2.S or ChAdOx1-S, 2 doses) | 6 months post-acute infection | Wild-type, Gamma, Delta, Omicron | NA | NA | Adjusted for sex, age, BMI, any comorbidity, the number of SARS-CoV-2 infections, and SARS-CoV-2 variant | -17 | -76 | 21 |
| Marra, 2023 (BNT162b2, CoronaVac, Ad26.COV2.S or ChAdOx1-S, 3 doses) | 6 months post-acute infection | Wild-type, Gamma, Delta, Omicron | NA | NA | Adjusted for sex, age, BMI, any comorbidity, the number of SARS-CoV-2 infections, and SARS-CoV-2 variant | 37 | -2 | 61 |
| Marra, 2023 (BNT162b2, CoronaVac, Ad26.COV2.S or ChAdOx1-S, 4 doses) | 6 months post-acute infection | Wild-type, Gamma, Delta, Omicron | NA | NA | Adjusted for sex, age, BMI, any comorbidity, the number of SARS-CoV-2 infections, and SARS-CoV-2 variant | 95 | 81 | 99 |
| Luo, 2023 (CoronaVac, BNT162b2 or N, ≥3 doses) | 6-12 months post-acute infection | Omicron | Number of doses | ≥3 doses | Adjusted for age, gender, BMI, infection days, vaccination status, vaccination after infection, comorbidities, infection days, treatment for the acute stage of illness, and patient's satisfaction of treatment | -11 | -24 | 2 |
| Ertesvag, 2023 (BNT162b2 or mRNA-1273, ≥1 dose) | 8 months post-infection | Delta | Timepoint and population | Vaccination as a predictor of any persisting symptoms among adolescents (≥16 years) 8-months post-Delta infection | Adjusted for female sex, age, reinfection, and any symptoms at baseline | -98 | -457 | 30 |
| de Arriba Fernández, 2023 (BNT162b2, mRNA-1273, Ad26.COV2.S or ChAdOx1-S, 1 dose) | NR | Delta, Omicron | NA | NA | Adjusted for age, sex, comorbidity history, body mass index, date of first, second and booster doses of a COVID-19 vaccine, type of COVID-19 vaccine, death, immunosuppressive treatment, diagnosis of persistent COVID-19, and diagnostic test results | 82 | 71 | 89 |

BMI: Body mass index; CI: Confidence interval; CDC: Centers for Disease Control and Prevention; CPRD: Clinical Practice Research Datalink; GP: General Practitioner; HHS: Health and Human Services; HR: Hazard ratio; ICD-10: International Classification of Diseases- 10; ICU: Intensive care units; IPTW: Inverse probability of treatment weighting; IQR: Interquartile range; mRNA: Messenger ribonucleic acid; NA: Not applicable; NR: Not reported; OR: Odds ratio; PASC: Post-acute sequelae of COVID-19; PCR: Polymerase Chain Reaction; VA: Veterans Affairs; VE: Vaccine effectiveness; VISN: Veterans Integrated Services Network; WHO: World Health Organization; WT: Wild-type.

**eFigure 4. Adjusted pre-infection aVE against ≥1 PCC (vs. unvaccinated) by proportion of sample hospitalized during acute infection**

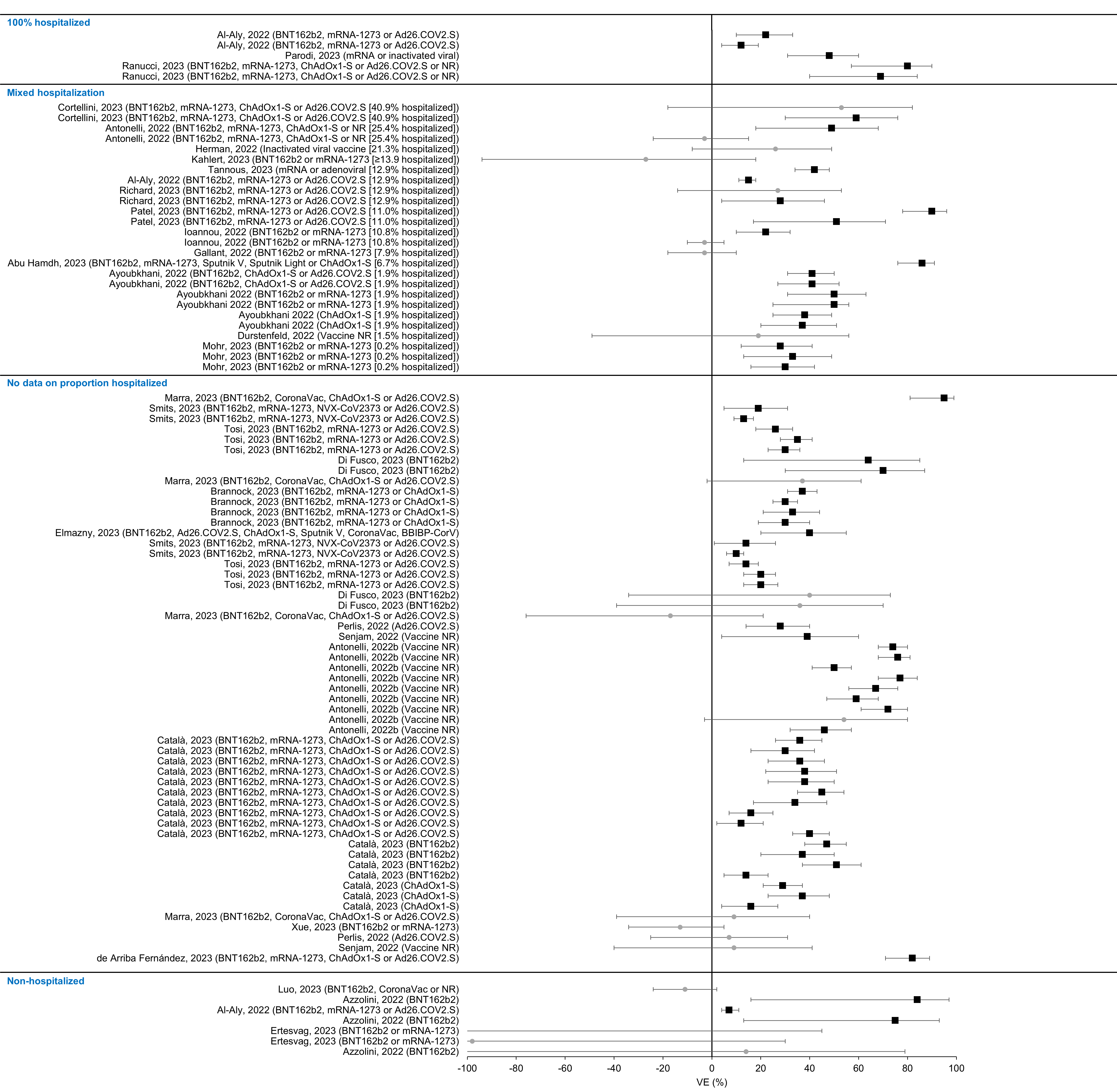

Of 93 VE estimates, 72 (77.4%) reported statistically significant (based on VE 95% CI) protective effect against ≥1 PCC.

CI: confidence interval; NR: not reported; PCC: post‑COVID‑19 conditions; VE: vaccine effectiveness.

References: Abu Hamdh, 2023^1^; Al-Aly, 2022^2^; Antonelli, 2022^6^; Antonelli, 2022b^7^; Ayoubkhani 2022b^12^; Azzolini, 2022^13^; Brannock, 2023^19^; Català, 2023^22^; Cortellini, 2023^25^; de Arriba Fernández, 2023^27^; Di Fusco, 2023^29^; Durstenfeld, 2022^30^; Elmazny, 2023^33^; Ertesvag, 2023^36^; Gallant, 2022^39^; Herman, 2022^43^; Ioannou, 2022^45^; Kahlert, 2023^49^; Luo, 2023^56^; Marra, 2023^57^; Mohr, 2023^62^; Parodi, 2023^68^; Patel, 2023^69^; Perlis, 2022^72^; Ranucci, 2023^76^; Richard, 2023^78^; Senjam, 2022^81^; Smits, 2023^82^; Tannous, 2023^84^; Tosi, 2023^86^; Xue, 2023^95^

**eTable 19. Summary of publications** **where adjusted pre-infection VE against ≥1 PCC (vs. unvaccinated) could be computed by proportion of sample hospitalized during acute infection (N=31 studies with 93 VE estimates)**

| **Author, year (vaccine and dose)** | **Proportion of sample hospitalized during acute infection** | **Variant** | **Subgroup** | **Subgroup details** | **Adjustments** | **VE** | **Lower 95% CI** | **Upper 95% CI** |
| --- | --- | --- | --- | --- | --- | --- | --- | --- |
| Luo, 2023 (BNT162b2, CoronaVac or NR, ≥3 doses) | 100% non-hospitalized | Omicron | NA | NA | Adjusted for age, gender, BMI, infection days, vaccination status, vaccination after infection, comorbidities, infection days, treatment for the acute stage of illness, and patient's satisfaction of treatment | -11 | -24 | 2 |
| Azzolini, 2022 (BNT162b2, 1 dose) | 100% non-hospitalized | Wild-type, Alpha, Delta, Omicron | NA | NA | Adjusted for sex, vaccine dose, allergies and comorbidities | 14 | -249 | 79 |
| Ertesvag, 2023 (BNT162b2 or mRNA-1273, ≥1 dose) | 100% non-hospitalized | Delta | Timepoint and population | Vaccination as a predictor of any persisting symptoms among adolescents (≥16 years) 8-months post-Delta infection | Adjusted for female sex, age, reinfection, and any symptoms at baseline | -98 | -457 | 30 |
| Ertesvag, 2023 (BNT162b2 or mRNA-1273, ≥1 dose) | 100% non-hospitalized | Delta | Timepoint | Vaccination as a predictor of any persisting symptoms 3-months post-Delta infection | Adjusted by female sex, age, reinfection, any symptoms at baseline. | -123 | -904 | 45 |
| Azzolini, 2022 (BNT162b2, 2 doses) | 100% non-hospitalized | Wild-type, Alpha, Delta, Omicron | NA | NA | Adjusted for sex, vaccine dose, allergies and comorbidities | 75 | 13 | 93 |
| Al-Aly, 2022 (BNT162b2, mRNA-1273 or Ad26.COV2.S, Primary series) | 100% non-hospitalized | Wild-type, Alpha, Delta | Setting | Non-hospitalized | Adjusted for age, race, area deprivation index, outpatient visits, inpatient visits, and unique medication prescriptions in previous year, routine blood chemistry panels, BMI, history of influenza vaccination, smoking status, comorbidities, systolic blood pressure, diastolic blood pressure, estimated glomerular filtration rate, and hospital system average hospital bed occupancy, total hospital bed capacity, positivity rate of tests, number of tests, and HHS region SARS-CoV-2 variant proportion in the week prior to enrolment, geographic region of VISN, and calendar week of enrolment | 7 | 4 | 11 |
| Azzolini, 2022 (BNT162b2, 3 doses) | 100% non-hospitalized | Wild-type, Alpha, Delta, Omicron | NA | NA | Adjusted for sex, vaccine dose, allergies and comorbidities | 84 | 16 | 97 |
| de Arriba Fernández, 2023 (BNT162b2, mRNA-1273, ChAdOx1-S or Ad26.COV2.S, 1 dose) | No data on proportion hospitalized | Delta, Omicron | NA | NA | Adjusted for age, sex, comorbidity history, body mass index, date of first, second and booster doses of a COVID-19 vaccine, type of COVID-19 vaccine, death, immunosuppressive treatment, diagnosis of persistent COVID-19, and diagnostic test results | 82 | 71 | 89 |
| Senjam, 2022 (Vaccine NR, 1 dose) | Hospitalization NR (no data on proportion hospitalized) | Delta | NA | NA | Adjusted for age, gender, educational level, BMI, workplace, blood group, smoking, alcohol consumption, presence of comorbidities, oxygen requirement, severity of COVID-19 disease, overall health status following COVUD-19 illness | 9 | -40 | 41 |
| Perlis, 2022 (Ad26.COV2.S, 1 dose) | Hospitalization NR (no data on proportion hospitalized) | Alpha, Delta, Omicron | NA | NA | Adjusted for age, gender, race and ethnicity, educational level, urbanicity, region and predominant variant | 7 | -25 | 31 |
| Xue, 2023 (BNT162b2 or mRNA-1273, 1 dose) | Hospitalization NR (no data on proportion hospitalized) | Alpha, Gamma, Delta, Omicron | NA | NA | Adjusted for age, gender, ethnicity, smoking status, alcohol consumption, educational level, since first vaccination, martial status, living area, level of physical activity, weight status, comorbidities. | -13 | -34 | 5 |
| Marra, 2023 (BNT162b2, CoronaVac, ChAdOx1-S or Ad26.COV2.S, 1 dose) | Hospitalization NR (no data on proportion hospitalized) | Wild-type, Gamma, Delta, Omicron | NA | NA | Adjusted for sex, age, BMI, any comorbidity, the number of SARS-CoV-2 infections, and SARS-CoV-2 variant | 9 | -39 | 40 |
| Català, 2023 (ChAdOx1-S, 1 dose) | Hospitalization NR (no data on proportion hospitalized) | Delta, Omicron | Vaccination group | SIDIAP (AstraZeneca vaccine) | Propensity score overlap weighting for age (5-year age bands for people aged ≥18 years), location, index date, previous observation years, number of previous GP visits, number of previous SARS-CoV-2 PCR tests, regional vaccination, testing | 16 | 4 | 27 |
| Català, 2023 (ChAdOx1-S, 1 dose) | Hospitalization NR (no data on proportion hospitalized) | Delta, Omicron | Vaccination group | CPRD GOLD (AstraZeneca vaccine) | Propensity score overlap weighting for age (5-year age bands for people aged ≥18 years), location, index date, previous observation years, number of previous GP visits, number of previous SARS-CoV-2 PCR tests, regional vaccination, testing, COVID-19 incidence rates | 37 | 23 | 48 |
| Català, 2023 (ChAdOx1-S, 1 dose) | Hospitalization NR (no data on proportion hospitalized) | Delta, Omicron | Vaccination group | CPRD AURUM (AstraZeneca vaccine) | Propensity score overlap weighting for age (5-year age bands for people aged ≥18 years), location, index date, previous observation years, number of previous GP visits, number of previous SARS-CoV-2 PCR tests, regional vaccination, testing, COVID-19 incidence rates | 29 | 21 | 37 |
| Català, 2023 (BNT162b2, 1 dose) | Hospitalization NR (no data on proportion hospitalized) | Delta, Omicron | Vaccination group | SIDIAP (Pfizer vaccine) | Propensity score overlap weighting for age (5-year age bands for people aged ≥18 years), location, index date, previous observation years, number of previous GP visits, number of previous SARS-CoV-2 PCR tests, regional vaccination, testing | 14 | 5 | 23 |
| Català, 2023 (BNT162b2, 1 dose) | Hospitalization NR (no data on proportion hospitalized) | Delta, Omicron | Vaccination group | CPRD GOLD (Pfizer vaccine) | Propensity score overlap weighting for age (5-year age bands for people aged ≥18 years), location, index date, previous observation years, number of previous GP visits, number of previous SARS-CoV-2 PCR tests, regional vaccination, testing, COVID-19 incidence rates | 51 | 37 | 61 |
| Català, 2023 (BNT162b2, 1 dose) | Hospitalization NR (no data on proportion hospitalized) | Delta, Omicron | Vaccination group | CORIVA (Pfizer vaccine) | Propensity score overlap weighting for age (5-year age bands for people aged ≥18 years), location, index date, previous observation years, number of previous GP visits, number of previous SARS-CoV-2 PCR tests, regional vaccination, testing | 37 | 20 | 50 |
| Català, 2023 (BNT162b2, 1 dose) | Hospitalization NR (no data on proportion hospitalized) | Delta, Omicron | Vaccination group | CPRD AURUM (Pfizer vaccine) | Propensity score overlap weighting for age (5-year age bands for people aged ≥18 years), location, index date, previous observation years, number of previous GP visits, number of previous SARS-CoV-2 PCR tests, regional vaccination, testing, COVID-19 incidence rates | 47 | 38 | 55 |
| Català, 2023 (BNT162b2, mRNA-1273, ChAdOx1-S or Ad26.COV2.S, 1 dose) | Hospitalization NR (no data on proportion hospitalized) | Delta, Omicron | Vaccination group | SIDIAP, GP diagnosis | Propensity score overlap weighting for age (5-year age bands for people aged ≥18 years), location, index date, previous observation years, number of previous GP visits, number of previous SARS-CoV-2 PCR tests, regional vaccination, testing | 40 | 33 | 48 |
| Català, 2023 (BNT162b2, mRNA-1273, ChAdOx1-S or Ad26.COV2.S, 1 dose) | Hospitalization NR (no data on proportion hospitalized) | Delta, Omicron | Vaccination group | SIDIAP, ≥28 days | Propensity score overlap weighting for age (5-year age bands for people aged ≥18 years), location, index date, previous observation years, number of previous GP visits, number of previous SARS-CoV-2 PCR tests, regional vaccination, testing | 12 | 2 | 21 |
| Català, 2023 (BNT162b2, mRNA-1273, ChAdOx1-S or Ad26.COV2.S, 1 dose) | Hospitalization NR (no data on proportion hospitalized) | Delta, Omicron | Vaccination group | SIDIAP | Propensity score overlap weighting for age (5-year age bands for people aged ≥18 years), location, index date, previous observation years, number of previous GP visits, number of previous SARS-CoV-2 PCR tests, regional vaccination, testing | 16 | 7 | 25 |
| Català, 2023 (BNT162b2, mRNA-1273, ChAdOx1-S or Ad26.COV2.S, 1 dose) | Hospitalization NR (no data on proportion hospitalized) | Delta, Omicron | Vaccination group | CPRD GOLD, ≥28 days | Propensity score overlap weighting for age (5-year age bands for people aged ≥18 years), location, index date, previous observation years, number of previous GP visits, number of previous SARS-CoV-2 PCR tests, regional vaccination, testing, COVID-19 incidence rates | 34 | 17 | 47 |
| Català, 2023 (BNT162b2, mRNA-1273, ChAdOx1-S or Ad26.COV2.S, 1 dose) | Hospitalization NR (no data on proportion hospitalized) | Delta, Omicron | Cohort | CPRD GOLD | Propensity score overlap weighting for age (5-year age bands for people aged ≥18 years), location, index date, previous observation years, number of previous GP visits, number of previous SARS-CoV-2 PCR tests, regional vaccination, testing, COVID-19 incidence rates | 45 | 35 | 54 |
| Català, 2023 (BNT162b2, mRNA-1273, ChAdOx1-S or Ad26.COV2.S, 1 dose) | Hospitalization NR (no data on proportion hospitalized) | Delta, Omicron | Vaccination group | CORIVA, ≥28 days | Propensity score overlap weighting for age (5-year age bands for people aged ≥18 years), location, index date, previous observation years, number of previous GP visits, number of previous SARS-CoV-2 PCR tests, regional vaccination, testing | 38 | 23 | 50 |
| Català, 2023 (BNT162b2, mRNA-1273, ChAdOx1-S or Ad26.COV2.S, 1 dose) | Hospitalization NR (no data on proportion hospitalized) | Delta, Omicron | Vaccination group | CORIVA | Propensity score overlap weighting for age (5-year age bands for people aged ≥18 years), location, index date, previous observation years, number of previous GP visits, number of previous SARS-CoV-2 PCR tests, regional vaccination, testing | 38 | 22 | 51 |
| Català, 2023 (BNT162b2, mRNA-1273, ChAdOx1-S or Ad26.COV2.S, 1 dose) | Hospitalization NR (no data on proportion hospitalized) | Delta, Omicron | Vaccination group | CPRD AURUM, GP diagnosis | Propensity score overlap weighting for age (5-year age bands for people aged ≥18 years), location, index date, previous observation years, number of previous GP visits, number of previous SARS-CoV-2 PCR tests, regional vaccination, testing, COVID-19 incidence rates | 36 | 23 | 46 |
| Català, 2023 (BNT162b2, mRNA-1273, ChAdOx1-S or Ad26.COV2.S, 1 dose) | Hospitalization NR (no data on proportion hospitalized) | Delta, Omicron | Vaccination group | CPRD AURUM, ≥28 days | Propensity score overlap weighting for age (5-year age bands for people aged ≥18 years), location, index date, previous observation years, number of previous GP visits, number of previous SARS-CoV-2 PCR tests, regional vaccination, testing, COVID-19 incidence rates | 30 | 16 | 42 |
| Català, 2023 (BNT162b2, mRNA-1273, ChAdOx1-S or Ad26.COV2.S, 1 dose) | Hospitalization NR (no data on proportion hospitalized) | Delta, Omicron | Vaccination group | CPRD AURUM | Propensity score overlap weighting for age (5-year age bands for people aged ≥18 years), location, index date, previous observation years, number of previous GP visits, number of previous SARS-CoV-2 PCR tests, regional vaccination, testing, COVID-19 incidence rates | 36 | 26 | 45 |
| Antonelli, 2022b (Vaccine NR, ≥1 dose) | Hospitalization NR (no data on proportion hospitalized) | Delta, Omicron | Age; Time since vaccination | ≥60; vaccinated <3 months prior to infection | Adjusted by age, sex, BMI, index of multiple deprivation, presence of comorbidities and vaccination status | 46 | 32 | 57 |
| Antonelli, 2022b (Vaccine NR, ≥1 dose) | Hospitalization NR (no data on proportion hospitalized) | Delta, Omicron | Age; Time since vaccination | ≥60; vaccinated 3–6 months prior to infection | Adjusted by age, sex, BMI, index of multiple deprivation, presence of comorbidities and vaccination status | 54 | -3 | 80 |
| Antonelli, 2022b (Vaccine NR, ≥1 dose) | Hospitalization NR (no data on proportion hospitalized) | Delta, Omicron | Age; Time since vaccination | ≥60; vaccinated >6 months prior to infection | Adjusted by age, sex, BMI, index of multiple deprivation, presence of comorbidities and vaccination status | 72 | 61 | 80 |
| Antonelli, 2022b (Vaccine NR, ≥1 dose) | Hospitalization NR (no data on proportion hospitalized) | Delta, Omicron | Age; Time since vaccination | 18–59; vaccinated <3 months prior to infection | Adjusted by age, sex, BMI, index of multiple deprivation, presence of comorbidities and vaccination status | 59 | 47 | 68 |
| Antonelli, 2022b (Vaccine NR, ≥1 dose) | Hospitalization NR (no data on proportion hospitalized) | Delta, Omicron | Age; Time since vaccination | 18–59; vaccinated 3–6 months prior to infection | Adjusted by age, sex, BMI, index of multiple deprivation, presence of comorbidities and vaccination status | 67 | 56 | 76 |
| Antonelli, 2022b (Vaccine NR, ≥1 dose) | Hospitalization NR (no data on proportion hospitalized) | Delta, Omicron | Age; Time since vaccination | 18–59; vaccinated >6 months prior to infection | Adjusted by age, sex, BMI, index of multiple deprivation, presence of comorbidities and vaccination status | 77 | 68 | 84 |
| Antonelli, 2022b (Vaccine NR, ≥1 dose) | Hospitalization NR (no data on proportion hospitalized) | Delta, Omicron | Time since vaccination | Vaccinated <3 months prior to infection | Adjusted by age, sex, BMI, index of multiple deprivation, presence of comorbidities and vaccination status | 50 | 41 | 57 |
| Antonelli, 2022b (Vaccine NR, ≥1 dose) | Hospitalization NR (no data on proportion hospitalized) | Delta, Omicron | Time since vaccination | Vaccinated 3–6 months prior to infection | Adjusted by age, sex, BMI, index of multiple deprivation, presence of comorbidities and vaccination status | 76 | 68 | 81 |
| Antonelli, 2022b (Vaccine NR, ≥1 dose) | Hospitalization NR (no data on proportion hospitalized) | Delta, Omicron | Time since vaccination | Vaccinated >6 months prior to infection | Adjusted by age, sex, BMI, index of multiple deprivation, presence of comorbidities and vaccination status | 74 | 68 | 80 |
| Senjam, 2022 (Vaccine NR, 2 doses) | Hospitalization NR (no data on proportion hospitalized) | Delta | NA | NA | Adjusted for severity of COVID-19, chronic medication use, presence of chronic conditions, self-perceived health status, monthly household income, smoking status, living status, educational level, current occupation, marital status, gender and age | 39 | 4 | 60 |
| Perlis, 2022 (Ad26.COV2.S, 2 doses) | Hospitalization NR (no data on proportion hospitalized) | Alpha, Delta, Omicron | NA | NA | Adjusted for age, gender, race and ethnicity, educational level, urbanicity, region and predominant variant | 28 | 14 | 40 |
| Marra, 2023 (BNT162b2, CoronaVac, ChAdOx1-S or Ad26.COV2.S, 2 doses) | Hospitalization NR (no data on proportion hospitalized) | Wild-type, Gamma, Delta, Omicron | NA | NA | Adjusted for sex, age, BMI, any comorbidity, the number of SARS-CoV-2 infections, and SARS-CoV-2 variant | -17 | -76 | 21 |
| Di Fusco, 2023 (BNT162b2, 2 doses) | Hospitalization NR (no data on proportion hospitalized) | Omicron | Number of symptoms | ≥2 symptoms | Adjusted for timepoint, vaccination status and interaction of time by vaccination status, participant pre-COVID-19 symptom onset score, sociodemographic characteristics (age, sex, regions, social vulnerability, race/ethnicity, high risk occupations), previously tested positive for COVID-19, severity of acute illness and immunocompromised status | 36 | -39 | 70 |
| Di Fusco, 2023 (BNT162b2, 2 doses) | Hospitalization NR (no data on proportion hospitalized) | Omicron | Number of symptoms | ≥3 symptoms | Adjusted for timepoint, vaccination status and interaction of time by vaccination status, participant pre-COVID-19 symptom onset score, sociodemographic characteristics (age, sex, regions, social vulnerability, race/ethnicity, high risk occupations), previously tested positive for COVID-19, severity of acute illness and immunocompromised status | 40 | -34 | 73 |
| Tosi, 2023 (BNT162b2, mRNA-1273 or Ad26.COV2.S, Primary series) | Hospitalization NR (no data on proportion hospitalized) | Omicron | Vaccination, patient frailty | Vaccinated, frail patient | Adjusted for age, BMI, race, ethnicity, smoking, active patient in the past 24 months, and rurality | 20 | 13 | 27 |
| Tosi, 2023 (BNT162b2, mRNA-1273 or Ad26.COV2.S, Primary series) | Hospitalization NR (no data on proportion hospitalized) | Omicron | Vaccination, patient frailty | Vaccinated, pre-frail patient | Adjusted for age, BMI, race, ethnicity, smoking, active patient in the past 24 months, and rurality | 20 | 13 | 26 |
| Tosi, 2023 (BNT162b2, mRNA-1273 or Ad26.COV2.S, Primary series) | Hospitalization NR (no data on proportion hospitalized) | Omicron | Vaccination, patient frailty | Vaccinated, robust patient | Adjusted for age, BMI, race, ethnicity, smoking, active patient in the past 24 months, and rurality | 14 | 7 | 19 |
| Smits, 2023 (BNT162b2, mRNA-1273, NVX-CoV2373 or Ad26.COV2.S, Primary series) | Hospitalization NR (no data on proportion hospitalized) | Omicron | Vaccination status | Long-COVID symptoms (28-365 days post-positive COVID-19 test) | Adjusted for vaccination status, comorbidities, sex, race, ethnicity, age, timing of infection, smoking status, influenza vaccination status, inpatient and outpatient encounters in previous two years | 10 | 6 | 13 |
| Smits, 2023 (BNT162b2, mRNA-1273, NVX-CoV2373 or Ad26.COV2.S, Primary series) | Hospitalization NR (no data on proportion hospitalized) | Omicron | Vaccination status | Long COVID diagnosis (ICD-10 codes; 28–365 days post-positive COVID-19 test) | Adjusted for vaccination status, comorbidities, sex, race, ethnicity, age, timing of infection, smoking status, influenza vaccination status, inpatient and outpatient encounters in previous two years | 14 | 1 | 26 |
| Elmazny, 2023 (BNT162b2, Ad26.COV2.S, ChAdOx1-S, Sputnik V, CoronaVac, BBIBP-CorV, Primary series) | Hospitalization NR (no data on proportion hospitalized) | WT, Alpha, Delta, Omicron | NA | NA | Adjusted for age, sex, and COVID-19 severity | 40 | 20 | 55 |
| Brannock, 2023 (BNT162b2, mRNA-1273 or ChAdOx1-S, Primary series) | Hospitalization NR (no data on proportion hospitalized) | Delta, Omicron | Methodology - logistic regression analysis | Clinic-based cohort IPTW-adjusted OR | Inverse probability of treatment weighting applied | 30 | 19 | 40 |
| Brannock, 2023 (BNT162b2, mRNA-1273 or ChAdOx1-S, Primary series) | Hospitalization NR (no data on proportion hospitalized) | Delta, Omicron | Methodology - proportional hazards | Clinic-based cohort IPTW-adjusted HR | Inverse probability of treatment weighting applied | 33 | 21 | 44 |
| Brannock, 2023 (BNT162b2, mRNA-1273 or ChAdOx1-S, Primary series) | Hospitalization NR (no data on proportion hospitalized) | Delta, Omicron | Methodology - logistic regression analysis | Model-based cohort IPTW-adjusted OR | Inverse probability of treatment weighting applied | 30 | 25 | 35 |
| Brannock, 2023 (BNT162b2, mRNA-1273 or ChAdOx1-S, Primary series) | Hospitalization NR (no data on proportion hospitalized) | Delta, Omicron | Methodology - proportional hazards | Model-based cohort IPTW-adjusted HR | Inverse probability of treatment weighting applied | 37 | 31 | 43 |
| Marra, 2023 (BNT162b2, CoronaVac, ChAdOx1-S or Ad26.COV2.S, 3 doses) | Hospitalization NR (no data on proportion hospitalized) | Wild-type, Gamma, Delta, Omicron | NA | NA | Adjusted for sex, age, BMI, any comorbidity, the number of SARS-CoV-2 infections, and SARS-CoV-2 variant | 37 | -2 | 61 |
| Di Fusco, 2023 (BNT162b2, 3 doses) | Hospitalization NR (no data on proportion hospitalized) | Omicron | Number of symptoms | ≥2 symptoms | Adjusted for timepoint, vaccination status and interaction of time by vaccination status, participant pre-COVID-19 symptom onset score, sociodemographic characteristics (age, sex, regions, social vulnerability, race/ethnicity, high risk occupations), previously tested positive for COVID-19, severity of acute illness and immunocompromised status | 70 | 30 | 87 |
| Di Fusco, 2023 (BNT162b2, 3 doses) | Hospitalization NR (no data on proportion hospitalized) | Omicron | Number of symptoms | ≥3 symptoms | Adjusted for timepoint, vaccination status and interaction of time by vaccination status, participant pre-COVID-19 symptom onset score, sociodemographic characteristics (age, sex, regions, social vulnerability, race/ethnicity, high risk occupations), previously tested positive for COVID-19, severity of acute illness and immunocompromised status | 64 | 13 | 85 |
| Tosi, 2023 (BNT162b2, mRNA-1273 or Ad26.COV2.S) | Hospitalization NR (no data on proportion hospitalized) | Omicron | Vaccination, patient frailty | Boosted, frail patient | Adjusted for age, BMI, race, ethnicity, smoking, active patient in the past 24 months, and rurality | 30 | 23 | 36 |
| Tosi, 2023 (BNT162b2, mRNA-1273 or Ad26.COV2.S) | Hospitalization NR (no data on proportion hospitalized) | Omicron | Vaccination, patient frailty | Boosted, pre-frail patient | Adjusted for age, BMI, race, ethnicity, smoking, active patient in the past 24 months, and rurality | 35 | 28 | 41 |
| Tosi, 2023 (BNT162b2, mRNA-1273 or Ad26.COV2.S) | Hospitalization NR (no data on proportion hospitalized) | Omicron | Vaccination, patient frailty | Boosted, robust patient | Adjusted for age, BMI, race, ethnicity, smoking, active patient in the past 24 months, and rurality | 26 | 18 | 33 |
| Smits, 2023 (BNT162b2, mRNA-1273, NVX-CoV2373 or Ad26.COV2.S, Primary series) | Hospitalization NR (no data on proportion hospitalized) | Omicron | Vaccination status | Long-COVID symptoms (28-365 days post-positive COVID-19 test) | Adjusted for vaccination status, comorbidities, sex, race, ethnicity, age, timing of infection, smoking status, influenza vaccination status, inpatient and outpatient encounters in previous two years | 13 | 9 | 17 |
| Smits, 2023 (BNT162b2, mRNA-1273, NVX-CoV2373 or Ad26.COV2.S, Primary series) | Hospitalization NR (no data on proportion hospitalized) | Omicron | Vaccination status | Long COVID diagnosis (ICD-10 codes; 28-365 days post-positive COVID-19 test) | Adjusted for vaccination status, comorbidities, sex, race, ethnicity, age, timing of infection, smoking status, influenza vaccination status, inpatient and outpatient encounters in previous two years | 19 | 5 | 31 |
| Marra, 2023 (BNT162b2, CoronaVac, ChAdOx1-S or Ad26.COV2.S, 4 doses) | Hospitalization NR (no data on proportion hospitalized) | Wild-type, Gamma, Delta, Omicron | NA | NA | Adjusted for sex, age, BMI, any comorbidity, the number of SARS-CoV-2 infections, and SARS-CoV-2 variant | 95 | 81 | 99 |
| Ioannou, 2022 (BNT162b2 or mRNA-1273, 1 dose) | Mixed (10.8%) | Wild-type, Alpha | NA | NA | Adjusted for age, sex, race, ethnicity, urban vs rural residence, Charlson Comorbidity Index score, VA Integrated Service Network, time period of infection/pandemic wave, number of primary care, mental health and specialty care encounters in the 2 years before infection | -3 | -10 | 5 |
| Antonelli, 2022 (BNT162b2, mRNA-1273, ChAdOx1-S or NR, 1 dose) | Mixed (25.4%) | Alpha, Delta | NA | NA | Adjusted for age, BMI, sex | -3 | -24 | 15 |
| Abu Hamdh, 2023 (BNT162b2, mRNA-1273, Sputnik V, Sputnik Light or ChAdOx1-S, ≥1 dose) | Mixed (6.7%) | NR | NA | NA | Adjusted for sex, age, comorbidities, hospital admission, dyspnoea, previous COVID-19 infection, fatigue | 86 | 76 | 91 |
| Durstenfeld, 2022 (Vaccine NR, ≥1 dose) | Mixed (1.5%) | Wild-type, Alpha, Delta, Omicron | NA | NA | Adjusted for age, COVID-19 wave, number of initial symptoms, sex, myocardial infarction, stroke, atrial fibrillation, sleep apnoea, COPD or asthma, and immunodeficiency, plus receipt of COVID-19 vaccination before or after COVID-19 diagnosis, ethnicity, subjective socioeconomic status, highest level of education, primary employment, pre-COVID depressive symptoms, anxiety symptoms, and financial security | 19.0 | -49 | 56 |
| Kahlert, 2023 (BNT162b2 or mRNA-1273, ≥1 dose) | Mixed (specific % NR, at least 13.9%) | Wild-type, Alpha, Delta, Omicron | NA | NA | Adjusted for age, sex, BMI, ethnicity, children in household, comorbidity, medication, smoker status, alcohol intake, medical profession, COVID-19 patient contact, ICU work, full-time work, SARS-CoV-2 variant | -27 | -94 | 18 |
| Mohr, 2023 (BNT162b2 or mRNA-1273, 2 doses) | Mixed (0.2% required hospitalization) | Wild-type, Alpha, Delta | Timepoint | 6 weeks | Adjusted for age, race and ethnicity, comorbidities, calendar month of diagnosis and weeks since symptoms started | 30 | 16 | 42 |
| Mohr, 2023 (BNT162b2 or mRNA-1273, 2 doses) | Mixed (0.2% required hospitalization) | Wild-type, Alpha, Delta | Timepoint | ≤16 weeks since vaccination | Adjusted for age, race and ethnicity, comorbidities, calendar month of diagnosis, and weeks since symptoms started | 33 | 13 | 49 |
| Mohr, 2023 (BNT162b2 or mRNA-1273, 2 doses) | Mixed (0.2% required hospitalization) | Wild-type, Alpha, Delta | Timepoint | >16 weeks since vaccination | Adjusted for age, race and ethnicity, comorbidities, calendar month of diagnosis, and weeks since symptoms started | 28 | 12 | 41 |
| Ayoubkhani 2022 (ChAdOx1-S, 2 doses) | Mixed (1.9%) | Delta | Vaccine and long COVID severity | AstraZeneca; activity limiting Long COVID | Adjusted for age, sex, white or non-white ethnicity, country/region of residence, area deprivation quintile group, and self-reported, pre-existing health/disability status and time from infection to follow-up for Long Covid | 37 | 20 | 51 |
| Ayoubkhani 2022 (ChAdOx1-S, 2 doses) | Mixed (1.9%) | Delta | Vaccine and long COVID severity | AstraZeneca; Long COVID of any severity | Adjusted for age, sex, white or non-white ethnicity, country/region of residence, area deprivation quintile group, and self-reported, pre-existing health/disability status and time from infection to follow-up for Long Covid | 38 | 25 | 49 |
| Ayoubkhani 2022 (BNT162b2 or mRNA-1273, 2 doses) | Mixed (1.9%) | Delta | Vaccine and long COVID severity | mRNA vaccine platform; activity-limiting Long COVID | Adjusted for age, sex, white or non-white ethnicity, country/region of residence, area deprivation quintile group, and self-reported, pre-existing health/disability status and time from infection to follow-up for Long Covid | 50 | 25 | 56 |
| Ayoubkhani 2022 (BNT162b2 or mRNA-1273, 2 doses) | Mixed (1.9%) | Delta | Vaccine and long COVID severity | mRNA vaccine platform; Long COVID of any severity | Adjusted for age, sex, white or non-white ethnicity, country/region of residence, area deprivation quintile group, and self-reported, pre-existing health/disability status and time from infection to follow-up for Long Covid | 50 | 31 | 63 |
| Ayoubkhani, 2022 (BNT162b2, ChAdOx1-S or Ad26.COV2.S, 2 doses) | Mixed (1.9%) | Delta | Vaccine and long COVID severity | Adenoviral and mRNA; activity-limiting Long COVID | Adjusted for age, sex, white or non-white ethnicity, country/region of residence, area deprivation quintile group, and self-reported, pre-existing health/disability status and time from infection to follow-up for Long Covid | 41 | 27 | 52 |
| Ayoubkhani, 2022 (BNT162b2, ChAdOx1-S or Ad26.COV2.S, 2 doses) | Mixed (1.9%) | Delta | Vaccine and long COVID severity | Adenoviral and mRNA; Long COVID of any severity | Adjusted for age, sex, white or non-white ethnicity, country/region of residence, area deprivation quintile group, and self-reported, pre-existing health/disability status and time from infection to follow-up for Long Covid | 41 | 31 | 50 |
| Ioannou, 2022 (BNT162b2 or mRNA-1273, 2 doses) | Mixed (10.8%) | Wild-type, Alpha | NA | NA | Adjusted for age, sex, race, ethnicity, urban vs rural residence, Charlson Comorbidity Index score, VA Integrated Service Network, time period of infection/pandemic wave, number of primary care, mental health and specialty care encounters in the 2 years before infection | 22 | 10 | 32 |
| Herman, 2022 (inactivated viral vaccine, 2 doses) | Mixed (21.3% hospitalized) | Delta, Omicron | NA | NA | Adjusted by shortness of breath | 26 | -8 | 49 |
| Antonelli, 2022 (BNT162b2, mRNA-1273, ChAdOx1-S or NR, 2 doses) | Mixed (25.4%) | Alpha, Delta | NA | NA | Adjusted for age, BMI, sex | 49 | 18 | 68 |
| Cortellini, 2023 (BNT162b2, mRNA-1273, ChAdOx1-S or Ad26.COV2.S, 2 doses) | Mixed (40.9%) | Alpha, Delta, Omicron | NA | NA | Inverse probability of treatment weighting applied | 59 | 30 | 76 |
| Patel, 2023 (BNT162b2, mRNA-1273 or Ad26.COV2.S, Primary series) | Mixed (11%) | Delta, Omicron | PASC definition | CDC; ≥28 days | Adjusted for age, sex, race, comorbidity count and use of any one of the following medications: anti-CD20 monoclonal antibodies, methotrexate, mycophenolate or glucocorticoids. | 51 | 17 | 71 |
| Patel, 2023 (BNT162b2, mRNA-1273 or Ad26.COV2.S, Primary series) | Mixed (11%) | Delta, Omicron | PASC definition | WHO; ≥90 days post-infection | Adjusted for age, sex, race, comorbidity count and use of any one of the following medications: anti-CD20 monoclonal antibodies, methotrexate, mycophenolate or glucocorticoids. | 90 | 78 | 96 |
| Al-Aly, 2022 (BNT162b2, mRNA-1273 or Ad26.COV2.S, Primary series) | Mixed (12.9% hospitalized) | Wild-type, Alpha, Delta | NA | Risk of incident post-acute sequelae | Adjusted for age, race, area deprivation index, outpatient visits, inpatient visits, and unique medication prescriptions in previous year, routine blood chemistry panels, BMI, history of influenza vaccination, smoking status, comorbidities, systolic blood pressure, diastolic blood pressure, estimated glomerular filtration rate, and hospital system average hospital bed occupancy, total hospital bed capacity, positivity rate of tests, number of tests, and HHS region SARS-CoV-2 variant proportion in the week prior to enrolment, geographic region of VISN, and calendar week of enrolment | 15 | 11 | 18 |
| Tannous, 2023 (mRNA or adenoviral, Primary series) | Mixed (12.9%) | Wild-type, Alpha, Delta | NA | NA | Adjusted for age, sex, race, ethnicity, Charlson Comorbidity Index, COVID-19 illness severity, and Texas state area deprivation index | 42 | 34 | 48 |
| Richard, 2023 (BNT162b2, mRNA-1273 or Ad26.COV2.S, Primary series) | Mixed (12.9%) | Wild-type, Alpha, Delta | Timepoint | ≥28 days | Adjusted for Delta variant period, maximum symptom severity during acute illness, days hospitalized for COVID-19, comorbidities, acute respiratory symptoms, age, sex and BMI category | 28 | 4 | 46 |
| Richard, 2023 (BNT162b2, mRNA-1273 or Ad26.COV2.S, Primary series) | Mixed (12.9%) | Wild-type, Alpha, Delta | Timepoint | ≥90 days | Adjusted for Delta variant period, maximum symptom severity during acute illness, days hospitalized for COVID-19, comorbidities, acute respiratory symptoms, age, sex and BMI category | 27 | -14 | 53 |
| Gallant, 2022 (BNT162b2 or mRNA-1273, ≥2 doses) | Mixed (7.4%) | Delta | NA | NA | Adjusted for age, sex, number of symptoms during acute COVID-19, and number of comorbidities. | -3 | -18 | 10 |
| Cortellini, 2023 (BNT162b2, mRNA-1273, ChAdOx1-S or Ad26.COV2.S, 3 doses) | Mixed (40.9%) | Alpha, Delta, Omicron | NA | NA | Inverse probability of treatment weighting applied | 53 | -18 | 82 |
| Ranucci, 2023 (BNT162b2, mRNA-1273, ChAdOx1-S or Ad26.COV2.S or NR, 2 doses) | 100% hospitalized | Wild-type, Alpha, Delta, Omicron | Outcome | Persistent major physical symptoms | Adjusted for age, severity of disease | 69 | 40 | 84 |
| Ranucci, 2023 (BNT162b2, mRNA-1273, ChAdOx1-S or Ad26.COV2.S or NR, 2 doses) | 100% hospitalized | Wild-type, Alpha, Delta, Omicron | Outcome | Persistent major neuropsychological symptoms | Adjusted for previous neurological symptoms | 80 | 57 | 90 |
| Parodi, 2023 (mRNA or inactivated viral, 2 doses) | 100% hospitalized | Gamma, Delta, Omicron | NA | NA | Adjusted for age, sex, hypertension and smoking | 48 | 31 | 60 |
| Al-Aly, 2022 (BNT162b2, mRNA-1273 or Ad26.COV2.S, Primary series) | 100% hospitalized | Wild-type, Alpha, Delta | Setting | Hospitalized | Adjusted for age, race, area deprivation index, outpatient visits, inpatient visits, and unique medication prescriptions in previous year, routine blood chemistry panels, BMI, history of influenza vaccination, smoking status, comorbidities, systolic blood pressure, diastolic blood pressure, estimated glomerular filtration rate, and hospital system average hospital bed occupancy, total hospital bed capacity, positivity rate of tests, number of tests, and HHS region SARS-CoV-2 variant proportion in the week prior to enrolment, geographic region of VISN, and calendar week of enrolment | 12 | 4 | 19 |
| Al-Aly, 2022 (BNT162b2, mRNA-1273 or Ad26.COV2.S, Primary series) | 100% hospitalized | Wild-type, Alpha, Delta | Setting | ICU | Adjusted for age, race, area deprivation index, outpatient visits, inpatient visits, and unique medication prescriptions in previous year, routine blood chemistry panels, BMI, history of influenza vaccination, smoking status, comorbidities, systolic blood pressure, diastolic blood pressure, estimated glomerular filtration rate, and hospital system average hospital bed occupancy, total hospital bed capacity, positivity rate of tests, number of tests, and HHS region SARS-CoV-2 variant proportion in the week prior to enrolment, geographic region of VISN, and calendar week of enrolment | 22 | 10 | 33 |

BMI: Body mass index; CDC: Centers for Disease Control and Prevention; CI: Confidence interval; CPRD: Clinical Practice Research Datalink; GP: General Practitioner; HHS: Health and Human Services; HR: Hazard ratio; ICD-10: International Classification of Diseases- 10; ICU: Intensive care units; IPTW: Inverse probability of treatment weighting; mRNA: Messenger ribonucleic acid; NA: Not applicable; NR: Not reported; OR: Odds ratio; PASC: Post-acute sequelae of COVID-19; PCR: Polymerase Chain Reaction; VA: Veterans Affairs; VE: Vaccine effectiveness; VISN: Veterans Integrated Services Network; VISN: Veterans Integrated Services Network; WHO: World Health Organization.

**eTable 20. Summary of pre-infection aVE against ≥1 PCC (vs. unvaccinated) by proportion of sample hospitalization during acute infection**

| **Proportion hospitalized during acute infection** | **Number of studies** | **Number of estimates** | **Number of statistically significant estimates** | **Proportion of estimates that are statistically significant (%)** | **Minimum statistically significant VE** | **Maximum statistically significant VE** |
| --- | --- | --- | --- | --- | --- | --- |
| 100% hospitalized | 2 | 5 | 5 | 100 | 12 | 80 |
| Mix of those hospitalized and not hospitalized | 13 | 24 | 17 | 71 | 15 | 90 |
| No data on proportion of sample hospitalized | 10 | 54 | 45 | 83 | 10 | 95 |
| 100% non-hospitalized | 3 | 6 | 3 | 50 | 7 | 84 |

VE: vaccine effectiveness.

**eTable 21. Summary of publications where adjusted VE (vs. unvaccinated) against ≥1 PCC could be computed when timing between vaccination and infection is reported, sorted by timing of vaccine relative to infection (N=8 studies with 24 VE estimates)**

| **Author, year (Vaccine, dose)** | **Vaccination timepoint** | **Timing since vaccination** | **Variant** | **Subgroup** | **Subgroup detail** | **Adjustments** | **VE** | **Lower 95% CI** | **Upper 95% CI** |
| --- | --- | --- | --- | --- | --- | --- | --- | --- | --- |
| Ertesvag, 2023 (BNT162b2 or mRNA-1273, ≥1 dose) | Before infection | Mean: 1.5 months to infection | Delta | Timing | 3-months post delta infection | Adjusted by female sex, age, reinfection, symptoms at baseline | -123 | -904 | 45 |
| Herman, 2022 (Inactivated viral vaccine, 2 doses) | Before infection | Fully vaccinated, mean: 85.9 days | Delta, Omicron | NA | NA | Adjusted by shortness of breath | 26 | -8 | 49 |
| Antonelli, 2022a (BNT162b2, mRNA-1273, ChAdOx1-S or NR, 1 dose) | Before infection | Mean (SD): 73 days (44) after first vaccination | Alpha, Delta | NA | NA | Adjusted for age, BMI, sex | -3 | -24 | 15 |
| Antonelli, 2022a (BNT162b2, mRNA-1273, ChAdOx1-S or NR, 2 doses) | Before infection | Mean (SD): 51 (30) days after second vaccination | Alpha, Delta | NA | NA | Adjusted for age, BMI, sex | 49 | 18 | 68 |
| Mohr, 2023 (BNT162b2 or mRNA-1273, 2 doses) | Before infection | >16 weeks since vaccination | Wild-type, Alpha, Delta | Timing since vaccination | >16 weeks | Adjusted for age, race and ethnicity, comorbidities, calendar month of diagnosis, and weeks since symptoms started | 28 | 12 | 41 |
| Mohr, 2023 (BNT162b2 or mRNA-1273, 2 doses) | Before infection | ≤16 weeks since vaccination | Wild-type, Alpha, Delta | Timing since vaccination | ≤16 weeks | Adjusted for age, race and ethnicity, comorbidities, calendar month of diagnosis, and weeks since symptoms started | 33 | 13 | 49 |
| Mohr, 2023 (BNT162b2 or mRNA-1273, 2 doses) | Before infection | 6 weeks since vaccination | Wild-type, Alpha, Delta | Timing since vaccination | 6 weeks | Adjusted for age, race and ethnicity, comorbidities, calendar month of diagnosis, and weeks since symptoms started | 30 | 16 | 42 |
| Di Fusco, 2023 (BNT162b2, 2 doses) | Before infection | Mean (SD): 6.9 (3.0) months since 2nd dose | Omicron | Number of symptoms | ≥3 long COVID symptoms | Adjusted for timepoint, vaccination status and interaction of time by vaccination status, participant pre-COVID-19 symptom onset score, sociodemographic characteristics (age, sex, regions, social vulnerability, race/ethnicity, high risk occupations), previously tested positive for COVID-19, severity of acute illness and immunocompromised status | 40 | -34 | 73 |
| Di Fusco, 2023 (BNT162b2, 2 doses) | Before infection | Mean (SD): 6.9 (3.0) months since 2nd dose | Omicron | Number of symptoms | ≥2 long COVID symptoms | Adjusted for timepoint, vaccination status and interaction of time by vaccination status, participant pre-COVID-19 symptom onset score, sociodemographic characteristics (age, sex, regions, social vulnerability, race/ethnicity, high risk occupations), previously tested positive for COVID-19, severity of acute illness and immunocompromised status | 36 | -39 | 70 |
| Di Fusco, 2023 (BNT162b2, 3 doses) | Before infection | Mean (SD) time from booster dose to infection: 2.3 (1.9) months | Omicron | Number of symptoms | ≥3 long COVID symptoms | Adjusted for timepoint, vaccination status and interaction of time by vaccination status, participant pre-COVID-19 symptom onset score, sociodemographic characteristics (age, sex, regions, social vulnerability, race/ethnicity, high risk occupations), previously tested positive for COVID-19, severity of acute illness and immunocompromised status | 64 | 13 | 85 |
| Di Fusco, 2023 (BNT162b2, 3 doses) | Before infection | Mean (SD) time from booster dose to infection: 2.3 (1.9) months | Omicron | Number of symptoms | ≥2 long COVID symptoms | Adjusted for timepoint, vaccination status and interaction of time by vaccination status, participant pre-COVID-19 symptom onset score, sociodemographic characteristics (age, sex, regions, social vulnerability, race/ethnicity, high risk occupations), previously tested positive for COVID-19, severity of acute illness and immunocompromised status | 70 | 30 | 87 |
| Antonelli, 2022b (Vaccine NR, ≥1 dose) | Before infection | Vaccinated >6 months prior to infection | Delta, Omicron | Timing since vaccination | >6 months post-vaccination | Adjusted by age, sex, BMI, index of multiple deprivation, presence of comorbidities and vaccination status | 74 | 68 | 80 |
| Antonelli, 2022b (Vaccine NR, ≥1 dose) | Before infection | Vaccinated 3-6 months prior to infection | Delta, Omicron | Timing since vaccination | 3-6 months post-vaccination | Adjusted by age, sex, BMI, index of multiple deprivation, presence of comorbidities and vaccination status | 76 | 68 | 81 |
| Antonelli, 2022b (Vaccine NR, ≥1 dose) | Before infection | Vaccinated <3 months prior to infection | Delta, Omicron | Timing since vaccination | <3 months post-vaccination | Adjusted by age, sex, BMI, index of multiple deprivation, presence of comorbidities and vaccination status | 50 | 41 | 57 |
| Antonelli, 2022b (Vaccine NR, ≥1 dose) | Before infection | Vaccinated >6 months prior to infection | Delta, Omicron | Timing since vaccination; age | >6 months post-vaccination; 18–59 years | Adjusted by age, sex, BMI, index of multiple deprivation, presence of comorbidities and vaccination status | 77 | 68 | 84 |
| Antonelli, 2022b (Vaccine NR, ≥1 dose) | Before infection | Vaccinated 3-6 months prior to infection | Delta, Omicron | Timing since vaccination; age | 3-6 months post-vaccination; 18–59 years | Adjusted by age, sex, BMI, index of multiple deprivation, presence of comorbidities and vaccination status | 67 | 56 | 76 |
| Antonelli, 2022b (Vaccine NR, ≥1 dose) | Before infection | Vaccinated <3 months prior to infection | Delta, Omicron | Timing since vaccination; age | <3 months post-vaccination; 18–59 years | Adjusted by age, sex, BMI, index of multiple deprivation, presence of comorbidities and vaccination status | 59 | 47 | 68 |
| Antonelli, 2022b (Vaccine NR, ≥1 dose) | Before infection | Vaccinated >6 months prior to infection | Delta, Omicron | Timing since vaccination; age | >6 months post-vaccination; ≥60 years | Adjusted by age, sex, BMI, index of multiple deprivation, presence of comorbidities and vaccination status | 72 | 61 | 80 |
| Antonelli, 2022b (Vaccine NR, ≥1 dose) | Before infection | Vaccinated 3-6 months prior to infection | Delta, Omicron | Timing since vaccination; age | 3-6 months post-vaccination; ≥60 years | Adjusted by age, sex, BMI, index of multiple deprivation, presence of comorbidities and vaccination status | 54 | -3 | 80 |
| Antonelli, 2022b (Vaccine NR, ≥1 dose) | Before infection | Vaccinated <3 months prior to infection | Delta, Omicron | Timing since vaccination; age | <3 months post-vaccination; ≥60 years | Adjusted by age, sex, BMI, index of multiple deprivation, presence of comorbidities and vaccination status | 46 | 32 | 57 |
| Peghin, 2022 (BNT162b2, mRNA-1273, ChAdOx1-S or Ad26.COV.S, ≥1 dose) | After infection | Patients received the first and second vaccine doses at a mean of 12.4 months (SD: 1.9 months) and 13.5 months (SD: 2.3 months), respectively, after onset of acute COVID-19. | Wild-type, Alpha | Sorted by | Acute COVID-19 severity | Adjusted for gender, age, acute COVID-19 severity and IgG antibodies | 12 | -63 | 53 |
| Peghin, 2022 (BNT162b2, mRNA-1273, ChAdOx1-S or Ad26.COV.S, ≥1 dose) | After infection | Patients received the first and second vaccine doses at a mean of 12.4 months (SD: 1.9 months) and 13.5 months (SD: 2.3 months), respectively, after onset of acute COVID-19. | Wild-type, Alpha | Sorted by | Acute COVID-19 management setting | Adjusted for gender, age, presence of symptoms at acute COVID-19 onset, acute COVID-19 management and IgG antibodies | 10 | -67 | 52 |
| Nehme, 2022 (BNT162b2 or mRNA-1273, 1 dose) | After infection | Mean (SD): 40.3 (29.2) days | Delta | NA | NA | Adjusted for time for infection, age, sex, smoking, and the following comorbidities present prior to the infection: overweight or obese, hypertension, respiratory disease, cardiovascular disease, diabetes, migraine, tension headache, sleeping disorder, anxiety, depression, hypothyroidism, anemia, and chronic fatigue. | 18 | -8 | 39 |
| Nehme, 2022 (BNT162b2 or mRNA-1273, 2 doses) | After infection | Mean (SD): 40.3 (29.2) days | Delta | NA | NA | Adjusted for severity of COVID-19, Chronic medication use, presence of chronic conditions, self-perceived health status, monthly household income, smoking status, living status, educational level, current occupation, marital status, gender and age | 40 | 17 | 57 |

BMI: Body mass index; CI: Confidence interval; mRNA: Messenger ribonucleic acid; NA: Not applicable; NR: Not reported; SD: Standard deviation; VE: Vaccine effectiveness

**eFigure 5.** **Adjusted VE (vs. unvaccinated) against ≥1 PCC for studies that included versus excluded repeat SARS-CoV-2 infections in the analysis, sorted by timing of vaccine relative to infection or PCC and inclusion vs. exclusion of repeat infections**

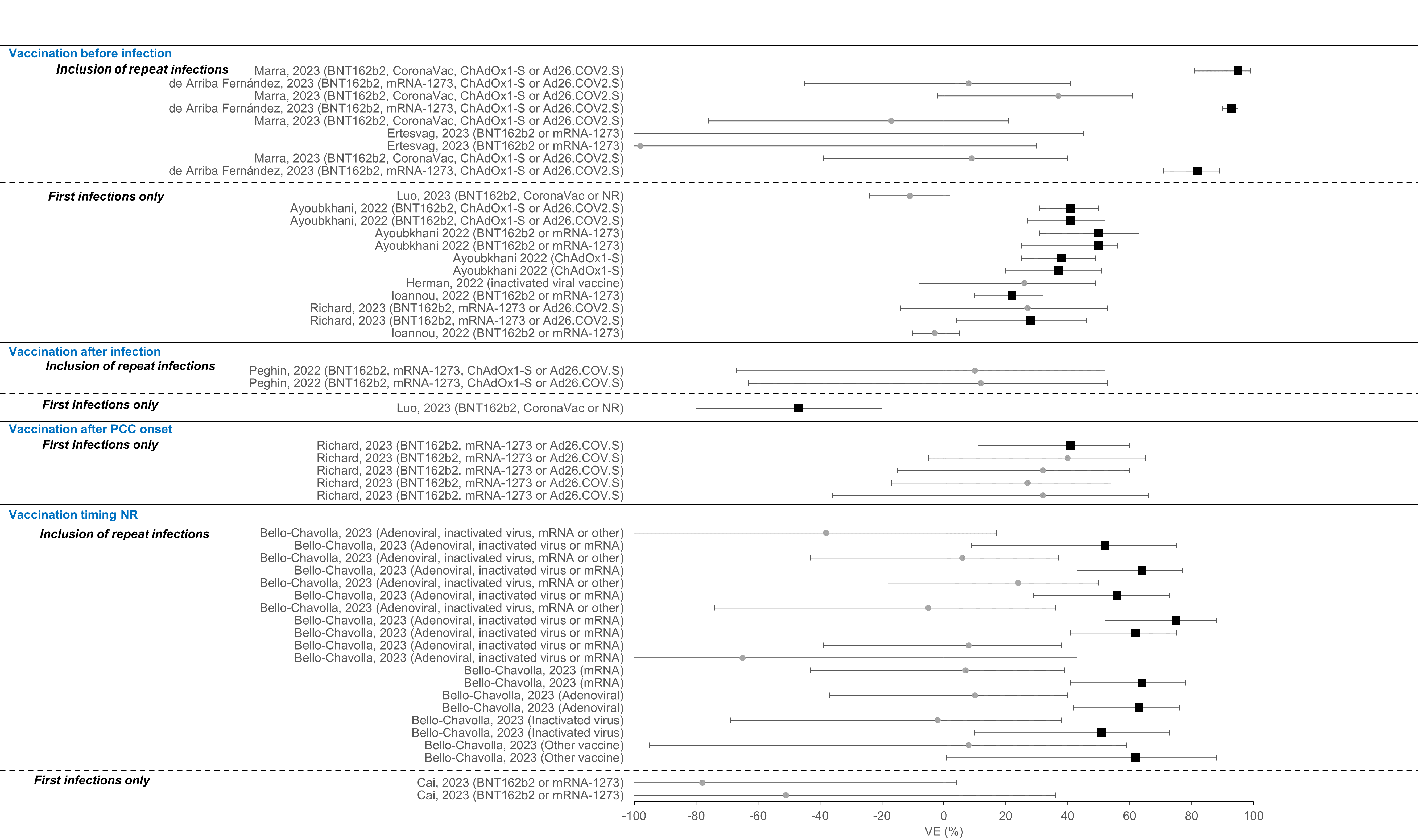

Of 50 VE estimates, 22 (44.0%) reported statistically significant (based on VE 95% CI) protective effects against ≥1 PCC.

CI: Confidence interval: NR: not reported; PCC: post‑COVID‑19 conditions; VE: vaccine effectiveness.

References: Ayoubkhani 2022b^12^; Bello-Chavolla, 2023^16^; Cai, 2023^21^; de Arriba Fernández, 2023^27^; Ertesvag, 2023^36^; Herman, 2022^43^; Ioannou, 2022^45^; Luo, 2023^56^; Marra, 2023^57^; Peghin, 2022^70^; Richard, 2023^78^

**eTable 22. Summary of publications where adjusted VE (vs. unvaccinated) against ≥1 PCC could be computed for studies reporting whether repeat SARS-CoV-2 infections were included or excluded, sorted by timing of vaccination relative to infection or PCC onset and inclusion vs. exclusion of repeat infections (N=11 studies with 50 VE estimates)**

| **Author, year (Vaccine, dose)** | **Vaccination timepoint** | **Re-infection details** | **Re-infection group** | **Variant** | **Subgroup** | **Subgroup details** | **Adjustments** | **VE** | **Lower 95% CI** | **Upper 95% CI** |
| --- | --- | --- | --- | --- | --- | --- | --- | --- | --- | --- |
| Marra, 2023 (BNT162b2, CoronaVac, ChAdOx1-S or Ad26.COV2.S, 4 doses) | Before infection | HCP with at least one SAR-CoV-2 infection during study period | Allows inclusion of repeat infection | Wild-type, Gamma, Delta, Omicron | NA | NA | Adjusted for sex, age, BMI, physical activity level, comorbidities, occupation (no direct patient contact or direct patient facing), number of COVID-19 vaccine doses received prior to infection, homologous or heterologous COVID-19 vaccine, the number of SARS-CoV-2 infections, and SARS-CoV-2 variant | 95 | 81 | 99 |
| de Arriba Fernández, 2023 (BNT162b2, mRNA-1273, ChAdOx1-S or Ad26.COV2.S, booster) | Before infection | Reinfection was suspected in patients with symptoms compatible with COVID-19, who presented SARS-CoV-2 infection, confirmed through AIDT, more than 90 days before | Allows inclusion of repeat infection | Delta, Omicron | NA | NA | Adjusted for age, sex, comorbidity history, body mass index, date of first, second and booster doses of a COVID-19 vaccine, type of COVID-19 vaccine, death, immunosuppressive treatment, diagnosis of persistent COVID-19, and diagnostic test results | 8 | -45 | 41 |
| Marra, 2023 (BNT162b2, CoronaVac, ChAdOx1-S or Ad26.COV2.S, 3 doses) | Before infection | HCP with at least one SAR-CoV-2 infection during study period | Allows inclusion of repeat infection | Wild-type, Gamma, Delta, Omicron | NA | NA | Adjusted for sex, age, BMI, physical activity level, comorbidities, occupation (no direct patient contact or direct patient facing), number of COVID-19 vaccine doses received prior to infection, homologous or heterologous COVID-19 vaccine, the number of SARS-CoV-2 infections, and SARS-CoV-2 variant | 37 | -2 | 61 |
| de Arriba Fernández, 2023 (BNT162b2, mRNA-1273, ChAdOx1-S or Ad26.COV2.S, primary series) | Before infection | Reinfection was suspected in patients with symptoms compatible with COVID-19, who presented SARS-CoV-2 infection, confirmed through AIDT, more than 90 days before | Allows inclusion of repeat infection | Delta, Omicron | NA | NA | Adjusted for age, sex, comorbidity history, body mass index, date of first, second and booster doses of a COVID-19 vaccine, type of COVID-19 vaccine, death, immunosuppressive treatment, diagnosis of persistent COVID-19, and diagnostic test results | 93 | 90 | 95 |
| Marra, 2023 (BNT162b2, CoronaVac, ChAdOx1-S or Ad26.COV2.S, 2 doses) | Before infection | HCP with at least one SAR-CoV-2 infection during study period | Allows inclusion of repeat infection | Wild-type, Gamma, Delta, Omicron | NA | NA | Adjusted for sex, age, BMI, physical activity level, comorbidities, occupation (no direct patient contact or direct patient facing), number of COVID-19 vaccine doses received prior to infection, homologous or heterologous COVID-19 vaccine, the number of SARS-CoV-2 infections, and SARS-CoV-2 variant | -17 | -76 | 21 |
| Ertesvag, 2023 (BNT162b2 or mRNA-1273, ≥1 dose) | Before infection | SARS-CoV-2 reinfections, confirmed by RT-PCR or antigen tests, identified n=2 delta reinfections in Oct-Nov 2021 and n=111 (N=67 >=16 year olds) omicron reinfections from 30th Dec 2021 to April 2022. | Allows inclusion of repeat infection | Delta | Timepoint and population | 3-months post-Delta infection | Adjusted for female sex, age, reinfection, and any symptoms at baseline | -123 | -904 | 45 |
| Ertesvag, 2023 (BNT162b2 or mRNA-1273, ≥1 dose) | Before infection | SARS-CoV-2 reinfections, confirmed by RT-PCR or antigen tests, identified n=2 delta reinfections in Oct-Nov 2021 and n=111 (N=67 >=16 year olds) omicron reinfections from 30th Dec 2021 to April 2022. | Allows inclusion of repeat infection | Delta | Timepoint and population | 8-months post-Delta infection [≥16 years]) | Adjusted for female sex, age, reinfection, and any symptoms at baseline | -98 | -457 | 30 |
| Marra, 2023 (BNT162b2, CoronaVac, ChAdOx1-S or Ad26.COV2.S, 1 dose) | Before infection | HCP with at least one SAR-CoV-2 infection during study period | Allows inclusion of repeat infection | Wild-type, Gamma, Delta, Omicron | NA | NA | Adjusted for sex, age, BMI, physical activity level, comorbidities, occupation (no direct patient contact or direct patient facing), number of COVID-19 vaccine doses received prior to infection, homologous or heterologous COVID-19 vaccine, the number of SARS-CoV-2 infections, and SARS-CoV-2 variant | 9 | -39 | 40 |
| de Arriba Fernández, 2023 (BNT162b2, mRNA-1273, ChAdOx1-S or Ad26.COV2.S, 1 dose) | Before infection | Reinfection was suspected in patients with symptoms compatible with COVID-19, who presented SARS-CoV-2 infection, confirmed through AIDT, more than 90 days before | Allows inclusion of repeat infection | Delta, Omicron | NA | NA | Adjusted for age, sex, comorbidity history, body mass index, date of first, second and booster doses of a COVID-19 vaccine, type of COVID-19 vaccine, death, immunosuppressive treatment, diagnosis of persistent COVID-19, and diagnostic test results | 82 | 71 | 89 |
| Luo, 2023 (BNT162b2, CoronaVac or NR, ≥3 doses) | Before infection | Excluded individuals with COVID symptoms attributable to other diseases (e.g. common cold or COVID-19 reinfection) | First infections only | Omicron | NA | NA | Adjusted for age, gender, BMI, infection days, vaccination status, vaccination after infection, comorbidities, infection days, treatment for the acute stage of illness, and patient's satisfaction of treatment | -11 | -24 | 2 |
| Ayoubkhani, 2022 (BNT162b2, ChAdOx1-S or Ad26.COV2.S, 2 doses) | Before infection | Activity limiting long COVID ≥12 weeks after first infection | First infections only | Delta | Vaccine and long COVID severity | Adenoviral and mRNA; Long COVID of any severity | Adjusted for age, sex, white or non-white ethnicity, country/region of residence, area deprivation quintile group, and self-reported, pre-existing health/disability status and time from infection to follow-up for Long Covid | 41 | 31 | 50 |
| Ayoubkhani, 2022 (BNT162b2, ChAdOx1-S or Ad26.COV2.S, 2 doses) | Before infection | Long COVID of any severity ≥12 weeks after first infection | First infections only | Delta | Vaccine and long COVID severity | Adenoviral and mRNA; activity-limiting Long COVID | Adjusted for age, sex, white or non-white ethnicity, country/region of residence, area deprivation quintile group, and self-reported, pre-existing health/disability status and time from infection to follow-up for Long Covid | 41 | 27 | 52 |
| Ayoubkhani 2022 (BNT162b2 or mRNA-1273, 2 doses) | Before infection | Activity limiting long COVID ≥12 weeks after first infection | First infections only | Delta | Vaccine and long COVID severity | mRNA vaccine platform; Long COVID of any severity | Adjusted for age, sex, white or non-white ethnicity, country/region of residence, area deprivation quintile group, and self-reported, pre-existing health/disability status and time from infection to follow-up for Long Covid | 50 | 31 | 63 |
| Ayoubkhani 2022 (BNT162b2 or mRNA-1273, 2 doses) | Before infection | Long COVID of any severity ≥12 weeks after first infection | First infections only | Delta | Vaccine and long COVID severity | mRNA vaccine platform; activity-limiting Long COVID | Adjusted for age, sex, white or non-white ethnicity, country/region of residence, area deprivation quintile group, and self-reported, pre-existing health/disability status and time from infection to follow-up for Long Covid | 50 | 25 | 56 |
| Ayoubkhani 2022 (ChAdOx1-S, 2 doses) COVID) | Before infection | Activity limiting long COVID ≥12 weeks after first infection | First infections only | Delta | Vaccine and long COVID severity | AstraZeneca; Long COVID of any severity | Adjusted for age, sex, white or non-white ethnicity, country/region of residence, area deprivation quintile group, and self-reported, pre-existing health/disability status and time from infection to follow-up for Long Covid | 38 | 25 | 49 |
| Ayoubkhani 2022 (ChAdOx1-S, 2 doses) COVID) limiting long COVID) | Before infection | Long COVID of any severity ≥12 weeks after first infection | First infections only | Delta | Vaccine and long COVID severity | AstraZeneca; activity limiting Long COVID | Adjusted for age, sex, white or non-white ethnicity, country/region of residence, area deprivation quintile group, and self-reported, pre-existing health/disability status and time from infection to follow-up for Long Covid | 37 | 20 | 51 |
| Herman, 2022 (inactivated viral vaccine, 2 doses) | Before infection | Those with suggested reinfection were excluded. | First infections only | Delta, Omicron | NA | NA | Adjusted by shortness of breath | 26 | -8 | 49 |
| Ioannou, 2022 (BNT162b2 or mRNA-1273, 2 doses) | Before infection | Participants included with no evidence of reinfection | First infections only | Wild-type, Alpha | NA | NA | Adjusted for age, sex, race, ethnicity, urban vs rural residence, Charlson Comorbidity Index score, VA Integrated Service Network, time period of infection/pandemic wave, number of primary care, mental health and specialty care encounters in the 2 years before infection | 22 | 10 | 32 |
| Richard, 2023 (BNT162b2, mRNA-1273 or Ad26.COV2.S, Primary series) | Before infection | Participants with suspected reinfection were excluded | First infections only | Wild-type, Alpha, Delta | Timepoint | ≥90 days | Adjusted for Delta variant period, maximum symptom severity during acute illness, days hospitalized for COVID-19, comorbidities, acute respiratory symptoms, age, sex and BMI category | 27 | -14 | 53 |
| Richard, 2023 (BNT162b2, mRNA-1273 or Ad26.COV2.S, Primary series) | Before infection | Participants with suspected reinfection were excluded | First infections only | Wild-type, Alpha, Delta | Timepoint | ≥28 days | Adjusted for Delta variant period, maximum symptom severity during acute illness, days hospitalized for COVID-19, comorbidities, acute respiratory symptoms, age, sex and BMI category | 28 | 4 | 46 |
| Ioannou, 2022 (BNT162b2 or mRNA-1273, 1 dose) | Before infection | Participants included with no evidence of reinfection | First infections only | Wild-type, Alpha | NA | NA | Adjusted for age, sex, race, ethnicity, urban vs rural residence, Charlson Comorbidity Index score, VA Integrated Service Network, time period of infection/pandemic wave, number of primary care, mental health and specialty care encounters in the 2 years before infection | -3 | -10 | 5 |
| Peghin, 2022 (BNT162b2, mRNA-1273, ChAdOx1-S or Ad26.COV.S, ≥1 dose) | After infection | Clinical re-infection defined as positive SARS-CoV-2 PCR or antigenic test more than 3 months after the onset of the primary infection with or without recurrence of symptoms compatible with COVID-19 (N=NR) | Allows inclusion of repeat infection | Wild-type, Alpha | Sorted by | Acute COVID-19 management setting) | Adjusted for gender, age, presence of symptoms at acute COVID-19 onset, acute COVID-19 management and IgG antibodies | 10 | -67 | 52 |
| Peghin, 2022 (BNT162b2, mRNA-1273, ChAdOx1-S or Ad26.COV.S, ≥1 dose) | After infection | Clinical re-infection defined as positive SARS-CoV-2 PCR or antigenic test more than 3 months after the onset of the primary infection with or without recurrence of symptoms compatible with COVID-19 (N=NR) | Allows inclusion of repeat infection | Wild-type, Alpha | Sorted by | Acute COVID-19 severity | Adjusted for gender, age, acute COVID-19 severity and IgG antibodies | 12 | -63 | 53 |
| Luo, 2023 (BNT162b2, CoronaVac or NR, dose NR) | After infection | Excluded individuals with COVID symptoms attributable to other diseases (e.g. common cold or COVID-19 reinfection) | First infections only | Omicron | NA | NA | Adjusted for age, gender, BMI, infection days, vaccination status, vaccination after infection, comorbidities, infection days, treatment for the acute stage of illness, and patient's satisfaction of treatment | -47 | -80 | -20 |
| Richard, 2023 (BNT162b2, mRNA-1273 or Ad26.COV.S, primary series) | After PCC onset | Participants with suspected reinfection were excluded | First infections only | Wild-type, Alpha, Delta | Follow-up | 12 months | Adjusted for Delta variant period, maximum symptom severity during acute illness, days hospitalized for COVID-19, comorbidities, acute respiratory symptoms, age, sex and BMI category | 41 | 11 | 60 |
| Richard, 2023 (BNT162b2, mRNA-1273 or Ad26.COV.S, primary series) | After PCC onset | Participants with suspected reinfection were excluded | First infections only | Wild-type, Alpha, Delta | Follow-up | 9 months | Adjusted for Delta variant period, maximum symptom severity during acute illness, days hospitalized for COVID-19, comorbidities, acute respiratory symptoms, age, sex and BMI category | 40 | -5 | 65 |
| Richard, 2023 (BNT162b2, mRNA-1273 or Ad26.COV.S, primary series) | After PCC onset | Participants with suspected reinfection were excluded | First infections only | Wild-type, Alpha, Delta | Follow-up | 6 months | Adjusted for Delta variant period, maximum symptom severity during acute illness, days hospitalized for COVID-19, comorbidities, acute respiratory symptoms, age, sex and BMI category | 32 | -15 | 60 |
| Richard, 2023 (BNT162b2, mRNA-1273 or Ad26.COV.S, primary series) | After PCC onset | Participants with suspected reinfection were excluded | First infections only | Wild-type, Alpha, Delta | Follow-up | 3 months | Adjusted for Delta variant period, maximum symptom severity during acute illness, days hospitalized for COVID-19, comorbidities, acute respiratory symptoms, age, sex and BMI category | 27 | -17 | 54 |
| Richard, 2023 (BNT162b2, mRNA-1273 or Ad26.COV.S, primary series) | After PCC onset | Participants with suspected reinfection were excluded | First infections only | Wild-type, Alpha, Delta | Follow-up | 1 month | Adjusted for Delta variant period, maximum symptom severity during acute illness, days hospitalized for COVID-19, comorbidities, acute respiratory symptoms, age, sex and BMI category | 32 | -36 | 66 |
| Bello-Chavolla, 2023 (Adenoviral, inactivated virus, mRNA or other, 4 doses) | NR | Multiple infections permitted (328 (82%) had SARS-CoV-2 infection once, 72 (18%) more than once) | Allows inclusion of repeat infection | Delta, Omicron | Long COVID definition | WHO definition | Adjusted by survey weights | -38 | -132 | 17 |
| Bello-Chavolla, 2023 (Adenoviral, inactivated virus, mRNA or other, 4 doses) | NR | Multiple infections permitted (328 (82%) had SARS-CoV-2 infection once, 72 (18%) more than once) | Allows inclusion of repeat infection | Delta, Omicron | Long COVID definition | PASC score ≥12 | Adjusted by survey weights | 52 | 9 | 75 |
| Bello-Chavolla, 2023 (Adenoviral, inactivated virus, mRNA or other, 3 doses | NR | Multiple infections permitted (328 (82%) had SARS-CoV-2 infection once, 72 (18%) more than once) | Allows inclusion of repeat infection | Delta, Omicron | Long COVID definition | WHO definition | Adjusted by survey weights | 6 | -43 | 37 |
| Bello-Chavolla, 2023 (Adenoviral, inactivated virus or mRNA, 3 doses) | NR | Multiple infections permitted (328 (82%) had SARS-CoV-2 infection once, 72 (18%) more than once) | Allows inclusion of repeat infection | Delta, Omicron | Long COVID definition | PASC score ≥12 | Adjusted by survey weights | 64 | 43 | 77 |
| Bello-Chavolla, 2023 (Adenoviral, inactivated virus, mRNA or other, 2 doses) | NR | Multiple infections permitted (328 (82%) had SARS-CoV-2 infection once, 72 (18%) more than once) | Allows inclusion of repeat infection | Delta, Omicron | Long COVID definition | WHO definition | Adjusted by survey weights | 24 | -18 | 50 |
| Bello-Chavolla, 2023 (Adenoviral, inactivated virus or mRNA, 2 doses) | NR | Multiple infections permitted (328 (82%) had SARS-CoV-2 infection once, 72 (18%) more than once) | Allows inclusion of repeat infection | Delta, Omicron | Long COVID definition | PASC score ≥12 | Adjusted by survey weights | 56 | 29 | 73 |
| Bello-Chavolla, 2023 (Adenoviral, inactivated virus, mRNA or other, 1 dose) | NR | Multiple infections permitted (328 (82%) had SARS-CoV-2 infection once, 72 (18%) more than once) | Allows inclusion of repeat infection | Delta, Omicron | Long COVID definition | WHO definition | Adjusted by survey weights | -5 | -74 | 36 |
| Bello-Chavolla, 2023 (Adenoviral, inactivated virus or mRNA, 1 dose) | NR | Multiple infections permitted (328 (82%) had SARS-CoV-2 infection once, 72 (18%) more than once) | Allows inclusion of repeat infection | Delta, Omicron | Long COVID definition | PASC score ≥12 | Adjusted by survey weights | 75 | 52 | 88 |
| Bello-Chavolla, 2023 (Adenoviral, inactivated virus or mRNA, ≥1 dose) | NR | Multiple infections permitted (328 (82%) had SARS-CoV-2 infection once, 72 (18%) more than once) | Allows inclusion of repeat infection | Delta, Omicron | Long COVID definition | Presence of any persistent COVID-19 symptom | Adjusted by survey weights | 62 | 41 | 75 |
| Bello-Chavolla, 2023 (Adenoviral, inactivated virus or mRNA, ≥1 dose) | NR | Multiple infections permitted (328 (82%) had SARS-CoV-2 infection once, 72 (18%) more than once) | Allows inclusion of repeat infection | Delta, Omicron | Long COVID definition | PASC defined by PASC score ≥12 points | Adjusted by survey weights | 8 | -39 | 38 |
| Bello-Chavolla, 2023 (Adenoviral, inactivated virus or mRNA, ≥1 dose) | NR | Multiple infections permitted (328 (82%) had SARS-CoV-2 infection once, 72 (18%) more than once) | Allows inclusion of repeat infection | Delta, Omicron | Long COVID definition | Incapacitating persistent post COVID-19 sequelae disaggregated by PASC scores using a 12-point threshold | Adjusted by survey weights | -65 | -503 | 43 |
| Bello-Chavolla, 2023 (mRNA, ≥1 dose) | NR | Multiple infections permitted (328 (82%) had SARS-CoV-2 infection once, 72 (18%) more than once) | Allows inclusion of repeat infection | Delta, Omicron | Vaccine; Long COVID definition | mRNA; WHO definition | Adjusted by survey weights | 7 | -43 | 39 |
| Bello-Chavolla, 2023 (mRNA, ≥1 dose) | NR | Multiple infections permitted (328 (82%) had SARS-CoV-2 infection once, 72 (18%) more than once) | Allows inclusion of repeat infection | Delta, Omicron | Vaccine; Long COVID definition | mRNA; PASC defined by PASC score ≥12 points | Adjusted by survey weights | 64 | 41 | 78 |
| Bello-Chavolla, 2023 (Adenoviral, ≥1 dose) | NR | Multiple infections permitted (328 (82%) had SARS-CoV-2 infection once, 72 (18%) more than once) | Allows inclusion of repeat infection | Delta, Omicron | Vaccine; Long COVID definition | Adenovirus vector; WHO definition | Adjusted by survey weights | 10 | -37 | 40 |
| Bello-Chavolla, 2023 (Adenoviral, ≥1 dose) | NR | Multiple infections permitted (328 (82%) had SARS-CoV-2 infection once, 72 (18%) more than once) | Allows inclusion of repeat infection | Delta, Omicron | Vaccine; Long COVID definition | Adenovirus vector; PASC defined by PASC score ≥12 points | Adjusted by survey weights | 63 | 42 | 76 |
| Bello-Chavolla, 2023 (Inactivated virus, ≥1 dose) | NR | Multiple infections permitted (328 (82%) had SARS-CoV-2 infection once, 72 (18%) more than once) | Allows inclusion of repeat infection | Delta, Omicron | Vaccine; Long COVID definition | Inactivated virus; WHO definition | Adjusted by survey weights | -2 | -69 | 38 |
| Bello-Chavolla, 2023 (Inactivated virus, ≥1 dose) | NR | Multiple infections permitted (328 (82%) had SARS-CoV-2 infection once, 72 (18%) more than once) | Allows inclusion of repeat infection | Delta, Omicron | Vaccine; Long COVID definition | Inactivated virus; PASC defined by PASC score ≥12 points | Adjusted by survey weights | 51 | 10 | 73 |
| Bello-Chavolla, 2023 (Other vaccine, ≥1 dose) | NR | Multiple infections permitted (328 (82%) had SARS-CoV-2 infection once, 72 (18%) more than once) | Allows inclusion of repeat infection | Delta, Omicron | Vaccine; Long COVID definition | Other; WHO definition | Adjusted by survey weights | 8 | -95 | 59 |
| Bello-Chavolla, 2023 (Other vaccine, ≥1 dose) | NR | Multiple infections permitted (328 (82%) had SARS-CoV-2 infection once, 72 (18%) more than once) | Allows inclusion of repeat infection | Delta, Omicron | Vaccine; Long COVID definition | Other; PASC defined by PASC score ≥12 points | Adjusted by survey weights | 62 | 1 | 88 |
| Cai, 2023 (BNT162b2 or mRNA-1273, ≥2 doses) | NR | At 12-month follow-up, reinfected individuals were excluded from follow-up | First infections only | Omicron | NA | NA | Adjusted for age, sex, COVID-19 vaccination status, number of comorbidities and severity during acute infection | -78 | -228 | 4 |
| Cai, 2023(BNT162b2 or mRNA-1273, 1 dose) | NR | Re-infection group | First infections only | Omicron | NA | NA | Adjusted for age, sex, COVID-19 vaccination status, number of comorbidities and severity during acute infection | -51 | -259 | 36 |

AIDT: Active infection diagnostic test; BMI: Body mass index; CDC: Centers for Disease Control and Prevention; CI: Confidence interval; CPRD: Clinical Practice Research Datalink; GP: General Practitioner; HCP: Healthcare professional; mRNA: Messenger ribonucleic acid; NA: Not applicable; NR: Not reported; PASC: Post-acute sequelae of COVID-19; PCC: Post- COVID condition; PCR: Polymerase Chain Reaction; VA: Veterans Affairs; VE: Vaccine effectiveness; WHO: World Health Organization.

References

1.	Abu Hamdh BN, Z. A prospective cohort study assessing the relationship between long-COVID symptom incidence in COVID-19 patients and COVID-19 vaccination. *Scientific reports*. 25 Mar 2023;13(1):4896. doi:<https://dx.doi.org/10.1038/s41598-023-30583-2>

2.	Al-Aly Z, Bowe B, Xie Y. Long COVID after breakthrough SARS-CoV-2 infection. *Nature Medicine*. 2022/07/01 2022;28(7):1461-1467. doi:10.1038/s41591-022-01840-0

3.	Alghamdi HY, Alrashed AM, Jawhari AM, Abdel-Moneim AS. Neuropsychiatric symptoms in post-COVID-19 long haulers. *Acta Neuropsychiatr*. Dec 2022;34(6):318-329. doi:<https://dx.doi.org/10.1017/neu.2022.13>

4.	Alradhi MSHA, Y. M. A. A Comparative Study to the Persistence Post COVID Syndrome and the Infection Period between Vaccinated and Non-vaccinated Patients in Al Basrah Province. *Indian Journal of Forensic Medicine and Toxicology*. October-December 2022;16(4):328-334. doi:<https://dx.doi.org/10.37506/ijfmt.v16i4.18609>

5.	Angarita-Fonseca AT-C, R.Benavides-Cordoba, V.Chero, S.Morales-Satan, M.Hernandez-Lopez, B.Salazar-Perez, R.Larrateguy, S.Sanchez-Ramirez, D. C. Exploring long COVID condition in Latin America: Its impact on patients' activities and associated healthcare use. *Frontiers in Medicine*. 2023;10 (no pagination)1168628. doi:<https://dx.doi.org/10.3389/fmed.2023.1168628>

6.	Antonelli M, Penfold RS, Merino J, et al. Risk factors and disease profile of post-vaccination SARS-CoV-2 infection in UK users of the COVID Symptom Study app: a prospective, community-based, nested, case-control study. *The Lancet Infectious Diseases*. January 2022;22(1):43-55. doi:<https://dx.doi.org/10.1016/S1473-3099%2821%2900460-6>

7.	Antonelli M, Pujol JC, Spector TD, Ourselin S, Steves CJ. Risk of long COVID associated with delta versus omicron variants of SARS-CoV-2. *The Lancet*. 2022;399(10343):2263-2264. doi:10.1016/S0140-6736(22)00941-2

8.	Arjun MCS, A. K.Pal, D.Das, K.Alekhya, G.Venkateshan, M.Mishra, B.Patro, B. K.Mohapatra, P. R.Subba, S. H. Characteristics and predictors of Long COVID among diagnosed cases of COVID-19. *PLoS ONE*. December 2022;17(12 December) (no pagination)e0278825. doi:<https://dx.doi.org/10.1371/journal.pone.0278825>

9.	Arjun MCS, A. K.Roy, P.Ravichandran, M.Mandal, S.Pal, D.Das, K.Gajjala, A.Venkateshan, M.Mishra, B.Patro, B. K.Mohapatra, P. R.Subba, S. H. Long COVID following Omicron wave in Eastern India-A retrospective cohort study. *Journal of Medical Virology*. January 2023;95(1) (no pagination)e28214. doi:<https://dx.doi.org/10.1002/jmv.28214>

10.	Augustin MS, M.Wuestenberg, H.Di Cristanziano, V.Pracht, E.Rauschning, D.Gruell, H.Klein, F.Hallek, M.Schommers, P.Lehmann, C. 15-Month Attributes of Post-Covid Syndrome in Therapeutically Vaccinated Outpatients. Conference Abstract. *Topics in Antiviral Medicine*. April 2023;31(2):285-286.

11.	Ayoubkhani DB, C.Pouwels, K. B.Glickman, M.Nafilyan, V.Zaccardi, F.Khunti, K.Alwan, N. A.Walker, A. S. Trajectory of long covid symptoms after covid-19 vaccination: community based cohort study. *The BMJ*. 2022;(no pagination)e069676. doi:<https://dx.doi.org/10.1136/bmj-2021-069676>

12.	Ayoubkhani D, Bosworth ML, King S, et al. Risk of Long COVID in People Infected With Severe Acute Respiratory Syndrome Coronavirus 2 After 2 Doses of a Coronavirus Disease 2019 Vaccine: Community-Based, Matched Cohort Study. *Open Forum Infectious Diseases*. September 2022;9(9) (no pagination)ofac464. doi:<https://dx.doi.org/10.1093/ofid/ofac464>

13.	Azzolini E, Levi R, Sarti R, et al. Association Between BNT162b2 Vaccination and Long COVID After Infections Not Requiring Hospitalization in Health Care Workers. *Jama*. Aug 16 2022;328(7):676-678. doi:10.1001/jama.2022.11691

14.	Babicki MK, J.Pieniawska-Smiech, K.Kaluzinska-Kolat, Z.Kolat, D.Mastalerz-Migas, A.Jankowski, P.Chudzik, M. Do COVID-19 Vaccinations Affect the Most Common Post-COVID Symptoms? Initial Data from the STOP-COVID Register-12-Month Follow-Up. *Viruses*. June 2023;15(6) (no pagination)1370. doi:<https://dx.doi.org/10.3390/v15061370>

15.	Ballouz TM, D.Kaufmann, M.Amati, R.Frei, A.von Wyl, V.Fehr, J. S.Albanese, E.Puhan, M. A. Post COVID-19 condition after Wildtype, Delta, and Omicron SARS-CoV-2 infection and prior vaccination: Pooled analysis of two population-based cohorts. *PLoS ONE*. February 2023;18(2 February) (no pagination)e0281429. doi:<https://dx.doi.org/10.1371/journal.pone.0281429>

16.	Bello-Chavolla OY, Fermín-Martínez CA, Fernández-Chirino L, et al. Nationally representative prevalence and determinants of post-acute sequelae of SARS-CoV-2 infection (Long COVID) amongst Mexican adults in 2022. *medRxiv*. 2023:2023.07.10.23292475. doi:10.1101/2023.07.10.23292475

17.	Berry DSF, Dalhuisen T, Marchena G, Tiemessen I, Geubbels E, Jaspers L. Association between post-infection COVID-19 vaccination and symptom severity of post COVID-19 condition among patients on Bonaire, Caribbean Netherlands: a retrospective cohort study. *medRxiv*. 2023:2023.06.20.23291649. doi:10.1101/2023.06.20.23291649

18.	Bostanci A, Gazi U, Tosun O, et al. Long-COVID-19 in Asymptomatic, Non-Hospitalized, and Hospitalized Populations: A Cross-Sectional Study. *Journal of Clinical Medicine*. 2023;12(7). doi:10.3390/jcm12072613 <https://mdpi-res.com/d_attachment/jcm/jcm-12-02613/article_deploy/jcm-12-02613-v2.pdf?version=1680248845>

19.	Brannock MD, Chew RF, Preiss AJ, et al. Long COVID Risk and Pre-COVID Vaccination: An EHR-Based Cohort Study from the RECOVER Program. Preprint. *medRxiv*. Oct 07 2022;07:07. doi:<https://dx.doi.org/10.1101/2022.10.06.22280795>

20.	Brunvoll SHN, A. B.Fagerland, M. W.Holland, P.Ellingjord-Dale, M.Dahl, J. A.Soraas, A. Post-acute symptoms 3-15 months after COVID-19 among unvaccinated and vaccinated individuals with a breakthrough infection. *International Journal of Infectious Diseases*. January 2022;126:10-13. doi:<https://dx.doi.org/10.1016/j.ijid.2022.11.009>

21.	Cai J, Lin K, Zhang H, et al. A one-year follow-up study of systematic impact of long COVID symptoms among patients post SARS-CoV-2 omicron variants infection in Shanghai, China. *Emerg Microbes Infect*. Dec 2023;12(2):2220578. doi:10.1080/22221751.2023.2220578

22.	Català M, Mercadé-Besora N, Kolde R, et al. The Effectiveness of COVID-19 Vaccines to Prevent Long COVID Symptoms: Staggered Cohort Analyses of Data from the UK, Spain, and Estonia. *SSRN*. 2023;doi:<https://dx.doi.org/10.2139/ssrn.4474215>

23.	Chelly S, Rouis S, Ezzi O, et al. Symptoms and risk factors for long COVID in Tunisian population. *BMC Health Services Research*. 2023/05/15 2023;23(1):487. doi:10.1186/s12913-023-09463-y

24.	Chilunga FP, Appelman B, van Vugt M, et al. Differences in incidence, nature of symptoms, and duration of long COVID among hospitalised migrant and non-migrant patients in the Netherlands: a retrospective cohort study. *The Lancet Regional Health – Europe*. 2023;29doi:10.1016/j.lanepe.2023.100630

25.	Cortellini A, Tabernero J, Mukherjee U, et al. SARS-CoV-2 omicron (B.1.1.529)-related COVID-19 sequelae in vaccinated and unvaccinated patients with cancer: results from the OnCovid registry. *The Lancet Oncology*. April 2023;24(4):335-346. doi:<https://dx.doi.org/10.1016/S1470-2045%2823%2900056-6>

26.	de Arriba Fernandez AAB, J. L.Espineira Frances, A.Cabeza Mora, A.Gutierrez Perez, A.Diaz Barreiros, M. A.Serra Majem, L. Assessment of SARS-CoV-2 Infection According to Previous Metabolic Status and Its Association with Mortality and Post-Acute COVID-19. *Nutrients*. July 2022;14(14) (no pagination)2925. doi:<https://dx.doi.org/10.3390/nu14142925>

27.	de Arriba Fernandez AB, J. L. A.Frances, A. E.Mora, A. C.Perez, A. G.Barreiros, M. Ad. Evaluation of persistent COVID and SARS-CoV-2 reinfection in a cohort of patients on the island of Gran Canaria, Spain. *Semergen*. 25 Jan 2023;49(5):101939. doi:<https://dx.doi.org/10.1016/j.semerg.2023.101939>

28.	de Bruijn S, Jan van Hoek A, Mutubuki EN, et al. Lower prevalence of Post-Covid-19 Condition following Omicron SARS-CoV-2 infection. *medRxiv*. 2023:2023.04.05.23288157. doi:10.1101/2023.04.05.23288157

29.	Di Fusco M, Sun X, Moran MM, et al. Impact of COVID-19 and effects of booster vaccination with BNT162b2 on six-month long COVID symptoms, quality of life, work productivity and activity impairment during Omicron. *Journal of Patient-Reported Outcomes*. 2023/07/24 2023;7(1):77. doi:10.1186/s41687-023-00616-5

30.	Durstenfeld MS, Peluso MJ, Peyser ND, et al. Factors Associated with Long COVID Symptoms in an Online Cohort Study. *Open Forum Infectious Diseases*. 01 Feb 2023;10(2) (no pagination)ofad047. doi:<https://dx.doi.org/10.1093/ofid/ofad047>

31.	El Otmani HN, S.Berrada, M.Bellakhdar, S.El Moutawakil, B.Abdoh Rafai, M. Prevalence, characteristics and risk factors in a Moroccan cohort of Long-Covid-19. *Neurological Sciences*. September 2022;43(9):5175-5180. doi:<https://dx.doi.org/10.1007/s10072-022-06138-0>

32.	Elias TP, Gebreamlak TW, Gebremeskel TT, et al. Determinants of Post-acute COVID-19 Syndrome among hospitalized severe COVID-19 patients: a 2-year follow up study. *medRxiv*. 2023:2023.06.13.23290674. doi:10.1101/2023.06.13.23290674

33.	Elmazny A, Magdy R, Hussein M, et al. Neuropsychiatric post-acute sequelae of COVID-19: prevalence, severity, and impact of vaccination. *European Archives of Psychiatry and Clinical Neuroscience*. 2023;doi:<https://dx.doi.org/10.1007/s00406-023-01557-2>

34.	Elnadi H, Al-Mustapha AI, Odetokun IA, et al. Prevalence of Post COVID-19 Condition among healthcare workers: self-reported online survey in four African countries. *medRxiv*. 2023:2023.06.22.23291768. doi:10.1101/2023.06.22.23291768

35.	Emecen AN, Keskin S, Turunc O, et al. The presence of symptoms within 6 months after COVID-19: a single-center longitudinal study. *Irish Journal of Medical Science*. April 2023;192(2):741-750. doi:<https://dx.doi.org/10.1007/s11845-022-03072-0>

36.	Ertesvag NU, Iversen A, Blomberg B, et al. Post COVID-19 condition after delta infection and omicron reinfection in children and adolescents. *eBioMedicine*. June 2023;92 (no pagination)104599. doi:<https://dx.doi.org/10.1016/j.ebiom.2023.104599>

37.	Fernandez-de-las-Penas CO-S, R.Fuensalida-Novo, S.Martin-Guerrero, J. D.Pellicer-Valero, O. J.Torres-Macho, J. Differences in Long-COVID Symptoms between Vaccinated and Non-Vaccinated (BNT162b2 Vaccine) Hospitalized COVID-19 Survivors Infected with the Delta Variant. *Vaccines*. September 2022;10(9) (no pagination)1481. doi:<https://dx.doi.org/10.3390/vaccines10091481>

38.	Gaber TAZKA, A.Unsworth, A.Martindale, J. Are mRNA covid 19 vaccines safe in long covid patients? A health care workers perspective. *British Journal of Medical Practitioners*. 2021;14(1) (no pagination)a008.

39.	Gallant MR-P, C.Lemaire-Paquette, S.Piche, A. SARS-CoV-2 infection outcomes associated with the Delta variant: A prospective cohort study. *Jammi*. 2022;8(1):49-56. doi:<https://dx.doi.org/10.3138/jammi-2022-0022>

40.	Hajjaji N, Lepoutre K, Lakhdar S, et al. 16 Months Follow Up of Patients' Behavior and Mild COVID-19 Patterns in a Large Cohort of Cancer Patients During the Pandemic. *Front Oncol*. 2022;12:901426. doi:10.3389/fonc.2022.901426

41.	Hastie CE, Lowe DJ, McAuley A, et al. Outcomes among confirmed cases and a matched comparison group in the Long-COVID in Scotland study. *Nat Commun*. Oct 12 2022;13(1):5663. doi:10.1038/s41467-022-33415-5

42.	Haupert SRS, X.Chen, C.Fritsche, L. G.Mukherjee, B. A Case-Crossover Phenome-wide association study (PheWAS) for understanding Post-COVID-19 diagnosis patterns. *Journal of Biomedical Informatics*. December 2022;136 (no pagination)104237. doi:<https://dx.doi.org/10.1016/j.jbi.2022.104237>

43.	Herman BW, M. C. S.Viwattanakulvanid, P. Vaccination status, favipiravir, and micronutrient supplementation roles in post-COVID symptoms: A longitudinal study. *PLoS ONE*. July 2022;17(7 July) (no pagination)e0271385. doi:<https://dx.doi.org/10.1371/journal.pone.0271385>

44.	Hirahata KN, N.Fujiwara, T. Characteristics of Long COVID: Cases from the First to the Fifth Wave in Greater Tokyo, Japan. *Journal of Clinical Medicine*. November 2022;11(21) (no pagination)6457. doi:<https://dx.doi.org/10.3390/jcm11216457>

45.	Ioannou GNB, A.Fox, A.Shahoumian, T.Hickok, A.O'Hare, A. M.Bohnert, A. S. B.Boyko, E. J.Maciejewski, M. L.Bowling, C. B.Viglianti, E.Iwashyna, T. J.Hynes, D. M. Rates and Factors Associated with Documentation of Diagnostic Codes for Long COVID in the National Veterans Affairs Health Care System. *JAMA Network Open*. 29 Jul 2022;5(7):E2224359. doi:<https://dx.doi.org/10.1001/jamanetworkopen.2022.24359>

46.	Jacobs MME, E.Ellis, C. Racial, ethnic, and sex disparities in the incidence and cognitive symptomology of long COVID-19. *Journal of the National Medical Association*. April 2023;115(2):233-243. doi:<https://dx.doi.org/10.1016/j.jnma.2023.01.016>

47.	Jaramillo M, Thyvalikakath TP, Eckert G, Srinivasan M. Characteristics of Chemosensory Perception in Long COVID and COVID Reinfection. *J Clin Med*. May 22 2023;12(10)doi:10.3390/jcm12103598

48.	Jassat W, Mudara C, Vika C, et al. A cohort study of post-COVID-19 condition across the Beta, Delta, and Omicron waves in South Africa: 6-month follow-up of hospitalized and nonhospitalized participants. *Int J Infect Dis*. Mar 2023;128:102-111. doi:10.1016/j.ijid.2022.12.036

49.	Kahlert CR, Strahm C, Güsewell S, et al. Post-Acute Sequelae After Severe Acute Respiratory Syndrome Coronavirus 2 Infection by Viral Variant and Vaccination Status: A Multicenter Cross-Sectional Study. *Clinical Infectious Diseases*. 2023;77(2):194-202. doi:10.1093/cid/ciad143

50.	Kim YB, S.Chang, H. H.Kim, S. W. Long COVID prevalence and impact on quality of life 2 years after acute COVID-19. Research Support, Non-U.S. Gov't. *Scientific Reports*. 07 11 2023;13(1):11207. doi:<https://dx.doi.org/10.1038/s41598-023-36995-4>

51.	Kubota GTS, F. H. C.da Fonseca, A. S.Rosa, T. D. S.da Silva, V. A.Gouveia, G. R.Faria, V.da Cunha, P. H. M.Brunoni, A. R.Teixeira, M.de Andrade, D. C. Pain paths among post-COVID-19 condition subjects: A prospective cross-sectional study with in-person evaluation. *European Journal of Pain (United Kingdom)*. May 2023;27(5):636-650. doi:<https://dx.doi.org/10.1002/ejp.2094>

52.	Kuodi PG, Y.Zayyad, H.Wertheim, O.Wiegler, K. B.Abu Jabal, K.Dror, A. A.Nazzal, S.Glikman, D.Edelstein, M. Association between BNT162b2 vaccination and reported incidence of post-COVID-19 symptoms: cross-sectional study 2020-21, Israel. *npj Vaccines*. December 2022;7(1) (no pagination)101. doi:<https://dx.doi.org/10.1038/s41541-022-00526-5>

53.	Edelstein M, Kuodi P, Gorelik Y, et al. Association between BNT162b2 vaccination and health-related quality of life up to 18 months post-SARS-CoV-2 infection in Israel: A cross sectional survey. 2023;doi:10.21203/rs.3.rs-2834514/v1

54.	Lam ICH, Wong CKH, Zhang R, et al. Long-term post-acute sequelae of COVID-19 infection: a retrospective, multi-database cohort study in Hong Kong and the UK. *eClinicalMedicine*. 2023;60doi:10.1016/j.eclinm.2023.102000

55.	Lippke S, Rinn R, Derksen C, Dahmen A. Patients' Post-/Long-COVID Symptoms, Vaccination and Functional Status-Findings from a State-Wide Online Screening Study. *Vaccines (Basel)*. Mar 17 2023;11(3)doi:10.3390/vaccines11030691

56.	Luo JZ, J.Tang, H. T.Wong, H. K.Lyu, A.Cheung, C. H.Bian, Z. Prevalence and risk factors of long COVID 6-12 months after infection with the Omicron variant among nonhospitalized patients in Hong Kong. *Journal of Medical Virology*. June 2023;95(6) (no pagination)e28862. doi:<https://dx.doi.org/10.1002/jmv.28862>

57.	Marra AR, Sampaio VS, Ozahata MC, et al. Risk factors for long coronavirus disease 2019 (long COVID) among healthcare personnel, Brazil, 2020-2022. *Infection Control and Hospital Epidemiology*. 2023;doi:<https://dx.doi.org/10.1017/ice.2023.95>

58.	McLaughlin M, Cerexhe L, Macdonald E, et al. A Cross-Sectional Study of Symptom Prevalence, Frequency, Severity, and Impact of Long-COVID in Scotland: Part I. *The American Journal of Medicine*. 2023/07/20/ 2023;doi:<https://doi.org/10.1016/j.amjmed.2023.07.004>

59.	McLaughlin M, Cerexhe L, Macdonald E, et al. A Cross-Sectional Study of Symptom Prevalence, Frequency, Severity, and Impact of Long-COVID in Scotland: Part II. *The American Journal of Medicine*. doi:10.1016/j.amjmed.2023.07.009

60.	Meza-Torres B, Delanerolle G, Okusi C, et al. Differences in Clinical Presentation With Long COVID After Community and Hospital Infection and Associations With All-Cause Mortality: English Sentinel Network Database Study. *JMIR Public Health Surveill*. Aug 16 2022;8(8):e37668. doi:10.2196/37668

61.	Mizrahi B, Sudry T, Flaks-Manov N, et al. Long covid outcomes at one year after mild SARS-CoV-2 infection: nationwide cohort study. *BMJ*. 2023;380:e072529. doi:10.1136/bmj-2022-072529

62.	Mohr NM, Plumb ID, Harland KK, et al. Presence of symptoms 6 weeks after COVID-19 among vaccinated and unvaccinated US healthcare personnel: a prospective cohort study. *BMJ Open*. 2023;13(2):e063141. doi:10.1136/bmjopen-2022-063141

63.	Munipalli BP, S.Mohabbat, A.Siddiqui, H.Li, Z.Abu Dabrh, A. M. Clinical differences in symptomology, characteristics, and risk factors in patients with post-acute sequelae of COVID-19: an experience from a tertiary-care academic center. *Journal of Investigative Medicine*. June 2023;71(5):495-501. doi:<https://dx.doi.org/10.1177/10815589231153604>

64.	Nascimento TCDCdVC, L.Ruiz, A. D.Ledo, C. B.Fernandes, V. P. L.Cardoso, L. F.Junior, J. M. V.Saretta, R.Kalil-Filho, R.Drager, L. F. Vaccination status and long COVID symptoms in patients discharged from hospital. *Scientific reports*. 11 Feb 2023;13(1):2481. doi:<https://dx.doi.org/10.1038/s41598-023-28839-y>

65.	Naveed Z, Velásquez García HA, Wong S, et al. Association of COVID-19 Infection With Incident Diabetes. *JAMA Network Open*. 2023;6(4):e238866-e238866. doi:10.1001/jamanetworkopen.2023.8866

66.	Nehme MB, O.Salamun, J.Jacquerioz, F.Courvoisier, D. S.Spechbach, H.Guessous, I. Symptoms After COVID-19 Vaccination in Patients with Post-Acute Sequelae of SARS-CoV-2. *Journal of General Internal Medicine*. May 2022;37(6):1585-1588. doi:<https://dx.doi.org/10.1007/s11606-022-07443-2>

67.	Nehme M, Vetter P, Chappuis F, Kaiser L, Guessous I. Prevalence of Post-Coronavirus Disease Condition 12 Weeks After Omicron Infection Compared With Negative Controls and Association With Vaccination Status. *Clin Infect Dis*. May 3 2023;76(9):1567-1575. doi:10.1093/cid/ciac947

68.	Parodi JB, Indavere A, Bobadilla Jacob P, et al. Impact of COVID-19 vaccination in post-COVID cardiac complications. *Vaccine*. 17 Feb 2023;41(8):1524-1528. doi:<https://dx.doi.org/10.1016/j.vaccine.2023.01.052>

69.	Patel NJ, Cook C, Vanni K, et al. Impact of vaccination on postacute sequelae of SARS CoV-2 infection in patients with rheumatic diseases. *Annals of the Rheumatic Diseases*. 01 Apr 2023;82(4):565-573. doi:<https://dx.doi.org/10.1136/ard-2022-223439>

70.	Peghin MDM, M.Palese, A.Gerussi, V.Bontempo, G.Graziano, E.Visintini, E.D'Elia, D.Dellai, F.Marrella, F.Fabris, M.Curcio, F.Sartor, A.Isola, M.Tascini, C. Post-COVID-19 syndrome and humoral response association after 1 year in vaccinated and unvaccinated patients. *Clinical Microbiology and Infection*. August 2022;28(8):1140-1148. doi:<https://dx.doi.org/10.1016/j.cmi.2022.03.016>

71.	Peghin M, De Martino M, Palese A, et al. Post-COVID-19 Syndrome 2 Years After the First Wave: The Role of Humoral Response, Vaccination and Reinfection. *Open Forum Infect Dis*. Jul 2023;10(7):ofad364. doi:10.1093/ofid/ofad364

72.	Perlis RH, Santillana M, Ognyanova K, et al. Prevalence and Correlates of Long COVID Symptoms Among US Adults. *JAMA Network Open*. 27 Oct 2022;5(10):E2238804. doi:<https://dx.doi.org/10.1001/jamanetworkopen.2022.38804>

73.	Pinato DJ, Ferrante D, Aguilar-Company J, et al. Vaccination against SARS-CoV-2 protects from morbidity, mortality and sequelae from COVID19 in patients with cancer. *Eur J Cancer*. Aug 2022;171:64-74. doi:10.1016/j.ejca.2022.04.036

74.	Qasmieh SA, Robertson MM, Teasdale CA, et al. The prevalence of SARS-CoV-2 infection and long COVID in U.S. adults during the BA.4/BA.5 surge, June-July 2022. *Prev Med*. Apr 2023;169:107461. doi:10.1016/j.ypmed.2023.107461

75.	Rahman A, Ikram MT, Arif A, et al. Prevalence and difference of COVID-19 symptoms, post-COVID conditions and duration of illness among the vaccinated and unvaccinated population: a cross-sectional study in Peshawar. *Ann Med Surg (Lond)*. May 2023;85(5):1678-1684. doi:10.1097/ms9.0000000000000606

76.	Ranucci M, Baryshnikova E, Anguissola M, et al. The Very Long COVID: Persistence of Symptoms after 12-18 Months from the Onset of Infection and Hospitalization. *J Clin Med*. Feb 28 2023;12(5)doi:10.3390/jcm12051915

77.	Resendez S, Brown SH, Sebastian Ruiz H, et al. Defining the Subtypes of Long COVID and Risk Factors for Prolonged Disease. *medRxiv*. 2023:2023.05.19.23290234. doi:10.1101/2023.05.19.23290234

78.	Richard SA, Pollett SD, Fries AC, et al. Persistent COVID-19 Symptoms at 6 Months after Onset and the Role of Vaccination before or after SARS-CoV-2 Infection. *JAMA Network Open*. 18 Jan 2023;6(1):E2251360. doi:<https://dx.doi.org/10.1001/jamanetworkopen.2022.51360>

79.	Robertson MM, Qasmieh SA, Kulkarni SG, et al. The Epidemiology of Long Coronavirus Disease in US Adults. *Clin Infect Dis*. May 3 2022;76(9):1636-1645. doi:10.1093/cid/ciac961

80.	Scherlinger MP, L.Chatelus, E.Arnaud, L.Gottenberg, J. E.Sibilia, J.Felten, R. Effect of SARS-CoV-2 Vaccination on Symptoms from Post-Acute Sequelae of COVID-19: Results from the Nationwide VAXILONG Study. *Vaccines*. January 2022;10(1) (no pagination)46. doi:<https://dx.doi.org/10.3390/vaccines10010046>

81.	Senjam SS, Balhara YPS, Kumar P, et al. A comprehensive assessment of self-reported post COVID-19 symptoms among beneficiaries of hospital employee scheme at a tertiary healthcare institution in Northern India. *International Journal of General Medicine*. 2022;15:7355.

82.	Smits PD, Rodriguez PJ, Gratzl S, et al. Effect of vaccination on time till Long COVID, a comparison of two ways to model effect of vaccination and two outcome definitions. *medRxiv*. 2023:2023.04.28.23289271. doi:10.1101/2023.04.28.23289271

83.	Strain WDS, O.Banerjee, A.Van der Togt, V.Hishmeh, L.Rossman, J. The Impact of COVID Vaccination on Symptoms of Long COVID: An International Survey of People with Lived Experience of Long COVID. *Vaccines*. May 2022;10(5) (no pagination)652. doi:<https://dx.doi.org/10.3390/vaccines10050652>

84.	Tannous J, Pan AP, Potter T, et al. Real-world effectiveness of COVID-19 vaccines and anti-SARS-CoV-2 monoclonal antibodies against postacute sequelae of SARS-CoV-2: Analysis of a COVID-19 observational registry for a diverse US metropolitan population. *BMJ Open*. 05 Apr 2023;13(4) (no pagination)e067611. doi:<https://dx.doi.org/10.1136/bmjopen-2022-067611>

85.	Taquet MD, Q.Harrison, P. J. Six-month sequelae of post-vaccination SARS-CoV-2 infection: A retrospective cohort study of 10,024 breakthrough infections. *Brain, Behavior, and Immunity*. July 2022;103:154-162. doi:<https://dx.doi.org/10.1016/j.bbi.2022.04.013>

86.	Tosi DT, F.Brito, Y.Sarasua, A.Ruiz, J.Hammel, I. S. Vaccination as a protective factor for PASC in Frail, Pre-frail, and Robust Veterans During the Omicron Wave. Conference Abstract. *Journal of the American Geriatrics Society*. April 2023;71(Supplement 1):S158-S159. doi:<https://dx.doi.org/10.1111/jgs.18336>

87.	Tran VTP, E.Saldanha, J.Pane, I.Ravaud, P. Efficacy of first dose of covid-19 vaccine versus no vaccination on symptoms of patients with long covid: target trial emulation based on ComPaRe e-cohort. *BMJ Med*. 2023;2(1):e000229. doi:<https://dx.doi.org/10.1136/bmjmed-2022-000229>

88.	Tsuchida T, Hirose M, Inoue Y, Kunishima H, Otsubo T, Matsuda T. Relationship between changes in symptoms and antibody titers after a single vaccination in patients with Long COVID. *J Med Virol*. Jul 2022;94(7):3416-3420. doi:10.1002/jmv.27689

89.	van der Maaden T, Mutubuki EN, de Bruijn S, et al. Prevalence and Severity of Symptoms 3 Months After Infection With SARS-CoV-2 Compared to Test-Negative and Population Controls in the Netherlands. *J Infect Dis*. Apr 26 2023;227(9):1059-1067. doi:10.1093/infdis/jiac474

90.	Wanga VC, J. R.Dimitrov, L. V.Gerdes, M. E.Whitfield, G. P.Bonacci, R. A.Nji, M. A. M.Hernandez-Romieu, A. C.Rogers-Brown, J. S.McLeod, T.Rushmore, J.Lutfy, C.Bushman, D.Koumans, E.Saydah, S.Goodman, A. B.Coleman King, S. M.Jackson, B. R.Cope, J. R. Long-Term Symptoms Among Adults Tested for SARS-CoV-2 - United States, January 2020-April 2021. *Mmwr*. 10 Sep 2021;Morbidity and mortality weekly report. 70(36):1235-1241. doi:<https://dx.doi.org/10.15585/mmwr.mm7036a1>

91.	Whittaker HR, Gulea C, Koteci A, et al. GP consultation rates for sequelae after acute covid-19 in patients managed in the community or hospital in the UK: population based study. *BMJ*. 2021;375:e065834. doi:10.1136/bmj-2021-065834

92.	Wisnivesky JPG, U.Bagiella, E.Goswami, R.Kale, M.Campbell, K. N.Meliambro, K.Chen, Z.Aberg, J. A.Lin, J. J. Association of Vaccination with the Persistence of Post-COVID Symptoms. *Journal of General Internal Medicine*. May 2022;37(7):1748-1753. doi:<https://dx.doi.org/10.1007/s11606-022-07465-w>

93.	Wong MCS, Huang J, Wong YY, et al. Epidemiology, Symptomatology, and Risk Factors for Long COVID Symptoms: Population-Based, Multicenter Study. *JMIR public health and surveillance*. 07 Mar 2023;9:e42315. doi:<https://dx.doi.org/10.2196/42315>

94.	Wynberg EH, A. X.Boyd, A.van Willigen, H. D. G.Verveen, A.Lebbink, R.van der Straten, K.Kootstra, N.van Gils, M. J.Russell, C.Leenstra, T.de Jong, M. D.de Bree, G. J.Prins, M. The effect of SARS-CoV-2 vaccination on post-acute sequelae of COVID-19 (PASC): A prospective cohort study. *Vaccine*. 30 Jul 2022;40(32):4424-4431. doi:<https://dx.doi.org/10.1016/j.vaccine.2022.05.090>

95.	Xue P, Merikanto I, Chung F, et al. Persistent short nighttime sleep duration is associated with a greater post-COVID risk in fully mRNA-vaccinated individuals. *Translational Psychiatry*. December 2023;13(1) (no pagination)32. doi:<https://dx.doi.org/10.1038/s41398-023-02334-4>

96.	Zhang D, Chung VC, Chan DC, et al. Determinants of post-COVID-19 symptoms among adults aged 55 or above with chronic conditions in primary care: data from a prospective cohort in Hong Kong. *Front Public Health*. 2023;11:1138147. doi:10.3389/fpubh.2023.1138147

97.	Zisis SN, Durieux JC, Mouchati C, Perez JA, McComsey GA. The Protective Effect of Coronavirus Disease 2019 (COVID-19) Vaccination on Postacute Sequelae of COVID-19: A Multicenter Study From a Large National Health Research Network. *Open Forum Infect Dis*. Jul 2022;9(7):ofac228. doi:10.1093/ofid/ofac228
